# The Psychosis Human Connectome Project: Design and rationale for studies of visual neurophysiology

**DOI:** 10.1101/2022.09.16.22280014

**Authors:** Michael-Paul Schallmo, Kimberly B. Weldon, Rohit S. Kamath, Hannah R. Moser, Samantha A. Montoya, Kyle W. Killebrew, Caroline Demro, Andrea N. Grant, Małgorzata Marjańska, Scott R. Sponheim, Cheryl A. Olman

## Abstract

Visual perception is abnormal in psychotic disorders such as schizophrenia. In addition to hallucinations, laboratory tests show differences in fundamental visual processes including contrast sensitivity, center-surround interactions, and perceptual organization. A number of hypotheses have been proposed to explain visual dysfunction in psychotic disorders, including an imbalance between excitation and inhibition. However, the precise neural basis of abnormal visual perception in people with psychotic psychopathology (PwPP) remains unknown. Here, we describe the behavioral and 7 tesla MRI methods we used to interrogate visual neurophysiology in PwPP as part of the Psychosis Human Connectome Project (HCP). In addition to PwPP (n = 66) and healthy controls (n = 43), we also recruited first-degree biological relatives (n = 44) in order to examine the role of genetic liability for psychosis in visual perception. Our visual tasks were designed to assess fundamental visual processes in PwPP, whereas MR spectroscopy enabled us to examine neurochemistry, including excitatory and inhibitory markers. We show that it is feasible to collect high-quality data across multiple psychophysical, functional MRI, and MR spectroscopy experiments with a sizable number of participants at a single research site. These data, in addition to those from our previously described 3 tesla experiments, will be made publicly available in order to facilitate further investigations by other research groups. By combining visual neuroscience techniques and HCP brain imaging methods, our experiments offer new opportunities to investigate the neural basis of abnormal visual perception in PwPP.

## Introduction and background

Abnormal visual perception is a symptom of psychosis spectrum disorders (e.g., schizophrenia), and includes both frank hallucinations and distorted perception of actual stimuli. The point prevalence of visual hallucinations in schizophrenia is 27%, and such anomalies are associated with greater disease severity and poorer outcomes (Waters et al., 2014). Laboratory assessments of visual perception have also shown more subtle abnormalities in people with psychotic psychopathology (for reviews, see Butler et al., 2008; King et al., 2017; Klein et al., 2020a; Notredame et al., 2014; Phillips and Silverstein, 2013; Yoon et al., 2013). While much is known about the neural mechanisms of visual perception in healthy individuals, it is not yet clear what differences in neural processing underlie visual dysfunction in people with psychotic psychopathology (PwPP), which limits the development of more effective treatment strategies for psychotic illness (e.g., to improve sensory functioning). A number of hypotheses have been proposed to explain visual disturbances in PwPP, including a disrupted balance of excitation and inhibition (Foss-Feig et al., 2017; Lewis et al., 2005; Lisman, 2012; Moghaddam and Javitt, 2012), thalamo-cortical dysconnectivity (Anticevic et al., 2014; Cheng et al., 2015; Damaraju et al., 2014; Dong et al., 2019; Giraldo-Chica and Woodward, 2017; Ramsay, 2019; Ramsay et al., 2017), abnormal visual gain control (Butler et al., 2008; Phillips and Silverstein, 2013), impaired top-down attentional processing (Gold et al., 2018; Luck et al., 2019a; Luck et al., 2019b), and disrupted predictive coding (Adams et al., 2013; Horga and Abi-Dargham, 2019; Sterzer et al., 2018). The current study was designed to provide a publicly available, multimodal neuroimaging data set to facilitate testing these (and other) hypotheses regarding the nature of visual dysfunction among PwPP.

There is additional motivation for studying visual functioning among PwPP, as the visual system offers a strong translational bridge between basic neuroscience in animal models and studies in humans with psychosis. There is significant homology between the visual systems of well-studied animal models (e.g., cats, ferrets, and non-human primates) and humans (Hubel and Wiesel, 1962; Van Essen et al., 1992). It is also relatively straightforward to adapt visual paradigms from animal models for use in human studies, and vice versa. This allows investigators to make strong inferences about the neurophysiological basis of perceptual anomalies seen in PwPP, which may be challenging when studying higher-order cognitive functions. In this study, we used visual paradigms designed to tap into several different aspects or levels of visual functioning. We provide further details on the background and motivation for the specific visual paradigms that we chose for this study in the Supplemental Information.

One theory that has been put forth to explain impaired visual perception in psychosis spectrum disorders is an imbalance between excitation and inhibition (E/I) within visual brain regions, with many suggesting excess excitation, reduced inhibition, or both (Foss-Feig et al., 2017; Lisman, 2012). The idea of E/I imbalance has the advantage of merging two neurochemical theories of schizophrenia (or psychotic disorders more generally) that have received widespread attention and support: the glutamate hypothesis (Javitt, 2004; Moghaddam and Javitt, 2012) and the γ-aminobutyric acid (GABA) hypothesis (Gonzalez-Burgos and Lewis, 2008; Hashimoto et al., 2008; Lewis et al., 2005). ^1^H magnetic resonance spectroscopy (MRS) is a popular method for investigating E/I functions non-invasively in the human brain that can be used to measure concentrations of various neurochemicals, including excitatory (i.e., glutamate) and inhibitory (i.e., GABA) neurotransmitters. Both standard and edited MRS techniques have been developed; the former provide measures of the full MR spectrum (after water suppression; Marjańska et al., 2017; Tkáč et al., 2001), whereas the latter are designed to measure concentrations of specific metabolites (e.g., GABA, via the MEGA-PRESS technique; Mescher et al., 1998) by canceling or ‘editing’ out other signals. MRS studies have provided somewhat mixed support for an E/I imbalance within visual regions in PwPP (Sydnor and Roalf, 2020; Taylor and Tso, 2015), with some suggesting abnormal glutamate and / or GABA levels in visual cortex among PwPP (Kelemen et al., 2013; Thakkar et al., 2017; Yoon et al., 2020; Yoon et al., 2010) and others reporting no differences versus controls (Kumar et al., 2020; Marsman et al., 2014).

There are some important methodological considerations for MR spectroscopy studies of E/I balance in PwPP, including the challenge of separating signals from neurochemicals of interest from those attributable to macromolecules (Cudalbu et al., 2021). In schizophrenia, faster metabolite transverse relaxation time constants (*T*_2_) have also been reported (Öngür et al., 2010). This raises the possibility that reports of lower metabolite concentrations among PwPP (versus controls) in MRS studies using sequences with longer echo times (TEs) might reflect a difference in *T*_2_, rather than true group differences in metabolite concentrations. In the current study, we used an ultra-short (8 ms) TE STEAM sequence (Marjańska et al., 2017; Tkáč et al., 2001), in order to address the potentially confounding effects of shorter *T*_2_ in PwPP. We also explicitly accounted for the contribution of macromolecules within our spectra based on measurements from inversion-recovery experiments (Marjańska et al., 2017).

Genetic factors play an important role in the development of psychotic disorders (Cardno et al., 1999; Cardno and Owen, 2014), but the link between genetic liability for psychotic illness and disordered visual perception remains unclear. Studying the first-degree biological relatives of PwPP (i.e., parents, siblings or children, who share on average 50% of their genes) may provide insight into the genetic basis of visual processing abnormalities in psychotic disorders. Visual task performance and physiological measures of visual processing in relatives may fall on a continuum between typical functioning among controls and the impairments observed among PwPP (Chkonia et al., 2010; Kéri et al., 2001; Klein et al., 2020b; Pokorny et al., 2021a; Pokorny et al., 2021b; Schallmo et al., 2013; Schallmo et al., 2015; Sponheim et al., 2006; Yeap et al., 2006). If consistent abnormalities in visual behavior and / or neural processing can be identified across both PwPP and their biological relatives, then such differences may be able to serve as endophenotypes (Calkins et al., 2008; Gottesman and Gould, 2003; Iacono et al., 2017), which could help elucidate the genetic basis of abnormal visual perception in PwPP, for example by aiding the development of animal models.

We chose to take a trans-diagnostic approach to studying visual functioning among PwPP, rather than focusing on specific diagnostic categories (e.g., schizophrenia vs. bipolar disorder). Because the reliability and validity of discrete categories, as defined by the Diagnostic and Statistical Manual of Mental Disorders (DSM), have been criticized (Kotov et al., 2017; Markon et al., 2011), we sought to examine the neurophysiology of visual dysfunction among PwPP more generally, as well as their first-degree biological relatives. Our approach is informed by the National Institute of Mental Health’s Research Domain Criteria (RDoC) framework (Cuthbert, 2014), and by the notion that there may be a spectrum of disrupted visual functioning among PwPP and their biological relatives that extends across diagnostic categories and includes genetic liability for psychosis. We hope that our work will help to clarify the etiology of visual dysfunction in psychosis spectrum conditions.

The primary focus of the present study was to examine behavioral and neurophysiological markers of visual functioning in PwPP and their biological relatives. This investigation was carried out using behavioral and 7 T imaging measures as part of the Psychosis Human Connectome Project. These data were collected from the same study population as the 3 T and clinical measures described in our recent publication (Demro et al., 2021). Similar to the original Young Adult HCP (Benson et al., 2018; Glasser et al., 2016; Van Essen et al., 2013; Vu et al., 2017), we collected 7 T fMRI data during a ‘sweeping bars’ paradigm to facilitate population receptive field (pRF) mapping and functional definition of retinotopic early visual area boundaries. We also conducted experiments using visual paradigms focused on aspects of visual perception that may differ among PwPP, as detailed above. Before beginning this study, we solicited input from a small number of experts in the fields of vision science and psychosis research in order to select visual paradigms that might provide valuable and complementary insight into such abnormalities. The paradigms that we selected (in addition to pRF mapping) were contrast surround suppression (CSS), contour object perception (COP), and a bi-stable structure-from-motion (SFM) task (behavioral data only), the details of which are described below. Acquiring our fMRI data at 7 T allowed us to achieve higher functional contrast-to-noise and higher spatial resolution versus comparable 3 T methods (Vu et al., 2017), which is advantageous for examining fMRI responses from retinotopically organized visual areas. Further, we conducted 7 T MR spectroscopy (MRS) experiments to measure neurochemical levels within occipital (OCC) and prefrontal (PFC) cortices, in order to examine hypotheses including whether abnormal E/I balance contributes to visual dysfunction in psychotic disorders (Foss-Feig et al., 2017; Lisman, 2012). The OCC region was chosen to include areas in early visual cortex that may be relevant to abnormal visual perception in PwPP, whereas the PFC region was selected for its potential role in higher cognitive dysfunction in PwPP. Using 7 T MRS provided higher signal-to-noise ratio (SNR) and greater ability to study individual metabolites (e.g., separating glutamate and glutamine, reliably quantifying GABA), in comparison to similar MRS methods at 3 T (Godlewska et al., 2017; Terpstra et al., 2016). Repeat scan data from all experiments were acquired from a subset of participants in order to examine longitudinal variability in our behavioral and physiological measures, and any associations with changes in psychiatric symptoms over time. This paper describes the motivation, methods, data quality, and general pattern of results (from controls) for each of our 7 T experimental paradigms. By making our code and data publicly available, we hope to facilitate further investigations of abnormal visual processing in PwPP, which may build upon the work described here.

## Methods

### Participants

Data were collected from three groups of participants: healthy controls with no family history of psychosis (n = 44; controls hereafter), first-degree biological relatives of a person with a history of psychotic psychopathology (n = 46; relatives hereafter), and people with a history of psychotic psychopathology (n = 68; PwPP hereafter). Of these, 1 control, 2 relatives, and 2 PwPP were excluded from our study after completing the experiments described below (e.g., found to have a visual abnormality). Data from excluded individuals are not presented here, unless otherwise noted. Our final sample size was 43 controls, 44 relatives, and 66 PwPP. Demographic and psychiatric symptom data for each group are presented in Table 1. In Supplemental Table 1, data for PwPP are presented within three diagnostic sub-groups (people with schizophrenia, n = 36; those with schizoaffective disorder, n = 10; and those with bipolar I disorder with psychotic features, n = 20).

**Table 1.**
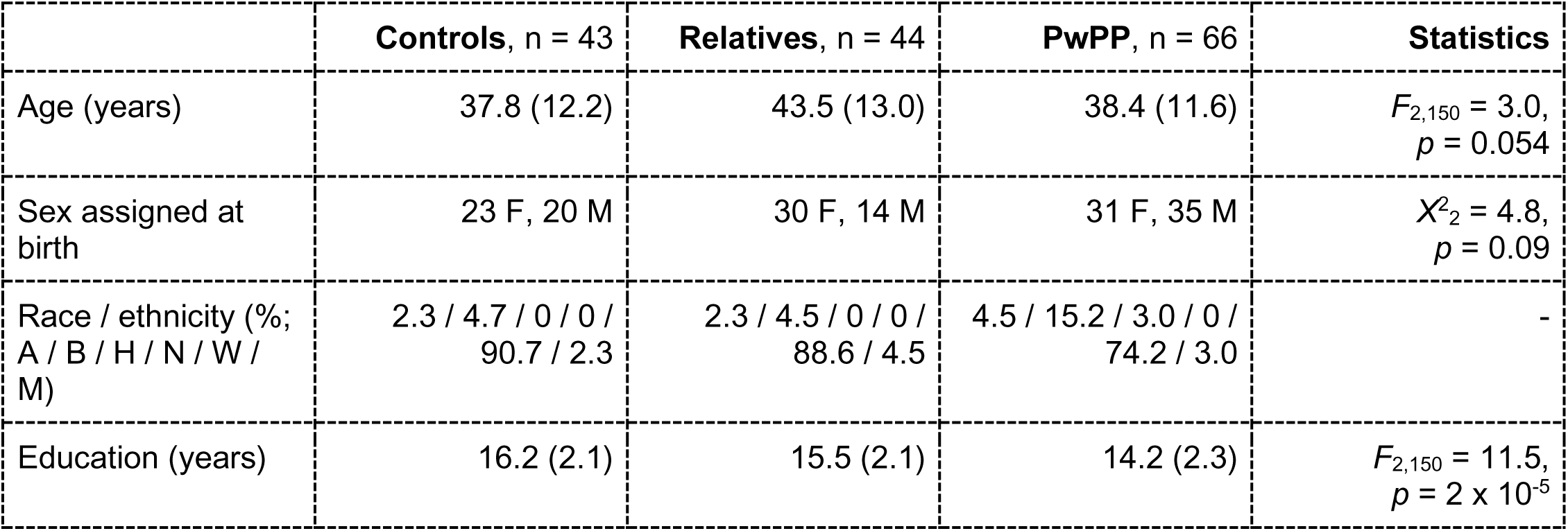
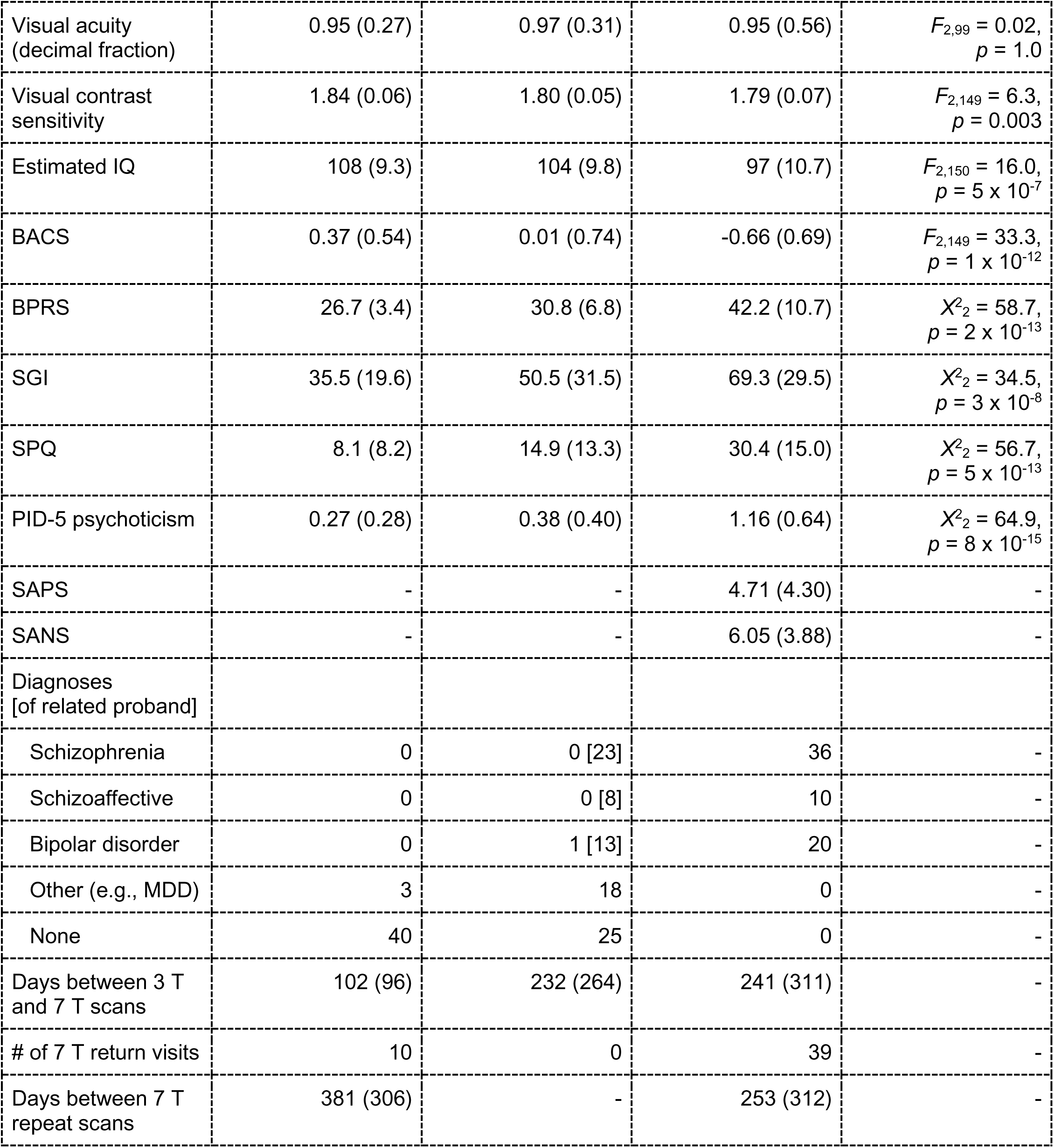
Participant demographics, cognitive, and symptom measures. Data are presented as mean (standard deviation), unless otherwise noted. Racial and ethnic designations (as defined by the National Institute of Health) are abbreviated as follows: A = Asian or Pacific Islander, B = Black (not of Hispanic origin), H = Hispanic, N = Native American or Alaskan Native, W = White (not of Hispanic origin), M = More than 1 race or ethnicity, or other. Visual acuity was assessed with a Snellen eye chart (Snellen, 1862); the decimal fraction is reported (e.g., 0.5 indicates 20/40). Visual contrast sensitivity was assessed with the Mars Letter Contrast Sensitivity test (Arditi, 2005). Estimated IQ was assessed using the Wechsler Adult Intelligence Scale (WAIS-IV; Wechsler, 2008). BACS = Brief Assessment of Cognition in Schizophrenia, Z-score (Keefe et al., 1999), BPRS = Brief Psychiatric Rating Scale Total Score (Ventura et al., 2000), SGI = Sensory Gating Inventory (Hetrick et al., 2012), SPQ = Schizotypal Personality Questionnaire (Raine, 1991), PID-5 psychoticism = psychoticism factor from the Personality Inventory for DSM-5 (Krueger et al., 2012), SANS = Scale for the Assessment of Negative Symptoms Total Global Score (Andreasen, 1982), SAPS = Scale for the Assessment of Positive Symptoms Total Global Score (Andreasen, 1984). Diagnoses were based on the Structured Clinical Interview for DSM-IV-TR disorders (SCID; First, 1997). Data collected at repeat scans were not included here. For the relative group, the number of related probands with a particular psychotic disorder diagnosis is listed in square brackets. The statistics column shows the test statistic and *p*-value for differences across all three groups in each measure. For measures where normality and / or homogeneity of variance were not observed, non-parametric Kruskal-Wallis tests (*Χ*^2^-values) were used in place of analyses of variance (*F*-values).

Inclusion criteria for the Psychosis Human Connectome Project have been described previously (Demro et al., 2021; Schallmo et al., 2021). These criteria included age 18-65 years, English as a primary language, the ability to provide written informed consent, no legal guardian, no diagnosed learning disability or IQ less than 70, no current or past central nervous system disease, no history of head injury with skull fracture or loss of consciousness longer than 30 minutes, no alcohol or drug abuse within the last 2 weeks, or dependence within the last 6 months, no electroconvulsive therapy within the last year, no tardive dyskinesia, no visual or hearing impairment, no condition that would physically inhibit task performance such as paralysis or severe arthritis. All participants in the PwPP group had a history of schizophrenia, schizoaffective disorder, or bipolar I disorder with psychotic features and were not adopted. Relatives had a biological parent, sibling, or child with a history of one of these disorders (but not necessarily an enrolled participant from the PwPP group) and were not adopted. Controls had no personal or immediate family history (i.e., parents, siblings, children) of psychosis spectrum disorders. Additional inclusion criteria for 7 T scanning included the ability to fit comfortably within the scanner bore (60 cm diameter) and the radio frequency (RF) head coil (head circumference less than 62 cm), weight less than 440 pounds, and corrected Snellen visual acuity of 20/40 (decimal fraction = 0.5) or better. All participants had completed two 3 T fMRI scanning sessions prior to 7 T scanning. Individuals who exceed a limit of 0.5 mm of head motion per TR in greater than 20% of TRs from all 3 T fMRI runs (approximately 2 hours of scanning) were excluded prior to 7 T scanning.

All participants provided written informed consent prior to their participation and were compensated approximately $20 / hour for their time. As part of the informed consent process, participants agreed to have their de-identified data shared publicly, including the dates of all research study visits. All experimental procedures complied with the Declaration of Helsinki and were approved by the University of Minnesota IRB. All participants had sufficient capacity to provide informed consent, as assessed by the University of California Brief Assessment of Capacity to Consent (Jeste et al., 2007).

### Study timeline

The time between 3 T and 7 T scanning is shown for each group in Table 1. Typically, the minimum delay was at least 1 week, owing to the need for quality assessment of the 3 T data for screening purposes (as described above, participants with poor 3 T data quality were excluded from 7 T scanning). We conducted repeat 7 T scans for a subset of PwPP and a very limited number of controls (Table 1). This was done in order to permit longitudinal assessment of neural and behavioral measures from the 7 T experiments (i.e., visual behavior, fMRI, MRS), and psychiatric symptom measures acquired on the day of 7 T scanning (i.e., BPRS, SANS, SAPS; see below). The time between the first and second 7 T scans for controls and PwPP is shown in Table 1.

The nomenclature for our different scanning sessions bears explanation. Our first iteration of the protocol was termed version ‘A.’ 7T-A scans began in July 2017 and did not include MRS data from prefrontal cortex, as the radiofrequency (RF) head coil used for these scans was not yet available. Other data are also missing from some early 7T-A scans due to technical issues (for information about the number of datasets available from each protocol version, see Supplemental Table 2). In January 2018, we began collecting data from the second iteration of our experimental protocol, which we refer to as 7T-B. Here, we added MRS data collection from a region of the dorsomedial prefrontal cortex, as described in the MR spectroscopy section below. We also made some small changes to the visual stimuli and paradigms in the CSS (psychophysics) and COP (psychophysics and fMRI) tasks (see below and Supplemental Information). We refer to repeat scans in which the 7T-B protocol was repeated as 7T-Z. In some cases, participants completed both 7T-A and 7T-B protocols; for these, the second scan is referred to as 7T-B. In other cases, individuals participated in both 7T-B and 7T-Z scans, but did not complete the full scanning protocol during 7T-B. In these cases, the scan data from the second visit are still labeled as 7T-Z, even though the 7T-B imaging data are incomplete or missing.

### Apparatus

Our psychophysical (quantitative behavioral) experiments were conducted in a darkened laboratory on an Apple Mac Pro and an Eizo FlexScan SX2462W monitor with a 60 Hz refresh rate (mean luminance = 61.2 cd/m^2^). A Bits# stimulus processor (Cambridge Research Systems) was also used; when in Mono++ mode, this provided 9.6-bit luminance resolution (for the CSS psychophysical task, see below). Participants were seated in a height-adjustable chair with an adjustable chin rest to maintain a stable head position at a viewing distance of 70 cm. Stimuli were generated and presented using PsychoPy (version 1.85.2; Peirce, 2007).

Monitor luminance was linearized using a PR655 (Photoresearch, JADAK, North Syracuse, NY) spectrophotometer. 7 T MR data were acquired on a Siemens MAGNETOM scanner. This scanner was equipped with an 8-kW RF power amplifier and body gradients with 70 mT/m maximum amplitude and 200 T/m/s maximum slew rate. The scanner software was upgraded from VB17 to VE12U in July 2019. Data quality before and after the upgrade was comparably high (e.g., temporal SNR, image SNR; data not shown). For both MRS and fMRI, participants were given head, neck, and lumbar padding, and instructed to minimize movement during scanning. For our MRS data acquisition, we used a custom-built RF head coil with two surface ^1^H quadrature transceivers, one for the front of the brain and one for the back of the brain. Participants were removed from the scanner in between occipital and prefrontal MRS scans (during the same scanning session), in order to switch the coil configuration. A 7 T MR-compatible motion tracking system (Metria Innovation Inc., Milwaukee, WI) was used to track a Moiré Phase Tracking marker attached to the participant’s face in order to measure head motion during MRS (but not during fMRI). Head motion was observed by an experimenter in real time. When head motion > 5 mm was observed, MRS scans were repeated, and participants were encouraged to remain still.

Participants were removed from the scanner in between MRS and fMRI data acquisition, in order to change scanning equipment. For fMRI data acquisition, we used a Nova Medical (Wilmington, MA) RF head coil (1 transmit and 32 receive channels). We used 5 mm thick dielectric pads (3:1 calcium titanate powder in water) positioned under the neck and beside the temples during the fMRI experiments, when possible (allowing for the participant’s comfort). Previous 7 T MRI studies as part of the HCP have shown this improves transmit *B*1 homogeneity in the cerebellum and temporal lobe regions (Vu et al., 2017).

During fMRI scanning, visual stimuli were presented using an NEC projector with a 60 Hz refresh rate (mean luminance = 271 cd/m^2^). Participants viewed stimuli projected onto a 3M (Maplewood, MN) Vikuiti acrylic screen at the back of the bore through a mirror mounted to the head coil, with a viewing distance of 100 cm. Participants used a Current Designs (Philadelphia, PA) MR-compatible 4-button response box during the visual tasks. Auditory stimuli were delivered through Sensimetrics (Gloucester, MA) earbuds, placed in the participant’s auditory canals by research staff and covered by medical tape, to minimize the possibility that the earbuds became dislodged while the participant was getting into position in the scanner. We confirmed that participants could hear auditory stimuli presented through the stimuli prior to scanning.

### Experimental protocol

#### Clinical measures

We collected questionnaire and interview-based measures related to clinical psychiatric status outside of the scanner, prior to the MR experiments. These included the Brief Psychiatric Rating Scale (BPRS, 24 item version; Ventura et al., 2000), the Scale for the Assessment of Negative Symptoms (SANS; Andreasen, 1982), the Scale for the Assessment of Positive Symptoms (SAPS; Andreasen, 1984), the Positive and Negative Affect Schedule (PANAS; Watson et al., 1988), and a questionnaire about sleep habits and recent use of prescription and non-prescription drugs. BPRS, PANAS, and the sleep and recent use questionnaire were collected for all individuals, while SAPS and SANS were collected only for PwPP. Because the BPRS, SAPS, and SANS reflect clinical symptom levels over the past month, these measures were collected on the day of 7 T scanning, unless the participant had completed them during another research visit within 30 days of the 7 T scan. These measures were collected at each visit for participants who completed two scanning sessions (e.g., 7T-B and 7T-Z). We observed a modest degree of variability in BPRS, SAPS, and SANS scores among PwPP between the first and second scanning sessions (Supplemental Table 3 and Supplemental Figure 2) indicating that symptoms levels varied to some extent over time within this group, as expected (Long and Brekke, 1999). Other clinical data used in this study were collected on a separate clinical research visit on a different day, prior to the 7 T session. These included the Sensory Gating Inventory (SGI; Hetrick et al., 2012), the Schizotypal Personality Questionnaire (SPQ; Raine, 1991), and the Personality Inventory for DSM-5 (PID-5; Krueger et al., 2012), which includes a psychoticism factor score. Full details of our clinical measures are reported in our companion paper (Demro et al., 2021). Table 1 shows a summary of the data from different clinical measures for each group.

Prior to completing visual psychophysics and 7 T MR experiments, the participant’s visual acuity was assessed using a Snellen eye chart (Snellen, 1862) from a viewing distance of 100 cm and a height of 140 cm from the floor. Acuity data are presented in Table 1. Participants wore corrective lenses as needed; MR compatible frames and lenses were provided, and were worn during all experiments on the 7 T scanning day. The experimental protocol was discontinued for participants who did not demonstrate visual acuity (including correction) of 20/40 (decimal fraction = 0.5) or better. Other visual measures were collected from all participants during an earlier, clinically focused study visit. These include: visual questionnaires from the NIH PhenX Toolkit (i.e., Contact Lens Use, Personal and Family History of Eye Disease and Treatments, Personal and Family History of Strabismus, Visual Function questions about eyesight and driving), Mars Letter Contrast Sensitivity test (Arditi, 2005), and the Farnsworth Dichotomous Color Vision test (Farnsworth, 1943), as described in our companion paper (Demro et al., 2021).

#### Tasks

##### Population receptive field (pRF)

Each participant completed at least one population receptive field (pRF) mapping scan (Dumoulin and Wandell, 2008) for the purpose of retinotopic mapping in visual areas. During the task, participants were asked to maintain fixation on a central point while a bar moved across the visual field at one of eight orientations (West (W) – East (E), North (N) - South (S), NW - SE, NE - SW, E - W, SE - NW, S - N, SW - NE; Figure 1A). The moving bar was populated with dynamic and highly salient visual stimuli from Kriegeskorte and colleagues (2008). The 96 images were divided into 7 categories: human face, human body, animal face, whole animal, food, manipulable objects, and places. An 8th category, noise, was created by Fourier phase-scrambling the three color channels of each image and re-combining the channels to make brightly colored textures with spatial frequencies similar to the object images. Each category was presented during two bar sweeps, chosen randomly. Reduced-contrast noise images were present as a background during all image categories to ensure coverage of the entire bar while minimizing crowding between object images. The bars flickered at either 2 or 12 Hz. The length of the bars spanned a 16° disk and subtended 2° of visual angle in width and traveled across the visual field out to 8° eccentricity. Each bar took 16 s to complete the movement across the visual field, and each bar sweep direction occurred twice in a single scan, once with a 2 Hz refresh rate and once at 12 Hz. There were 4 s of rest between each bar sweep and 4 s of rest at the beginning and end of each scan, such that each scan took 324 s.

**Figure 1.**
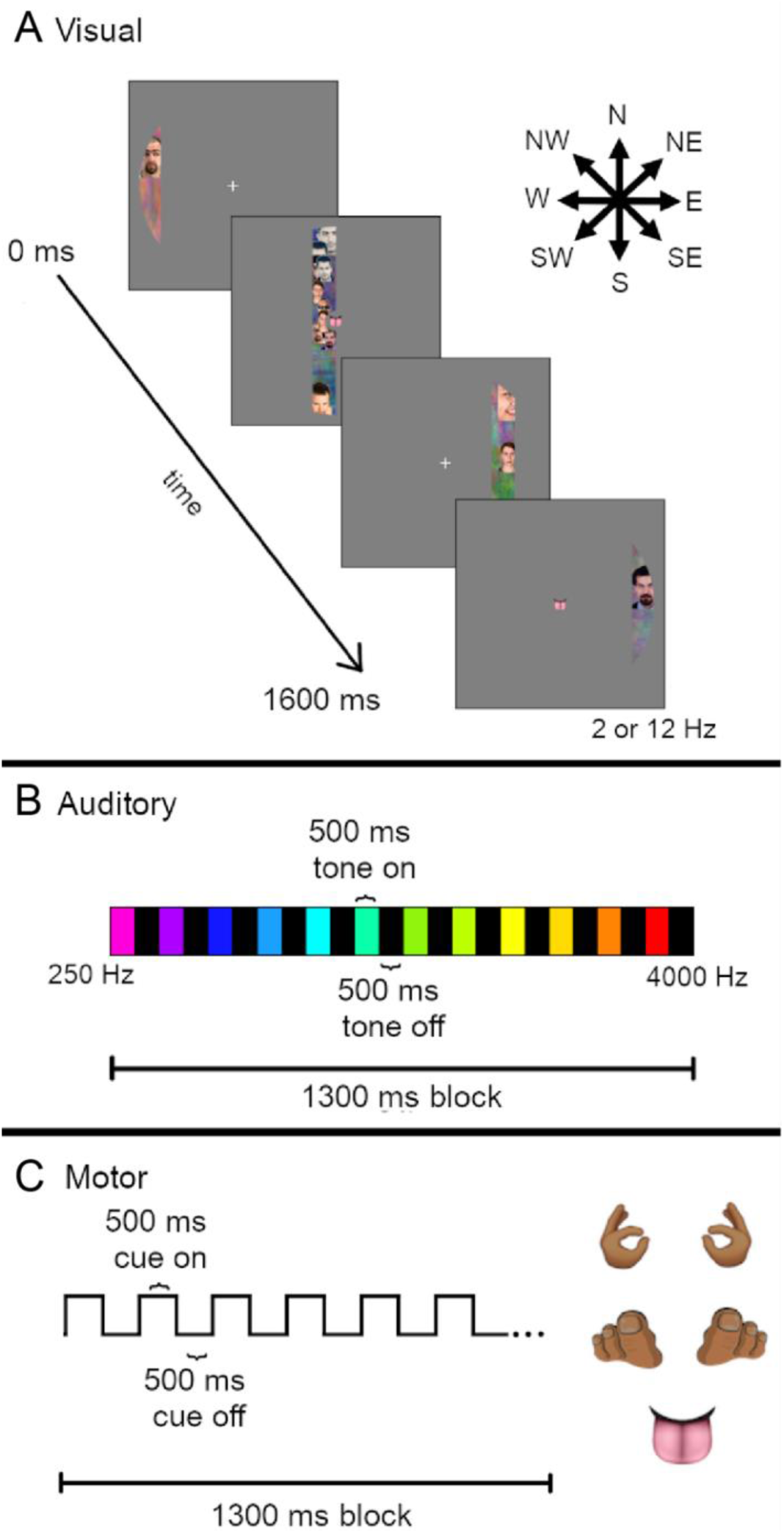
Population Receptive Field (pRF) mapping task. A) Visual stimuli (dynamic sweeping bars, flickering at 2 or 12 Hz temporal frequency) were presented which traveled in one of eight directions (arrows, top right). Sweep duration was 16 s. B) Auditory stimuli (pure tones, 250 - 4000 Hz) were presented in ascending or descending order during 13 s blocks. Tone duration was 500 ms with 500 ms of silence (scanner noise only) in between. C) Motor task cue stimuli were presented at the center of the screen (i.e., fixation point, see A). Five different body part images were presented: left and right hands, left and right toes, and tongue. The participants’ task was to tap the relevant body part in time with the cue presentation (500 ms on, 500 ms off, 13 s block duration).

During the scan, participants heard auditory tones that varied in pitch (13 tones between 250 - 4000 Hz; Figure 1B) for the purpose of tonotopic mapping (Allen et al., 2022; Norman-Haignere et al., 2013). Auditory tones were presented through the headphones in the presence of constant scanner sounds (i.e., no ‘gap’ in the scanning sequence). In an auditory block, each of the 13 tones were presented in either ascending (i.e., 250 Hz to 4000 Hz) or descending (i.e., 4000 Hz to 250 Hz) order for 500 ms with a 500 ms inter-stimulus intervals, such that a full auditory block was 13 s long. For each run of the pRF task, there were 8 auditory blocks in the first half of the scan (ascending tone order in run 1, descending in run 2) followed by 8 auditory blocks in the second half of the scan (descending in run 1, ascending in run 2). There was a 10 s silent period (scanner noise only) at the beginning and end of each run, and 96 s in between the ascending and descending tone sets.

Participants were asked to complete a motor tapping task during the pRF paradigm, to facilitate functional mapping of motor cortex (Lotze et al., 2000; Olman et al., 2012). In this task, they were asked to press the left or right buttons on the button box, curl the toes on their left or right feet, and tap their tongue to the roof of their mouth in time with the presentation of a flashing body part cue image presented at fixation (Figure 1C). Motor cues were presented for 500 ms followed by a 500 ms presentation of a fixation cross, with a motor block duration of 13 s. It was not an intentional aspect of the design that both the motor blocks and the auditory sweeps lasted 13 s. Originally, 12 tones were included in the auditory design; the sweeps were not intended to cycle at the same frequency as the motor task. In the process of optimizing the auditory stimulus, a 13th tone was generated and the fact that this created cycles with the same duration as blocks in the motor condition was unfortunately overlooked.

For most participants, two 324 s pRF mapping runs were completed. Because adequate maps may be extracted from the data from a single run, the second scan was omitted from the end of a scanning session if the participant was experiencing fatigue or discomfort (3 of 52 scanning sessions for controls had only one pRF scan, 10 of 40 for relatives, and 38 of 84 for PwPP).

Participants viewed the task before entering the scanner, to practice tapping in response to the motor cues. Once participants were settled in the scanner, a sound check was performed. The sound check consisted of playing the tones that would occur during the pRF task while an EPI scan was being acquired. After that scan participants were asked to indicate whether they could hear the tones over the noise of the scanner.

Example pRF results from individuals and a group of control participants are shown in the Results section. As this manuscript focuses on describing data acquisition methods and data quality, details of the data processing and analysis for the pRF experiment are relegated to the Supplemental Information.

##### Contrast surround suppression (CSS) task

For the CSS task, participants were asked to determine which of two briefly presented sinusoidal luminance gratings—presented to the left or right of a fixation mark—had higher contrast. Examples of the CSS stimuli are shown in Figure 2A.

**Figure 2.**
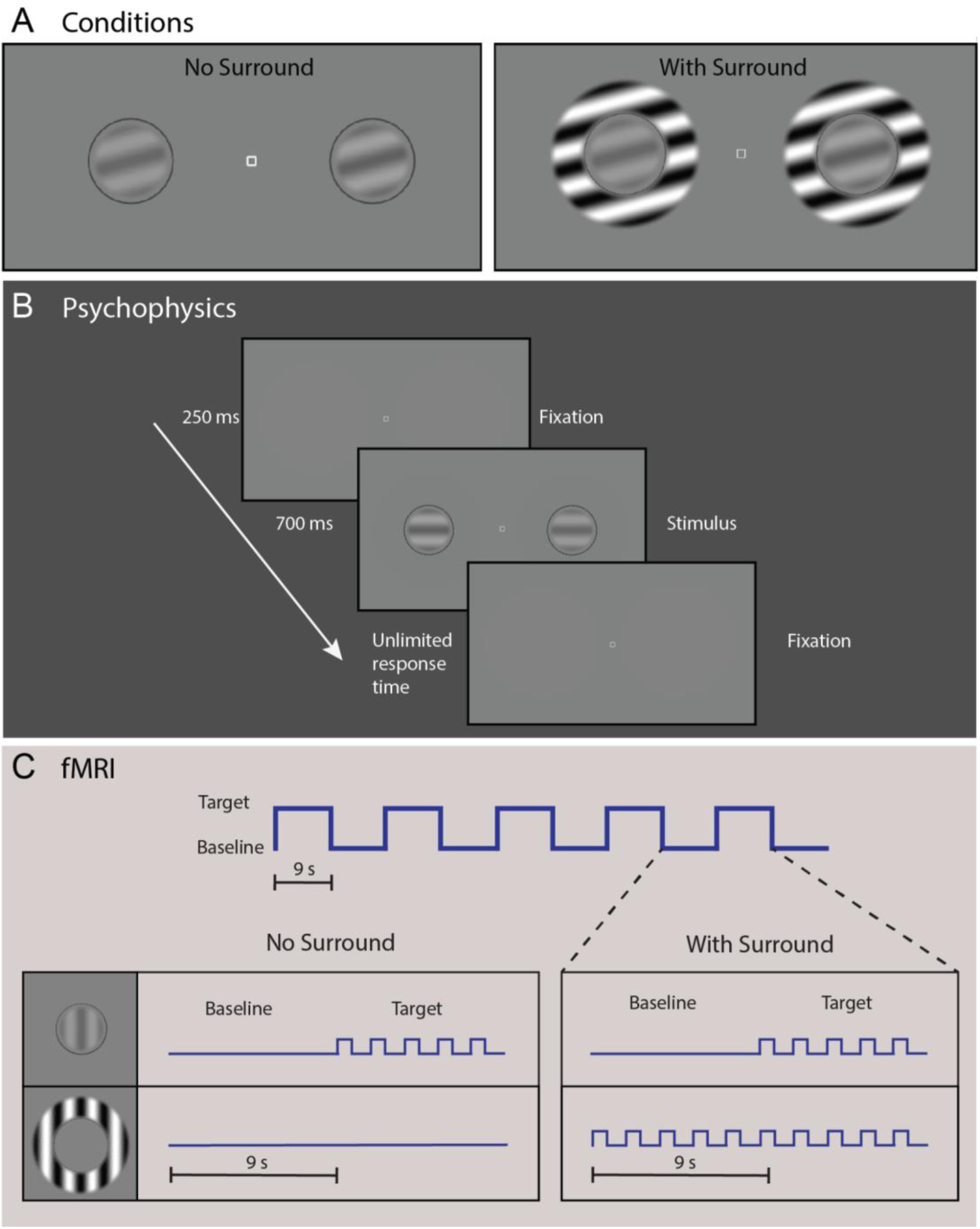
Contrast surround suppression (CSS) task. A) Circular grating stimuli were present alone (left) or inside surrounding annular gratings (right; to induce the surround suppression illusion). Panel B shows the sequence for a single trial in our CSS psychophysical experiment. Participants were instructed to fixate on the center square and report which grating appeared ’stronger,’ or higher contrast (left or right). In panel C, we illustrate the blocked experimental design for both the No Surround and With Surround conditions from our CSS fMRI task. Target and baseline stimuli were presented in 9 s blocks. Five stimulus presentations (trials) composed one block, with 5 pairs of target and baseline blocks composing one condition (90 s). Surrounding stimuli (0% or 100% contrast for No Surround and With Surround, respectively) were held constant across both baseline and target blocks within each condition.

In the psychophysical experiment, gratings were presented on a mean gray background at seven pedestal contrast levels: 0, 0.65, 1.25, 2.5, 5, 10, 20%. The range of pedestals was chosen based on previous studies (Boynton et al., 1999; Legge and Foley, 1980; Schallmo et al., 2016; Yu et al., 2003; Zenger-Landolt and Heeger, 2003) to allow investigation of threshold and suprathreshold contrast discrimination. Two gratings (2° diameter) were presented, each at 3° eccentricity to the left and right of a central fixation point (0.2° square; white with black outline) along the horizontal meridian. Gratings were contrast reversing at 4 Hz, and had a spatial frequency of 1.1 cycles/°.

Four target orientations were used: vertical, horizontal, and diagonal ± 45°. In some trials, the 10% contrast pedestal was presented with a surrounding sinusoidal grating annulus with inner and outer diameters of 2.5° and 4°, respectively (contrast = 100%; parallel to the target grating). The edges of the target and surrounding stimuli were blurred with a raised cosine function. A small fiducial circle (2.1° diameter with 0.05° gap between target grating and circle line; dark gray, luminance = 31.1 cd/m^2^) outside the target location was used to reduce spatial uncertainty about the target position. On each trial, a contrast increment was added to one of the two target gratings. The contrast increment was adjusted across trials within a range of 0.13 - 40%, according to a Psi adaptive staircase method (Kingdom and Prins, 2010) implemented in PsychoPy, to find the minimum contrast increment that could be perceived with 80% accuracy (i.e., contrast discrimination threshold for that pedestal contrast). For twelve early participants (part of the 7T-A protocol), we ran a pilot version of the task with slight differences (see Supplemental Information for more details about task differences; for the number of data sets collected for each version of the task, see Supplemental Table 2). Pilot task data are not included in the results presented below.

Participants were instructed to maintain fixation at the center of the screen while using their peripheral vision to determine which target grating was higher contrast. CSS task timing is illustrated in Figure 2B. Each trial began with an audio tone (250 ms) and presentation of the central fixation square and the two target gratings for 700 ms. Participants indicated their response on a keyboard using the left or right arrow key. The response period was not limited in duration, with a minimum inter-trial interval of 400 ms. The task was divided into eight blocks, one per pedestal contrast condition plus one block with the 10% pedestal plus the surrounding annulus. Each block included 90 trials from three separate interleaved staircases (30 trials / staircase), which yielded 3 independent threshold estimates per condition. Each block also contained 20 catch trials (not included in the staircases), in which the contrast increment was set to the maximum value (45%), in order to assess off-task performance. Each block lasted approximately 4 min, with self-timed rests between blocks. Participants were also instructed that they could take a brief pause within a block by withholding their response to the current trial. Task instructions and practice example trials were presented at the beginning of the psychophysical experiment. Total task duration (including instructions and practice) was approximately 40 min.

In the fMRI experiment, gratings were presented at five pedestal contrast levels: 0, 10, 20, 40, 80%. This range of pedestals was chosen based on previous fMRI studies (Zenger-Landolt and Heeger, 2003). Gratings were presented with the same size, positions, and spatial frequency as in psychophysical experiments. Grating orientation was randomized across stimulus presentations in a range of 0° to 180° in 15° increments. For each of the five pedestal contrasts, target gratings were presented either with or without a surrounding sinusoidal grating annulus with inner and outer diameters of 1.25° and 2°, respectively (100% contrast; same orientation as center).

CSS fMRI task timing is shown in Figure 2C. Each trial began with the presentation of the stimuli for 750 ms followed by a blank screen with a central fixation square presented for 1.05 s (1.8 s total trial duration). Trials were presented within blocks of 5 trials (9 s per block). Within each block, the target contrast pedestal was either 0, 10, 20, 40, or 80% for all 5 trials. Ten blocks (5 ‘target on’ with pedestal contrast at either 0, 10, 20, 40, or 80%, 5 ‘target off’ with pedestal contrast at 0%) comprised each condition. Within each condition, ‘target on’ and ‘target off’ blocks were presented in an alternating order, starting with a ‘target on’ block and ending with a ‘target off.’ This yielded an on-off block presentation order with 5 cycles per 90 s condition. Target pedestal contrast (10-80%) and surround contrast (0 or 100%) were held constant within each condition, for a total of 8 stimulus conditions in the main experiment. The experiment was divided into 3 fMRI runs, each 5 minutes long, with 3 conditions presented in each run. Each condition was presented only once across all runs, and the order of the 8 conditions in the main experiment was randomized across participants to one of 4 possible pseudo-random presentation sequences.

The first condition in the first fMRI run was always a functional localizer condition (also 90 s long, also divided into 10 blocks of 9 s each), which was designed to define regions within primary visual cortex representing the target stimuli. Here, we used a differential localizer technique (Olman et al., 2007), in which blocks of target gratings without surrounds (pedestal contrast = 80%, surround contrast = 0%) alternated with surround-only blocks (target pedestal contrast = 0%, surround contrast = 100%). This allowed us to isolate voxels in retinotopic early visual areas that responded more strongly to target stimuli than to surrounding gratings, as in previous work (Qiu et al., 2016; Schallmo et al., 2016; Schallmo et al., 2018; Schallmo et al., 2020). This first functional localizer condition was contiguous with the rest of the first CSS functional run (i.e., not a separate fMRI run).

In all of the CSS fMRI conditions, contrast increments were added to one of the two target gratings on each trial, as in our psychophysical experiment. This was done to equate task difficulty across conditions, and to keep the participants engaged and attending to the target stimuli. On each trial, participants indicated on which side the increment appeared using the left- or right-most button on a 4-button response box (Current Designs, Philadelphia, PA). The fixation square turned green upon correct responses. The response period (during the blank after each stimulus presentation) timed out after 600 ms. During our fMRI task, contrast increments were controlled by a 3-down, 1-up staircase using PsychoPy, with separate staircases for each of the 10 target pedestal (0, 10, 20, 40, or 80%) and surround (0 or 100%) combinations. There were 30 ‘target on’ and 30 ‘target off’ trials (on separate staircases) within each condition. The minimum contrast increment was 1%, and maximum contrast increment within each staircase was 25% of the target pedestal contrast, or 3% contrast, whichever was greater. At the beginning of the fMRI experiment, participants were briefly reminded of the task instructions, and were told that the task would not wait for them to respond, unlike during psychophysics.

Example CSS results from individuals and a group of controls are shown in the Results. Full details of the CSS data processing and analysis are provided in the Supplemental Information.

##### Contour object perception (COP) task

Visual stimuli in the COP experiment (Figure 3A-D) were based on those previously used by Silverstein and others (Silverstein et al., 2009; Silverstein et al., 2006; Silverstein et al., 2015; Silverstein et al., 2000). Stimuli consisted of a grid of 170 Gabor line elements, 14° visual angle wide by 11.3° tall. Gabor elements had a Gaussian envelope with *SD* = 0.067° and a spatial frequency of 5 cycles/°, with 2 cycles visible within each ∼0.4° wide (6 *SD*) Gabor. Of the 170 Gabor elements, 155 comprised the background, with a minimum spacing of 0.8° visual angle. The remaining 15 elements positioned around the center of the display formed an egg-shaped contour object that either pointed towards the left or right. This egg contour was 5.9° wide by 4.7° tall. Gabor elements that composed the egg were positioned with a minimum spacing of 1.09° and a maximum spacing of 1.13° relative to one another along the contour axis. One thousand exemplar stimulus grids were procedurally generated and saved for presentation during the task (chosen randomly). To manipulate the detectability of the contour stimuli, the relative orientation of each Gabor element within the contour was jittered with respect to the axis of the egg. This was done in steps of ± 3° between 0° (perfectly aligned) and 45° (completely scrambled). SNR, defined as the average spacing between adjacent background elements divided by the average spacing between adjacent contour object elements, was 0.87.

**Figure 3.**
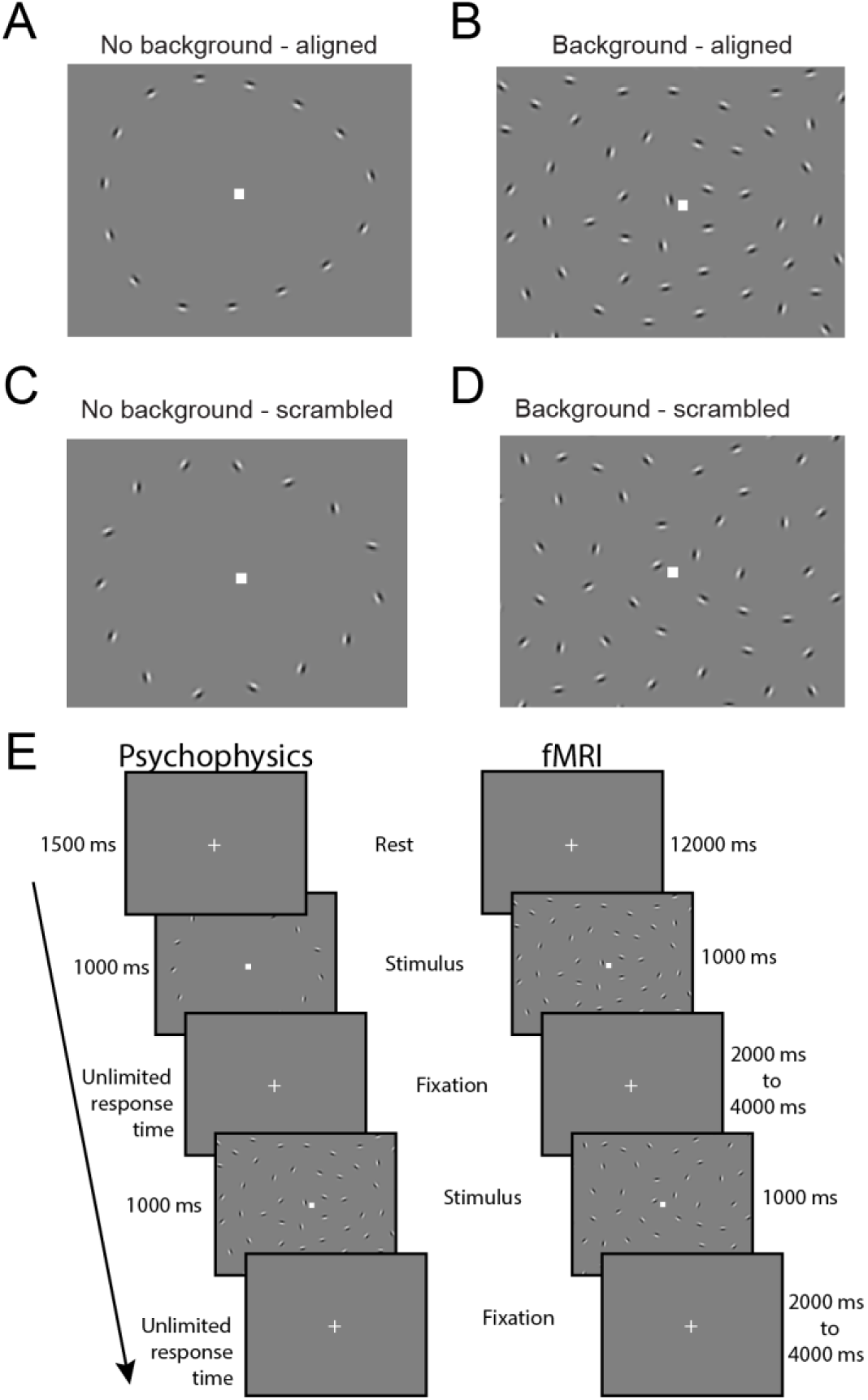
Contour object perception (COP) task. Panels A-D show example stimuli from 4 conditions: No background - aligned, Background - aligned, No background - scrambled, and Background - scrambled. Panel E (left column) shows the timing for a single trial from our COP psychophysical task. Participants were asked to report which direction an egg-shaped contour object pointed (left or right). In the right column, we show the timing for our COP fMRI paradigm, which included an initial 12 s rest period, followed by 1 s stimulus presentations with a jittered 2-4 s inter-stimulus interval.

In our COP psychophysical experiment, three different types of trials were presented with different stimuli: scrambled (45° jitter) contour stimuli presented without background elements (i.e., catch trials, used to assess off-task performance), aligned (0° jitter) contour stimuli presented within a field of background elements (to measure discrimination accuracy for fully aligned contours), and jittered contour stimuli presented with background elements. In this third condition, the alignment of the Gabor elements was manipulated across trials to identify the specific degree of orientation, or jitter threshold, at which the participant would discriminate directionality of the egg (left or right) with 70% accuracy. Contour jitter varied in increments of 3° from 0° to 45°. Contour jitter varied across trials based on a Psi adaptive staircase method (Kingdom and Prins, 2010), implemented within PsychoPy.

The COP psychophysical task was divided into two blocks, with each block consisting of three independent interleaved staircases of 30 trials of the jittered contour stimuli. Each block also included 20 additional trials of the scrambled contour stimuli with no background, and 20 of the aligned contour stimuli with a background. The experiment began with instructions asking participants to fixate on a square at the center of the screen and use their peripheral vision to decide whether the egg-shaped contour object was pointing towards the left or the right. They were told to make their best guess if they were unsure in which direction the egg was pointing. Blocks began with a central fixation square presented for 1500 ms, followed by presentation of the stimulus for 1000 ms, and an unlimited response period during which participants used the left and right arrow keys on a keyboard to provide an answer (Figure 3E). The order of trials was randomized within each block, and participants were allowed to take a self-timed rest between blocks. They were also told that they could take a short pause during the block if needed by withholding their response until they were ready to proceed. Each block lasted about five minutes, with a total task duration including instructions of approximately 15 minutes.

The fMRI version of the COP task was analogous to that used in our psychophysics experiment, with a few differences (Figure 3E). Four types of stimuli were presented, and these were divided into different conditions: jittered or fully scrambled contours, presented either with or without background elements. Scrambled contours with backgrounds were included in the fMRI task in order to measure responses to a field of Gabors in which the contour was very difficult (if not impossible) to perceive. For the jittered with background condition, contour stimuli were presented with their degree of orientation jitter controlled by a 3-up, 1-down adaptive staircase. Jitter level began at 0° at the start of the fMRI task run, and changed in increments of ± 3°. These staircases are expected to converge at the jitter level for which a participant could detect the directionality of the egg with 79% accuracy (Garcia-Perez, 1998). Contour jitter in the jittered without background condition was matched to the jittered with background condition across blocks. The contour stimuli in the scrambled conditions were completely scrambled (45° of jitter). Stimuli were presented in trials with a 1 s stimulus duration, followed by a randomized inter-stimulus interval of 2-4 s. In between each trial, a white fixation square was presented on a mean gray background with no background Gabors present. Trials were organized into two types of blocks (24 s long): those with and those without background elements. Each block included six trials; four jittered and two scrambled trials per block. The experiment began with a block of stimuli with background elements, and alternated between the two block types. Six blocks were administered per fMRI run. There was an additional 12 sec of rest before and after the 6 main experiment blocks in each run. A single fMRI task run lasted 6 or 9 min in total (see below), and each participant completed 2 fMRI runs within a single scanning session.

The instructions provided to the participant during the COP fMRI task were identical to those given during the psychophysics experiment, with the exception of being asked to provide a response as soon as possible after stimulus presentation.

The COP fMRI paradigm also included a functional localizer condition. Data from the functional localizer condition were acquired to permit identification of retinotopic regions of visual cortex that represented the spatial position of the egg stimuli. This condition consisted of repeated presentations of egg-shaped contours of Gabor elements, without background elements. In order to facilitate strong fMRI responses in early visual cortex, Gabor elements reversed contrast at a frequency of 2 Hz. Alternating blocks of rest (fixation only) and localizer stimuli were administered, each lasting 12 s. During the localizer stimulus blocks, the contrast reversing egg randomly changed directionality (left or right) every 2 s. Seven blocks of rest and 6 blocks of localizer stimuli were presented during the functional localizer condition, which lasted about three minutes. The functional localizer condition was presented at the beginning of the first COP fMRI run, prior to the first main experimental block, which meant that this first COP run was longer than the second (9 minutes in total, rather than 6).

Both the psychophysical and fMRI paradigms for the COP task were modified slightly after an initial piloting phase (about 6 months after the study’s onset, when the change from protocol version 7T-A to 7T-B was made). This was done to make the task easier, especially for PwPP, by making the spacing between background and contour Gabor elements wider (SNR was increased from 0.75 to 0.87), thereby making the contour elements easier to perceive. For full details, please see the Supplemental Methods, and Supplemental Table 2.

Example COP results from individuals and a group of control participants are shown in the Results. Full details of the data processing and analysis for the COP experiment are included in the Supplemental Information.

##### Structure-from-motion (SFM) task

We collected psychophysical behavioral data during a SFM task using the rotating cylinder illusion; SFM fMRI data were not acquired, due to time constraints. The visual stimuli in our SFM psychophysical task (Figure 4A) were standard rotating cylinders (Treue et al., 1991; Ullman, 1979). The rotating cylinder is a classic illusion in which small visual elements (here, black and white squares) move back and forth across a rectangular area in order to induce the perception of a 3-dimensional cylinder rotating in the depth plane. The rotating cylinder stimuli used here were composed of 400 small black and white squares (each 0.25° wide; 200 black and 200 white) that alternated between moving from the left to right and right to left across the width of a rectangular area, and each positioned pseudo-randomly along the height of the rectangular region (height = 10°, width = 7°). The squares sped up as they approached the center of the rectangle and slowed down as they approached each edge. This was done to simulate the perceived speed of the squares as if they were positioned on a transparent cylinder rotating in the depth plane (simulated rotation speed of 90°/s).

**Figure 4.**
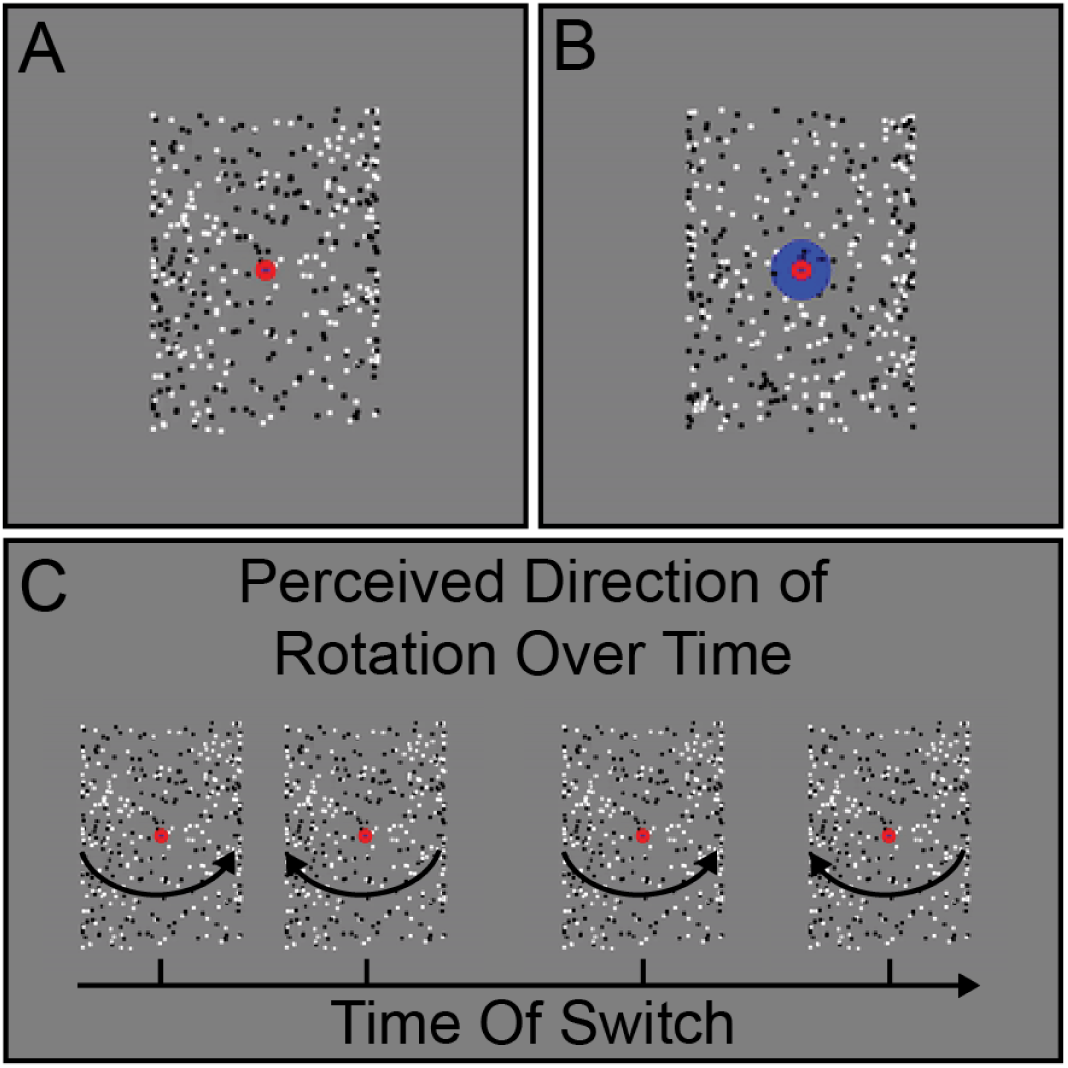
Structure-from-motion (SFM) task. Example stimuli for the real-switch task (A) and bi-stable task (B) conditions are shown. Panel C shows hypothetical examples of perceived switches in stimulus rotation direction across time during the bi-stable task.

Two versions of the stimuli with subtle but important differences were used for the two different task conditions, referred to as the bi-stable and real switch tasks. In both tasks the same black and white squares moved back and forth across the rectangular area with speed, size, and position being constant across the two tasks. In the bi-stable task, a small red fixation point (0.6° diameter) was positioned in front of all squares in the center of the rectangular area (Figure 4A). When dots collided in the bi-stable task, they occluded one another randomly, to remove this potential depth cue. In the absence of inherent depth cues, the direction of motion is ambiguous, and perception spontaneously alternates between the front surface rotating to the left and to the right. In the real switch task, we simulated physical switches of the rotation direction in depth by changing which set of dots (i.e., those rotating left to right or right to left; surface #1 and surface #2 of the cylinder) were presented in the front or back. To do so, we used the same red fixation dot overlaid on another larger blue circle (1.8° diameter) and alternated which set of dots passed in front of or behind each other and the larger fixation circle (Figure 4B). This provided an explicit depth cue thus biasing one percept (e.g., front rotating to the left) to become dominant. Switches occurred every 9-13 s (order and timing were pseudo-randomized, but fixed and identical for all participants). The real switch task was added to the experiment during the 7T-A data collection phase, about 2 months before the switch to the final 7T-B protocol. This permitted us to assess participants’ ability to perceive and respond to real switches in rotation direction. Thus, data are missing from a number of the early participants (7T-A) who did not complete this condition.

In both SFM task conditions, participants were asked to fixate on the small central red circle and told to use their peripheral vision to determine the direction of rotation of the front surface of the cylinder, either left or right (Figure 4C). Participants were instructed to respond using the left or right arrow keys to indicate which direction of rotation they perceived. Importantly, they were told to respond immediately to their initial percept at the beginning of the stimulus presentation, and then again each time their perception changed. Each participant first completed a short (30 s) practice version of the real switch task. Participants then ran one block of the real switch task followed by 5 blocks of the bi-stable task. Each block was 2 minutes long; the rotating cylinder was presented for the entirety of the block.

Example SFM results from individuals and a group of control participants are shown in the Results. Additional details of the SFM analysis are included in the Supplemental Information.

##### Eye tracking

Eye tracking data were acquired during our psychophysical experiments using an SR Research (Ottawa, Canada) Eyelink 1000 infrared eye tracker (1000 Hz sampling rate, binocular acquisition). The camera was mounted on the table in front of the participant, below the monitor. Nine point calibration and validation were performed prior to each psychophysical task, and drift correction was performed in between task blocks.

During fMRI, eye tracking data were acquired from a subset of participants using either an SR Research Eyelink 1000 (mounted at the back of the scanner bore), or an Avotec (Stuart, FL) Nano infrared eye tracker (mounted inside the Nova RF head coil). Information on the number of eye tracking data sets collected in each task across participant groups is presented in Supplemental Table 4. Note that due to logistical and safety issues during the COVID-19 pandemic, eye tracking data were not collected during fMRI experiments between March, 2020 and July, 2021.

#### Functional magnetic resonance imaging (fMRI)

FMRI data acquisition parameters followed the 7 T scanning protocol in the original Young Adult HCP (Glasser et al., 2016; Van Essen et al., 2013; Vu et al., 2017; Vu et al., 2015), as described below. Full details of our scanning protocols are included in the Supplemental Materials. Gradient echo (GE) fMRI data were acquired with TR = 1000 ms, TE = 22.2 ms, echo spacing = 0.64 ms, flip angle = 45°, resolution = 1.6 mm isotropic, partial Fourier = 7/8, 85 oblique-axial slices, field of view = 208 x 208 mm^2^, phase encode (PE) direction = anterior-posterior, multi-band acceleration factor = 5, parallel imaging acceleration factor = 2. A single GE scan (3 TRs) was acquired with identical parameters as above, but with an opposite PE direction (posterior-anterior), to facilitate geometric distortion compensation (Schallmo et al., 2021). A *T*_1_-weighted structural scan was acquired with TR = 3000 ms, TE = 3.27 ms, echo spacing = 8.1 ms, flip angle = 5°, resolution = 1 mm isotropic, partial Fourier = 6/8, 176 oblique-axial slices, field of view = 256 x 256 mm^2^, parallel imaging acceleration factor = 2. A *B*_0_ field map was acquired with TR = 642 ms, TEs = 4.08 & 5.1 ms, flip angle = 32°, resolution = 1.6 mm isotropic, partial Fourier = 6/8, 85 oblique-axial slices, and field of view = 208 x 208 mm^2^. A pair of spin echo (SE) scans (3 TRs each) with opposite PE directions (anterior-posterior and posterior-anterior) were acquired with TR = 3000 ms, TE = 60 ms, echo spacing = 0.64 ms, flip angles = 90° & 180°, resolution = 1.6 mm isotropic, partial Fourier = 7/8, 85 oblique-axial slices, field of view = 208 x 208 mm^2^, multi-band acceleration factor = 5, parallel imaging acceleration factor = 2. The imaging field-of-view positioning (yellow box in Figure 5A) was standardized using Siemens AutoAlign. An example sagittal image from a GE EPI scan in a single participant is shown in Figure 5B.

**Figure 5.**
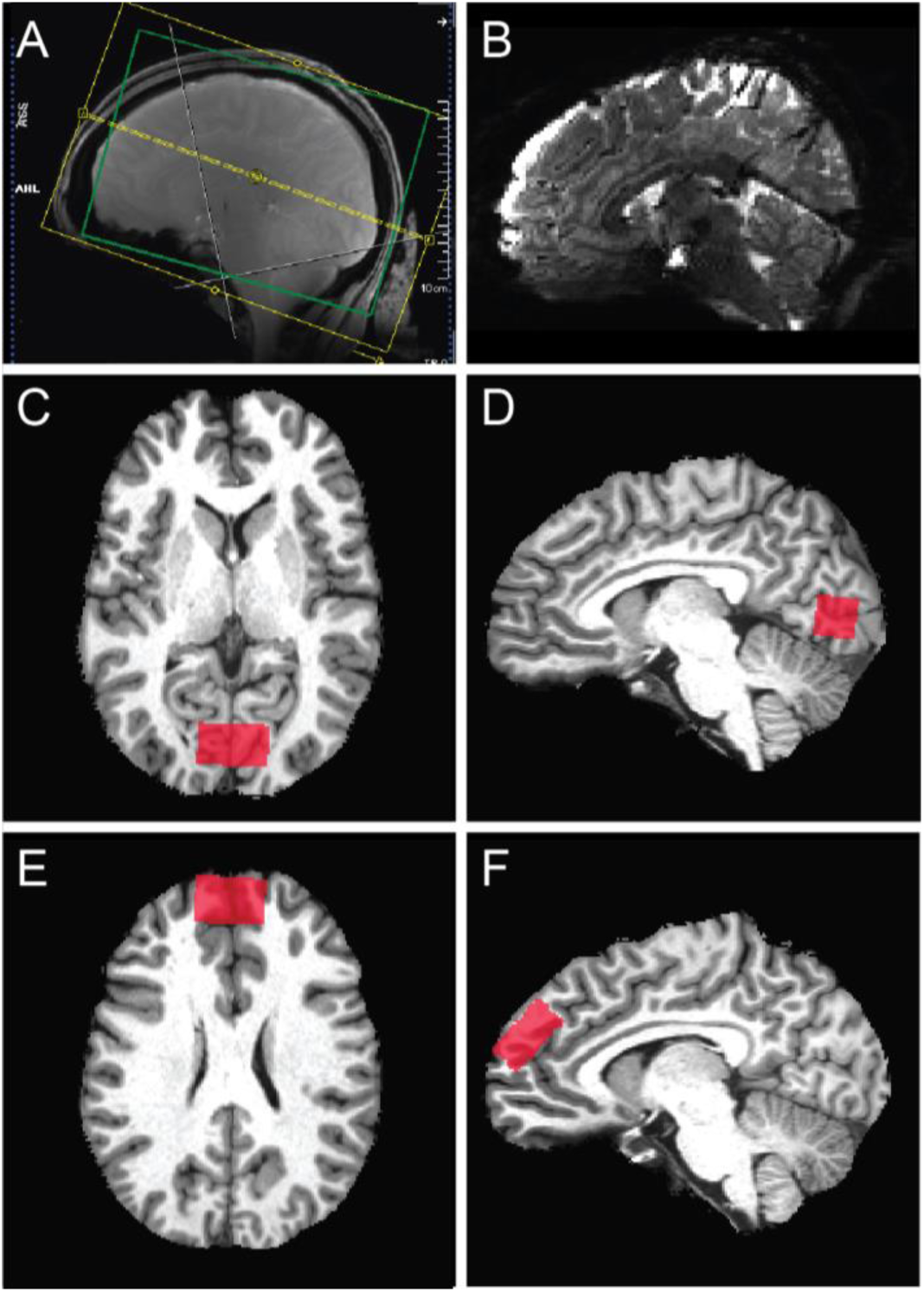
Functional MRI and MR spectroscopy acquisitions. A) Field of view (yellow) and adjustment volume (green) for fMRI. B) Example sagittal image from a GE EPI scan in a single participant. Panels C-F show example MRS volume placement (red) for occipital (C & and prefrontal (E & F) volumes-of-interest in axial (C & and sagittal (D & F) images from individual participants.

In order to reduce magnetic field inhomogeneity, *B*_0_ shimming was performed within a 130 x 170 x 120 mm^3^ oblique-axial region (i.e., adjustment volume; green box in Figure 5A), centered on the brain. We measured the linewidth of water in the Siemens Interactive Shim tab; for values > 80 Hz, shim currents were re-calculated to obtain a better shim solution. We used a local software program (*shimcache*) to apply the computed shim values before each scan to ensure there was no loss of *B*_0_ shim values during scanning.

#### Magnetic resonance spectroscopy (MRS)

##### Occipital cortex (OCC)

For our MRS experiments in OCC, we acquired data using a stimulated echo acquisition mode (STEAM) sequence (Marjańska et al., 2017) with the following parameters: TR = 5 s, TE = 8 ms, mixing time (TM) = 32 ms, hsinc pulse = 1.28 ms, volume-of-interest (VOI) size = 30 mm (left-right) x 18 mm (anterior-posterior) x 18 mm (inferior-superior), transmitter frequency = 3 ppm, 3D outer volume suppression interleaved with variable power and optimized relaxation delay (VAPOR) water suppression (Tkáč et al., 2001). Chemical shift displacement error was 4% per ppm. We acquired 2048 complex data points with a 6000 Hz spectral bandwidth. The OCC VOI was placed within the medial occipital lobe, superior to the cerebellar tentorium, posterior to the occipitoparietal junction, and anterior to the sagittal sinus (Figure 5C & D). For repeat (7T-Z) scans, VOIs were placed in the same anatomical position as in the first scan using a saved copy of the scan protocol along with AutoAlign. The full details of our scanning protocols are included in the Supplemental Materials.

Our acquisition protocol was as follows. We began by acquiring a *T*_1_-weighted anatomical scan to facilitate voxel placement, using the following parameters: TR = 2500 ms, TE = 3.2 ms, echo spacing = 7.9 ms, flip angle = 5°, resolution = 1.3 mm isotropic, partial Fourier = 6/8, 64 mid-sagittal slices, field of view = 160 x 160 mm^2^. After positioning the OCC voxel based on the individual participant’s occipital anatomy, we performed shimming within the VOI using FAST(EST) MAP (Gruetter, 1993; Gruetter and Tkáč, 2000) to obtain a linewidth of water ≤ 15 Hz (measured using Spectroscopy card in Siemens software), repositioning the voxel slightly as necessary. The *B*_1_ field for the 90° pulse and the water suppression flip angles were calibrated by adjusting the transmit voltage for each VOI in each participant. We then conducted a brief STEAM scan (4 TRs) to review the quality of water suppression. Next, we acquired our primary metabolite STEAM spectra (96 shots or TRs; 8 min), from which metabolite concentrations were quantified. Finally, three additional reference scans were acquired without water suppression to permit eddy current correction and absolute quantification of metabolite concentrations relative to water. The first was the same as the previous STEAM scan, but with the transmitter frequency = 4.7 ppm, and without VAPOR water suppression (1 TR). The second was the same as the first, but included 4 TRs to permit phase cycling. The third was the same as the first (1 TR), but did not include outer volume suppression.

##### Prefrontal cortex (PFC)

We acquired STEAM data within PFC using the same protocol as for OCC, except that the VOI size = 30 mm (left-right) by 30 mm (anterior-posterior) by 15 mm (inferior-superior). The PFC voxel was oriented obliquely within the sagittal plane, and placed within the dorsomedial prefrontal cortex, dorsal and anterior to the cingulate gyrus (Figure 5E & F). PFC data acquisition did not begin until January, 2018, when the frontal MRS coil became available; MRS data sets acquired prior to this included OCC data only. For an initial group of 30 participants with PFC data, we acquired 128 shots (TRs) of metabolite STEAM data. After ensuring that data quality was comparable between PFC and OCC, we reduced this number to 96 shots for subsequent participants, in order to shorten the scan time and reduce participant burden. OCC data were always acquired prior to PFC data within a scanning session. Participants were removed from the scanner in between OCC and PFC scans, in order to switch from posterior to anterior transceiver coils. The entire MRS experiment duration was approximately 80 min (40 min each for OCC and PFC). MRS data were always acquired prior to fMRI scanning. Participants were removed from the scanner between MRS and fMRI scans and given a short (∼30 min) break, during which we prepared the scanner environment for the fMRI experiments.

### Data processing

We provide a summary of our own internal fMRI and MRS data processing pipelines in the Supplemental Information. This processing enabled us to assess data quality and the general pattern of results, as presented in the Results section.

### Data quality assessment

#### Psychophysical & fMRI data quality

We assessed multiple retrospective quality metrics for the psychophysical and fMRI data from our visual tasks. For the CSS and COP psychophysical tasks, poor task engagement was defined as achieving less than 80% (CSS) or 85% accuracy (COP) across all catch trials. For the SFM psychophysical experiment, poor task engagement was defined based on performance in the real switch task as having fewer than 7 correct responses (< 63.6% accuracy) made within 4 seconds of a physical stimulus change.

For fMRI data quality, we first defined excessive head motion during a given task as having ≥ 0.5 mm of motion across > 20% of TR pairs. This was quantified using AFNI’s *gen_ss_review_scripts.py*, based on the Euclidean norm of the six head motion parameters from AFNI’s *3dvolreg*. We also quantified the fraction of stimulus presentations in each fMRI task for which a behavioral response was recorded (either correct or incorrect, assuming a response was required), as a measure of task engagement. For the pRF fMRI task, we defined poor task engagement as having made no button press responses (at all) during > 10% of left- and right-hand finger tapping blocks in the motor tapping task. During the CSS and the COP fMRI tasks, we defined poor task engagement as not responding to > 10% of all stimulus presentations.

#### MRS data quality

We examined a number of metrics to assess the quality of our MRS data. First, we measured the linewidth of the unsuppressed water signal (in Hz), which provides a measure of the shim quality (i.e., *B*_0_ homogeneity) within the selected VOI. Poorer shim quality will reduce the fidelity with which different peaks can be resolved in the spectrum. Specifically, we quantified the linewidth of the unsuppressed water signal (FWHM in Hz) within the MRS VOI during the scanning session by fitting a mixed Gaussian-Lorentzian function in the Spectroscopy tab on the Siemens console, as described above, to a phased water signal. This linewidth value was assessed prospectively (i.e., prior to MRS data acquisition), and manually recorded by the scanner operators. We set an *a priori* threshold for poor shim quality as a water linewidth of > 15 Hz. When linewidth values exceeded this limit, shimming was performed again to obtain a better shim solution. Head motion during MRS data acquisition was also examined prospectively using a Metria motion tracking system, as noted above. A very small minority of MRS data sets (n = 3) were acquired with shim values > 15 Hz (e.g., cases in which better shim solutions could not be found). Additionally, the spectrum linewidth and SNR as quantified using LCModel were also used as retrospective MRS data quality metrics. LCModel defines SNR as the maximum signal (*N*-acetylaspartate peak) minus baseline divided by twice the root mean square of the residuals. We set data quality thresholds for these two metrics based on *post hoc* inspection of the data. Poor data quality was defined as > 5 Hz spectrum linewidth, or SNR < 40.

### Data analysis

For this report, we preprocessed all datasets to compute data quality metrics and compare across groups. We present behavioral, fMRI, and spectroscopy results for controls in order to show representative results, rather than an exhaustive analysis. Analyses were performed in MATLAB (version 2016a) unless otherwise noted.

Details of our analysis methods are provided in the Supplemental Information.

### Code and data availability

Our experimental task code and our data processing code are available from GitHub (github.com/mpschallmo/PsychosisHCP). Unprocessed (i.e., DICOM) imaging data and associated behavioral data files will be available from the Human Connectome Project (db.humanconnectome.org; 3 T data released in February, 2022; 7 T data release planned for December, 2022). Note that the publicly available data from each scanning session are in native (i.e., scanner) space. Details of our procedures for integrating data across scanning sessions are provided in the Supplemental Information. Clinical and other (non-imaging associated) behavioral data, as well as notes for each scanning session, will be available from the National Data Archive (nda.nih.gov/edit_collection.html?id=3162). Processed data will be made available by the investigators upon request.

## Results

In order to characterize visual perceptual functioning in PwPP, we acquired neurophysiological data from a group of 43 healthy controls, 44 biological relatives, and 66 PwPP. The dataset includes visual psychophysical (i.e., behavioral task) data and 7 T functional MRI data, using tasks focused on basic aspects of visual perception (e.g., retinotopy, context processing, object perception). We also acquired 7 T MR spectroscopy data in the occipital and frontal lobes, in order to characterize the concentrations of different metabolites in these same participants. A subset of PwPP and controls were recruited to return for an identical repeat session a few months after their initial experimental session (see Table 1 for information about the time between first and repeat sessions). Table 2 summarizes the number of unique participants and repeat sessions for each experiment. Below, we provide a description of data quality and example results from each of the various experiments.

**Table 2.**
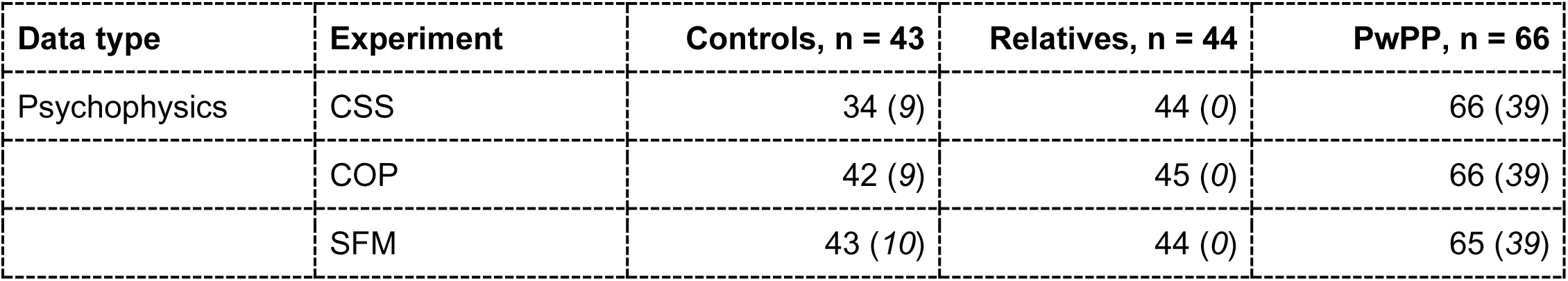
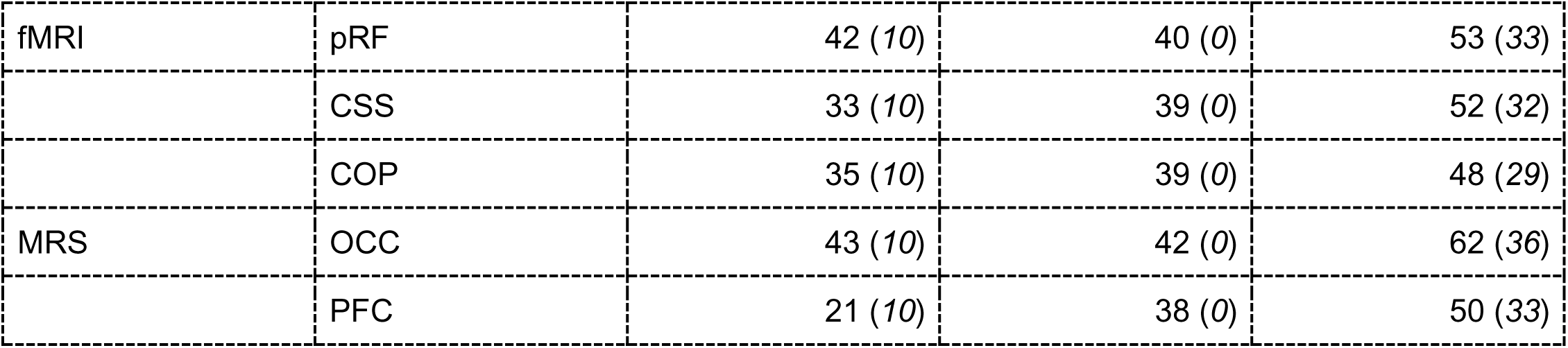
Number of data sets collected for each experiment. Values are reported as: unique individuals (*repeated data sets*). CSS = contrast surround suppression, COP = contour object perception, SFM = structure from motion, pRF = population receptive field, fMRI = functional magnetic resonance imaging, MRS = magnetic resonance spectroscopy, OCC = occipital cortex, PFC = prefrontal cortex

### Psychophysical & fMRI results

We defined a set of data quality metrics for our psychophysical and fMRI task experiments based on *a priori* thresholds for poor task performance. These included excessive head motion or failing to respond during fMRI tasks, and low accuracy during psychophysical catch trials. In general, the data we collected were of good quality as measured by these metrics, with 81% of our > 800 psychophysical and fMRI data sets passing quality control checks (Figure 6; green bar). Statistical comparisons of these quality metrics are provided in Supplemental Table 5. Summaries of data quality metrics for each participant group in each experiment are presented in Supplemental Figure 3 (psychophysics) & Supplemental Figure 4 (fMRI). Data quality and example results for each of the experiments are described below. In Supplemental Figure 5 we provide additional information regarding our fMRI data (the average head motion per TR, as well as temporal outliers per TR, across both groups and experiments). A chart detailing data quality for each experimental session in each participant is provided in Supplemental Figure 6.

**Figure 6.**
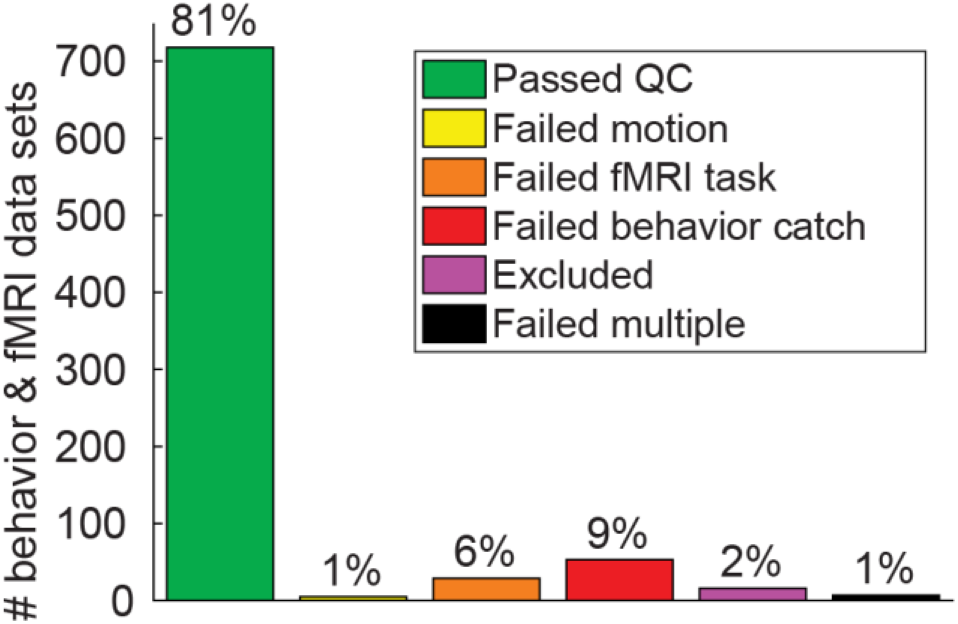
Behavioral and fMRI data quality. Chart shows the number and percentage of behavioral (psychophysics) and fMRI data sets that passed or failed various data quality checks across 4 experiments (pRF, CSS, COP, & SFM).

#### pRF results

Our population receptive field (pRF) mapping fMRI experiment used sweeping bars as visual stimuli to permit retinotopic mapping. Figure 7A shows polar angle (left) and eccentricity (right) maps from one individual (top) as well as a group of n = 49 control participants (bottom). Visual stimuli were presented at two different temporal frequencies (2 and 12 Hz), which permitted functional examination of temporal frequency selectivity across visual cortex (Figure 7B). Our pRF paradigm also included auditory tone sweep sequences (250 - 4000 Hz), to enable tonotopic mapping in auditory cortical regions (Figure 7C). Participants completed a motor tapping task during the pRF experiment, with 5 body part images presented at fixation (left and right thumbs, left and right toes, and tongue; see Methods). This permitted the functional identification of somatomotor cortical regions that were responsive during the tapping task (Figure 7D). Data quality in our pRF experiment, as assessed by head motion and behavioral response rate, was generally high (Supplemental Figure 4, left column), but differed significantly across groups (Supplemental Table 5), and was lowest among PwPP.

**Figure 7.**
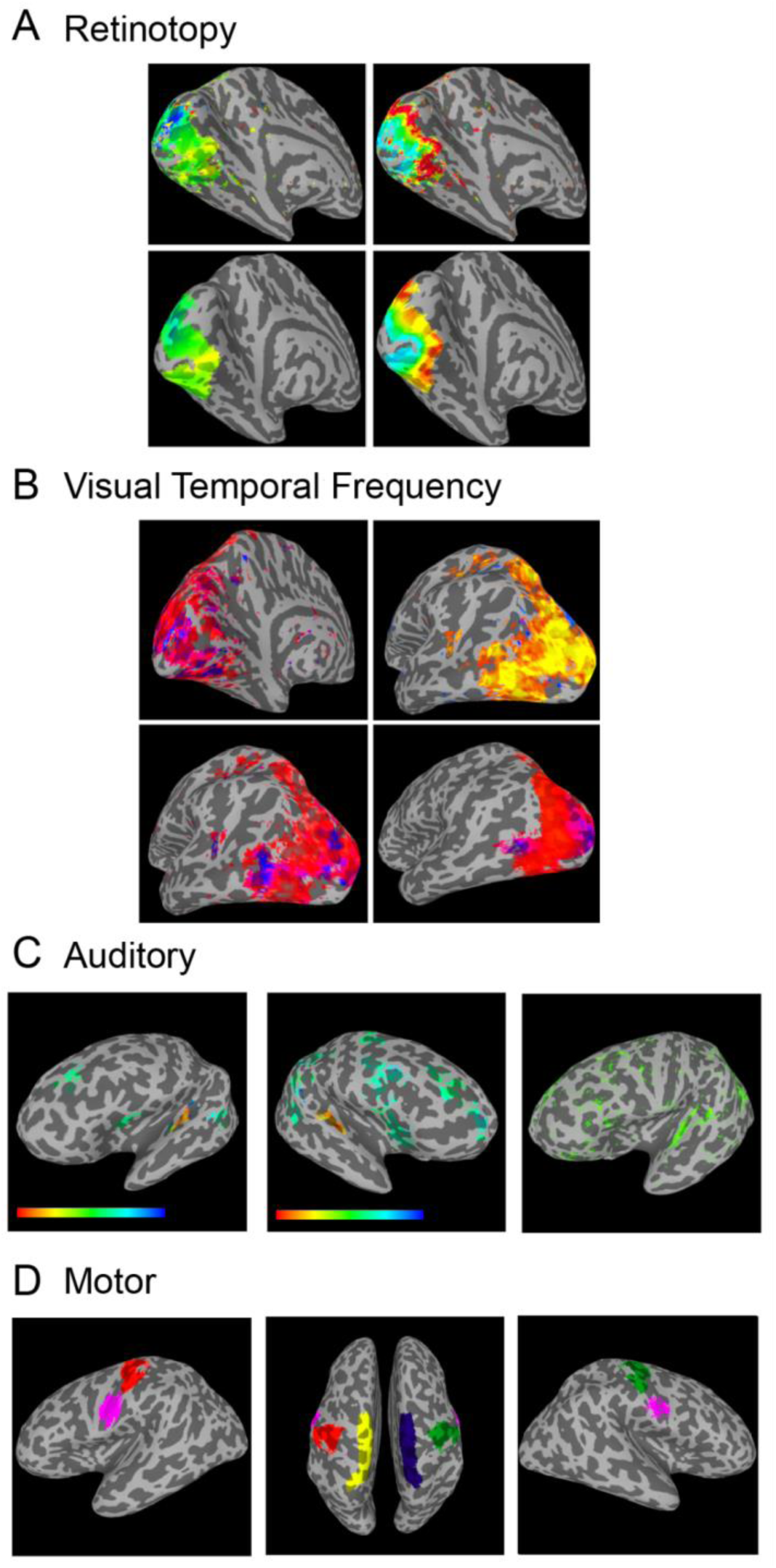
Results from the pRF fMRI task. A) Retinotopic fMRI results from the sweeping bars paradigm. Polar angle maps are shown from a single participant (top left), as well as from a group of N = 49 controls (bottom left). Eccentricity maps from a single participant (top right) and the same group of controls (bottom right) are also shown. Colors indicate retinotopic selectivity (polar angle: blue = lower vertical meridian, green = right horizontal meridian, yellow = upper vertical meridian; eccentricity: blue = fovea, red = periphery). B) Responses to visual stimuli presented at 2 vs. 12 Hz were compared, to examine temporal frequency selectivity. Top right image shows all active regions from a single participant, top left and bottom left show 2 Hz - 12 Hz contrast from the same participant (medial and lateral views, respectively; red = selectivity for 2 Hz, blue = selectivity for 12 Hz). Bottom right image shows 2 Hz - 12 Hz contrast from a group of N = 49 controls. Note that human MT complex (hMT+) can be seen in the middle and right panels as a region selective for high temporal frequency stimuli (blue) in the lateral occipital lobe. C) Tonotopic fMRI responses to the auditory sweep stimuli. Left and middle images show individual tonotopic responses in left and right hemispheres from a single participant, right image shows data from a group of N = 49 controls. Color bar indicates voxel peak auditory frequency selectivity (red = 250 Hz, blue = 4000 Hz), from a Fourier analysis. Note that spurious regions of activation outside of auditory cortex are present (e.g., central sulcus) due to the accidental confound of having 13 s blocks for both motor and auditory tasks. D) Regions showing selective fMRI responses during the motor tapping task from a group of N = 49 controls. Different colors indicate selectivity for different body parts (left and right thumbs = green & red, left and right toes = blue and yellow, tongue = pink). The relative position of different body part ROIs follows the expected… (continued on next page) ’homunculus’ pattern in the region of the central sulcus. ROIs were defined using a contrast between the body part of interest and all other body parts. Voxels are included in the group level ROI if they were included in the individual level ROI for > 50% of participants.

#### CSS results

Our contrast surround suppression (CSS) experiment permitted us to examine two visual phenomena: contrast-response functions and surround suppression. To this end, we acquired both psychophysical and 7 T fMRI data during a CSS task. Example CSS psychometric data are shown in Figure 8A, which illustrates how contrast discrimination thresholds were defined from our psychophysical data. Threshold versus contrast data from control participants (n = 32) are shown in Figure 8B, and illustrate the expected ‘dipper’ shape (open symbols); thresholds decrease slightly between pedestal contrasts of 0% (detection) and 0.6%, before increasing again at higher pedestal values. Further, there is a clear surround suppression effect in the 10% contrast pedestal data (the only with-surround pedestal condition we examined in the behavioral dataset): contrast discrimination thresholds are higher with versus without surrounding stimuli (Figure 8B; filled vs. open symbols), indicating surround suppression of contrast perception.

**Figure 8.**
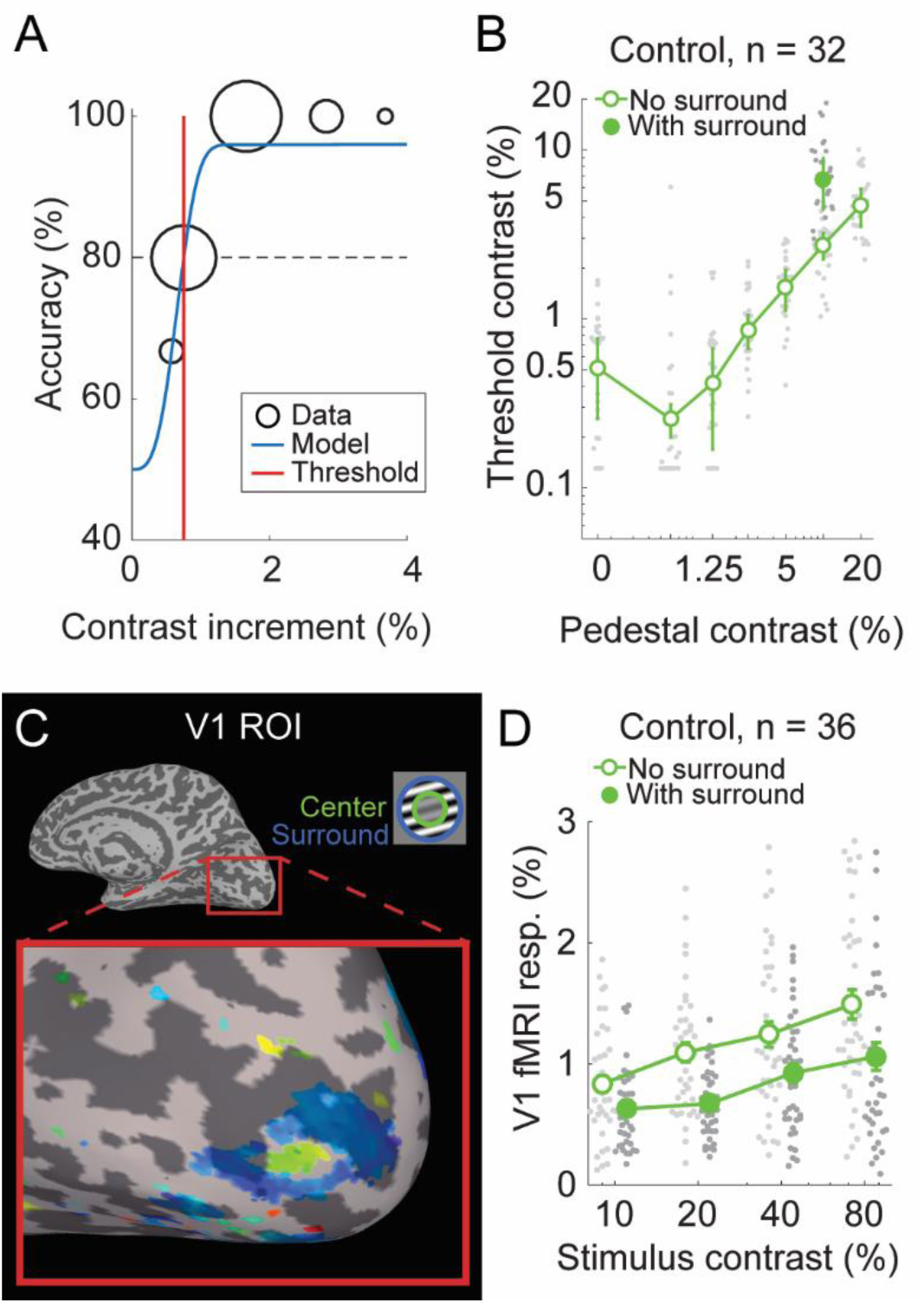
CSS task results. A) Example psychometric data from a single pedestal contrast condition in one participant. Data (black) were fit with a Weibull function (blue) to obtain a discrimination threshold (red) at which stimulus contrast could be perceived with 80% accuracy (dashed line). B) Group threshold-versus-contrast data for N = 32 control participants in 8 different stimulus conditions (7 pedestal contrasts [x-axis], plus 10% pedestal with surround [filled circle]). These data show the expected ’dipper’ function, with thresholds in the 0.6% pedestal condition being lower than for detection (0% pedestal). Gray dots show individual data points, green dots show group geometric means, error bars show median absolute deviation. C) Functional localizer data from the CSS fMRI task in a single participant. A region of primary visual cortex is highlighted, which shows spatial selectivity for the center stimulus region (green) versus the surround (blue). Color indicates the phase of the fMRI response from a Fourier analysis (Engel, 1997). D) CSS fMRI responses from V1 center-selective ROIs in the no surround (open, offset left) and with surround (closed, offset right) conditions across 4 pedestal contrasts, in a group of N = 36 controls. Gray dots show individual data points, green dots show group means, error bars show S.E.M.

An example ROI in primary visual cortex (V1) from the CSS fMRI task in a single participant is shown in Figure 8C. The differential (i.e., center vs. surround) localizer produced the expected retinotopic activation pattern on the V1 cortical surface, with voxels that respond selectively to the center stimuli (green) surrounded by voxels responding to the surround (blue). 7 T fMRI responses in V1 from controls (n = 36) are shown in Figure 8D. These data show the expected increase in V1 fMRI responses with increasing stimulus contrast, as well as the expected surround suppression effect; V1 fMRI responses from center-selective ROIs are lower with versus without surrounding stimuli (Figure 8D; filled versus open symbols). Data quality in the CSS task, as assessed by psychophysical catch trials, fMRI head motion, and fMRI behavioral task responses, was generally high (Supplemental Figure 3 & Supplemental Figure 4), but was significantly different across groups (Supplemental Table 5), and lowest among PwPP.

#### COP results

We acquired psychophysical (outside the scanner) and 7 T fMRI data during a contour object perception (COP) task. The degree of collinearity (i.e., orientation jitter) among Gabor contour elements and the presence or absence of irrelevant background elements were manipulated in order to examine contour integration and figure-ground segmentation (see Methods). Figure 9A shows an example psychometric function from the COP task, which illustrates how decreasing contour jitter was associated with higher shape discrimination accuracy, as well as how jitter thresholds (70% accuracy) were quantified. Threshold data from a group of n = 33 control participants are shown in Figure 9B. Figure 9C shows an example ROI identified using the COP localizer data in right V1 from a single participant. As expected, the COP localizer yields a stripe of activation across V1 (orange), roughly perpendicular to the calcarine sulcus. In Figure 9D, we show an example of 7 T fMRI responses from V1 in n = 30 controls across our 4 COP task conditions. This illustrates that the addition of background stimuli yielded higher fMRI responses (as would be expected with a larger number of stimuli on the screen), whereas contour jitter had little effect on the V1 fMRI response amplitude. We saw high data quality overall in our COP experiments (Supplemental Figure 3 & Supplemental Figure 4), as assessed by psychophysical catch trials, fMRI head motion, and fMRI behavioral task responses. Of these, only head motion differed significantly across groups (Supplemental Table 5) and was highest (worst) among PwPP.

**Figure 9.**
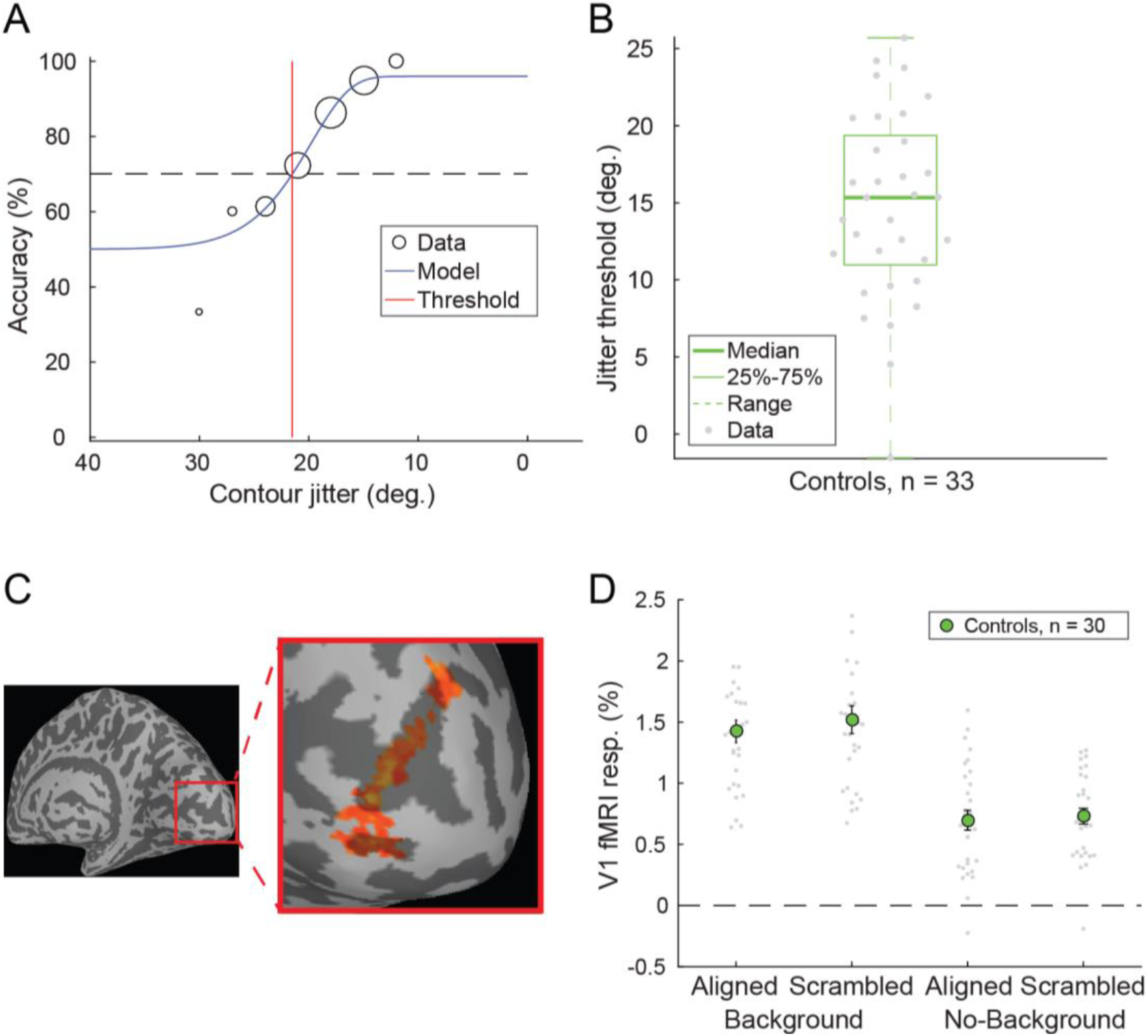
COP task results. A) Example COP psychophysical data (black) from a single participant were fit with a Weibull function (blue) to obtain a jitter threshold (red), which reflecting 70% contrast discrimination accuracy (dashed line). B) Jitter thresholds for N = 33 controls. Thick line shows median, box shows 25-75%, whiskers show 1.5 x interquartile range, gray dots show individual data points. C) COP functional localizer data from area V1 in the right hemisphere from an example participant. Color indicates statistical significance from a Fourier analysis (Engel, 1997) showing voxels selective for the COP localizer stimulus > blank. D) COP fMRI responses from retinotopic contour ROIs in area V1 across 4 stimulus conditions in a group of N = 30 controls. The presence of background stimuli increased the fMRI response (as expected, with more stimuli on the screen), whereas we saw little effect of contour alignment in the V1 fMRI response.

#### SFM results

We obtained psychophysical (but not fMRI) data from a structure-from-motion (SFM) task using the bi-stable rotating cylinder illusion. This task provided us with a behavioral measure of the stability of visual motion and form integration during the perception of a bi-stable illusion. Participants reported the perceived direction of rotation (clockwise or counterclockwise in depth) for an array of dots without explicit depth cues. This bi-stable stimulus yielded spontaneous alternations in the perceived direction of rotation. Example behavioral responses time courses from n = 32 healthy control participants are shown in Figure 10A. In Figure 10B, we show the average switch rates within the same group. On average, percept direction alternated about every 10 s, but there was substantial variability in bi-stable switch rates between individuals, as expected. Participants also completed a ‘real switch’ task as an experimental control condition, in which explicit depth cues were used to define physical changes in the rotation direction of the stimulus. A fair number of participants showed poor behavioral performance on the real switch task, which we interpret as difficulty in understanding and performing the task as instructed. Participants with poor performance were excluded from our analyses (see Methods). Performance in the SFM real switch task did not differ significantly across groups (Supplemental Table 5).

**Figure 10.**
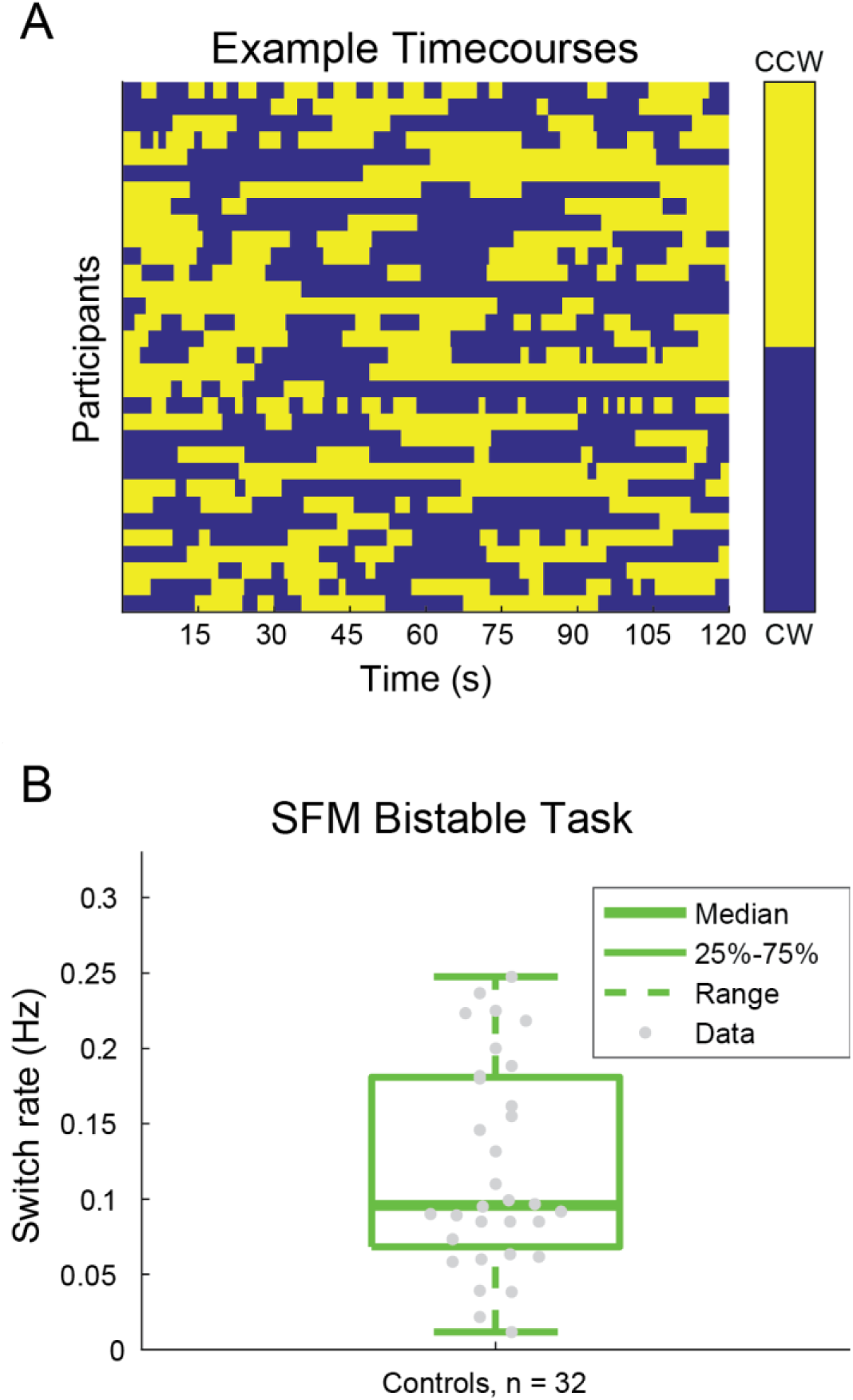
SFM results. A) Example behavioral response time courses from the bi-stable task, showing perceived switches between clockwise (CW, blue) and counterclockwise (CCW, yellow) rotation in depth. Data from a single 120 s block are shown for N = 32 control participants. B) Bi-stable switch rates from the same group of participants. Thick line shows group median, box shows 25-75%, whiskers show 1.5 x interquartile range, dots show individual participants.

### MRS results

To assess data quality in our MRS experiments, we defined a set of quality metrics, both *a priori* (linewidth of the unsuppressed water peak) and *post hoc* (spectrum linewidth and SNR). We observed high quality overall in our MRS data, with 92% of all of our > 300 MRS data sets passing all quality thresholds (Figure 11; green bar). MRS data quality metrics for each group and VOI are shown in Supplemental Figure 7, which indicates that data quality is generally comparable across groups and VOIs (Supplemental Table 6 shows statistical comparisons between groups for MRS quality metrics). A detailed summary of data quality for each VOI from each MRS scanning session in all participants is shown in Supplemental Figure 8.

**Figure 11.**
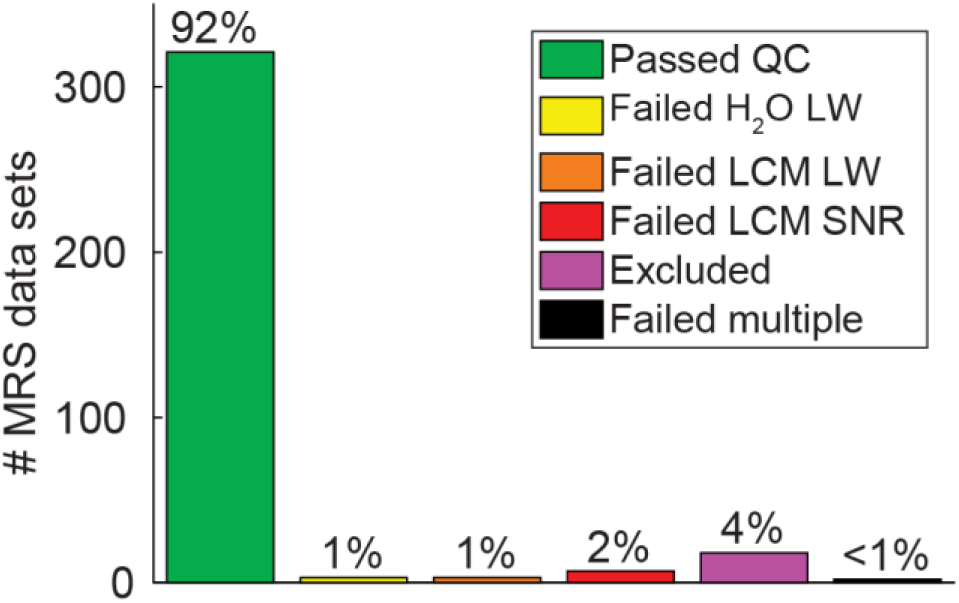
MRS data quality. Chart shows the number and percentage of MRS data sets that passed or failed various data quality checks across 2 VOIs (OCC and PFC). H2O LW = water linewidth, LCM LW = LCModel linewidth, LCM SNR = LCModel signal-to-noise ratio.

Example MR spectra from individual participants in both OCC and PFC VOIs are shown in Figure 12A & C, respectively. Here we show example spectra (black) fit by LCModel (red), and the residual error after fitting, to illustrate the fit quality. Example voxel placement, and the proportion of gray matter, white matter, and CSF within each voxel are also illustrated. We fit a combination of 18 different metabolites using LCModel; examples of individual fits for glutamate and GABA are shown at the bottom of Figure 12A & C.

**Figure 12.**
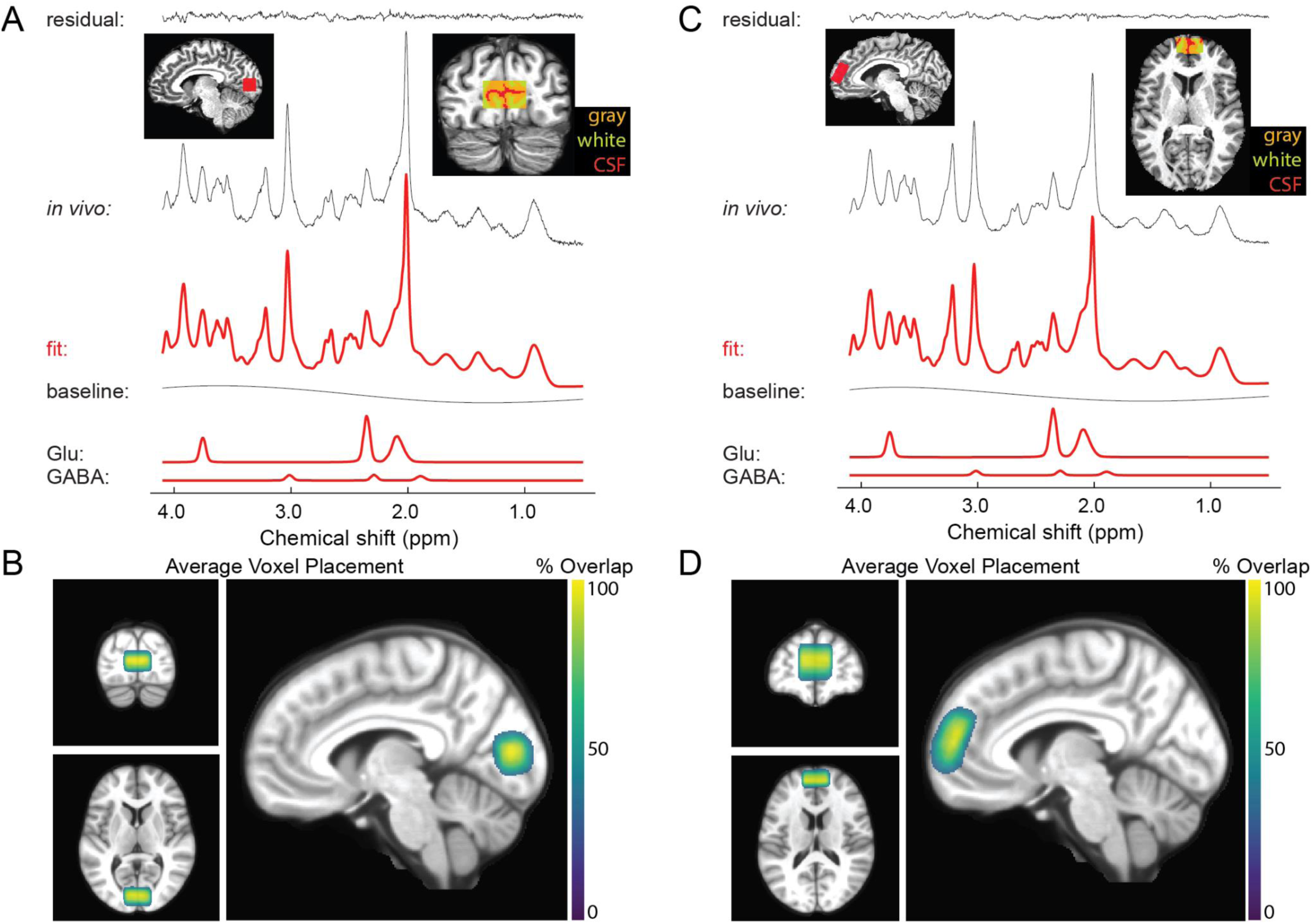
Example MRS spectra and average VOI positions. A) OCC spectrum from a single participant (black) fitted with LCModel (red). Top left inset shows a sagittal view of OCC VOI placement. Top right inset shows a coronal view of the OCC VOI, with gray matter (orange), white matter (yellow), and cerebrospinal fluid (CSF, red) highlighted. Bottom rows show glutamate (Glu) and GABA components, as fitted by LCModel. B) Average OCC VOI placement across N = 193 data sets (including repeats). Color shows overlap across participants. VOI masks were transformed from individual to MNI space, and thresholded at 30% overlap. C and D show the same as A & B, but for the PFC VOI (N = 147 data sets in D).

To visualize the consistency of voxel placement across participants, we transformed voxel masks for each VOI in each scanning session into MNI canonical space, and then computed the percent overlap in space across participants and sessions. This is shown for both OCC and PFC voxels in Figure 12B & D, respectively, and indicates a fairly high degree of consistency in voxel placement across individuals.

Finally, in Figure 13 we show the quantification of glutamate (A) and GABA (B) in both OCC and PFC voxels across controls (n = 42 in OCC, 30 in PFC; fewer PFC data sets were collected due to a delay in hardware availability, as noted in the Methods). These plots illustrate that metabolite concentrations, scaled relative to water, are within the expected range for our healthy adult population, as measured by this MRS technique (Marjańska et al., 2017). We note that previous studies have also observed numerically higher glutamate levels in prefrontal as compared to occipital cortex (Marsman et al., 2014; Zhang and Shen, 2015), though such differences were not statistically significant.

**Figure 13.**
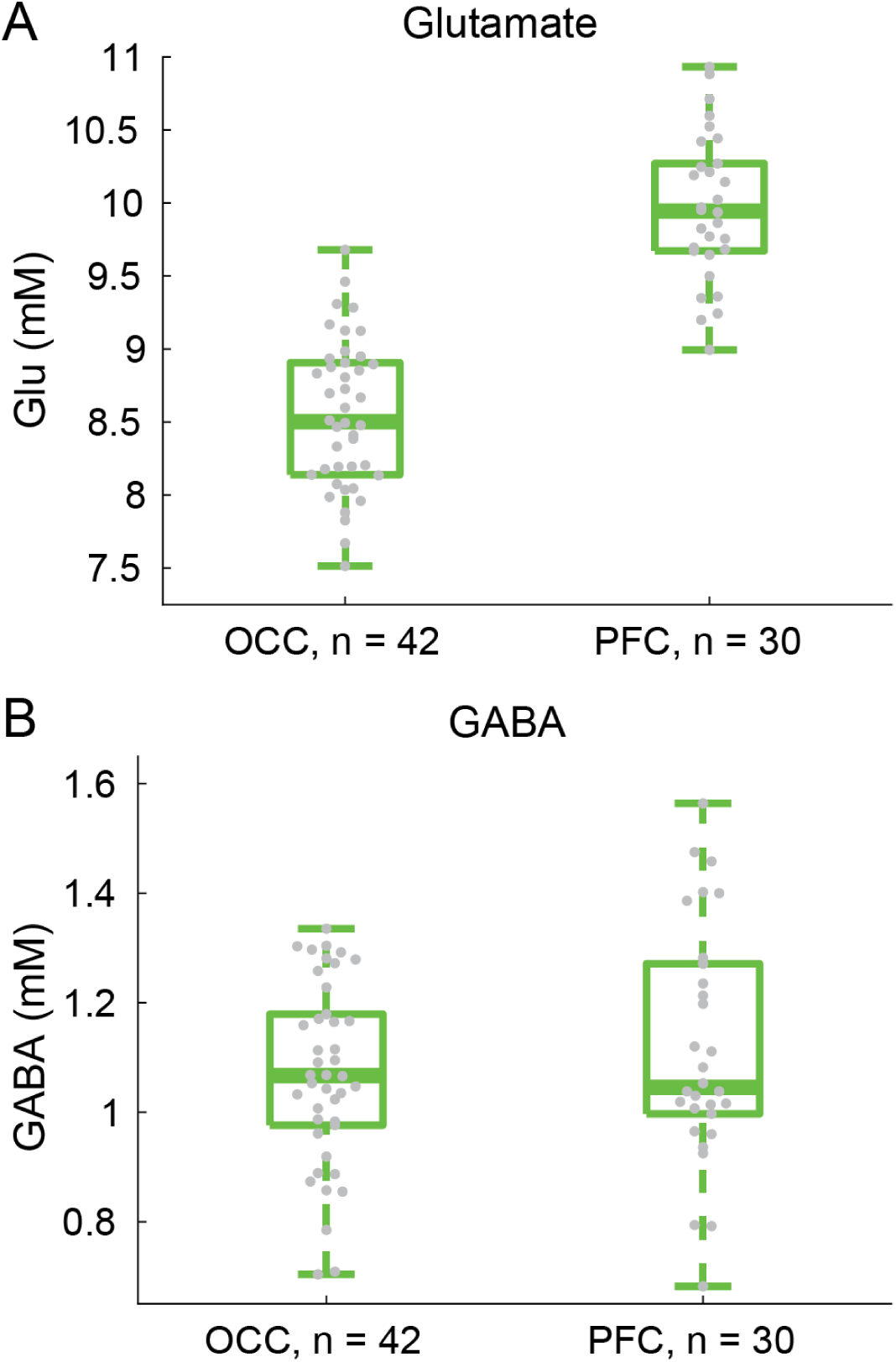
MRS results showing (A) glutamate and (B) GABA concentrations in OCC and PFC, from a group of N = 42 & 30 participants, respectively. Fewer PFC data sets were collected due to a delay in hardware availability, as noted in the Methods. Thick lines show group medians, boxes show 25-75%, whiskers show 1.5 x interquartile range, gray dots show data points from individual participants.

## Discussion

This report of the data collected as part of our Psychosis HCP project is intended to facilitate the use of this dataset by the research community. We are pleased that, overall, this project yielded a substantial number of data sets, and relatively few were hampered by the expected confounds of head motion, fatigue, difficulty complying with task instructions, or equipment malfunction. For this report, we have included results of analyses that extract a small number behavioral or physiological parameters from a subset of control participants, in order to provide a sense of effect sizes and measurement error in the dataset. No results from PwPP or relatives are included in this report; more thorough analyses of these datasets will be included in future publications focused on the findings from group comparisons. The primary goal of this report is to document methods and promote the availability of this dataset.

This dataset is one of many available through db.humanconnectome.org as part of the Human Connectome Project. In addition to the 7 T visual pRF data acquired as part of the original Young Adult HCP (Benson et al., 2018), other HCP studies have focused on the structure and function of the visual system in conditions such as macular degeneration, low vision, blindness, and sight restoration. Further HCP data that are relevant to the current study are available from other projects focused on topics including early psychosis and the genetic basis of mental disorders (e.g., Amish Connectome).

We have reported here only on experiments performed during visits for 7 T scanning sessions. Our 3 T dataset has been described in a previous publication (Demro et al., 2021). All participants at 7 T also participated in 3 T scanning sessions, and although we have not detailed the potential approaches in this report, it is entirely feasible to design analyses that integrate 3 T resting state or cortical parcellation derived from 3 T data with functional responses or neurotransmitter concentrations measured at 7 T. Thus, we hope that this multimodal dataset, once fully released in December 2022, provides the basis for many multimodal analyses by diverse research groups.

Because symptoms vary over time for PwPP, and performance on visual tasks such as contour object perception has been found to correlate with symptom severity (Keane et al., 2018; Silverstein et al., 2000), a subset of PwPP participating in this study were scanned on 2 occasions, separated by at least 1 month. Although not all PwPP returned for a second visit, we successfully obtained 39 pairs of repeat datasets in this group. The BPRS, SANS, and SAPS were collected at both 7 T visits, to provide a snapshot of clinical symptomatology that can be analyzed in conjunction with visual task performance, fMRI responses, and prefrontal or occipital metabolite concentrations. The goal of this aspect of the project is to provide data that may help elucidate the neural changes underlying shifts in symptom levels and visual task performance.

Naturally, acquiring a dataset this size is not without challenges, particularly when the project coincides with a worldwide viral pandemic. We have provided detailed information in the Methods and Supplemental Information describing which datasets are complete and incomplete, and which experimental protocols were changed over time. The authors are more than happy to consult with potential users of this dataset to help them navigate the details of exactly which data are available for which subsets of participants and experiments.

In sum, this dataset offers novel opportunities to investigate specific neurophysiological responses during visual experiments designed to study particular neural mechanisms (e.g., contour facilitation, contrast surround suppression) among PwPP, their relatives, and healthy controls, both cross-sectionally and across time. Our data may also facilitate investigating the role of neurochemical functioning (via MRS) in these groups. Researchers may also wish to explore relationships with structural and resting state functional connectivity measures acquired in the same participants at 3 T (Demro et al., 2021). For each usage case, approximately 150 - 200 high-quality datasets (including repeated scans) are publicly available to support future analyses. We know of very few other 7 T fMRI or MRS data sets that are currently publicly available, and even fewer that are designed to investigate visual functioning (Allen et al., 2022; Benson et al., 2018; Sengupta et al., 2017) and / or psychotic psychopathology.

## Conclusion

We show that it is feasible to collect a diverse array of high quality brain imaging data in visual cortex at ultra-high field (7 tesla) from a relatively large sample of healthy controls (N = 43), unaffected relatives (N = 44), and PwPP (N = 66) at a single research site. We demonstrate how cutting-edge multimodal imaging and behavioral methods can be applied to investigate visual neurophysiology in PwPP. By applying visual neuroscience methods along with standardized imaging methods from the Human Connectome Project (Benson et al., 2018; Glasser et al., 2016; Van Essen et al., 2013), our datasets offer new opportunities to investigate the role(s) of structural and functional connectivity in abnormal visual processing among PwPP. Hypotheses of neural dysfunction in PwPP that might be examined include a disruption in the balance of excitation and inhibition (Foss-Feig et al., 2017; Lewis et al., 2005; Lisman, 2012; Moghaddam and Javitt, 2012), thalamo-cortical dysconnectivity (Anticevic et al., 2014; Cheng et al., 2015; Damaraju et al., 2014; Dong et al., 2019; Giraldo-Chica and Woodward, 2017; Ramsay, 2019; Ramsay et al., 2017), abnormal visual gain control (Butler et al., 2008; Phillips and Silverstein, 2013), impaired top-down attentional modulation (Gold et al., 2018; Luck et al., 2019a; Luck et al., 2019b), and disrupted predictive coding (Adams et al., 2013; Horga and Abi-Dargham, 2019; Sterzer et al., 2018). By making our data and code publicly available, we hope to facilitate new investigations of visual processing in PwPP, and invite interested researchers to reach out to us for collaboration and support.

## Supporting information

FMRI Protocol

MRS Protocol

## Data Availability

Unprocessed (i.e., DICOM) imaging data and associated behavioral data files will be available from the Human Connectome Project (db.humanconnectome.org; 3 T data released in February, 2022; 7 T data release planned for December, 2022). Clinical and other (non-imaging associated) behavioral data, as well as notes for each scanning session, will be available from the National Data Archive (nda.nih.gov/edit_collection.html?id=3162).

http://db.humanconnectome.org/

https://nda.nih.gov/edit_collection.html?id=3162

## Acknowledgments

We thank Jesslyn (Li Shen) Chong, Victoria Espensen-Sturges, and Marisa J. Sanchez for their assistance with data collection and processing. Timothy J. Lano also contributed to data collection and project management. We thank Kamar S. Abdullahi, Phillip C. Burton, and Bryon A. Mueller for help with data processing. We also thank Stephen A. Engel and Essa Yacoub for supporting the design of the study, and Essa Yacoub for comments on the manuscript. Finally, we are grateful to Pamela D. Butler, Steven C. Dakin, Brian P. Keane, Damien J. Mannion, Steven M. Silverstein, and Duje Tadin for their input on the experimental design.

This work was supported by funding from the National Institutes of Health (U01 MH108150). Salary support for MPS was provided in part by K01 MH120278, and salary support for CAO was provided in part by R01 MH111447. Support for MR scanning at the University of Minnesota Center for Magnetic Resonance Research was provided by P41 EB015894 and P30 NS076408. This work used tools from the University of Minnesota Clinical and Translational Science Institute that were supported by UL1 TR002494.

## Supplemental information

### Supplemental background

One of the most fundamental functions in the visual system is contrast perception, which is impaired in psychosis spectrum disorders including schizophrenia and bipolar disorder (Butler et al., 2007; Butler et al., 2005; Calderone et al., 2013; Fernandes et al., 2019; Keri et al., 2002; Lalor et al., 2012; Martinez et al., 2008; Skottun and Skoyles, 2007; Slaghuis and Bishop, 2001; Yoon et al., 2009). Visual contrast is the difference in luminance between adjacent pixels or image regions. Electrophysiological studies of early visual cortex in animal models show that neurons have nonlinear contrast-response functions (i.e., the relationship between input, or visual stimulus contrast, and output, or neural response), with response saturation occurring at high contrast (Albrecht and Hamilton, 1982; Sclar et al., 1990). This is often referred to as contrast gain control and is thought to reflect a balance between local excitatory and inhibitory processes (Butler et al., 2008; Carandini and Heeger, 2012; Katzner et al., 2011). Similar nonlinearities are observed in human contrast perception (Boynton et al., 1999; Legge and Foley, 1980; Yu et al., 2003). Studies in humans have used psychophysical and functional MRI methods to link performance in visual contrast perception tasks to the magnitude of neural responses in early visual areas such as primary visual cortex (V1; Boynton et al., 1999; Olman et al., 2004; Zenger-Landolt and Heeger, 2003). With regard to psychotic disorders, early work indicated that impaired contrast perception might reflect a specific magnocellular deficit (Butler et al., 2007; Butler et al., 2005; Keri et al., 2002; Martinez et al., 2008), whereas more recent studies have suggested that contrast perception may be impaired more generally (i.e., for both magnocellular and parvocellular pathways; Calderone et al., 2013; Lalor et al., 2012; Skottun and Skoyles, 2007). A few studies have applied neuroimaging tools to investigate impaired contrast perception in PwPP, with some evidence suggesting reduced neural responses in early visual cortex (Butler et al., 2007; Calderone et al., 2013; Lalor et al., 2012; Martinez et al., 2008).

Another visual function that appears disrupted among PwPP is visual context perception. Spatial context phenomena refer to the modulation (either enhancement or suppression) of the perception of a centralized visual target by surrounding stimuli. For example, surround suppression is a visual effect (illusion) in which the perceived salience of a center stimulus (e.g., the contrast of a sinusoidal grating) is reduced in the presence of a surrounding stimulus (e.g., a high contrast annular grating; Cai et al., 2008; Chubb et al., 1989; Petrov and McKee, 2006; Schallmo and Murray, 2016; Snowden and Hammett, 1998; Xing and Heeger, 2000, 2001; Yu et al., 2001; Yu et al., 2003). This perceptual suppression corresponds with suppressed neural activity (as measured by fMRI in humans) in early visual areas such as V1 (Chen, 2014; Joo et al., 2012; Nurminen et al., 2009; Pihlaja et al., 2008; Poltoratski et al., 2017; Schallmo et al., 2016; Self et al., 2016; Vanegas et al., 2015; Williams et al., 2003; Zenger-Landolt and Heeger, 2003), consistent with electrophysiological studies in animal models showing suppression of neural responses to stimuli inside the classical receptive field by surrounding stimuli (Angelucci and Bressloff, 2006; Bair et al., 2003; Cavanaugh et al., 2002; Shushruth et al., 2013; Walker et al., 1999; Webb et al., 2005). Studies of surround suppression in people with psychotic disorders have generally shown weaker suppression effects (i.e., reduced illusion strength, or more veridical perception), especially among people with schizophrenia (Barch et al., 2012; Dakin et al., 2005; Schallmo et al., 2015; Serrano-Pedraza et al., 2014; Tadin et al., 2006; Tibber et al., 2013; Yang et al., 2013b; Yoon et al., 2009), and to perhaps a lesser extent among people with bipolar disorder (Schallmo et al., 2015; Yang et al., 2013a); for a meta-analysis, see (Linares et al., 2020). Relatively few studies have examined the physiological basis of reduced surround suppression in psychotic psychopathology; those few have suggested there might be impaired inhibition by GABA (Yoon et al., 2010) and / or reduced neural suppression (Anderson et al., 2017; Seymour et al., 2013) in early visual cortex.

Aspects of perceptual organization, including visual contour integration and perception of more complex forms / objects, also appear to be disrupted among PwPP. Contour perception involves detecting visual edges or boundaries, which is a critical step for distinguishing visual objects from background stimuli (i.e., figure-ground segmentation), and is important for navigating a visual environment (Loffler, 2008). Contour detection is often studied using tasks that require participants to integrate spatially separated elements; studies in healthy adults generally show that contour perception follows the Gestalt principles of proximity and good continuation (Loffler, 2008; Wertheimer, 1938). Perception of visual forms or objects involves the integration of one or more visual edges or contours, which facilitates perception of an isolated and / or closed visual shape. Processing of visual contours and shapes is reflected in neural responses in human V1 and in higher visual areas such as the lateral occipital complex (LOC) as measured by fMRI (Altmann et al., 2003; Keane et al., 2021; Murray et al., 2002; Qiu et al., 2016), in agreement with electrophysiological studies in animal models (Bauer and Heinze, 2002; Li et al., 2006; Li et al., 2008). Behavioral studies have repeatedly demonstrated impaired detection and discrimination of fragmented visual contours among people with schizophrenia (Grove et al., 2018; Keane et al., 2016; Pokorny et al., 2021b; Robol et al., 2013; Schallmo et al., 2013; Silverstein et al., 2006; Silverstein et al., 2012; Silverstein et al., 2000), as well as impaired perception of illusory contours (e.g., Kanizsa figures; Keane et al., 2014; Keane et al., 2018). A few fMRI studies have linked impaired contour integration in schizophrenia to abnormal neural processing in mid-level visual areas such as LOC (Silverstein et al., 2009; Silverstein et al., 2015). There is also evidence for impaired perception of more complex visual objects or forms among PwPP, including fragmented objects (Pokorny et al., 2021a), Mooney faces (Rivolta et al., 2014; Uhlhaas et al., 2006), and global motion percepts (Bennett et al., 2016; Carter et al., 2017; Chen et al., 2005). However, our understanding of the neural basis of impaired perceptual organization among PwPP remains somewhat limited.

It has been proposed that abnormal visual perception among PwPP may depend on an imbalance between excitation / inhibition (E/I) in brain regions related to visual perception (Foss-Feig et al., 2017; Lisman, 2012). E/I functions have been investigated behaviorally among healthy adults using bi-stable visual paradigms, such as binocular rivalry or the rotating cylinder illusion (Mentch et al., 2019; Robertson et al., 2016; van Loon et al., 2013). Bi-stable paradigms involve presenting a single visual stimulus with two competing perceptual interpretations, with the participant’s perceptual experience spontaneously alternating between the two. Abnormal bi-stable perception has been reported among PwPP as compared with controls (Fox, 1965; Miller et al., 2003; Nagamine et al., 2009; Ngo et al., 2011; Schmack et al., 2017; Schmack et al., 2015; Xiao et al., 2018; Ye et al., 2019), which may suggest an E/I imbalance in visual cortex.

### Supplemental methods

#### Task versions

Between July and December 2017, an initial group of 27 participants (20 controls, 7 PwPP) completed our initial experimental protocol, which we refer to as 7T-A. Following examination of the data from this initial pilot phase, modifications were made to the CSS (psychophysics) and COP (psychophysics and fMRI) paradigms, which are detailed below. The number of data sets collected from each version of these tasks is described in Supplemental Table 2.

For the CSS experiment, as part of the transition to 7T-B we added a 0.65% contrast pedestal condition (with no surround) to the psychophysical task to better capture the expected ‘dipper’ phenomenon in the threshold versus contrast data (i.e., thresholds at very low pedestal contrasts that are lower than the detection threshold; see Figure 8B). We also increased the maximum value of the contrast increment from 20% to 40%, in order to better address cases in which participants struggled to discriminate stimuli with relatively large contrast increments. As the contrast increment during catch trials was fixed at the maximum value, this change also increased the increment on catch trials from 20% to 40%.

For the COP experiment, we reduced the background clutter as part of the change to the 7T-B protocol, in order to make the task easier. To this end, we decreased the number of background Gabor elements (from 207 to 155) and increased their minimum spacing (from 0.7° to 0.8°). This had the effect of increasing the SNR for the contour stimuli (i.e., background versus contour element spacing) from 0.75 to 0.87.

#### Data processing

For both fMRI and MRS, T_1_-weighted structural MRI data acquired in a separate scanning session at 3T were used as an anatomical reference scan (i.e., for co-registration, below). Anatomical data (T_1_- and T_2_-weighted scans acquired at 3T) were processed using the HCP minimal preprocessing pipeline (version 3.22.0; Glasser et al., 2013), which includes gradient nonlinearity correction via *gradunwarp*. Both T_1_ and T_2_ data were used to generate white matter and pial surface models using FreeSurfer (version 5.3.0; Fischl, 2012). We then removed non-brain regions from the T_1_ data using the *3dSkullStrip* command from AFNI (version 18.2.04; Cox, 1996), followed by correcting for inhomogeneities in the intensity profile of gray matter and white matter voxels using AFNI’s *3dUnifize*.

#### FMRI processing

Our fMRI data processing pipeline is summarized in Supplemental Figure 1, and began with a few simple quality control (QC) assessments. This was performed in an automated fashion using a custom python script. For each fMRI session, we first determined whether behavioral and fMRI data sets were complete (i.e., all data files were present in the expected locations), whether scans were acquired in the expected order, and whether any scans were repeated. We also determined whether date and time stamps in behavioral and fMRI data matched. For data sets that passed the QC checks above, we automatically generated shell scripts for data processing, the steps for which are described below. Data sets that failed one or both QC checks were flagged for manual intervention and held out of automated processing.

**Supplemental Figure 1.**
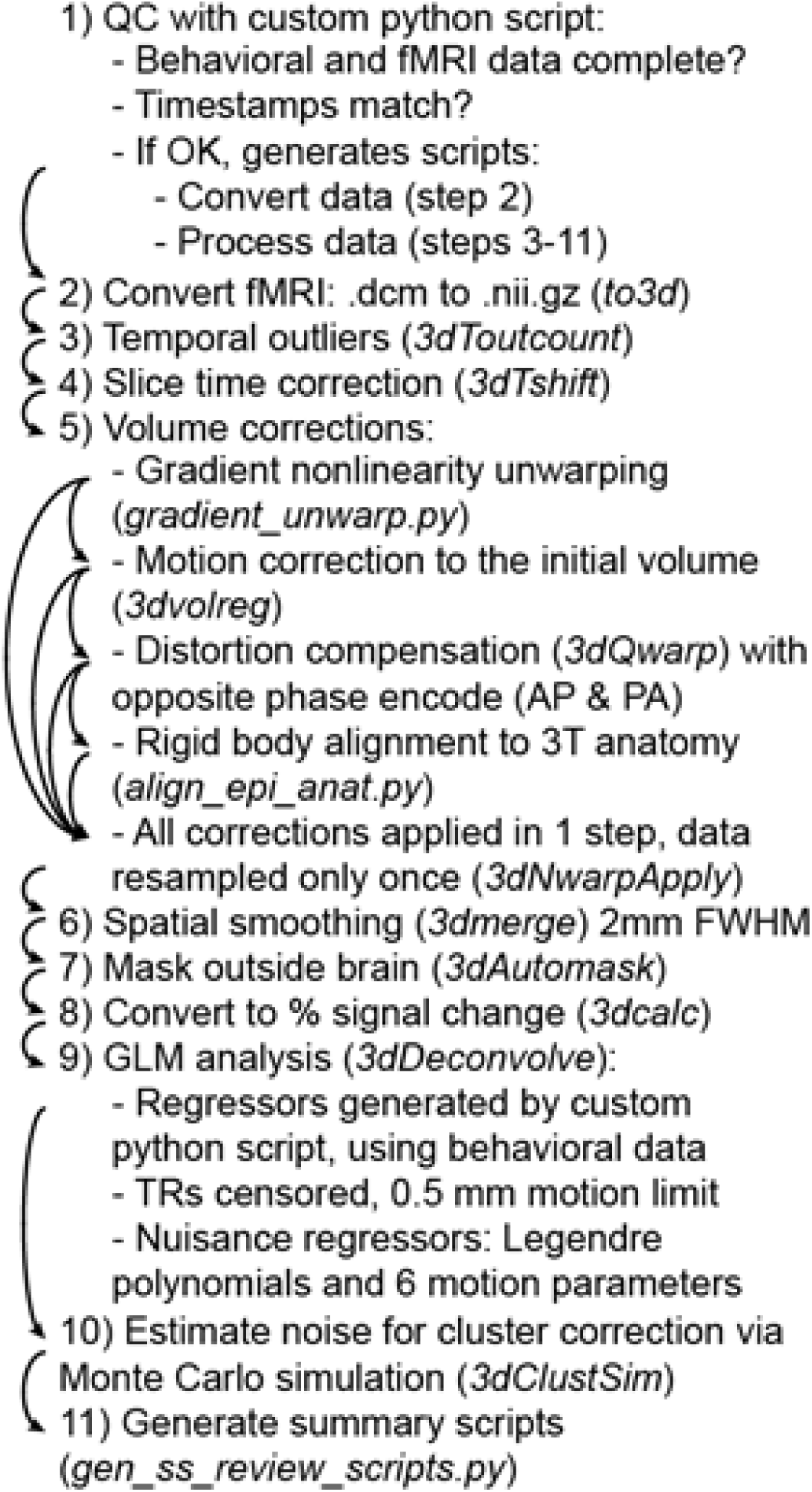
Our AFNI-based data processing pipeline. Numbers and arrows indicate sequential data processing steps. Function names are given in italics.

The first step in our fMRI data processing was to convert the 7T fMRI data from DICOM to compressed (g-zipped) nifti format using AFNI’s *to3d* function. We identified temporal outliers in the fMRI time series data using AFNI’s *3dToutcount*, which included masking out non-brain regions (via AFNI’s *3dAutomask*) and de-trending the time series using 4th order Legendre polynomials. For each TR, the fraction of voxels within the mask that were flagged as outliers (based on calculating the median absolute deviation of that voxel’s time series) was found. We then performed slice time correction using AFNI’s *3dTshift*, with the slice timing information extracted automatically from the header.

Next, we performed geometric corrections and transformations, which included 4 steps: 1) gradient nonlinearity unwarping (*gradunwarp* version 1.0.3; <u>github.com/Washington-University/gradunwarp</u>), comparable to that implemented in the HCP pipeline (Glasser et al., 2013). 2) Motion correction using AFNI’s *3dvolreg*, with the first AP functional scan (PRF 1) as the base. 3) Compensation for geometric distortions due to B_0_ inhomogeneities using AFNI’s *3dQwarp*. We used a pair of gradient echo EPI scans with opposite phase encoding directions (posterior-anterior and anterior-posterior) acquired sequentially to map the distortion. We have recently shown that this technique yielded superior distortion compensation across the whole brain within our data set, as compared to B_0_ field mapping or spin echo opposite phase encoding methods (Schallmo et al., 2021). 4) Co-registration of the 7T fMRI data to the 3T T_1_-weighted anatomical scan using AFNI’s *align_epi_anat.py*. Here, we used a local Pearson correlation (lpc) cost function (Saad et al., 2009) and rigid-body (6-parameter) alignment. All of these corrections were calculated sequentially, but were applied to the 7T fMRI data in a single step (i.e., data were resampled only once) using AFNI’s *3dNwarpApply*, in order to avoid additional blurring that would occur from multiple resampling steps.

Following geometric corrections, we performed spatial smoothing (2 mm full-width half-max [FWHM]) using AFNI’s *3dmerge*. Then, non-brain regions of the fMRI data were masked out using AFNI’s *3dAutomask*. We converted our fMRI data to percent signal change using AFNI’s *3dcalc*.

To quantify fMRI response magnitude, we performed general linear model (GLM) analyses using AFNI’s *3dDeconvolve*. This included task regressors generated in Python based on the behavioral data collected during fMRI scanning. We censored out (i.e., removed from analysis) TRs in which head motion was greater than 0.5 mm, as identified during the earlier motion correction step. Nuisance regressors included Legendre polynomials to capture low frequency signal oscillations (polynomial order was selected for each task automatically using AFNI’s default, which is 1 + [number of TRs / 150]). The 6 motion parameters estimated during motion correction were also included as nuisance regressors.

Following the GLM analysis, we obtained an estimate of the spatial profile of noise in our fMRI data using Monte Carlo simulation via AFNI’s *3dClustSim* (Cox et al., 2017; Eklund et al., 2016). Finally, summary scripts were generated using AFNI’s *gen_ss_review_scripts.py*, which included information about the number of temporal outliers and TRs censored for excessive head motion in each fMRI scan.

#### MRS processing

We used a custom python script to perform some basic, automated QC assessments of our MRS data. These checks were performed separately for both the OCC and PFC VOIs in each MRS dataset. We first assessed whether or not all expected MRS data files were present, and verified that the correct number of shots (TRs) were acquired based on the number of DICOM files. Next, we verified that the VOI was correct based on automated parsing of the DICOM file header. Finally, we tabulated VOI position and orientation based on DICOM header information, as well as shim quality (water linewidth in Hz) and transmit voltage from our (manually entered) scanning notes.

We processed our MRS data using the *matspec* toolbox (<u>github.com/romainVala/matspec</u>) in MATLAB (versions 2014a and 2009a). First, we performed eddy current correction for both our STEAM metabolite spectra and water reference data. To correct a known artifact, we then removed the first data point from each TR in the metabolite spectrum, and replaced it with a zero at the end of the spectrum. We then performed frequency and phase correction by finding the maximum amplitude for the spectrum at each TR within a range of 1.65 - 2.25 parts per million (ppm, i.e., the *N*-acetyl aspartate peak), and adjusting the frequency for all points in the spectrum such that the frequency of this maximum was 2.01 ppm. The phase for the maximum value was adjusted to zero on each TR. TRs with known signal artifacts (e.g., due to head motion) that could not be corrected in this way were removed manually. During frequency and phase correction, 4 Hz line broadening was applied; afterward, frequency and phase adjustments were applied to the original spectra without line broadening. We repeated our frequency and phase correction procedure across three iterations to improve correction quality. In the first two iterations, corrections were performed using the absolute value of the spectral data, while in the third iteration corrections were performed using the real portion of the complex data. A small number of data sets (n = 3 in OCC and n = 5 in PFC) were manually excluded from our analyses due to artifacts observed during data processing (e.g., failed water suppression).

Concentrations for different metabolites were quantified from the MRS data for each VOI in each scanning session using LCModel version 6.3-1N (Provencher, 2001). Our basis set was derived from previous work (Marjańska et al., 2017), and included the following metabolites: ascorbic acid, aspartic acid, creatine, GABA, glucose, glutamate, glutamine, glutathione, glycerophosphorylcholine, lactate, *myo*-inositol, *N*-acetyl aspartate (NAA), *N*-acetyl aspartylglutamate (NAAG), phosphocreatine, phosphorylcholine, phosphorylethanolamine, *scyllo*-inositol, taurine, as well as lipids and macromolecules. Macromolecule signals included in the basis set were obtained from inversion-recovery experiments (Behar et al., 1994) in the OCC region of 4 healthy young adults from a previous study (Marjańska et al., 2017). The Cramér-Rao Lower Bound (CRLB) provided an estimate of the lower limit of the variance for the fit concentration values (Cavassila et al., 2001; Landheer and Juchem, 2021). We scaled metabolite concentrations (millimolar) relative to the unsuppressed water signal, after correcting for differences in gray matter (GM), white matter (WM), and CSF fractions within the region of each participant’s MRS voxel. For this purpose, we used assumed values for water content in these tissue types (GM = 0.8, WM = 0.71, CSF = 0.97), and the different *T_1_* and *T*_2_ relaxation time constants of water within each compartment (*T_1_* gray matter = 2130 ms, *T_1_* white matter = 1220 ms, *T_1_* CSF = 4425 ms, *T_2_* gray matter = 50 ms, *T_2_* white matter = 55 ms, *T_2_* CSF = 141 ms), based on previous work (Marjańska et al., 2017). Tissue fractions were quantified in each VOI in each participant using individual gray matter and white matter surface models from FreeSurfer, after alignment of the in-session *T*_1_ anatomy (partial coverage due to MRS surface coil) to the whole-brain *T*_1_ data acquired at 3 T (processed with FreeSurfer). *T_1_* and *T_2_* relaxation times for the various metabolites were not accounted for in our analysis, as their effects are expected to be very small given the long TR(s) and the short TE (8 ms; Marjańska et al., 2017).

#### Data analysis

##### pRF analysis

PRF analyses of data (demeaned and smoothed by a 2 mm kernel) were conducted in AFNI, using a pRF implementation for the AFNI distribution (see Silson et al., 2018). We created a 2D+time binary mask of the stimulus input (191 x 191 voxels) as well as a convolution reference time series for the functional data using AFNI’s *3dDeconvolve* function with GAM as the response model. The AFNI model used these inputs and both Simplex and Powell algorithms to find the optimized time series/parameter sets. For each voxel, the model outputs the (*x, y*) coordinates representing the center of the receptive field, *sigma*, representing the diameter (size) of the receptive field, and *R*^2^, the explained variance of the fit used to statistically threshold the data.

##### Temporal frequency functional localizer

We used AFNI’s *3dDeconvolve* command to run a general linear model (GLM) analysis in order to estimate the voxelwise magnitude of the hemodynamic responses evoked by the 2 conditions: 2 Hz and 12 Hz flicker frequency. The functional ROI was defined by contrasting 2 Hz minus 12 Hz.

##### Functional mapping of auditory cortex

We used Fourier analysis methods (Engel, 1997) to map fMRI responses in auditory cortex to the auditory tone sweep stimuli. Since each of the 2 runs started with either an increasing tone-sweep or decreasing tone-sweep, one of the runs was time-shifted 4 frames and time-reversed so the runs could be averaged together. Then, we filtered out the lowest frequency, and the coherence and phase were calculated from the time series data in each voxel. Coherence (similar to an unsigned correlation) was defined as the amplitude at a frequency divided by the absolute value of the square root of the sum of squared amplitudes at all frequencies (Engel, 1997).

##### Functional mapping of motor cortex

For mapping motor cortex, we used AFNI’s *3dDeconvolve* command to run a GLM analysis in order to estimate the voxelwise magnitude of the hemodynamic responses evoked by each of the 5 conditions: right fingers (RF), left fingers (LF), right toes (RT), left toes (LT), and Tongue. Functional ROIs for each condition were defined by contrasting the target body part with all other body parts (e.g., for RF: RF > RT + RT + LT + Tongue). For each ROI in each participant, we chose the largest cluster (or largest 2 clusters for the Tongue condition) that overlapped with precentral/paracentral gyrus as defined by that participant’s FreeSurfer parcellation. The group-level ROI represents the voxels significantly modulated in at least 60% of participants for that contrast.

##### CSS analysis

We fit the behavioral data from our CSS psychophysical contrast discrimination task with a psychometric (Weibull) function in order to obtain discrimination thresholds (Figure 8A). Contrast increment versus accuracy data were combined across the 3 independent staircases and fit within each stimulus condition (i.e., pedestal contrast) separately, yielding 8 independent threshold estimates (evaluated at 80% accuracy). This allowed us to examine threshold-versus-contrast curves, which show a characteristic ‘dipper’ shape (i.e., lowest thresholds at low, non-zero pedestal contrast; Figure 8B).

Functional MRI data from our CSS task were analyzed within regions-of-interest (ROIs) in area V1. These V1 ROIs were defined using the functional localizer data (i.e., the first 90 s task condition from the first scanning run). We used a Fourier analysis (Engel, 1997; Schallmo et al., 2016) to identify voxels that responded selectively to the center > surrounding stimulus (green voxels in Figure 8C). Center-selective voxels were identified based on a coherence (similar to an unsigned correlation) threshold ≥ 0.2, and phase values between 7π/8 to 11π/8 (i.e., in phase with the center stimulus presentation, accounting for the delayed hemodynamic response). V1 ROIs were defined in AFNI using a cluster correction method (Cox et al., 2017; Eklund et al., 2016), with a whole-brain significance threshold of *p* < 0.01. V1 clusters were identified in each hemisphere in each individual and scanning session using a V1 anatomical mask (Wang et al., 2015) from FreeSurfer and manual inspection of the participant’s functional activation map and occipital anatomy in SUMA and AFNI. V1 responses in the CSS fMRI task were quantified in terms of percent signal change using a Fourier analysis in MATLAB, based on the stimulus presentation frequency (5 cycles of target-on, target-off per 90 s stimulus condition).

##### COP analyses

Our analyses of the psychophysical and fMRI data from the COP task matched that in the CSS task above, with the following differences. Data from the 3 staircases in each block were combined for fitting purposes, yielding 2 independent threshold estimates for contour jitter versus accuracy (1 in each block; Figure 9A). Thresholds were assessed at 70% accuracy in the COP psychophysical task. This was done in order to enable quantification of thresholds for participants with relatively low ceiling performance (i.e., contour discrimination accuracy ∼80% for aligned contours with 0° jitter). V1 ROIs in the COP task were defined using a Fourier analysis (contour > blank), with a correlation threshold ≥ 0.3 and phase values 0 to 3π/8 and 15π/8 to 2π (Figure 9C). V1 responses in the COP fMRI task were quantified in terms of percent signal change (i.e., beta weights) using a GLM analysis in AFNI (see FMRI Processing in Supplemental Methods).

##### SFM analysis

Switch rates (in Hz) for bi-stable percepts were quantified by dividing the number of switches reported in each 2 min block and dividing by 120 sec, and then averaging across all 5 bi-stable stimulus blocks. Repeated responses indicating the same perceived direction were ignored.

##### MRS analysis

In order to visualize average MRS VOI positions in OCC and PFC, we transformed binary ROI masks from the individual participant space to MNI space using FreeSurfer’s *mri_convert* function and the *talairach.xfm* from each participant. We then computed the voxel-wise average VOI position using AFNI’s *3dMean*. Average VOIs were visualized on a canonical brain image in AFNI (Figure 12B & D), thresholded at > 30% overlap across participants. MRS results from LCModel were analyzed in MATLAB.

### Supplemental results

**Supplemental Table 1.**
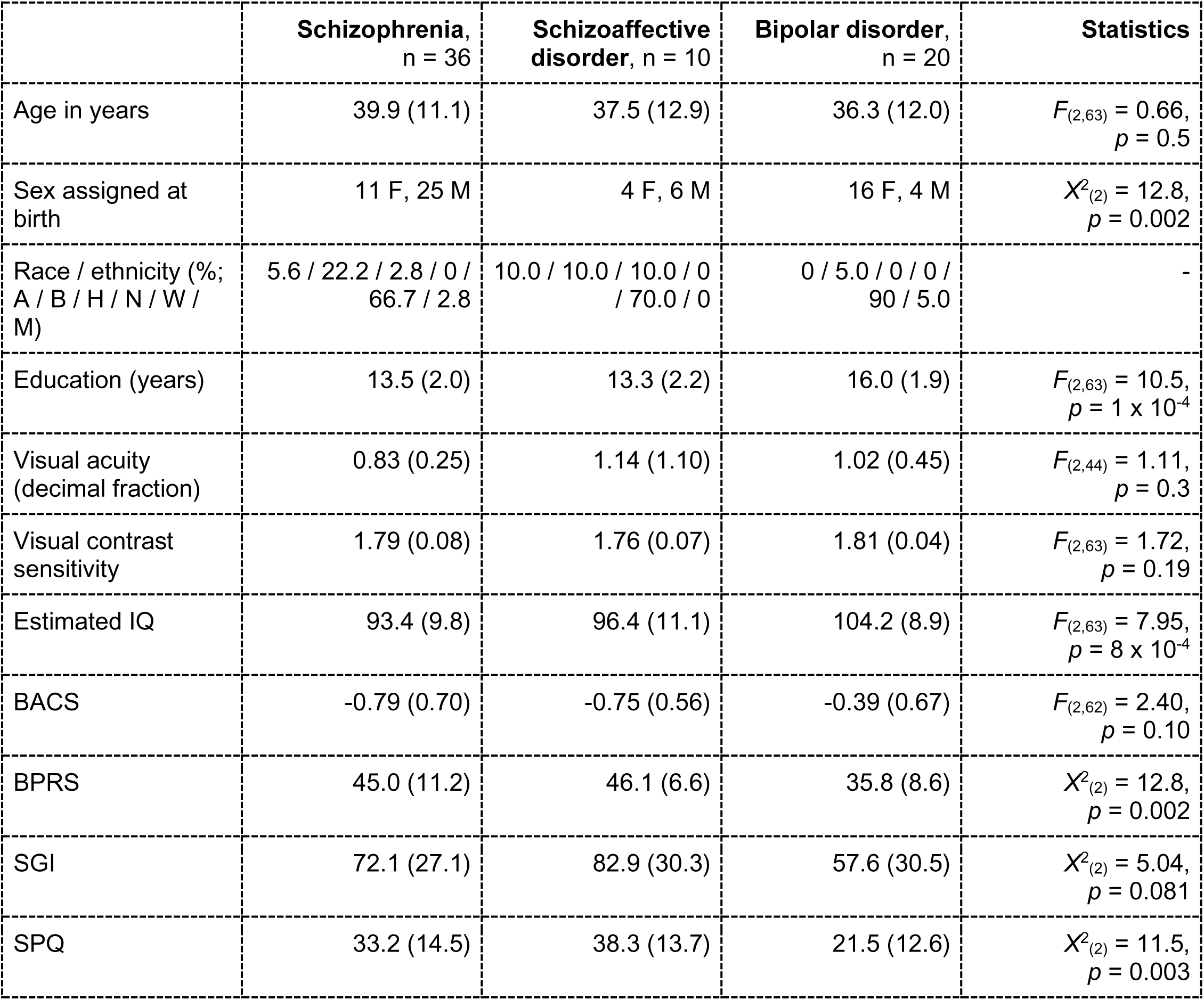
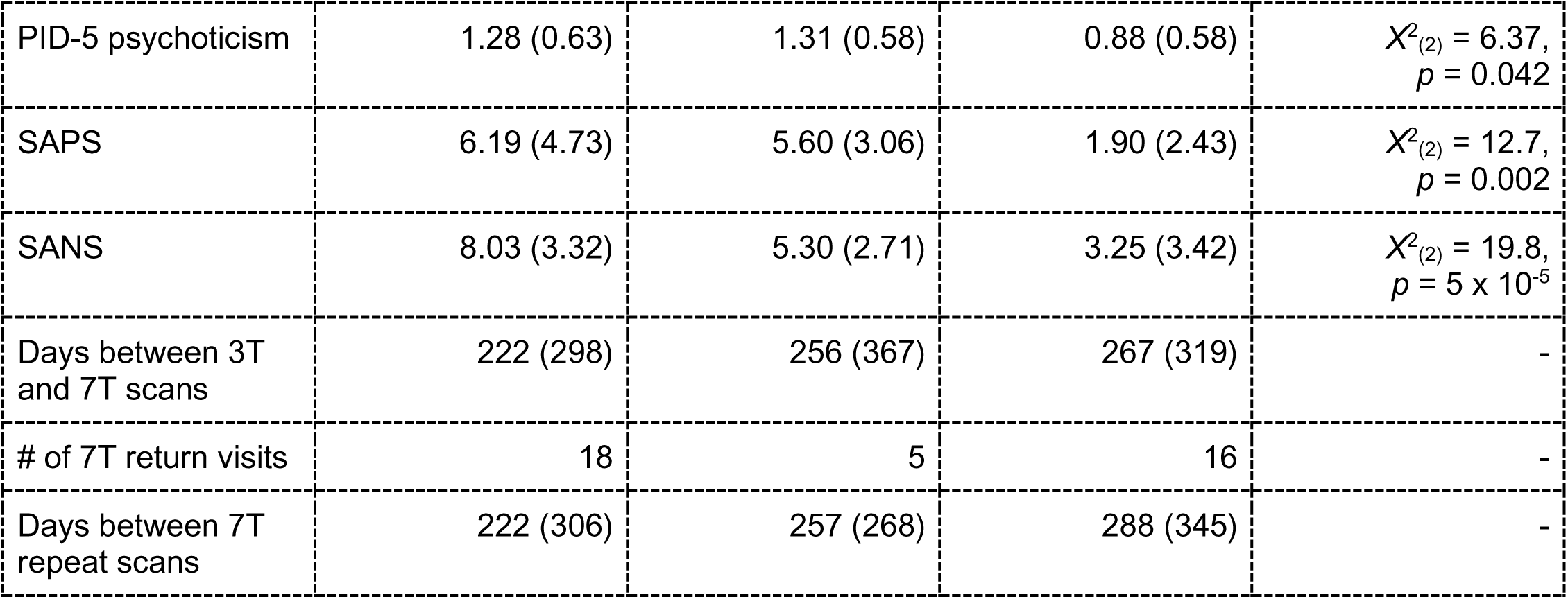
Demographics, cognitive, and symptom measures for PwPP, by diagnostic group. Data are presented as mean (standard deviation), unless otherwise noted. Racial and ethnic designations (as defined by the National Institute of Health) are abbreviated as follows: A = Asian or Pacific Islander, B = Black (not of Hispanic origin), H = Hispanic, N = Native American or Alaskan Native, W = White (not of Hispanic origin), M = More than 1 race or ethnicity, or other. Visual acuity was assessed with a Snellen eye chart (Snellen, 1862); the decimal fraction is reported (e.g., 0.5 indicates 20/40). Visual contrast sensitivity was assessed with the Mars Letter Contrast Sensitivity test (Arditi, 2005). Estimated IQ was assessed using the Wechsler Adult Intelligence Scale (WAIS-IV; Wechsler, 2008). BACS = Brief Assessment of Cognition in Schizophrenia, Z-score (Keefe et al., 1999), BPRS = Brief Psychiatric Rating Scale (Ventura et al., 2000), SGI = Sensory Gating Inventory (Hetrick et al., 2012), SPQ = Schizotypal Personality Questionnaire (Raine, 1991), PID-5 psychoticism = psychoticism factor from the Personality Inventory for DSM-5 (Krueger et al., 2012), SANS = Scale for the Assessment of Negative Symptoms (Andreasen, 1982), SAPS = Scale for the Assessment of Positive Symptoms (Andreasen, 1984). Diagnoses were based on the Structured Clinical Interview for DSM-IV-TR disorders (SCID; First, 1997). Data collected at repeat scans were not included here. For the relative group, the number of related probands with a particular psychotic disorder diagnosis is listed in square brackets. The statistics column shows the test statistic and *p*-value for differences across all three groups in each measure. For measures where normality and / or homogeneity of variance were not observed, non-parametric Kruskal-Wallis tests (*Χ*^2^-values) were used in place of analyses of variance (*F*-values).

|  | **Schizophrenia,**<br>n = 36 | **Schizoaffective**<br>**disorder, n = 10** | **Bipolar disorder,**<br>n = 20 | **Statistics** |
| --- | --- | --- | --- | --- |
| Age in years | 39.9 (11.1) | 37.5 (12.9) | 36.3 (12.0) | $F_{(2,63)} = 0.66,$<br>$p = 0.5$ |
| Sex assigned at birth | 11 F, 25 M | 4 F, 6 M | 16 F, 4 M | $X^2_{(2)} = 12.8,$<br>$p = 0.002$ |
| Race / ethnicity (%;<br>A / B / H / N / W /<br>M) | 5.6 / 22.2 / 2.8 / 0 /<br>66.7 / 2.8 | 10.0 / 10.0 / 10.0 / 0<br>/ 70.0 / 0 | 0 / 5.0 / 0 / 0 /<br>90 / 5.0 | - |
| Education (years) | 13.5 (2.0) | 13.3 (2.2) | 16.0 (1.9) | $F_{(2,63)} = 10.5,$<br>$p = 1 \times 10^{-4}$ |
| Visual acuity<br>(decimal fraction) | 0.83 (0.25) | 1.14 (1.10) | 1.02 (0.45) | $F_{(2,44)} = 1.11,$<br>$p = 0.3$ |
| Visual contrast<br>sensitivity | 1.79 (0.08) | 1.76 (0.07) | 1.81 (0.04) | $F_{(2,63)} = 1.72,$<br>$p = 0.19$ |
| Estimated IQ | 93.4 (9.8) | 96.4 (11.1) | 104.2 (8.9) | $F_{(2,63)} = 7.95,$<br>$p = 8 \times 10^{-4}$ |
| BACS | -0.79 (0.70) | -0.75 (0.56) | -0.39 (0.67) | $F_{(2,62)} = 2.40,$<br>$p = 0.10$ |
| BPRS | 45.0 (11.2) | 46.1 (6.6) | 35.8 (8.6) | $X^2_{(2)} = 12.8,$<br>$p = 0.002$ |
| SGI | 72.1 (27.1) | 82.9 (30.3) | 57.6 (30.5) | $X^2_{(2)} = 5.04,$<br>$p = 0.081$ |
| SPQ | 33.2 (14.5) | 38.3 (13.7) | 21.5 (12.6) | $X^2_{(2)} = 11.5,$<br>$p = 0.003$ |
| PID-5 psychoticism | 1.28 (0.63) | 1.31 (0.58) | 0.88 (0.58) | $X^2_{(2)} = 6.37,$<br>$p = 0.042$ |
| SAPS | 6.19 (4.73) | 5.60 (3.06) | 1.90 (2.43) | $X^2_{(2)} = 12.7,$<br>$p = 0.002$ |
| SANS | 8.03 (3.32) | 5.30 (2.71) | 3.25 (3.42) | $X^2_{(2)} = 19.8,$<br>$p = 5 \times 10^{-5}$ |
| Days between 3T<br>and 7T scans | 222 (298) | 256 (367) | 267 (319) | - |
| # of 7T return visits | 18 | 5 | 16 | - |
| Days between 7T<br>repeat scans | 222 (306) | 257 (268) | 288 (345) | - |

**Supplemental Table 2.** Number of data sets collected under the protocols we refer to as 7T-A and 7T-B / Z in each experiment and participant group. Note that the number of unique participants is lower than the total number of data sets, as some participants returned for a follow-up scan, as reported in Table 1. Psych. = psychophysics.

|  |  | **Controls** | **Relatives** | **PwPP** |
| --- | --- | --- | --- | --- |
| 7T-A | CSS psych. | 10 | 0 | 5 |
|  | COP psych. | 17 | 0 | 5 |
|  | COP fMRI | 12 | 0 | 3 |
| 7T-B / Z | CSS psych. | 33 | 44 | 100 |
|  | COP psych. | 34 | 44 | 100 |
|  | COP fMRI | 33 | 39 | 74 |

**Supplemental Table 3.** Longitudinal variability of clinical measures. Data presented are from n = 38 PwPP who completed two scanning sessions (e.g., 7T-B & 7T-Z) and for whom BPRS (Ventura et al., 2000), SANS (Andreasen, 1982), and SAPS (Andreasen, 1984) data were acquired during both visits. Rows show data for these three measures, columns show the metrics quantifying change between scans after taking the absolute value (abs.) of the difference between data from scan 1 and scan 2. SD = standard deviation.

|  | Mean abs. change | SD abs. change | Median abs. change |
| --- | --- | --- | --- |
| **BPRS total score** | 6.37 | 5.01 | 4.5 |
| **SANS total global score** | 2.37 | 2.42 | 1.5 |
| **SAPS total global score** | 2.00 | 1.93 | 2 |

**Supplemental Figure 2.**
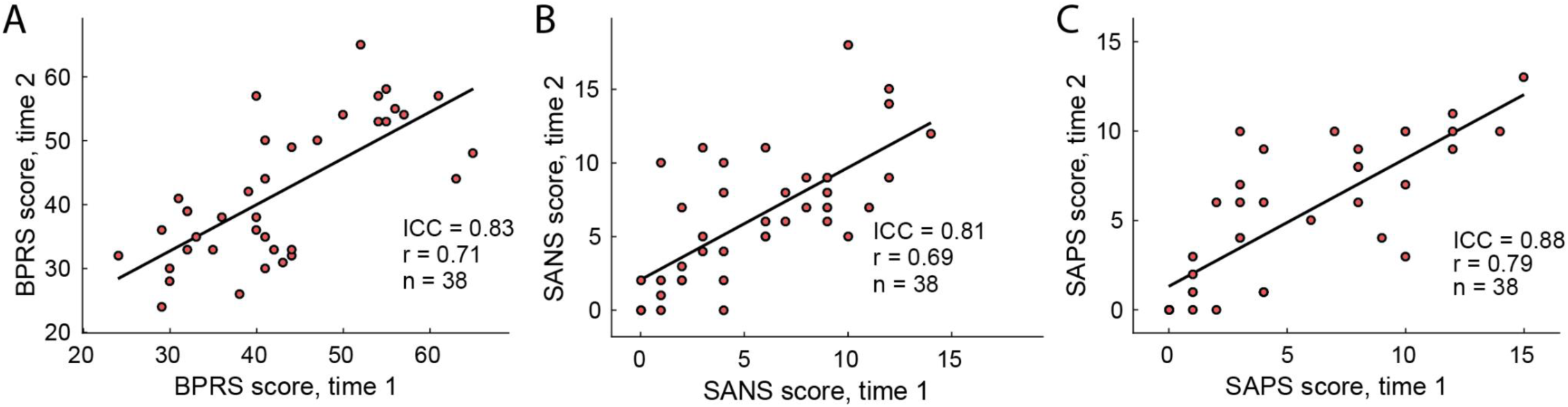
Scatter plots of clinical measures over time. Data presented are from n = 38 PwPP who completed two scanning sessions (e.g., 7T-B & 7T-Z) and for whom BPRS (Ventura et al., 2000), SANS (Andreasen, 1982), and SAPS (Andreasen, 1984) data were acquired during both visits. A) BPRS total score, B) SANS total global score, C) SAPS total global score. X-axes show data from scanning session 1, y-axes show data from session 2. Black line shows the linear trend. ICC = intraclass correlation coefficient, ICC(3,k), (Koo and Li, 2016); r = Pearson’s r-value.

**Supplemental Table 4.** Number of data sets with eye tracking data. Note that the number of unique participants is lower than the total number of data sets, as some participants returned for a follow-up scan, as reported in Table 1.

|  |  | **Controls** | **Relatives** | **PwPP** |
| --- | --- | --- | --- | --- |
| Psychophysics | CSS | 24 | 33 | 81 |
|  | COP | 24 | 34 | 81 |
|  | SFM | 26 | 35 | 85 |
| fMRI | pRF | 19 | 20 | 42 |
|  | CSS | 18 | 19 | 40 |
|  | COP | 13 | 17 | 34 |

**Supplemental Table 5.** Statistical comparisons of psychophysics and fMRI data quality metrics between groups. As most metrics were not normally distributed (see Supplemental Figure 3, Supplemental Figure 4, & Supplemental Figure 5), Kruskal-Wallis nonparametric 1-way ANOVAs were used to quantify between-group differences.

|  | **pRF** | **CSS** | **COP** | **SFM** |
| --- | --- | --- | --- | --- |
| Psychophysical catch trial accuracy | - | $X^2_{(2)} = 8.99$,<br>$p = 0.011$ | $X^2_{(2)} = 4.30$,<br>$p = 0.12$ | $X^2_{(2)} = 2.41$,<br>$p = 0.3$ |
| fMRI head motion (# TRs censored) | $X^2_{(2)} = 7.71$,<br>$p = 0.021$ | $X^2_{(2)} = 11.8$,<br>$p = 0.003$ | $X^2_{(2)} = 14.4$,<br>$p = 8 \times 10^{-4}$ | - |
| fMRI behavioral responses (% of trials) | $X^2_{(2)} = 10.1$,<br>$p = 0.006$ | $X^2_{(2)} = 6.08$,<br>$p = 0.048$ | $X^2_{(2)} = 3.45$,<br>$p = 0.18$ | - |
| fMRI head motion (avg. per TR) | $X^2_{(2)} = 8.99$,<br>$p = 0.011$ | $X^2_{(2)} = 10.1$,<br>$p = 0.006$ | $X^2_{(2)} = 10.0$,<br>$p = 0.007$ | - |
| fMRI temporal outliers (% of voxels per TR) | $X^2_{(2)} = 6.87$,<br>$p = 0.032$ | $X^2_{(2)} = 11.1$,<br>$p = 0.004$ | $X^2_{(2)} = 10.8$,<br>$p = 0.005$ | - |

**Supplemental Figure 3.**
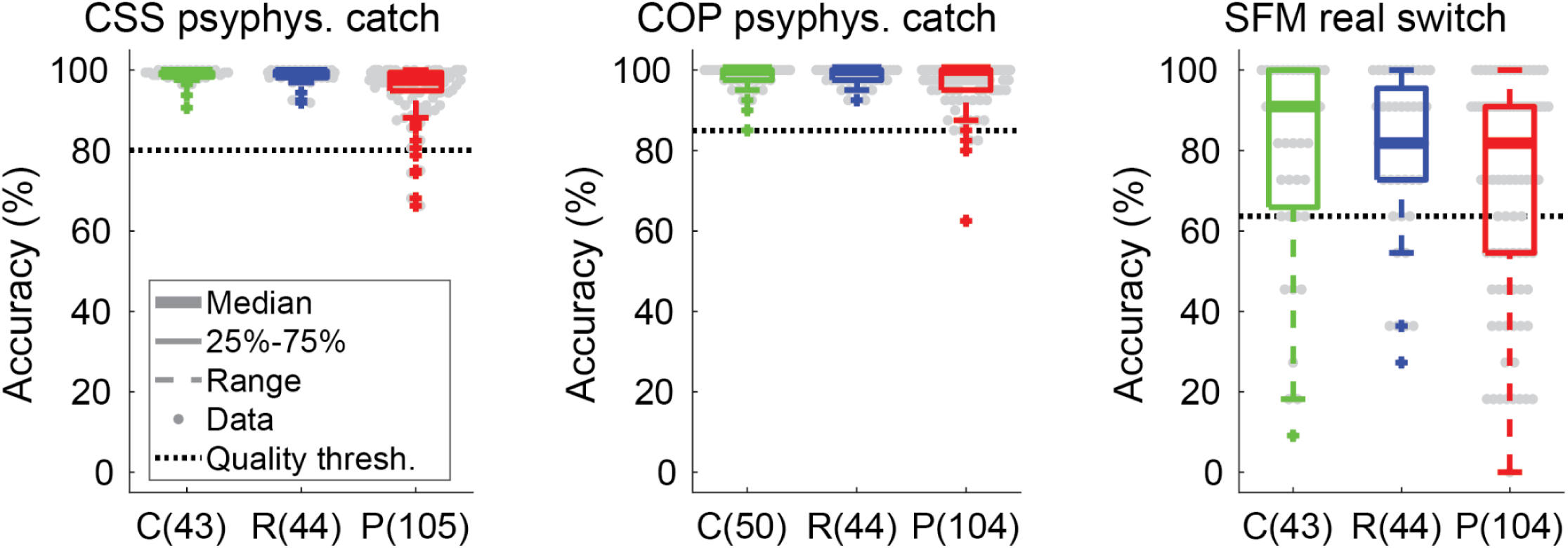
Psychophysical data quality checks across groups and experiments. Panels show catch trial accuracy from the CSS (left) and COP (middle) psychophysical (psyphys.) tasks, as well as accuracy for responses made in < 4 s in the SFM real switch psychophysical task (right). The number of data sets (not unique individuals) per group and experiment are shown in parentheses. C = healthy controls (green), R = first-degree biological relatives (blue), P = people with psychotic psychopathology (red). Thick lines show group medians, boxes show 25-75%, whiskers show 1.5 x interquartile range, dots show individual data points, dashed black lines show data quality thresholds.

**Supplemental Figure 4.**
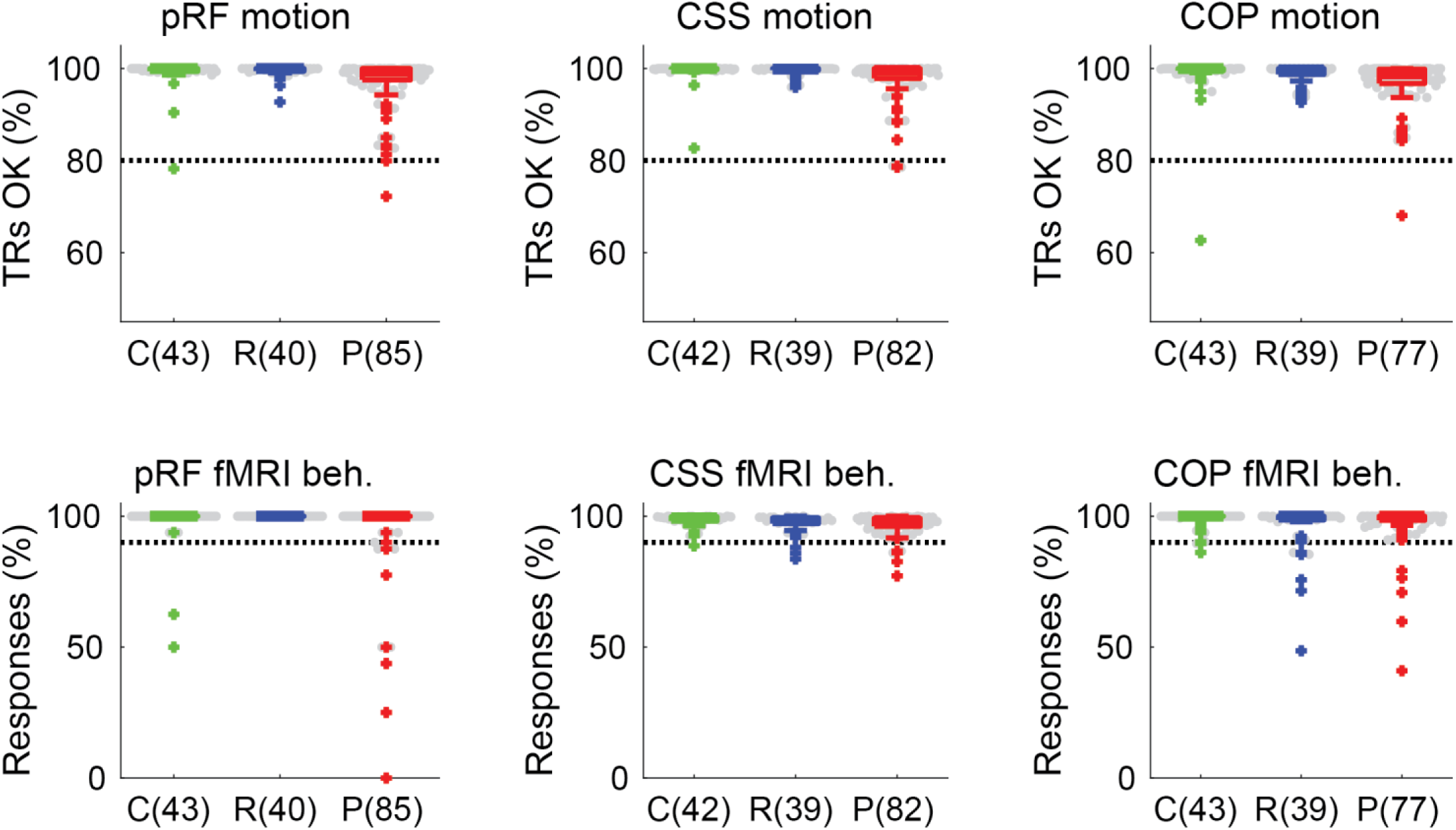
FMRI data quality checks across groups and experiments. Top row shows the proportion of TRs with < 0.5 mm of head motion in the pRF (left), CSS (middle), and COP (right) fMRI tasks. Bottom row shows the same, but for the proportion of trials in which a behavioral (beh.) response was recorded (regardless of accuracy). The number of data sets (not unique individuals) per group and experiment are shown in parentheses. C = healthy controls (green), R = first-degree biological relatives (blue), P = people with psychotic psychopathology (red). Thick lines show group medians, boxes show 25-75%, whiskers show 1.5 x interquartile range, dots show individual data points, dashed black lines show data quality thresholds.

**Supplemental Figure 5.**
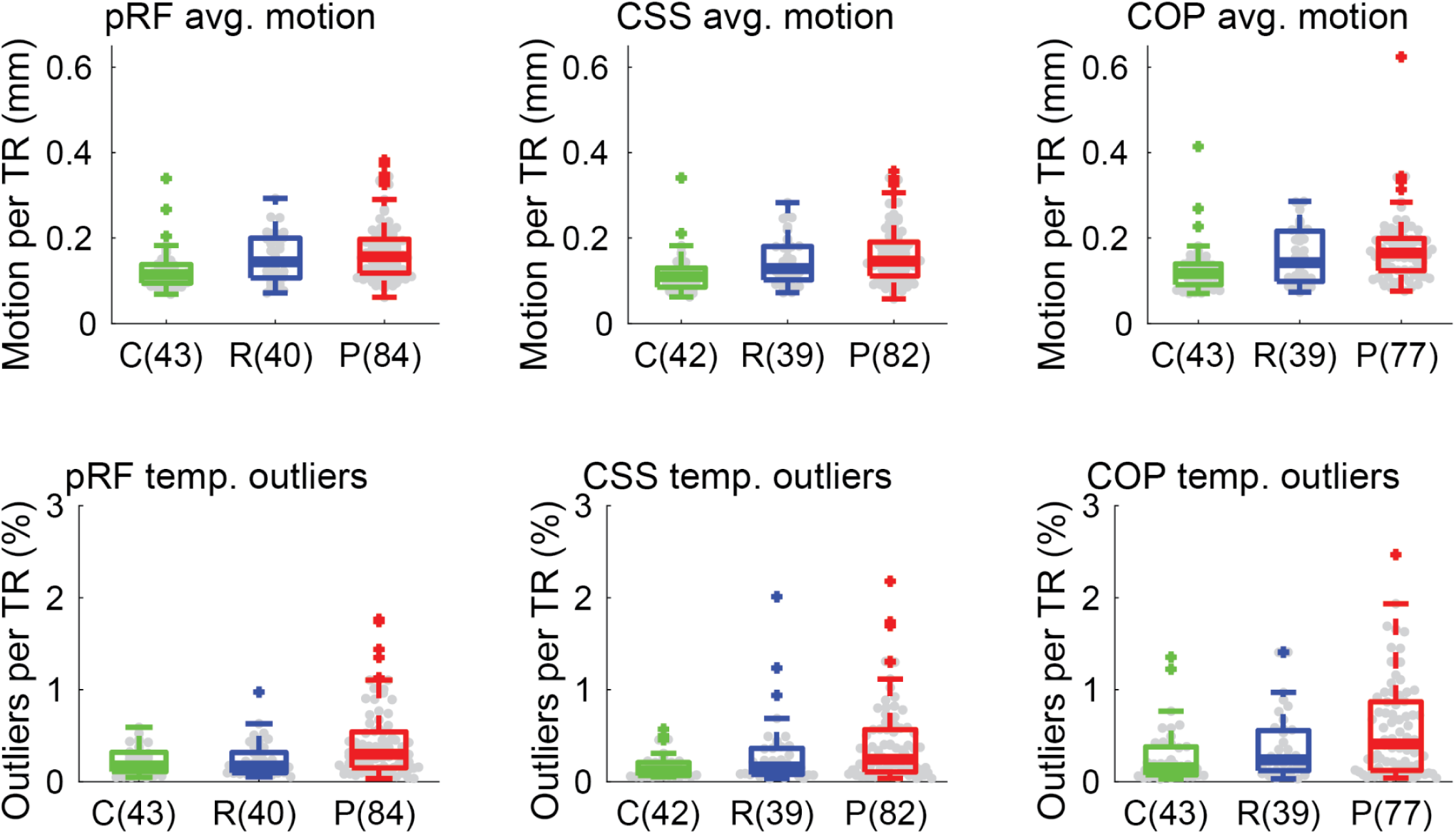
Motion per TR and temporal outliers per TR, across groups and fMRI experiments. Top row shows the average (avg.) head motion per TR for the pRF (left), CSS (middle), and COP (right) fMRI tasks. The bottom row shows the same, but for the percent of voxels labeled as temporal (temp.) outliers per TR. The number of data sets (not unique individuals) per group and experiment are shown in parentheses. C = healthy controls (green), R = first-degree biological relatives (blue), P = people with psychotic psychopathology (red). Thick lines show group medians, boxes show 25-75%, whiskers show 1.5 x interquartile range, dots show individual data points.

**Supplemental Figure 6.**
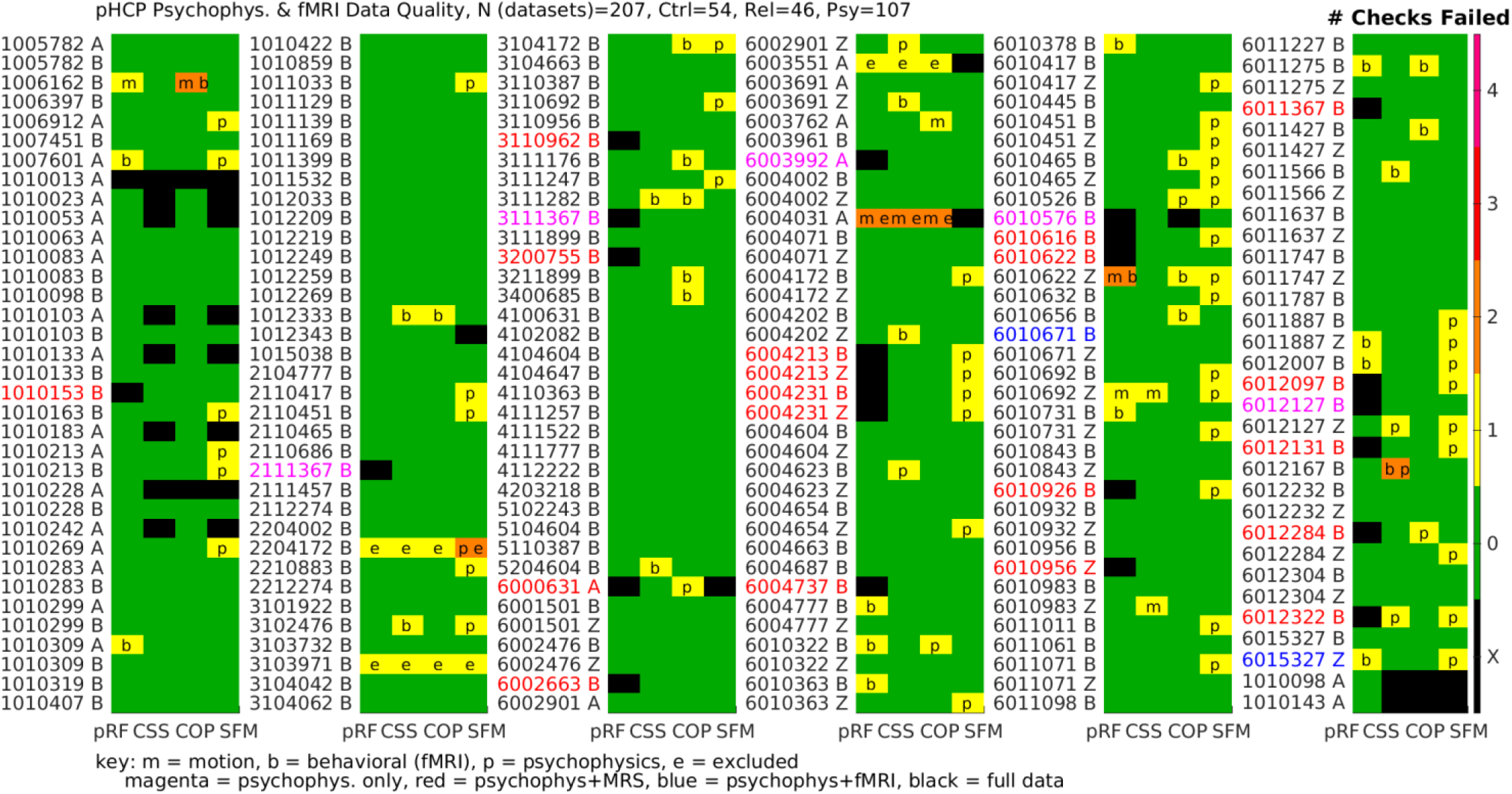
Psychophysics and fMRI data quality detailed summary. Rows show individual data sets labeled by participant ID number and protocol (A, B, or Z). The font color for the row labels indicates whether there is a full data set (black), psychophysics & fMRI only (blue), psychophysics & MRS only (red), or psychophysics only (magenta). Columns show data for the 4 different visual paradigms (pRF = population receptive field modeling, CSS = contrast surround suppression, COP = contour object perception, SFM = structure-from-motion). Colors within the table show the number of data quality checks that were failed (black = missing data). Letters within the table indicate which checks were failed (m = motion, b = fMRI behavioral task, p = psychophysical catch trials, e = excluded data). Note that data sets that were collected but subsequently excluded (e) appear here for the sake of completeness, but are not included elsewhere in the manuscript unless otherwise noted.

**Supplemental Table 6.** Statistical comparisons of MRS data quality metrics between groups. Kruskal-Wallis nonparametric 1-way ANOVAs were used to quantify between-group differences.

|  | **OCC** | **PFC** |
| --- | --- | --- |
| H<sub>2</sub>O linewidth | $X^2_{(2)} = 1.12,$<br>$p = 0.6$ | $X^2_{(2)} = 3.02,$<br>$p = 0.2$ |
| LCModel linewidth | $X^2_{(2)} = 3.24,$<br>$p = 0.2$ | $X^2_{(2)} = 2.33,$<br>$p = 0.3$ |
| SNR | $X^2_{(2)} = 6.87,$<br>$p = 0.032$ | $X^2_{(2)} = 2.08,$<br>$p = 0.4$ |

**Supplemental Figure 7.**
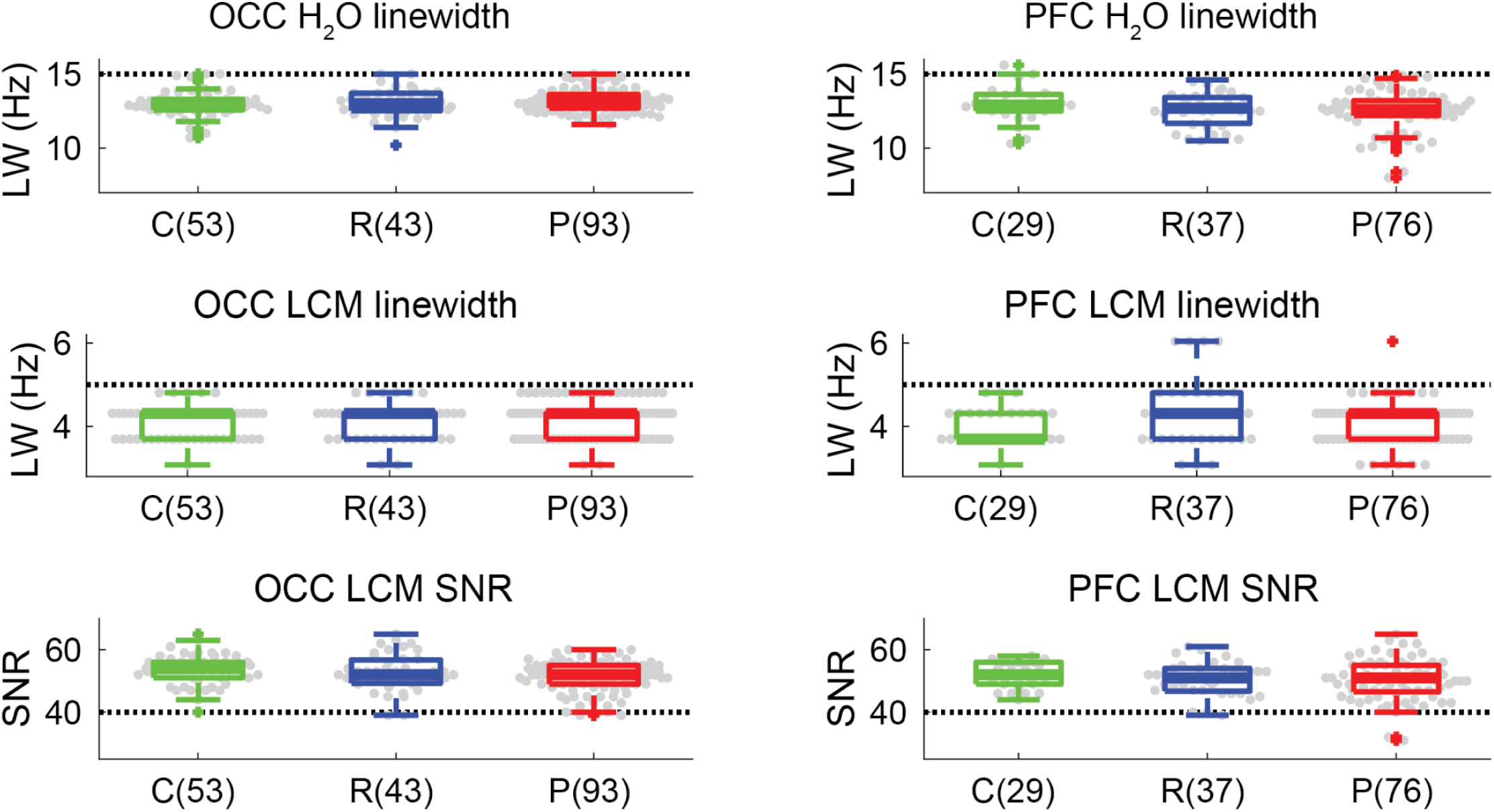
MRS data quality checks across groups and experiments. Top row shows the linewidth of water (H_2_O) in the OCC (left) and PFC (right) VOIs. Middle row shows the same, but for the linewidth of the fitted spectrum from LCModel (LCM), whereas the bottom row shows SNR from LCModel. The number of data sets (not unique individuals) per group and experiment are shown in parentheses. C = healthy controls (green), R = first-degree biological relatives (blue), P = people with psychotic psychopathology (red). Thick lines show group medians, boxes show 25-75%, whiskers show 1.5 x interquartile range, dots show individual data points, dashed black lines show data quality thresholds. Participants with missing quality data (e.g., linewidth of water was not recorded during scanning) are not shown. Note that in the PFC LCModel SNR plot (bottom right), an outlier data point (SNR = 13) from a single relative is not shown, in order to better visualize the distribution of the other data points.

**Supplemental Figure 8.**
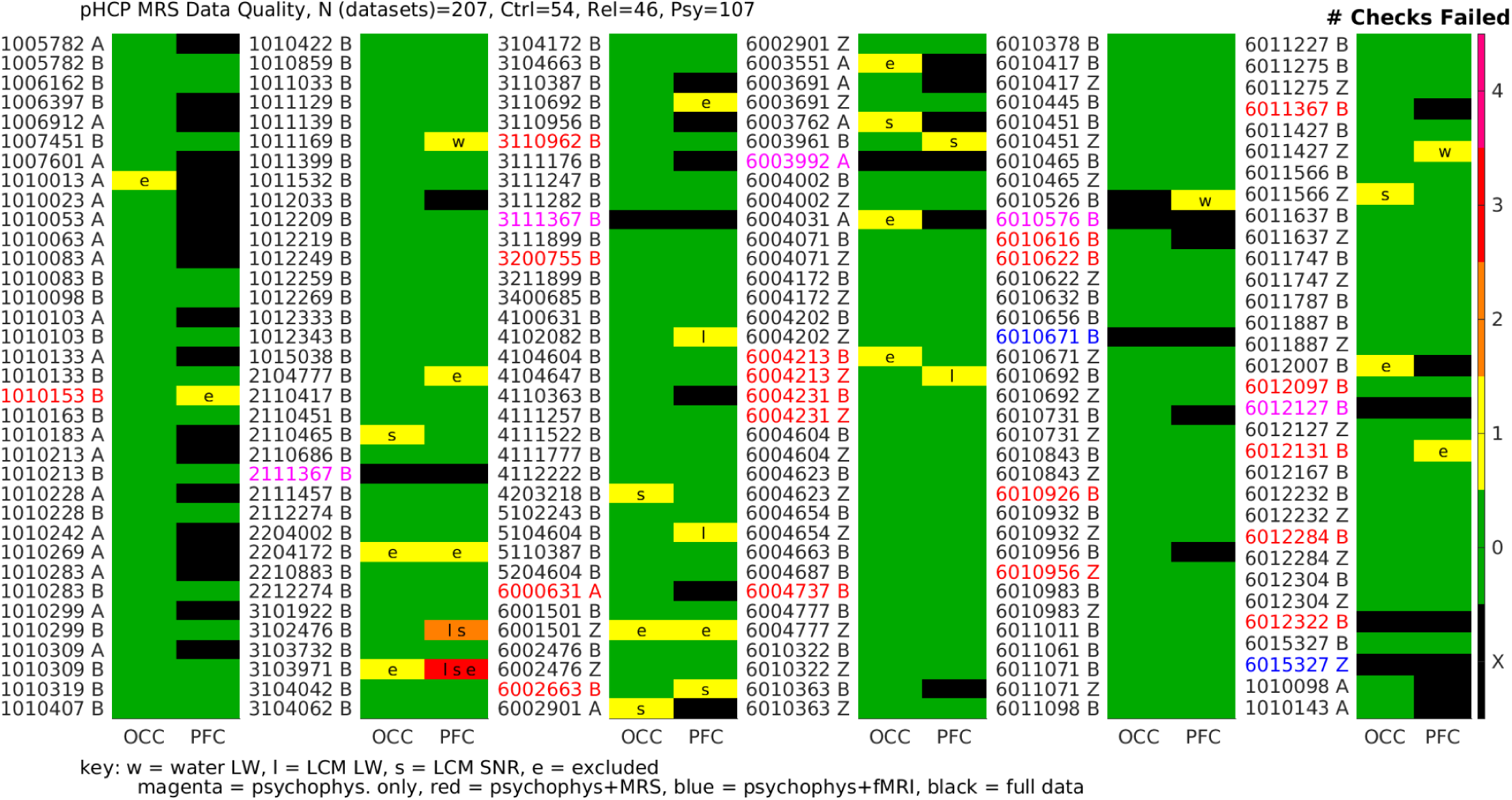
MRS data quality detailed summary. Rows show individual data sets labeled by participant ID number and protocol (A, B, or Z). The font color for the row labels indicates whether there is a full data set (black), psychophysics & fMRI only (blue), psychophysics & MRS only (red), or psychophysics only (magenta). Columns show data for the 2 different MRS VOIs (OCC = occipital cortex, PFC = prefrontal cortex). Colors within the table show the number of data quality checks that were failed (black = missing data). Letters within the table indicate which checks were failed (w = linewidth of water, l = linewidth from LCModel, s = SNR from LCModel, e = excluded data). Note that data sets that were collected but subsequently excluded (e) appear here for the sake of completeness, but are not included elsewhere in the manuscript unless otherwise noted.

