## Supplementary material for "The Psychosis Human Connectome Project: Design and rationale for studies of visual neurophysiology": FMRI Protocol

**Table of contents**

\\USER
PHCP
functional
final
Localizer
AAHScout_32ch
Localizer_aligned
BOLD_oppPE_AP
BOLD_PRF1_AP
BOLD_CSS_task1_AP
BOLD_CSS_task2_AP
BOLD_CSS_task3_AP
FieldMap
t1_mpr_tra_iso
BOLD_AP_SE
BOLD_REV_AP_SE
BOLD_COP_task1_AP
BOLD_COP_task2_AP
BOLD_PRF2_AP

| \\USER\PHCP\functional\final\Localizer |
|---|
| TA: 6.1 s PM: FIX Voxel size: 0.5×0.5×5.0 mmPAT: Off Rel. SNR: 1.00 : fl |

### Properties

| | |
|---|---|
| Prio recon | Off |
| Load images to viewer | On |
| Inline movie | Off |
| Auto store images | On |
| Load images to stamp segments | Off |
| Load images to graphic segments | Off |
| Auto open inline display | Off |
| Auto close inline display | Off |
| Start measurement without further preparation | Off |
| Wait for user to start | Off |
| Start measurements | Single measurement |

### Routine

| | |
|---|---|
| Slice group | 1 |
| Slices | 1 |
| Dist. factor | 20 % |
| Position | L0.0 A50.0 H0.0 mm |
| Orientation | Sagittal |
| Phase enc. dir. | A >> P |
| Slice group | 2 |
| Slices | 1 |
| Dist. factor | 20 % |
| Position | Isocenter |
| Orientation | Transversal |
| Phase enc. dir. | A >> P |
| Slice group | 3 |
| Slices | 1 |
| Dist. factor | 20 % |
| Position | Isocenter |
| Orientation | Coronal |
| Phase enc. dir. | R >> L |
| AutoAlign | --- |
| Phase oversampling | 0 % |
| FoV read | 250 mm |
| FoV phase | 100.0 % |
| Slice thickness | 5.0 mm |
| TR | 6.6 ms |
| TE | 2.67 ms |
| Averages | 1 |
| Concatenations | 3 |
| Filter | Elliptical filter |
| Coil elements | A32 |

### Contrast - Common

| | |
|---|---|
| TR | 6.6 ms |
| TE | 2.67 ms |
| TD | 0 ms |
| MTC | Off |
| Magn. preparation | None |
| Flip angle | 10 deg |
| Fat suppr. | None |
| Water suppr. | None |
| SWI | Off |

### Contrast - Dynamic

| | |
|---|---|
| Averages | 1 |
| Averaging mode | Short term |
| Reconstruction | Magnitude |
| Measurements | 1 |

### Contrast - Dynamic

| | |
|---|---|
| Multiple series | Each measurement |

### Resolution - Common

| | |
|---|---|
| FoV read | 250 mm |
| FoV phase | 100.0 % |
| Slice thickness | 5.0 mm |
| Base resolution | 256 |
| Phase resolution | 90 % |
| Phase partial Fourier | Off |
| Interpolation | On |

### Resolution - iPAT

| | |
|---|---|
| PAT mode | None |

### Resolution - Filter Image

| | |
|---|---|
| Image Filter | Off |
| Distortion Corr. | Off |
| Prescan Normalize | Off |
| Normalize | Off |
| B1 filter | Off |

### Resolution - Filter Rawdata

| | |
|---|---|
| Raw filter | Off |
| Elliptical filter | On |

### Geometry - Common

| | |
|---|---|
| Slice group | 1 |
| Slices | 1 |
| Dist. factor | 20 % |
| Position | L0.0 A50.0 H0.0 mm |
| Orientation | Sagittal |
| Phase enc. dir. | A >> P |
| Slice group | 2 |
| Slices | 1 |
| Dist. factor | 20 % |
| Position | Isocenter |
| Orientation | Transversal |
| Phase enc. dir. | A >> P |
| Slice group | 3 |
| Slices | 1 |
| Dist. factor | 20 % |
| Position | Isocenter |
| Orientation | Coronal |
| Phase enc. dir. | R >> L |
| FoV read | 250 mm |
| FoV phase | 100.0 % |
| Slice thickness | 5.0 mm |
| TR | 6.6 ms |
| Multi-slice mode | Sequential |
| Series | Interleaved |
| Concatenations | 3 |

### Geometry - AutoAlign

| | |
|---|---|
| Slice group | 1 |
| Position | L0.0 A50.0 H0.0 mm |
| Orientation | Sagittal |
| Phase enc. dir. | A >> P |
| Slice group | 2 |
| Position | Isocenter |
| Orientation | Transversal |

### Geometry - AutoAlign

| | |
|---|---|
| Phase enc. dir. | A >> P |
| Slice group | 3 |
| Position | Isocenter |
| Orientation | Coronal |
| Phase enc. dir. | R >> L |
| AutoAlign | --- |
| Initial Position | L0.0 A50.0 H0.0 |
| L | 0.0 mm |
| A | 50.0 mm |
| H | 0.0 mm |
| Initial Rotation | 0.00 deg |
| Initial Orientation | Sagittal |

### Geometry - Saturation

| | |
|---|---|
| Saturation mode | Standard |
| Fat suppr. | None |
| Water suppr. | None |
| Special sat. | None |

### Geometry - Tim Planning Suite

| | |
|---|---|
| Set-n-Go Protocol | Off |
| Table position | H |
| Table position | 0 mm |
| Inline Composing | Off |

### Geometry - Tim CT

| | |
|---|---|
| Tim CT mode | Off |
| Slices | 1 |
| Slice thickness | 5.0 mm |
| Dist. factor | 20 % |
| FoV read | 250 mm |
| FoV phase | 100.0 % |
| Segments | 1 |

### System - Miscellaneous

| | |
|---|---|
| Positioning mode | FIX |
| Table position | F |
| Table position | 0 mm |
| MSMA | S - C - T |
| Sagittal | R >> L |
| Coronal | A >> P |
| Transversal | F >> H |
| Coil Combine Mode | Adaptive Combine |
| Save uncombined | Off |
| Matrix Optimization | Off |
| AutoAlign | --- |
| Coil Select Mode | Off - AutoCoilSelect |

### System - Adjustments

| | |
|---|---|
| B0 Shim mode | Tune up |
| B1 Shim mode | TrueForm |
| Confirm freq. adjustment | Off |
| Assume Dominant Fat | Off |
| Assume Silicone | Off |
| Adjustment Tolerance | Auto |

### System - Adjust Volume

| | |
|---|---|
| Position | Isocenter |
| Orientation | Transversal |
| Rotation | 0.00 deg |
| A >> P | 263 mm |
| R >> L | 350 mm |
| F >> H | 350 mm |
| Reset | Off |

### System - Tx/Rx

| | |
|---|---|
| Frequency 1H | 297.207726 MHz |
| Correction factor | 1 |
| Gain | High |
| Img. Scale Cor. | 1.000 |
| Reset | Off |
| ? Ref. amplitude 1H | 0.000 V |

### Physio - Signal1

| | |
|---|---|
| 1st Signal/Mode | None |
| TR | 6.6 ms |
| Concatenations | 3 |
| Segments | 1 |

### Physio - Cardiac

| | |
|---|---|
| Tagging | None |
| Magn. preparation | None |
| Fat suppr. | None |
| Dark blood | Off |
| FoV read | 250 mm |
| FoV phase | 100.0 % |
| Phase resolution | 90 % |

### Physio - PACE

| | |
|---|---|
| Resp. control | Off |
| Concatenations | 3 |

### Inline - Common

| | |
|---|---|
| Subtract | Off |
| Measurements | 1 |
| StdDev | Off |
| Liver registration | Off |
| Save original images | On |

### Inline - MIP

| | |
|---|---|
| MIP-Sag | Off |
| MIP-Cor | Off |
| MIP-Tra | Off |
| MIP-Time | Off |
| Save original images | On |

### Inline - Soft Tissue

| | |
|---|---|
| Wash - In | Off |
| Wash - Out | Off |
| TTP | Off |
| PEI | Off |
| MIP - time | Off |
| Measurements | 1 |

### Inline - Composing

| | |
|---|---|
| Inline Composing | Off |
| Distortion Corr. | Off |

### Inline - MapIt

| | |
|---|---|
| Save original images | On |
| MapIt | None |
| Flip angle | 10 deg |
| Measurements | 1 |
| Contrasts | 1 |
| TR | 6.6 ms |
| TE | 2.67 ms |

### Sequence - Part 1

| | |
|---|---|
| Introduction | On |

### Sequence - Part 1

| | |
|---|---|
| Dimension | 2D |
| Phase stabilisation | Off |
| Asymmetric echo | Allowed |
| Contrasts | 1 |
| Flow comp. | No |
| Multi-slice mode | Sequential |
| Bandwidth | 320 Hz/Px |

### Sequence - Part 2

| | |
|---|---|
| Segments | 1 |
| Acoustic noise reduction | None |
| RF pulse type | Normal |
| Gradient mode | Normal |
| Excitation | Slice-sel. |
| RF spoiling | On |

### Sequence - Nuclei

| | |
|---|---|
| TX/RX Nucleus | 1H |
| TX/RX delta frequency | 0 Hz |
| TX Nucleus | None |
| TX delta frequency | 0 Hz |
| Coil elements | A32 |

### Sequence - Assistant

| | |
|---|---|
| Mode | Off |

**\\USER\PHCP\functional\final\AAHScout_32ch**

TA: 0:18 PM: REF Voxel size: 1.6×1.6×1.6 mmPAT: 2 Rel. SNR: 1.00 : fl

### Properties

| | |
|---|---|
| Prio recon | Off |
| Load images to viewer | On |
| Inline movie | Off |
| Auto store images | On |
| Load images to stamp segments | On |
| Load images to graphic segments | On |
| Auto open inline display | Off |
| Auto close inline display | Off |
| Start measurement without further preparation | Off |
| Wait for user to start | Off |
| Start measurements | Single measurement |

### Routine

| | |
|---|---|
| Slab group | 1 |
| Slabs | 1 |
| Dist. factor | 20 % |
| Position | L0.0 A50.0 H0.0 mm |
| Orientation | Sagittal |
| Phase enc. dir. | A >> P |
| Phase oversampling | 0 % |
| Slice oversampling | 0.0 % |
| Slices per slab | 128 |
| FoV read | 260 mm |
| FoV phase | 100.0 % |
| Slice thickness | 1.6 mm |
| TR | 3.25 ms |
| TE | 1.53 ms |
| Averages | 1 |
| Concatenations | 1 |
| Filter | None |
| Coil elements | A32 |

### Contrast - Common

| | |
|---|---|
| TR | 3.25 ms |
| TE | 1.53 ms |
| Flip angle | 16 deg |

### Contrast - Dynamic

| | |
|---|---|
| Averages | 1 |
| Averaging mode | Short term |
| Reconstruction | Magnitude |
| Measurements | 1 |

### Resolution - Common

| | |
|---|---|
| FoV read | 260 mm |
| FoV phase | 100.0 % |
| Slice thickness | 1.6 mm |
| Base resolution | 160 |
| Phase resolution | 100 % |
| Slice resolution | 69 % |
| Phase partial Fourier | 6/8 |
| Slice partial Fourier | 6/8 |
| Trajectory | Cartesian |

### Resolution - iPAT

| | |
|---|---|
| PAT mode | GRAPPA |
| Accel. factor PE | 2 |
| Ref. lines PE | 24 |
| Accel. factor 3D | 1 |

### Resolution - iPAT

| | |
|---|---|
| Reference scan mode | Integrated |

### Resolution - Filter Image

| | |
|---|---|
| Image Filter | Off |
| Distortion Corr. | Off |
| Prescan Normalize | Off |
| Normalize | Off |
| B1 filter | Off |

### Resolution - Filter Rawdata

| | |
|---|---|
| Raw filter | Off |
| Elliptical filter | Off |

### Geometry - Common

| | |
|---|---|
| Slab group | 1 |
| Slabs | 1 |
| Dist. factor | 20 % |
| Position | L0.0 A50.0 H0.0 mm |
| Orientation | Sagittal |
| Phase enc. dir. | A >> P |
| Slice oversampling | 0.0 % |
| Slices per slab | 128 |
| FoV read | 260 mm |
| FoV phase | 100.0 % |
| Slice thickness | 1.6 mm |
| TR | 3.25 ms |
| Multi-slice mode | Sequential |
| Series | Ascending |
| Concatenations | 1 |

### Geometry - AutoAlign

| | |
|---|---|
| Slab group | 1 |
| Position | L0.0 A50.0 H0.0 mm |
| Orientation | Sagittal |
| Phase enc. dir. | A >> P |
| Initial Position | Isocenter |
| L | 0.0 mm |
| P | 0.0 mm |
| H | 0.0 mm |
| Initial Rotation | 0.00 deg |
| Initial Orientation | Transversal |

### Geometry - Tim Planning Suite

| | |
|---|---|
| Set-n-Go Protocol | Off |
| Table position | H |
| Table position | 0 mm |
| Inline Composing | Off |

### System - Miscellaneous

| | |
|---|---|
| Positioning mode | REF |
| Table position | H |
| Table position | 0 mm |
| MSMA | S - C - T |
| Sagittal | R >> L |
| Coronal | A >> P |
| Transversal | F >> H |
| Coil Combine Mode | Adaptive Combine |
| Save uncombined | Off |
| Matrix Optimization | Off |
| Coil Select Mode | Off - AutoCoilSelect |

### System - Adjustments

| | |
|---|---|
| B0 Shim mode | Tune up |
| B1 Shim mode | TrueForm |
| Confirm freq. adjustment | Off |
| Assume Dominant Fat | Off |
| Assume Silicone | Off |
| Adjustment Tolerance | Auto |

### System - Adjust Volume

| | |
|---|---|
| ! Position | L0.0 A46.1 F28.8 mm |
| ! Orientation | T > C-0.7 |
| ! Rotation | 0.00 deg |
| ! A >> P | 263 mm |
| ! R >> L | 350 mm |
| ! F >> H | 350 mm |
| Reset | Off |

### System - Tx/Rx

| | |
|---|---|
| Frequency 1H | 297.207726 MHz |
| Correction factor | 1 |
| Gain | High |
| Img. Scale Cor. | 1.000 |
| Reset | Off |
| ? Ref. amplitude 1H | 0.000 V |

### Physio - PACE

| | |
|---|---|
| Resp. control | Off |
| Concatenations | 1 |

### Inline - Common

| | |
|---|---|
| Flip angle | 16 deg |
| Measurements | 1 |
| Time to center | 7.7 s |

### Inline - Inline

| | |
|---|---|
| Subtract | Off |
| Measurements | 1 |
| StdDev | Off |
| Save original images | On |

### Inline - MIP

| | |
|---|---|
| MIP-Sag | Off |
| MIP-Cor | Off |
| MIP-Tra | Off |
| MIP-Time | Off |
| Save original images | On |

### Inline - Composing

| | |
|---|---|
| Inline Composing | Off |
| Distortion Corr. | Off |

### Inline - MapIt

| | |
|---|---|
| Save original images | On |
| MapIt | None |
| Flip angle | 16 deg |
| Measurements | 1 |
| Contrasts | 1 |
| TR | 3.25 ms |
| TE | 1.53 ms |

### Sequence - Part 1

| | |
|---|---|
| Introduction | On |
| Dimension | 3D |
| Asymmetric echo | Weak |

### Sequence - Part 1

| | |
|---|---|
| Contrasts | 1 |
| Multi-slice mode | Sequential |
| Bandwidth | 540 Hz/Px |

### Sequence - Part 2

| | |
|---|---|
| RF pulse type | Fast |
| Gradient mode | Normal |
| Excitation | Non-sel. |
| RF spoiling | On |

### Sequence - Nuclei

| | |
|---|---|
| TX/RX Nucleus | 1H |
| TX/RX delta frequency | 0 Hz |
| TX Nucleus | None |
| TX delta frequency | 0 Hz |
| Coil elements | A32 |

### Sequence - Assistant

| | |
|---|---|
| Mode | Off |

| \\USER\PHCP\functional\final\Localizer_aligned |
|---|
| TA: 0:24 PM: FIX Voxel size: 0.5×0.5×5.0 mmPAT: Off Rel. SNR: 1.00 : fl |

### Properties

| | |
|---|---|
| Prio recon | Off |
| Load images to viewer | On |
| Inline movie | Off |
| Auto store images | On |
| Load images to stamp segments | On |
| Load images to graphic segments | On |
| Auto open inline display | Off |
| Auto close inline display | Off |
| Start measurement without further preparation | On |
| Wait for user to start | Off |
| Start measurements | Single measurement |

### Routine

| | |
|---|---|
| Slice group | 1 |
| Slices | 5 |
| Dist. factor | 400 % |
| Position | Isocenter |
| Orientation | Coronal |
| Phase enc. dir. | R >> L |
| Slice group | 2 |
| Slices | 5 |
| Dist. factor | 600 % |
| Position | Isocenter |
| Orientation | Transversal |
| Phase enc. dir. | L >> R |
| Slice group | 3 |
| Slices | 5 |
| Dist. factor | 300 % |
| Position | Isocenter |
| Orientation | Sagittal |
| Phase enc. dir. | A >> P |
| AutoAlign | Head > Brain |
| Phase oversampling | 0 % |
| FoV read | 250 mm |
| FoV phase | 100.0 % |
| Slice thickness | 5.0 mm |
| TR | 6.6 ms |
| TE | 2.67 ms |
| Averages | 1 |
| Concatenations | 15 |
| Filter | Elliptical filter |
| Coil elements | A32 |

### Contrast - Common

| | |
|---|---|
| TR | 6.6 ms |
| TE | 2.67 ms |
| TD | 0 ms |
| MTC | Off |
| Magn. preparation | None |
| Flip angle | 10 deg |
| Fat suppr. | None |
| Water suppr. | None |
| SWI | Off |

### Contrast - Dynamic

| | |
|---|---|
| Averages | 1 |
| Averaging mode | Short term |
| Reconstruction | Magnitude |
| Measurements | 1 |

### Contrast - Dynamic

| | |
|---|---|
| Multiple series | Each measurement |

### Resolution - Common

| | |
|---|---|
| FoV read | 250 mm |
| FoV phase | 100.0 % |
| Slice thickness | 5.0 mm |
| Base resolution | 256 |
| Phase resolution | 90 % |
| Phase partial Fourier | Off |
| Interpolation | On |

### Resolution - iPAT

| | |
|---|---|
| PAT mode | None |

### Resolution - Filter Image

| | |
|---|---|
| Image Filter | Off |
| Distortion Corr. | Off |
| Prescan Normalize | Off |
| Normalize | Off |
| B1 filter | Off |

### Resolution - Filter Rawdata

| | |
|---|---|
| Raw filter | Off |
| Elliptical filter | On |

### Geometry - Common

| | |
|---|---|
| Slice group | 1 |
| Slices | 5 |
| Dist. factor | 400 % |
| Position | Isocenter |
| Orientation | Coronal |
| Phase enc. dir. | R >> L |
| Slice group | 2 |
| Slices | 5 |
| Dist. factor | 600 % |
| Position | Isocenter |
| Orientation | Transversal |
| Phase enc. dir. | L >> R |
| Slice group | 3 |
| Slices | 5 |
| Dist. factor | 300 % |
| Position | Isocenter |
| Orientation | Sagittal |
| Phase enc. dir. | A >> P |
| FoV read | 250 mm |
| FoV phase | 100.0 % |
| Slice thickness | 5.0 mm |
| TR | 6.6 ms |
| Multi-slice mode | Sequential |
| Series | Interleaved |
| Concatenations | 15 |

### Geometry - AutoAlign

| | |
|---|---|
| Slice group | 1 |
| Position | Isocenter |
| Orientation | Coronal |
| Phase enc. dir. | R >> L |
| Slice group | 2 |
| Position | Isocenter |
| Orientation | Transversal |

### Geometry - AutoAlign

| | |
|---|---|
| Phase enc. dir. | L >> R |
| Slice group | 3 |
| Position | Isocenter |
| Orientation | Sagittal |
| Phase enc. dir. | A >> P |
| AutoAlign | Head > Brain |
| Initial Position | Isocenter |
| L | 0.0 mm |
| P | 0.0 mm |
| H | 0.0 mm |
| Initial Rotation | 0.00 deg |
| Initial Orientation | Coronal |

### Geometry - Saturation

| | |
|---|---|
| Saturation mode | Standard |
| Fat suppr. | None |
| Water suppr. | None |
| Special sat. | None |

### Geometry - Tim Planning Suite

| | |
|---|---|
| Set-n-Go Protocol | Off |
| Table position | H |
| Table position | 0 mm |
| Inline Composing | Off |

### Geometry - Tim CT

| | |
|---|---|
| Tim CT mode | Off |
| Slices | 5 |
| Slice thickness | 5.0 mm |
| Dist. factor | 300 % |
| FoV read | 250 mm |
| FoV phase | 100.0 % |
| Segments | 1 |

### System - Miscellaneous

| | |
|---|---|
| Positioning mode | FIX |
| Table position | F |
| Table position | 0 mm |
| MSMA | S - C - T |
| Sagittal | R >> L |
| Coronal | A >> P |
| Transversal | F >> H |
| Coil Combine Mode | Adaptive Combine |
| Save uncombined | Off |
| Matrix Optimization | Off |
| AutoAlign | Head > Brain |
| Coil Select Mode | Off - AutoCoilSelect |

### System - Adjustments

| | |
|---|---|
| B0 Shim mode | Tune up |
| B1 Shim mode | TrueForm |
| Confirm freq. adjustment | Off |
| Assume Dominant Fat | Off |
| Assume Silicone | Off |
| Adjustment Tolerance | Auto |

### System - Adjust Volume

| | |
|---|---|
| Position | Isocenter |
| Orientation | Transversal |
| Rotation | 0.00 deg |
| A >> P | 263 mm |
| R >> L | 350 mm |
| F >> H | 350 mm |
| Reset | Off |

### System - Tx/Rx

| | |
|---|---|
| Frequency 1H | 297.207726 MHz |
| Correction factor | 1 |
| Gain | High |
| Img. Scale Cor. | 1.000 |
| Reset | Off |
| ? Ref. amplitude 1H | 0.000 V |

### Physio - Signal1

| | |
|---|---|
| 1st Signal/Mode | None |
| TR | 6.6 ms |
| Concatenations | 15 |
| Segments | 1 |

### Physio - Cardiac

| | |
|---|---|
| Tagging | None |
| Magn. preparation | None |
| Fat suppr. | None |
| Dark blood | Off |
| FoV read | 250 mm |
| FoV phase | 100.0 % |
| Phase resolution | 90 % |

### Physio - PACE

| | |
|---|---|
| Resp. control | Off |
| Concatenations | 15 |

### Inline - Common

| | |
|---|---|
| Subtract | Off |
| Measurements | 1 |
| StdDev | Off |
| Liver registration | Off |
| Save original images | On |

### Inline - MIP

| | |
|---|---|
| MIP-Sag | Off |
| MIP-Cor | Off |
| MIP-Tra | Off |
| MIP-Time | Off |
| Save original images | On |

### Inline - Soft Tissue

| | |
|---|---|
| Wash - In | Off |
| Wash - Out | Off |
| TTP | Off |
| PEI | Off |
| MIP - time | Off |
| Measurements | 1 |

### Inline - Composing

| | |
|---|---|
| Inline Composing | Off |
| Distortion Corr. | Off |

### Inline - MapIt

| | |
|---|---|
| Save original images | On |
| MapIt | None |
| Flip angle | 10 deg |
| Measurements | 1 |
| Contrasts | 1 |
| TR | 6.6 ms |
| TE | 2.67 ms |

### Sequence - Part 1

| | |
|---|---|
| Introduction | On |

### Sequence - Part 1

| | |
|---|---|
| Dimension | 2D |
| Phase stabilisation | Off |
| Asymmetric echo | Allowed |
| Contrasts | 1 |
| Flow comp. | No |
| Multi-slice mode | Sequential |
| Bandwidth | 320 Hz/Px |

### Sequence - Part 2

| | |
|---|---|
| Segments | 1 |
| Acoustic noise reduction | None |
| RF pulse type | Normal |
| Gradient mode | Normal |
| Excitation | Slice-sel. |
| RF spoiling | On |

### Sequence - Nuclei

| | |
|---|---|
| TX/RX Nucleus | 1H |
| TX/RX delta frequency | 0 Hz |
| TX Nucleus | None |
| TX delta frequency | 0 Hz |
| Coil elements | A32 |

### Sequence - Assistant

| | |
|---|---|
| Mode | Off |

**\\USER\PHCP\functional\final\BOLD_oppPE_AP**

TA: 1:22 PM: FIX Voxel size: 1.6×1.6×1.6 mmPAT: 2 Rel. SNR: 1.00 : epfid

### Properties

| | |
|---|---|
| Prio recon | Off |
| Load images to viewer | Off |
| Inline movie | Off |
| Auto store images | On |
| Load images to stamp segments | Off |
| Load images to graphic segments | Off |
| Auto open inline display | On |
| Auto close inline display | Off |
| Start measurement without further preparation | Off |
| Wait for user to start | On |
| Start measurements | Single measurement |

### Routine

| | |
|---|---|
| Slice group | 1 |
| Slices | 85 |
| Dist. factor | 0 % |
| Position | L0.0 P13.6 H7.5 mm |
| Orientation | T > C-20.0 |
| Phase enc. dir. | A >> P |
| AutoAlign | Head > Brain |
| Phase oversampling | 0 % |
| FoV read | 208 mm |
| FoV phase | 100.0 % |
| Slice thickness | 1.60 mm |
| TR | 1000 ms |
| TE | 22.20 ms |
| Multi-band accel. factor | 5 |
| Filter | None |
| Coil elements | A32 |

### Contrast - Common

| | |
|---|---|
| TR | 1000 ms |
| TE | 22.20 ms |
| MTC | Off |
| Magn. preparation | None |
| Flip angle | 45 deg |
| Fat suppr. | Fat sat. |

### Contrast - Dynamic

| | |
|---|---|
| Averaging mode | Long term |
| Reconstruction | Magnitude |
| Measurements | 3 |
| Delay in TR | 0 ms |
| Multiple series | Off |

### Resolution - Common

| | |
|---|---|
| FoV read | 208 mm |
| FoV phase | 100.0 % |
| Slice thickness | 1.60 mm |
| Base resolution | 130 |
| Phase resolution | 100 % |
| Phase partial Fourier | 7/8 |
| Interpolation | Off |

### Resolution - iPAT

| | |
|---|---|
| PAT mode | GRAPPA |
| Accel. factor PE | 2 |
| Ref. lines PE | 96 |
| Reference scan mode | GRE |

### Resolution - Filter Image

| | |
|---|---|
| Distortion Corr. | Off |
| Prescan Normalize | Off |

### Resolution - Filter Rawdata

| | |
|---|---|
| Raw filter | Off |
| Elliptical filter | Off |
| Hamming | Off |

### Geometry - Common

| | |
|---|---|
| Slice group | 1 |
| Slices | 85 |
| Dist. factor | 0 % |
| Position | L0.0 P13.6 H7.5 mm |
| Orientation | T > C-20.0 |
| Phase enc. dir. | A >> P |
| FoV read | 208 mm |
| FoV phase | 100.0 % |
| Slice thickness | 1.60 mm |
| TR | 1000 ms |
| Multi-slice mode | Interleaved |
| Series | Interleaved |
| Multi-band accel. factor | 5 |

### Geometry - AutoAlign

| | |
|---|---|
| Slice group | 1 |
| Position | L0.0 P13.6 H7.5 mm |
| Orientation | T > C-20.0 |
| Phase enc. dir. | A >> P |
| AutoAlign | Head > Brain |
| Initial Position | L0.0 P13.6 H7.5 |
| L | 0.0 mm |
| P | 13.6 mm |
| H | 7.5 mm |
| Initial Rotation | -0.52 deg |
| Initial Orientation | T > C |
| T > C | -20.0 |
| > S | 0.0 |

### Geometry - Saturation

| | |
|---|---|
| Fat suppr. | Fat sat. |
| Special sat. | None |

### Geometry - Tim Planning Suite

| | |
|---|---|
| Set-n-Go Protocol | Off |
| Table position | H |
| Table position | 0 mm |
| Inline Composing | Off |

### System - Miscellaneous

| | |
|---|---|
| Positioning mode | FIX |
| Table position | H |
| Table position | 0 mm |
| MSMA | S - C - T |
| Sagittal | R >> L |
| Coronal | A >> P |
| Transversal | F >> H |
| Coil Combine Mode | Sum of Squares |
| Matrix Optimization | Off |
| AutoAlign | Head > Brain |
| Coil Select Mode | Off - AutoCoilSelect |

### System - Adjustments

| | |
|---|---|
| B0 Shim mode | Advanced |
| B1 Shim mode | TrueForm |
| Confirm freq. adjustment | Off |
| Assume Dominant Fat | Off |
| Assume Silicone | Off |
| Adjustment Tolerance | Auto |

### System - Adjust Volume

| | |
|---|---|
| ! Position | L0.7 P13.2 H9.5 mm |
| ! Orientation | T > C-15.6 > S0.3 |
| ! Rotation | 0.00 deg |
| ! A >> P | 170 mm |
| ! R >> L | 130 mm |
| ! F >> H | 120 mm |
| Reset | Off |

### System - Tx/Rx

| | |
|---|---|
| Frequency 1H | 297.207726 MHz |
| Correction factor | 1 |
| Gain | High |
| Img. Scale Cor. | 1.000 |
| Reset | Off |
| ? Ref. amplitude 1H | 0.000 V |

### Physio - Signal1

| | |
|---|---|
| 1st Signal/Mode | None |
| TR | 1000 ms |
| Multi-band accel. factor | 5 |

### BOLD

| | |
|---|---|
| GLM Statistics | Off |
| Dynamic t-maps | Off |
| Ignore meas. at start | 0 |
| Ignore after transition | 0 |
| Model transition states | On |
| Temp. highpass filter | On |
| Threshold | 4.00 |
| Paradigm size | 20 |
| Meas[1] | Baseline |
| Meas[2] | Baseline |
| Meas[3] | Baseline |
| Meas[4] | Baseline |
| Meas[5] | Baseline |
| Meas[6] | Baseline |
| Meas[7] | Baseline |
| Meas[8] | Baseline |
| Meas[9] | Baseline |
| Meas[10] | Baseline |
| Meas[11] | Active |
| Meas[12] | Active |
| Meas[13] | Active |
| Meas[14] | Active |
| Meas[15] | Active |
| Meas[16] | Active |
| Meas[17] | Active |
| Meas[18] | Active |
| Meas[19] | Active |
| Meas[20] | Active |
| Motion correction | Off |
| Spatial filter | Off |
| Measurements | 3 |
| Delay in TR | 0 ms |
| Multiple series | Off |

### Sequence - Part 1

| | |
|---|---|
| Introduction | Off |
| Contrasts | 1 |
| Flow comp. | No |
| Multi-slice mode | Interleaved |
| Free echo spacing | Off |
| Echo spacing | 0.64 ms |
| Bandwidth | 1924 Hz/Px |

### Sequence - Part 2

| | |
|---|---|
| EPI factor | 130 |
| Gradient mode | Normal |
| RF spoiling | Off |

### Sequence - Special

| | |
|---|---|
| Excite pulse duration | 5760 us |
| Single-band images | On |
| MB LeakBlock kernel | Off |
| MB dual kernel | Off |
| MB RF phase scramble | On |
| SENSE1 coil combine | Off |
| Invert RO/PE polarity | On |
| PF omits higher k-space | Off |
| Disable freq. update | Off |
| Force equal slice timing | Off |
| Online multi-band recon. | Online |
| FFT scale factor | 0.60 |
| Fat saturation FA | 110.0 deg |
| GRE iPAT ref. FA | 12.0 deg |
| Physio recording | Off |
| Triggering scheme | Standard |

| \\USER\PHCP\functional\final\BOLD_PRF1_AP |
|---|
| TA: 6:43 PM: FIX Voxel size: 1.6×1.6×1.6 mmPAT: 2 Rel. SNR: 1.00 : epfid |

### Properties

| | |
|---|---|
| Prio recon | Off |
| Load images to viewer | Off |
| Inline movie | Off |
| Auto store images | On |
| Load images to stamp segments | Off |
| Load images to graphic segments | Off |
| Auto open inline display | On |
| Auto close inline display | Off |
| Start measurement without further preparation | Off |
| Wait for user to start | On |
| Start measurements | Single measurement |

### Routine

| | |
|---|---|
| Slice group | 1 |
| Slices | 85 |
| Dist. factor | 0 % |
| Position | L0.0 P13.6 H7.5 mm |
| Orientation | T > C-20.0 |
| Phase enc. dir. | A >> P |
| AutoAlign | Head > Brain |
| Phase oversampling | 0 % |
| FoV read | 208 mm |
| FoV phase | 100.0 % |
| Slice thickness | 1.60 mm |
| TR | 1000 ms |
| TE | 22.20 ms |
| Multi-band accel. factor | 5 |
| Filter | None |
| Coil elements | A32 |

### Contrast - Common

| | |
|---|---|
| TR | 1000 ms |
| TE | 22.20 ms |
| MTC | Off |
| Magn. preparation | None |
| Flip angle | 45 deg |
| Fat suppr. | Fat sat. |

### Contrast - Dynamic

| | |
|---|---|
| Averaging mode | Long term |
| Reconstruction | Magnitude |
| Measurements | 324 |
| Delay in TR | 0 ms |
| Multiple series | Off |

### Resolution - Common

| | |
|---|---|
| FoV read | 208 mm |
| FoV phase | 100.0 % |
| Slice thickness | 1.60 mm |
| Base resolution | 130 |
| Phase resolution | 100 % |
| Phase partial Fourier | 7/8 |
| Interpolation | Off |

### Resolution - iPAT

| | |
|---|---|
| PAT mode | GRAPPA |
| Accel. factor PE | 2 |
| Ref. lines PE | 96 |
| Reference scan mode | GRE |

### Resolution - Filter Image

| | |
|---|---|
| Distortion Corr. | Off |
| Prescan Normalize | Off |

### Resolution - Filter Rawdata

| | |
|---|---|
| Raw filter | Off |
| Elliptical filter | Off |
| Hamming | Off |

### Geometry - Common

| | |
|---|---|
| Slice group | 1 |
| Slices | 85 |
| Dist. factor | 0 % |
| Position | L0.0 P13.6 H7.5 mm |
| Orientation | T > C-20.0 |
| Phase enc. dir. | A >> P |
| FoV read | 208 mm |
| FoV phase | 100.0 % |
| Slice thickness | 1.60 mm |
| TR | 1000 ms |
| Multi-slice mode | Interleaved |
| Series | Interleaved |
| Multi-band accel. factor | 5 |

### Geometry - AutoAlign

| | |
|---|---|
| Slice group | 1 |
| Position | L0.0 P13.6 H7.5 mm |
| Orientation | T > C-20.0 |
| Phase enc. dir. | A >> P |
| AutoAlign | Head > Brain |
| Initial Position | L0.0 P13.6 H7.5 |
| L | 0.0 mm |
| P | 13.6 mm |
| H | 7.5 mm |
| Initial Rotation | -0.52 deg |
| Initial Orientation | T > C |
| T > C | -20.0 |
| > S | 0.0 |

### Geometry - Saturation

| | |
|---|---|
| Fat suppr. | Fat sat. |
| Special sat. | None |

### Geometry - Tim Planning Suite

| | |
|---|---|
| Set-n-Go Protocol | Off |
| Table position | H |
| Table position | 0 mm |
| Inline Composing | Off |

### System - Miscellaneous

| | |
|---|---|
| Positioning mode | FIX |
| Table position | H |
| Table position | 0 mm |
| MSMA | S - C - T |
| Sagittal | R >> L |
| Coronal | A >> P |
| Transversal | F >> H |
| Coil Combine Mode | Sum of Squares |
| Matrix Optimization | Off |
| AutoAlign | Head > Brain |
| Coil Select Mode | Off - AutoCoilSelect |

### System - Adjustments

| | |
|---|---|
| B0 Shim mode | Advanced |
| B1 Shim mode | TrueForm |
| Confirm freq. adjustment | Off |
| Assume Dominant Fat | Off |
| Assume Silicone | Off |
| Adjustment Tolerance | Auto |

### System - Adjust Volume

| | |
|---|---|
| ! Position | L0.7 P13.2 H9.5 mm |
| ! Orientation | T > C-15.6 > S0.3 |
| ! Rotation | 0.00 deg |
| ! A >> P | 170 mm |
| ! R >> L | 130 mm |
| ! F >> H | 120 mm |
| Reset | Off |

### System - Tx/Rx

| | |
|---|---|
| Frequency 1H | 297.207726 MHz |
| Correction factor | 1 |
| Gain | High |
| Img. Scale Cor. | 1.000 |
| Reset | Off |
| ? Ref. amplitude 1H | 0.000 V |

### Physio - Signal1

| | |
|---|---|
| 1st Signal/Mode | None |
| TR | 1000 ms |
| Multi-band accel. factor | 5 |

### BOLD

| | |
|---|---|
| GLM Statistics | Off |
| Dynamic t-maps | Off |
| Ignore meas. at start | 0 |
| Ignore after transition | 0 |
| Model transition states | On |
| Temp. highpass filter | On |
| Threshold | 4.00 |
| Paradigm size | 20 |
| Meas[1] | Baseline |
| Meas[2] | Baseline |
| Meas[3] | Baseline |
| Meas[4] | Baseline |
| Meas[5] | Baseline |
| Meas[6] | Baseline |
| Meas[7] | Baseline |
| Meas[8] | Baseline |
| Meas[9] | Baseline |
| Meas[10] | Baseline |
| Meas[11] | Active |
| Meas[12] | Active |
| Meas[13] | Active |
| Meas[14] | Active |
| Meas[15] | Active |
| Meas[16] | Active |
| Meas[17] | Active |
| Meas[18] | Active |
| Meas[19] | Active |
| Meas[20] | Active |
| Motion correction | Off |
| Spatial filter | Off |
| Measurements | 324 |
| Delay in TR | 0 ms |
| Multiple series | Off |

### Sequence - Part 1

| | |
|---|---|
| Introduction | Off |
| Contrasts | 1 |
| Flow comp. | No |
| Multi-slice mode | Interleaved |
| Free echo spacing | Off |
| Echo spacing | 0.64 ms |
| Bandwidth | 1924 Hz/Px |

### Sequence - Part 2

| | |
|---|---|
| EPI factor | 130 |
| Gradient mode | Normal |
| RF spoiling | Off |

### Sequence - Special

| | |
|---|---|
| Excite pulse duration | 5760 us |
| Single-band images | On |
| MB LeakBlock kernel | Off |
| MB dual kernel | Off |
| MB RF phase scramble | On |
| SENSE1 coil combine | Off |
| Invert RO/PE polarity | Off |
| PF omits higher k-space | Off |
| Disable freq. update | Off |
| Force equal slice timing | Off |
| Online multi-band recon. | Online |
| FFT scale factor | 0.60 |
| Fat saturation FA | 110.0 deg |
| GRE iPAT ref. FA | 12.0 deg |
| Physio recording | Off |
| Triggering scheme | Standard |

**\\USER\PHCP\functional\final\BOLD_CSS_task1_AP**

TA: 6:16 PM: FIX Voxel size: 1.6×1.6×1.6 mmPAT: 2 Rel. SNR: 1.00 : epfid

### Properties

| | |
|---|---|
| Prio recon | Off |
| Load images to viewer | Off |
| Inline movie | Off |
| Auto store images | On |
| Load images to stamp segments | Off |
| Load images to graphic segments | Off |
| Auto open inline display | On |
| Auto close inline display | Off |
| Start measurement without further preparation | Off |
| Wait for user to start | On |
| Start measurements | Single measurement |

### Routine

| | |
|---|---|
| Slice group | 1 |
| Slices | 85 |
| Dist. factor | 0 % |
| Position | L0.0 P13.6 H7.5 mm |
| Orientation | T > C-20.0 |
| Phase enc. dir. | A >> P |
| AutoAlign | Head > Brain |
| Phase oversampling | 0 % |
| FoV read | 208 mm |
| FoV phase | 100.0 % |
| Slice thickness | 1.60 mm |
| TR | 1000 ms |
| TE | 22.20 ms |
| Multi-band accel. factor | 5 |
| Filter | None |
| Coil elements | A32 |

### Contrast - Common

| | |
|---|---|
| TR | 1000 ms |
| TE | 22.20 ms |
| MTC | Off |
| Magn. preparation | None |
| Flip angle | 45 deg |
| Fat suppr. | Fat sat. |

### Contrast - Dynamic

| | |
|---|---|
| Averaging mode | Long term |
| Reconstruction | Magnitude |
| Measurements | 297 |
| Delay in TR | 0 ms |
| Multiple series | Off |

### Resolution - Common

| | |
|---|---|
| FoV read | 208 mm |
| FoV phase | 100.0 % |
| Slice thickness | 1.60 mm |
| Base resolution | 130 |
| Phase resolution | 100 % |
| Phase partial Fourier | 7/8 |
| Interpolation | Off |

### Resolution - iPAT

| | |
|---|---|
| PAT mode | GRAPPA |
| Accel. factor PE | 2 |
| Ref. lines PE | 96 |
| Reference scan mode | GRE |

### Resolution - Filter Image

| | |
|---|---|
| Distortion Corr. | Off |
| Prescan Normalize | Off |

### Resolution - Filter Rawdata

| | |
|---|---|
| Raw filter | Off |
| Elliptical filter | Off |
| Hamming | Off |

### Geometry - Common

| | |
|---|---|
| Slice group | 1 |
| Slices | 85 |
| Dist. factor | 0 % |
| Position | L0.0 P13.6 H7.5 mm |
| Orientation | T > C-20.0 |
| Phase enc. dir. | A >> P |
| FoV read | 208 mm |
| FoV phase | 100.0 % |
| Slice thickness | 1.60 mm |
| TR | 1000 ms |
| Multi-slice mode | Interleaved |
| Series | Interleaved |
| Multi-band accel. factor | 5 |

### Geometry - AutoAlign

| | |
|---|---|
| Slice group | 1 |
| Position | L0.0 P13.6 H7.5 mm |
| Orientation | T > C-20.0 |
| Phase enc. dir. | A >> P |
| AutoAlign | Head > Brain |
| Initial Position | L0.0 P13.6 H7.5 |
| L | 0.0 mm |
| P | 13.6 mm |
| H | 7.5 mm |
| Initial Rotation | -0.52 deg |
| Initial Orientation | T > C |
| T > C | -20.0 |
| > S | 0.0 |

### Geometry - Saturation

| | |
|---|---|
| Fat suppr. | Fat sat. |
| Special sat. | None |

### Geometry - Tim Planning Suite

| | |
|---|---|
| Set-n-Go Protocol | Off |
| Table position | H |
| Table position | 0 mm |
| Inline Composing | Off |

### System - Miscellaneous

| | |
|---|---|
| Positioning mode | FIX |
| Table position | H |
| Table position | 0 mm |
| MSMA | S - C - T |
| Sagittal | R >> L |
| Coronal | A >> P |
| Transversal | F >> H |
| Coil Combine Mode | Sum of Squares |
| Matrix Optimization | Off |
| AutoAlign | Head > Brain |
| Coil Select Mode | Off - AutoCoilSelect |

### System - Adjustments

| | |
|---|---|
| B0 Shim mode | Advanced |
| B1 Shim mode | TrueForm |
| Confirm freq. adjustment | Off |
| Assume Dominant Fat | Off |
| Assume Silicone | Off |
| Adjustment Tolerance | Auto |

### System - Adjust Volume

| | |
|---|---|
| ! Position | L0.7 P13.2 H9.5 mm |
| ! Orientation | T > C-15.6 > S0.3 |
| ! Rotation | 0.00 deg |
| ! A >> P | 170 mm |
| ! R >> L | 130 mm |
| ! F >> H | 120 mm |
| Reset | Off |

### System - Tx/Rx

| | |
|---|---|
| Frequency 1H | 297.207726 MHz |
| Correction factor | 1 |
| Gain | High |
| Img. Scale Cor. | 1.000 |
| Reset | Off |
| ? Ref. amplitude 1H | 0.000 V |

### Physio - Signal1

| | |
|---|---|
| 1st Signal/Mode | None |
| TR | 1000 ms |
| Multi-band accel. factor | 5 |

### BOLD

| | |
|---|---|
| GLM Statistics | Off |
| Dynamic t-maps | Off |
| Ignore meas. at start | 0 |
| Ignore after transition | 0 |
| Model transition states | On |
| Temp. highpass filter | On |
| Threshold | 4.00 |
| Paradigm size | 20 |
| Meas[1] | Baseline |
| Meas[2] | Baseline |
| Meas[3] | Baseline |
| Meas[4] | Baseline |
| Meas[5] | Baseline |
| Meas[6] | Baseline |
| Meas[7] | Baseline |
| Meas[8] | Baseline |
| Meas[9] | Baseline |
| Meas[10] | Baseline |
| Meas[11] | Active |
| Meas[12] | Active |
| Meas[13] | Active |
| Meas[14] | Active |
| Meas[15] | Active |
| Meas[16] | Active |
| Meas[17] | Active |
| Meas[18] | Active |
| Meas[19] | Active |
| Meas[20] | Active |
| Motion correction | Off |
| Spatial filter | Off |
| Measurements | 297 |
| Delay in TR | 0 ms |
| Multiple series | Off |

### Sequence - Part 1

| | |
|---|---|
| Introduction | Off |
| Contrasts | 1 |
| Flow comp. | No |
| Multi-slice mode | Interleaved |
| Free echo spacing | Off |
| Echo spacing | 0.64 ms |
| Bandwidth | 1924 Hz/Px |

### Sequence - Part 2

| | |
|---|---|
| EPI factor | 130 |
| Gradient mode | Normal |
| RF spoiling | Off |

### Sequence - Special

| | |
|---|---|
| Excite pulse duration | 5760 us |
| Single-band images | On |
| MB LeakBlock kernel | Off |
| MB dual kernel | Off |
| MB RF phase scramble | On |
| SENSE1 coil combine | Off |
| Invert RO/PE polarity | Off |
| PF omits higher k-space | Off |
| Disable freq. update | Off |
| Force equal slice timing | Off |
| Online multi-band recon. | Online |
| FFT scale factor | 0.60 |
| Fat saturation FA | 110.0 deg |
| GRE iPAT ref. FA | 12.0 deg |
| Physio recording | Off |
| Triggering scheme | Standard |

**\\USER\PHCP\functional\final\BOLD_CSS_task2_AP**

TA: 6:16 PM: FIX Voxel size: 1.6×1.6×1.6 mmPAT: 2 Rel. SNR: 1.00 : epfid

### Properties

| | |
|---|---|
| Prio recon | Off |
| Load images to viewer | Off |
| Inline movie | Off |
| Auto store images | On |
| Load images to stamp segments | Off |
| Load images to graphic segments | Off |
| Auto open inline display | On |
| Auto close inline display | Off |
| Start measurement without further preparation | Off |
| Wait for user to start | On |
| Start measurements | Single measurement |

### Routine

| | |
|---|---|
| Slice group | 1 |
| Slices | 85 |
| Dist. factor | 0 % |
| Position | L0.0 P13.6 H7.5 mm |
| Orientation | T > C-20.0 |
| Phase enc. dir. | A >> P |
| AutoAlign | Head > Brain |
| Phase oversampling | 0 % |
| FoV read | 208 mm |
| FoV phase | 100.0 % |
| Slice thickness | 1.60 mm |
| TR | 1000 ms |
| TE | 22.20 ms |
| Multi-band accel. factor | 5 |
| Filter | None |
| Coil elements | A32 |

### Contrast - Common

| | |
|---|---|
| TR | 1000 ms |
| TE | 22.20 ms |
| MTC | Off |
| Magn. preparation | None |
| Flip angle | 45 deg |
| Fat suppr. | Fat sat. |

### Contrast - Dynamic

| | |
|---|---|
| Averaging mode | Long term |
| Reconstruction | Magnitude |
| Measurements | 297 |
| Delay in TR | 0 ms |
| Multiple series | Off |

### Resolution - Common

| | |
|---|---|
| FoV read | 208 mm |
| FoV phase | 100.0 % |
| Slice thickness | 1.60 mm |
| Base resolution | 130 |
| Phase resolution | 100 % |
| Phase partial Fourier | 7/8 |
| Interpolation | Off |

### Resolution - iPAT

| | |
|---|---|
| PAT mode | GRAPPA |
| Accel. factor PE | 2 |
| Ref. lines PE | 96 |
| Reference scan mode | GRE |

### Resolution - Filter Image

| | |
|---|---|
| Distortion Corr. | Off |
| Prescan Normalize | Off |

### Resolution - Filter Rawdata

| | |
|---|---|
| Raw filter | Off |
| Elliptical filter | Off |
| Hamming | Off |

### Geometry - Common

| | |
|---|---|
| Slice group | 1 |
| Slices | 85 |
| Dist. factor | 0 % |
| Position | L0.0 P13.6 H7.5 mm |
| Orientation | T > C-20.0 |
| Phase enc. dir. | A >> P |
| FoV read | 208 mm |
| FoV phase | 100.0 % |
| Slice thickness | 1.60 mm |
| TR | 1000 ms |
| Multi-slice mode | Interleaved |
| Series | Interleaved |
| Multi-band accel. factor | 5 |

### Geometry - AutoAlign

| | |
|---|---|
| Slice group | 1 |
| Position | L0.0 P13.6 H7.5 mm |
| Orientation | T > C-20.0 |
| Phase enc. dir. | A >> P |
| AutoAlign | Head > Brain |
| Initial Position | L0.0 P13.6 H7.5 |
| L | 0.0 mm |
| P | 13.6 mm |
| H | 7.5 mm |
| Initial Rotation | -0.52 deg |
| Initial Orientation | T > C |
| T > C | -20.0 |
| > S | 0.0 |

### Geometry - Saturation

| | |
|---|---|
| Fat suppr. | Fat sat. |
| Special sat. | None |

### Geometry - Tim Planning Suite

| | |
|---|---|
| Set-n-Go Protocol | Off |
| Table position | H |
| Table position | 0 mm |
| Inline Composing | Off |

### System - Miscellaneous

| | |
|---|---|
| Positioning mode | FIX |
| Table position | H |
| Table position | 0 mm |
| MSMA | S - C - T |
| Sagittal | R >> L |
| Coronal | A >> P |
| Transversal | F >> H |
| Coil Combine Mode | Sum of Squares |
| Matrix Optimization | Off |
| AutoAlign | Head > Brain |
| Coil Select Mode | Off - AutoCoilSelect |

### System - Adjustments

| | |
|---|---|
| B0 Shim mode | Advanced |
| B1 Shim mode | TrueForm |
| Confirm freq. adjustment | Off |
| Assume Dominant Fat | Off |
| Assume Silicone | Off |
| Adjustment Tolerance | Auto |

### System - Adjust Volume

| | |
|---|---|
| ! Position | L0.7 P13.2 H9.5 mm |
| ! Orientation | T > C-15.6 > S0.3 |
| ! Rotation | 0.00 deg |
| ! A >> P | 170 mm |
| ! R >> L | 130 mm |
| ! F >> H | 120 mm |
| Reset | Off |

### System - Tx/Rx

| | |
|---|---|
| Frequency 1H | 297.207726 MHz |
| Correction factor | 1 |
| Gain | High |
| Img. Scale Cor. | 1.000 |
| Reset | Off |
| ? Ref. amplitude 1H | 0.000 V |

### Physio - Signal1

| | |
|---|---|
| 1st Signal/Mode | None |
| TR | 1000 ms |
| Multi-band accel. factor | 5 |

### BOLD

| | |
|---|---|
| GLM Statistics | Off |
| Dynamic t-maps | Off |
| Ignore meas. at start | 0 |
| Ignore after transition | 0 |
| Model transition states | On |
| Temp. highpass filter | On |
| Threshold | 4.00 |
| Paradigm size | 20 |
| Meas[1] | Baseline |
| Meas[2] | Baseline |
| Meas[3] | Baseline |
| Meas[4] | Baseline |
| Meas[5] | Baseline |
| Meas[6] | Baseline |
| Meas[7] | Baseline |
| Meas[8] | Baseline |
| Meas[9] | Baseline |
| Meas[10] | Baseline |
| Meas[11] | Active |
| Meas[12] | Active |
| Meas[13] | Active |
| Meas[14] | Active |
| Meas[15] | Active |
| Meas[16] | Active |
| Meas[17] | Active |
| Meas[18] | Active |
| Meas[19] | Active |
| Meas[20] | Active |
| Motion correction | Off |
| Spatial filter | Off |
| Measurements | 297 |
| Delay in TR | 0 ms |
| Multiple series | Off |

### Sequence - Part 1

| | |
|---|---|
| Introduction | Off |
| Contrasts | 1 |
| Flow comp. | No |
| Multi-slice mode | Interleaved |
| Free echo spacing | Off |
| Echo spacing | 0.64 ms |
| Bandwidth | 1924 Hz/Px |

### Sequence - Part 2

| | |
|---|---|
| EPI factor | 130 |
| Gradient mode | Normal |
| RF spoiling | Off |

### Sequence - Special

| | |
|---|---|
| Excite pulse duration | 5760 us |
| Single-band images | On |
| MB LeakBlock kernel | Off |
| MB dual kernel | Off |
| MB RF phase scramble | On |
| SENSE1 coil combine | Off |
| Invert RO/PE polarity | Off |
| PF omits higher k-space | Off |
| Disable freq. update | Off |
| Force equal slice timing | Off |
| Online multi-band recon. | Online |
| FFT scale factor | 0.60 |
| Fat saturation FA | 110.0 deg |
| GRE iPAT ref. FA | 12.0 deg |
| Physio recording | Off |
| Triggering scheme | Standard |

| \\USER\PHCP\functional\final\BOLD_CSS_task3_AP |
|---|
| TA: 6:16 PM: FIX Voxel size: 1.6×1.6×1.6 mmPAT: 2 Rel. SNR: 1.00 : epfid |

### Properties

| | |
|---|---|
| Prio recon | Off |
| Load images to viewer | Off |
| Inline movie | Off |
| Auto store images | On |
| Load images to stamp segments | Off |
| Load images to graphic segments | Off |
| Auto open inline display | On |
| Auto close inline display | Off |
| Start measurement without further preparation | Off |
| Wait for user to start | On |
| Start measurements | Single measurement |

### Routine

| | |
|---|---|
| Slice group | 1 |
| Slices | 85 |
| Dist. factor | 0 % |
| Position | L0.0 P13.6 H7.5 mm |
| Orientation | T > C-20.0 |
| Phase enc. dir. | A >> P |
| AutoAlign | Head > Brain |
| Phase oversampling | 0 % |
| FoV read | 208 mm |
| FoV phase | 100.0 % |
| Slice thickness | 1.60 mm |
| TR | 1000 ms |
| TE | 22.20 ms |
| Multi-band accel. factor | 5 |
| Filter | None |
| Coil elements | A32 |

### Contrast - Common

| | |
|---|---|
| TR | 1000 ms |
| TE | 22.20 ms |
| MTC | Off |
| Magn. preparation | None |
| Flip angle | 45 deg |
| Fat suppr. | Fat sat. |

### Contrast - Dynamic

| | |
|---|---|
| Averaging mode | Long term |
| Reconstruction | Magnitude |
| Measurements | 297 |
| Delay in TR | 0 ms |
| Multiple series | Off |

### Resolution - Common

| | |
|---|---|
| FoV read | 208 mm |
| FoV phase | 100.0 % |
| Slice thickness | 1.60 mm |
| Base resolution | 130 |
| Phase resolution | 100 % |
| Phase partial Fourier | 7/8 |
| Interpolation | Off |

### Resolution - iPAT

| | |
|---|---|
| PAT mode | GRAPPA |
| Accel. factor PE | 2 |
| Ref. lines PE | 96 |
| Reference scan mode | GRE |

### Resolution - Filter Image

| | |
|---|---|
| Distortion Corr. | Off |
| Prescan Normalize | Off |

### Resolution - Filter Rawdata

| | |
|---|---|
| Raw filter | Off |
| Elliptical filter | Off |
| Hamming | Off |

### Geometry - Common

| | |
|---|---|
| Slice group | 1 |
| Slices | 85 |
| Dist. factor | 0 % |
| Position | L0.0 P13.6 H7.5 mm |
| Orientation | T > C-20.0 |
| Phase enc. dir. | A >> P |
| FoV read | 208 mm |
| FoV phase | 100.0 % |
| Slice thickness | 1.60 mm |
| TR | 1000 ms |
| Multi-slice mode | Interleaved |
| Series | Interleaved |
| Multi-band accel. factor | 5 |

### Geometry - AutoAlign

| | |
|---|---|
| Slice group | 1 |
| Position | L0.0 P13.6 H7.5 mm |
| Orientation | T > C-20.0 |
| Phase enc. dir. | A >> P |
| AutoAlign | Head > Brain |
| Initial Position | L0.0 P13.6 H7.5 |
| L | 0.0 mm |
| P | 13.6 mm |
| H | 7.5 mm |
| Initial Rotation | -0.52 deg |
| Initial Orientation | T > C |
| T > C | -20.0 |
| > S | 0.0 |

### Geometry - Saturation

| | |
|---|---|
| Fat suppr. | Fat sat. |
| Special sat. | None |

### Geometry - Tim Planning Suite

| | |
|---|---|
| Set-n-Go Protocol | Off |
| Table position | H |
| Table position | 0 mm |
| Inline Composing | Off |

### System - Miscellaneous

| | |
|---|---|
| Positioning mode | FIX |
| Table position | H |
| Table position | 0 mm |
| MSMA | S - C - T |
| Sagittal | R >> L |
| Coronal | A >> P |
| Transversal | F >> H |
| Coil Combine Mode | Sum of Squares |
| Matrix Optimization | Off |
| AutoAlign | Head > Brain |
| Coil Select Mode | Off - AutoCoilSelect |

### System - Adjustments

| | |
|---|---|
| B0 Shim mode | Advanced |
| B1 Shim mode | TrueForm |
| Confirm freq. adjustment | Off |
| Assume Dominant Fat | Off |
| Assume Silicone | Off |
| Adjustment Tolerance | Auto |

### System - Adjust Volume

| | |
|---|---|
| ! Position | L0.7 P13.2 H9.5 mm |
| ! Orientation | T > C-15.6 > S0.3 |
| ! Rotation | 0.00 deg |
| ! A >> P | 170 mm |
| ! R >> L | 130 mm |
| ! F >> H | 120 mm |
| Reset | Off |

### System - Tx/Rx

| | |
|---|---|
| Frequency 1H | 297.207726 MHz |
| Correction factor | 1 |
| Gain | High |
| Img. Scale Cor. | 1.000 |
| Reset | Off |
| ? Ref. amplitude 1H | 0.000 V |

### Physio - Signal1

| | |
|---|---|
| 1st Signal/Mode | None |
| TR | 1000 ms |
| Multi-band accel. factor | 5 |

### BOLD

| | |
|---|---|
| GLM Statistics | Off |
| Dynamic t-maps | Off |
| Ignore meas. at start | 0 |
| Ignore after transition | 0 |
| Model transition states | On |
| Temp. highpass filter | On |
| Threshold | 4.00 |
| Paradigm size | 20 |
| Meas[1] | Baseline |
| Meas[2] | Baseline |
| Meas[3] | Baseline |
| Meas[4] | Baseline |
| Meas[5] | Baseline |
| Meas[6] | Baseline |
| Meas[7] | Baseline |
| Meas[8] | Baseline |
| Meas[9] | Baseline |
| Meas[10] | Baseline |
| Meas[11] | Active |
| Meas[12] | Active |
| Meas[13] | Active |
| Meas[14] | Active |
| Meas[15] | Active |
| Meas[16] | Active |
| Meas[17] | Active |
| Meas[18] | Active |
| Meas[19] | Active |
| Meas[20] | Active |
| Motion correction | Off |
| Spatial filter | Off |
| Measurements | 297 |
| Delay in TR | 0 ms |
| Multiple series | Off |

### Sequence - Part 1

| | |
|---|---|
| Introduction | Off |
| Contrasts | 1 |
| Flow comp. | No |
| Multi-slice mode | Interleaved |
| Free echo spacing | Off |
| Echo spacing | 0.64 ms |
| Bandwidth | 1924 Hz/Px |

### Sequence - Part 2

| | |
|---|---|
| EPI factor | 130 |
| Gradient mode | Normal |
| RF spoiling | Off |

### Sequence - Special

| | |
|---|---|
| Excite pulse duration | 5760 us |
| Single-band images | On |
| MB LeakBlock kernel | Off |
| MB dual kernel | Off |
| MB RF phase scramble | On |
| SENSE1 coil combine | Off |
| Invert RO/PE polarity | Off |
| PF omits higher k-space | Off |
| Disable freq. update | Off |
| Force equal slice timing | Off |
| Online multi-band recon. | Online |
| FFT scale factor | 0.60 |
| Fat saturation FA | 110.0 deg |
| GRE iPAT ref. FA | 12.0 deg |
| Physio recording | Off |
| Triggering scheme | Standard |

| \\USER\PHCP\functional\final\FieldMap |
|---|
| TA: 2:14 PM: FIX Voxel size: 2.2×2.2×3.0 mmRel. SNR: 1.00 : fm_r |

### Properties

| | |
|---|---|
| Prio recon | Off |
| Load images to viewer | Off |
| Inline movie | Off |
| Auto store images | On |
| Load images to stamp segments | Off |
| Load images to graphic segments | Off |
| Auto open inline display | Off |
| Auto close inline display | Off |
| Start measurement without further preparation | Off |
| Wait for user to start | Off |
| Start measurements | Single measurement |

### Routine

| | |
|---|---|
| Slice group | 1 |
| Slices | 85 |
| Dist. factor | 0 % |
| Position | L0.0 P13.6 H7.5 mm |
| Orientation | T > C-20.0 |
| Phase enc. dir. | A >> P |
| AutoAlign | Head > Brain |
| Phase oversampling | 0 % |
| FoV read | 280 mm |
| FoV phase | 100.0 % |
| Slice thickness | 3.0 mm |
| TR | 668.0 ms |
| TE 1 | 4.08 ms |
| TE 2 | 5.1 ms |
| Averages | 1 |
| Concatenations | 1 |
| Filter | None |
| Coil elements | A32 |

### Contrast - Common

| | |
|---|---|
| TR | 668.0 ms |
| TE 1 | 4.08 ms |
| TE 2 | 5.1 ms |
| MTC | Off |
| Flip angle | 32 deg |
| Fat suppr. | None |

### Contrast - Dynamic

| | |
|---|---|
| Averages | 1 |
| Averaging mode | Short term |
| Reconstruction | Magn./Phase |
| Measurements | 1 |
| Multiple series | Off |

### Resolution - Common

| | |
|---|---|
| FoV read | 280 mm |
| FoV phase | 100.0 % |
| Slice thickness | 3.0 mm |
| Base resolution | 130 |
| Phase resolution | 100 % |
| Phase partial Fourier | 6/8 |
| Interpolation | Off |

### Resolution - Filter Image

| | |
|---|---|
| Image Filter | Off |
| Distortion Corr. | Off |

### Resolution - Filter Image

| | |
|---|---|
| Prescan Normalize | Off |
| Normalize | Off |
| B1 filter | Off |

### Resolution - Filter Rawdata

| | |
|---|---|
| Raw filter | Off |
| Elliptical filter | Off |

### Geometry - Common

| | |
|---|---|
| Slice group | 1 |
| Slices | 85 |
| Dist. factor | 0 % |
| Position | L0.0 P13.6 H7.5 mm |
| Orientation | T > C-20.0 |
| Phase enc. dir. | A >> P |
| FoV read | 280 mm |
| FoV phase | 100.0 % |
| Slice thickness | 3.0 mm |
| TR | 668.0 ms |
| Multi-slice mode | Interleaved |
| Series | Interleaved |
| Concatenations | 1 |

### Geometry - AutoAlign

| | |
|---|---|
| Slice group | 1 |
| Position | L0.0 P13.6 H7.5 mm |
| Orientation | T > C-20.0 |
| Phase enc. dir. | A >> P |
| AutoAlign | Head > Brain |
| Initial Position | L0.0 P13.6 H7.5 |
| L | 0.0 mm |
| P | 13.6 mm |
| H | 7.5 mm |
| Initial Rotation | 0.00 deg |
| Initial Orientation | T > C |
| T > C | -20.0 |
| > S | 0.0 |

### Geometry - Saturation

| | |
|---|---|
| Fat suppr. | None |
| Special sat. | None |

### Geometry - Tim Planning Suite

| | |
|---|---|
| Set-n-Go Protocol | Off |
| Table position | H |
| Table position | 0 mm |
| Inline Composing | Off |

### System - Miscellaneous

| | |
|---|---|
| Positioning mode | FIX |
| Table position | H |
| Table position | 0 mm |
| MSMA | S - C - T |
| Sagittal | R >> L |
| Coronal | A >> P |
| Transversal | F >> H |
| Coil Combine Mode | Adaptive Combine |
| Save uncombined | Off |
| Matrix Optimization | Off |
| AutoAlign | Head > Brain |

#### System - Miscellaneous

| | |
|---|---|
| Coil Select Mode | Off - AutoCoilSelect |

#### System - Adjustments

| | |
|---|---|
| B0 Shim mode | Advanced |
| B1 Shim mode | TrueForm |
| Confirm freq. adjustment | Off |
| Assume Dominant Fat | Off |
| Assume Silicone | Off |
| Adjustment Tolerance | Auto |

#### System - Adjust Volume

| | |
|---|---|
| ! Position | L0.7 P13.2 H9.5 mm |
| ! Orientation | T > C-15.6 > S0.3 |
| ! Rotation | 0.00 deg |
| ! A >> P | 170 mm |
| ! R >> L | 130 mm |
| ! F >> H | 120 mm |
| Reset | Off |

#### System - Tx/Rx

| | |
|---|---|
| Frequency 1H | 297.207726 MHz |
| Correction factor | 1 |
| Gain | High |
| Img. Scale Cor. | 1.000 |
| Reset | Off |
| ? Ref. amplitude 1H | 0.000 V |

#### Sequence - Part 1

| | |
|---|---|
| Introduction | On |
| Dimension | 2D |
| Asymmetric echo | Off |
| Contrasts | 2 |
| Flow comp. | Yes |
| Multi-slice mode | Interleaved |
| Bandwidth | 401 Hz/Px |

#### Sequence - Part 2

| | |
|---|---|
| RF pulse type | Normal |
| Gradient mode | Fast |
| RF spoiling | On |

#### Sequence - Assistant

| | |
|---|---|
| Mode | Off |

| \\USER\PHCP\functional\final\t1_mpr_tra_iso |
|---|
| TA: 4:17 PM: FIX Voxel size: 1.0×1.0×1.0 mmPAT: 3 Rel. SNR: 1.00 : tfl |

### Properties

| | |
|---|---|
| Prio recon | Off |
| Load images to viewer | On |
| Inline movie | Off |
| Auto store images | On |
| Load images to stamp segments | Off |
| Load images to graphic segments | Off |
| Auto open inline display | Off |
| Auto close inline display | Off |
| Start measurement without further preparation | Off |
| Wait for user to start | Off |
| Start measurements | Single measurement |

### Routine

| | |
|---|---|
| Slab group | 1 |
| Slabs | 1 |
| Dist. factor | 50 % |
| Position | Isocenter |
| Orientation | Sagittal |
| Phase enc. dir. | A >> P |
| AutoAlign | Head > Brain |
| Phase oversampling | 0 % |
| Slice oversampling | 0.0 % |
| Slices per slab | 176 |
| FoV read | 256 mm |
| FoV phase | 100.0 % |
| Slice thickness | 1.00 mm |
| TR | 3000.0 ms |
| TE | 3.27 ms |
| Averages | 1 |
| Concatenations | 1 |
| Filter | Distortion Corr.(3D) |
| Coil elements | A32 |

### Contrast - Common

| | |
|---|---|
| TR | 3000.0 ms |
| TE | 3.27 ms |
| Magn. preparation | Non-sel. IR |
| TI | 1500 ms |
| Flip angle | 5.0 deg |
| Fat suppr. | None |
| Water suppr. | None |

### Contrast - Dynamic

| | |
|---|---|
| Averages | 1 |
| Averaging mode | Long term |
| Reconstruction | Magnitude |
| Measurements | 1 |
| Multiple series | Each measurement |

### Resolution - Common

| | |
|---|---|
| FoV read | 256 mm |
| FoV phase | 100.0 % |
| Slice thickness | 1.00 mm |
| Base resolution | 256 |
| Phase resolution | 100 % |
| Slice resolution | 100 % |
| Phase partial Fourier | 6/8 |
| Slice partial Fourier | Off |
| Interpolation | Off |

### Resolution - iPAT

| | |
|---|---|
| PAT mode | GRAPPA |
| Accel. factor PE | 3 |
| Ref. lines PE | 32 |
| Accel. factor 3D | 1 |
| Reference scan mode | Integrated |

### Resolution - Filter Image

| | |
|---|---|
| Image Filter | Off |
| Distortion Corr. | On |
| Mode | 3D |
| Unfiltered images | On |
| Prescan Normalize | Off |
| Normalize | Off |
| B1 filter | Off |

### Resolution - Filter Rawdata

| | |
|---|---|
| Raw filter | Off |
| Elliptical filter | Off |

### Geometry - Common

| | |
|---|---|
| Slab group | 1 |
| Slabs | 1 |
| Dist. factor | 50 % |
| Position | Isocenter |
| Orientation | Sagittal |
| Phase enc. dir. | A >> P |
| Slice oversampling | 0.0 % |
| Slices per slab | 176 |
| FoV read | 256 mm |
| FoV phase | 100.0 % |
| Slice thickness | 1.00 mm |
| TR | 3000.0 ms |
| Multi-slice mode | Single shot |
| Series | Ascending |
| Concatenations | 1 |

### Geometry - AutoAlign

| | |
|---|---|
| Slab group | 1 |
| Position | Isocenter |
| Orientation | Sagittal |
| Phase enc. dir. | A >> P |
| AutoAlign | Head > Brain |
| Initial Position | Isocenter |
| L | 0.0 mm |
| P | 0.0 mm |
| H | 0.0 mm |
| Initial Rotation | 0.00 deg |
| Initial Orientation | Sagittal |

### Geometry - Navigator

### Geometry - Tim Planning Suite

| | |
|---|---|
| Set-n-Go Protocol | Off |
| Table position | H |
| Table position | 0 mm |
| Inline Composing | Off |

### System - Miscellaneous

| | |
|---|---|
| Positioning mode | FIX |
| Table position | H |

#### System - Miscellaneous

| | |
|---|---|
| Table position | 0 mm |
| MSMA | S - C - T |
| Sagittal | R >> L |
| Coronal | A >> P |
| Transversal | F >> H |
| Coil Combine Mode | Adaptive Combine |
| Save uncombined | Off |
| Matrix Optimization | Off |
| AutoAlign | Head > Brain |
| Coil Select Mode | Off - AutoCoilSelect |

#### System - Adjustments

| | |
|---|---|
| B0 Shim mode | Tune up |
| B1 Shim mode | TrueForm |
| Confirm freq. adjustment | Off |
| Assume Dominant Fat | Off |
| Assume Silicone | Off |
| Adjustment Tolerance | Auto |

#### System - Adjust Volume

| | |
|---|---|
| ! Position | L0.6 P15.0 H19.2 mm |
| ! Orientation | T > C-15.5 |
| ! Rotation | 0.00 deg |
| ! A >> P | 107 mm |
| ! R >> L | 104 mm |
| ! F >> H | 65 mm |
| Reset | Off |

#### System - Tx/Rx

| | |
|---|---|
| Frequency 1H | 297.207726 MHz |
| Correction factor | 1 |
| Gain | High |
| Img. Scale Cor. | 1.000 |
| Reset | Off |
| ? Ref. amplitude 1H | 0.000 V |

#### Physio - Signal1

| | |
|---|---|
| 1st Signal/Mode | None |
| TR | 3000.0 ms |
| Concatenations | 1 |

#### Physio - Cardiac

| | |
|---|---|
| Magn. preparation | Non-sel. IR |
| TI | 1500 ms |
| Fat suppr. | None |
| Dark blood | Off |
| FoV read | 256 mm |
| FoV phase | 100.0 % |
| Phase resolution | 100 % |

#### Physio - PACE

| | |
|---|---|
| Resp. control | Off |
| Concatenations | 1 |

#### Inline - Common

| | |
|---|---|
| Subtract | Off |
| Measurements | 1 |
| StdDev | Off |
| Save original images | On |

#### Inline - MIP

| | |
|---|---|
| MIP-Sag | Off |
| MIP-Cor | Off |

#### Inline - MIP

| | |
|---|---|
| MIP-Tra | Off |
| MIP-Time | Off |
| Save original images | On |

#### Inline - Composing

| | |
|---|---|
| Inline Composing | Off |
| Distortion Corr. | On |
| Mode | 3D |
| Unfiltered images | On |

#### Inline - MapIt

| | |
|---|---|
| Save original images | On |
| MapIt | None |
| Flip angle | 5.0 deg |
| Measurements | 1 |
| TR | 3000.0 ms |
| TE | 3.27 ms |

#### Sequence - Part 1

| | |
|---|---|
| Introduction | On |
| Dimension | 3D |
| Elliptical scanning | Off |
| Reordering | Linear |
| Asymmetric echo | Allowed |
| Flow comp. | No |
| Multi-slice mode | Single shot |
| Echo spacing | 8.1 ms |
| Bandwidth | 180 Hz/Px |

#### Sequence - Part 2

| | |
|---|---|
| RF pulse type | Normal |
| Gradient mode | Normal |
| Excitation | Slab-sel. |
| RF spoiling | On |
| Incr. Gradient spoiling | Off |
| Turbo factor | 176 |

#### Sequence - Nuclei

| | |
|---|---|
| TX/RX Nucleus | 1H |
| TX/RX delta frequency | 0 Hz |
| TX Nucleus | None |
| TX delta frequency | 0 Hz |
| Coil elements | A32 |

#### Sequence - Assistant

| | |
|---|---|
| Mode | Off |

| \\USER\PHCP\functional\final\BOLD_AP_SE |
|---|
| TA: 1:31 PM: FIX Voxel size: 1.6×1.6×1.6 mmPAT: 2 Rel. SNR: 1.00 : epse |

### Properties

| | |
|---|---|
| Prio recon | Off |
| Load images to viewer | On |
| Inline movie | Off |
| Auto store images | On |
| Load images to stamp segments | Off |
| Load images to graphic segments | Off |
| Auto open inline display | Off |
| Auto close inline display | Off |
| Start measurement without further preparation | Off |
| Wait for user to start | Off |
| Start measurements | Single measurement |

### Routine

| | |
|---|---|
| Slice group | 1 |
| Slices | 85 |
| Dist. factor | 0 % |
| Position | L0.0 P13.6 H7.5 mm |
| Orientation | T > C-20.0 |
| Phase enc. dir. | A >> P |
| AutoAlign | Head > Brain |
| Phase oversampling | 0 % |
| FoV read | 208 mm |
| FoV phase | 100.0 % |
| Slice thickness | 1.60 mm |
| TR | 3000 ms |
| TE | 60.00 ms |
| Multi-band accel. factor | 5 |
| Filter | None |
| Coil elements | A32 |

### Contrast - Common

| | |
|---|---|
| TR | 3000 ms |
| TE | 60.00 ms |
| MTC | Off |
| Magn. preparation | None |
| Flip angle | 90 deg |
| Refocus flip angle | 180 deg |
| Fat suppr. | None |
| Grad. rev. fat suppr. | Disabled |

### Contrast - Dynamic

| | |
|---|---|
| Averaging mode | Long term |
| Reconstruction | Magnitude |
| Measurements | 3 |
| Delay in TR | 0 ms |
| Multiple series | Off |

### Resolution - Common

| | |
|---|---|
| FoV read | 208 mm |
| FoV phase | 100.0 % |
| Slice thickness | 1.60 mm |
| Base resolution | 130 |
| Phase resolution | 100 % |
| Phase partial Fourier | 7/8 |
| Interpolation | Off |

### Resolution - iPAT

| | |
|---|---|
| PAT mode | GRAPPA |
| Accel. factor PE | 2 |

### Resolution - iPAT

| | |
|---|---|
| Ref. lines PE | 96 |
| Reference scan mode | GRE |

### Resolution - Filter Image

| | |
|---|---|
| Distortion Corr. | Off |
| Prescan Normalize | Off |

### Resolution - Filter Rawdata

| | |
|---|---|
| Raw filter | Off |
| Elliptical filter | Off |
| Hamming | Off |

### Geometry - Common

| | |
|---|---|
| Slice group | 1 |
| Slices | 85 |
| Dist. factor | 0 % |
| Position | L0.0 P13.6 H7.5 mm |
| Orientation | T > C-20.0 |
| Phase enc. dir. | A >> P |
| FoV read | 208 mm |
| FoV phase | 100.0 % |
| Slice thickness | 1.60 mm |
| TR | 3000 ms |
| Multi-slice mode | Interleaved |
| Series | Interleaved |
| Multi-band accel. factor | 5 |

### Geometry - AutoAlign

| | |
|---|---|
| Slice group | 1 |
| Position | L0.0 P13.6 H7.5 mm |
| Orientation | T > C-20.0 |
| Phase enc. dir. | A >> P |
| AutoAlign | Head > Brain |
| Initial Position | L0.0 P13.6 H7.5 |
| L | 0.0 mm |
| P | 13.6 mm |
| H | 7.5 mm |
| Initial Rotation | -0.51 deg |
| Initial Orientation | T > C |
| T > C | -20.0 |
| > S | 0.0 |

### Geometry - Saturation

| | |
|---|---|
| Fat suppr. | None |
| Grad. rev. fat suppr. | Disabled |
| Special sat. | None |

### Geometry - Tim Planning Suite

| | |
|---|---|
| Set-n-Go Protocol | Off |
| Table position | H |
| Table position | 0 mm |
| Inline Composing | Off |

### System - Miscellaneous

| | |
|---|---|
| Positioning mode | FIX |
| Table position | H |
| Table position | 0 mm |
| MSMA | S - C - T |
| Sagittal | R >> L |
| Coronal | A >> P |

### System - Miscellaneous

| | |
|---|---|
| Transversal | F >> H |
| Coil Combine Mode | Sum of Squares |
| Matrix Optimization | Off |
| AutoAlign | Head > Brain |
| Coil Select Mode | Off - AutoCoilSelect |

### System - Adjustments

| | |
|---|---|
| B0 Shim mode | Advanced |
| B1 Shim mode | TrueForm |
| Confirm freq. adjustment | Off |
| Assume Dominant Fat | Off |
| Assume Silicone | Off |
| Adjustment Tolerance | Auto |

### System - Adjust Volume

| | |
|---|---|
| ! Position | L0.7 P13.2 H9.5 mm |
| ! Orientation | T > C-15.6 > S0.3 |
| ! Rotation | 0.00 deg |
| ! A >> P | 170 mm |
| ! R >> L | 130 mm |
| ! F >> H | 120 mm |
| Reset | Off |

### System - Tx/Rx

| | |
|---|---|
| Frequency 1H | 297.207726 MHz |
| Correction factor | 1 |
| Gain | High |
| Img. Scale Cor. | 1.000 |
| Reset | Off |
| ? Ref. amplitude 1H | 0.000 V |

### Physio - Signal1

| | |
|---|---|
| 1st Signal/Mode | None |
| TR | 3000 ms |
| Multi-band accel. factor | 5 |

### BOLD

| | |
|---|---|
| GLM Statistics | Off |
| Dynamic t-maps | Off |
| Ignore meas. at start | 0 |
| Ignore after transition | 0 |
| Model transition states | On |
| Temp. highpass filter | On |
| Threshold | 4.00 |
| Paradigm size | 20 |
| Meas[1] | Baseline |
| Meas[2] | Baseline |
| Meas[3] | Baseline |
| Meas[4] | Baseline |
| Meas[5] | Baseline |
| Meas[6] | Baseline |
| Meas[7] | Baseline |
| Meas[8] | Baseline |
| Meas[9] | Baseline |
| Meas[10] | Baseline |
| Meas[11] | Active |
| Meas[12] | Active |
| Meas[13] | Active |
| Meas[14] | Active |
| Meas[15] | Active |
| Meas[16] | Active |
| Meas[17] | Active |
| Meas[18] | Active |
| Meas[19] | Active |

### BOLD

| | |
|---|---|
| Meas[20] | Active |
| Motion correction | Off |
| Spatial filter | Off |
| Measurements | 3 |
| Delay in TR | 0 ms |
| Multiple series | Off |

### Sequence - Part 1

| | |
|---|---|
| Introduction | Off |
| Contrasts | 1 |
| Multi-slice mode | Interleaved |
| Free echo spacing | Off |
| Echo spacing | 0.64 ms |
| Bandwidth | 1924 Hz/Px |

### Sequence - Part 2

| | |
|---|---|
| EPI factor | 130 |
| Gradient mode | Normal |

### Sequence - Special

| | |
|---|---|
| Excite pulse duration | 5120 us |
| Refocus pulse duration | 10240 us |
| Single-band images | On |
| MB LeakBlock kernel | Off |
| MB dual kernel | Off |
| MB RF phase scramble | Off |
| Time-shifted MB RF | Off |
| SENSE1 coil combine | Off |
| Invert RO/PE polarity | Off |
| PF omits higher k-space | Off |
| Disable freq. update | Off |
| Force equal slice timing | Off |
| Online multi-band recon. | Online |
| FFT scale factor | 1.00 |
| GRE iPAT ref. FA | 12.0 deg |
| Physio recording | Off |
| Triggering scheme | Standard |

**\\USER\PHCP\functional\final\BOLD_REV_AP_SE**

TA: 1:31 PM: FIX Voxel size: 1.6×1.6×1.6 mmPAT: 2 Rel. SNR: 1.00 : epse

### Properties

| | |
|---|---|
| Prio recon | Off |
| Load images to viewer | On |
| Inline movie | Off |
| Auto store images | On |
| Load images to stamp segments | Off |
| Load images to graphic segments | Off |
| Auto open inline display | Off |
| Auto close inline display | Off |
| Start measurement without further preparation | Off |
| Wait for user to start | Off |
| Start measurements | Single measurement |

### Routine

| | |
|---|---|
| Slice group | 1 |
| Slices | 85 |
| Dist. factor | 0 % |
| Position | L0.0 P13.6 H7.5 mm |
| Orientation | T > C-20.0 |
| Phase enc. dir. | A >> P |
| AutoAlign | Head > Brain |
| Phase oversampling | 0 % |
| FoV read | 208 mm |
| FoV phase | 100.0 % |
| Slice thickness | 1.60 mm |
| TR | 3000 ms |
| TE | 60.00 ms |
| Multi-band accel. factor | 5 |
| Filter | None |
| Coil elements | A32 |

### Contrast - Common

| | |
|---|---|
| TR | 3000 ms |
| TE | 60.00 ms |
| MTC | Off |
| Magn. preparation | None |
| Flip angle | 90 deg |
| Refocus flip angle | 180 deg |
| Fat suppr. | None |
| Grad. rev. fat suppr. | Disabled |

### Contrast - Dynamic

| | |
|---|---|
| Averaging mode | Long term |
| Reconstruction | Magnitude |
| Measurements | 3 |
| Delay in TR | 0 ms |
| Multiple series | Off |

### Resolution - Common

| | |
|---|---|
| FoV read | 208 mm |
| FoV phase | 100.0 % |
| Slice thickness | 1.60 mm |
| Base resolution | 130 |
| Phase resolution | 100 % |
| Phase partial Fourier | 7/8 |
| Interpolation | Off |

### Resolution - iPAT

| | |
|---|---|
| PAT mode | GRAPPA |
| Accel. factor PE | 2 |

### Resolution - iPAT

| | |
|---|---|
| Ref. lines PE | 96 |
| Reference scan mode | GRE |

### Resolution - Filter Image

| | |
|---|---|
| Distortion Corr. | Off |
| Prescan Normalize | Off |

### Resolution - Filter Rawdata

| | |
|---|---|
| Raw filter | Off |
| Elliptical filter | Off |
| Hamming | Off |

### Geometry - Common

| | |
|---|---|
| Slice group | 1 |
| Slices | 85 |
| Dist. factor | 0 % |
| Position | L0.0 P13.6 H7.5 mm |
| Orientation | T > C-20.0 |
| Phase enc. dir. | A >> P |
| FoV read | 208 mm |
| FoV phase | 100.0 % |
| Slice thickness | 1.60 mm |
| TR | 3000 ms |
| Multi-slice mode | Interleaved |
| Series | Interleaved |
| Multi-band accel. factor | 5 |

### Geometry - AutoAlign

| | |
|---|---|
| Slice group | 1 |
| Position | L0.0 P13.6 H7.5 mm |
| Orientation | T > C-20.0 |
| Phase enc. dir. | A >> P |
| AutoAlign | Head > Brain |
| Initial Position | L0.0 P13.6 H7.5 |
| L | 0.0 mm |
| P | 13.6 mm |
| H | 7.5 mm |
| Initial Rotation | -0.51 deg |
| Initial Orientation | T > C |
| T > C | -20.0 |
| > S | 0.0 |

### Geometry - Saturation

| | |
|---|---|
| Fat suppr. | None |
| Grad. rev. fat suppr. | Disabled |
| Special sat. | None |

### Geometry - Tim Planning Suite

| | |
|---|---|
| Set-n-Go Protocol | Off |
| Table position | H |
| Table position | 0 mm |
| Inline Composing | Off |

### System - Miscellaneous

| | |
|---|---|
| Positioning mode | FIX |
| Table position | H |
| Table position | 0 mm |
| MSMA | S - C - T |
| Sagittal | R >> L |
| Coronal | A >> P |

### System - Miscellaneous

| | |
|---|---|
| Transversal | F >> H |
| Coil Combine Mode | Sum of Squares |
| Matrix Optimization | Off |
| AutoAlign | Head > Brain |
| Coil Select Mode | Off - AutoCoilSelect |

### System - Adjustments

| | |
|---|---|
| B0 Shim mode | Advanced |
| B1 Shim mode | TrueForm |
| Confirm freq. adjustment | Off |
| Assume Dominant Fat | Off |
| Assume Silicone | Off |
| Adjustment Tolerance | Auto |

### System - Adjust Volume

| | |
|---|---|
| ! Position | L0.7 P13.2 H9.5 mm |
| ! Orientation | T > C-15.6 > S0.3 |
| ! Rotation | 0.00 deg |
| ! A >> P | 170 mm |
| ! R >> L | 130 mm |
| ! F >> H | 120 mm |
| Reset | Off |

### System - Tx/Rx

| | |
|---|---|
| Frequency 1H | 297.207726 MHz |
| Correction factor | 1 |
| Gain | High |
| Img. Scale Cor. | 1.000 |
| Reset | Off |
| ? Ref. amplitude 1H | 0.000 V |

### Physio - Signal1

| | |
|---|---|
| 1st Signal/Mode | None |
| TR | 3000 ms |
| Multi-band accel. factor | 5 |

### BOLD

| | |
|---|---|
| GLM Statistics | Off |
| Dynamic t-maps | Off |
| Ignore meas. at start | 0 |
| Ignore after transition | 0 |
| Model transition states | On |
| Temp. highpass filter | On |
| Threshold | 4.00 |
| Paradigm size | 20 |
| Meas[1] | Baseline |
| Meas[2] | Baseline |
| Meas[3] | Baseline |
| Meas[4] | Baseline |
| Meas[5] | Baseline |
| Meas[6] | Baseline |
| Meas[7] | Baseline |
| Meas[8] | Baseline |
| Meas[9] | Baseline |
| Meas[10] | Baseline |
| Meas[11] | Active |
| Meas[12] | Active |
| Meas[13] | Active |
| Meas[14] | Active |
| Meas[15] | Active |
| Meas[16] | Active |
| Meas[17] | Active |
| Meas[18] | Active |
| Meas[19] | Active |

### BOLD

| | |
|---|---|
| Meas[20] | Active |
| Motion correction | Off |
| Spatial filter | Off |
| Measurements | 3 |
| Delay in TR | 0 ms |
| Multiple series | Off |

### Sequence - Part 1

| | |
|---|---|
| Introduction | Off |
| Contrasts | 1 |
| Multi-slice mode | Interleaved |
| Free echo spacing | Off |
| Echo spacing | 0.64 ms |
| Bandwidth | 1924 Hz/Px |

### Sequence - Part 2

| | |
|---|---|
| EPI factor | 130 |
| Gradient mode | Normal |

### Sequence - Special

| | |
|---|---|
| Excite pulse duration | 5120 us |
| Refocus pulse duration | 10240 us |
| Single-band images | On |
| MB LeakBlock kernel | Off |
| MB dual kernel | Off |
| MB RF phase scramble | Off |
| Time-shifted MB RF | Off |
| SENSE1 coil combine | Off |
| Invert RO/PE polarity | On |
| PF omits higher k-space | Off |
| Disable freq. update | Off |
| Force equal slice timing | Off |
| Online multi-band recon. | Online |
| FFT scale factor | 1.00 |
| GRE iPAT ref. FA | 12.0 deg |
| Physio recording | Off |
| Triggering scheme | Standard |

| \\USER\PHCP\functional\final\BOLD_COP_task1_AP |
|---|
| TA: 9:07 PM: FIX Voxel size: 1.6×1.6×1.6 mmPAT: 2 Rel. SNR: 1.00 : epfid |

### Properties

| | |
|---|---|
| Prio recon | Off |
| Load images to viewer | Off |
| Inline movie | Off |
| Auto store images | On |
| Load images to stamp segments | Off |
| Load images to graphic segments | Off |
| Auto open inline display | Off |
| Auto close inline display | Off |
| Start measurement without further preparation | Off |
| Wait for user to start | On |
| Start measurements | Single measurement |

### Routine

| | |
|---|---|
| Slice group | 1 |
| Slices | 85 |
| Dist. factor | 0 % |
| Position | L0.0 P13.6 H7.5 mm |
| Orientation | T > C-20.0 |
| Phase enc. dir. | A >> P |
| AutoAlign | Head > Brain |
| Phase oversampling | 0 % |
| FoV read | 208 mm |
| FoV phase | 100.0 % |
| Slice thickness | 1.60 mm |
| TR | 1000 ms |
| TE | 22.20 ms |
| Multi-band accel. factor | 5 |
| Filter | None |
| Coil elements | A32 |

### Contrast - Common

| | |
|---|---|
| TR | 1000 ms |
| TE | 22.20 ms |
| MTC | Off |
| Magn. preparation | None |
| Flip angle | 45 deg |
| Fat suppr. | Fat sat. |

### Contrast - Dynamic

| | |
|---|---|
| Averaging mode | Long term |
| Reconstruction | Magnitude |
| Measurements | 468 |
| Delay in TR | 0 ms |
| Multiple series | Off |

### Resolution - Common

| | |
|---|---|
| FoV read | 208 mm |
| FoV phase | 100.0 % |
| Slice thickness | 1.60 mm |
| Base resolution | 130 |
| Phase resolution | 100 % |
| Phase partial Fourier | 7/8 |
| Interpolation | Off |

### Resolution - iPAT

| | |
|---|---|
| PAT mode | GRAPPA |
| Accel. factor PE | 2 |
| Ref. lines PE | 96 |
| Reference scan mode | GRE |

### Resolution - Filter Image

| | |
|---|---|
| Distortion Corr. | Off |
| Prescan Normalize | Off |

### Resolution - Filter Rawdata

| | |
|---|---|
| Raw filter | Off |
| Elliptical filter | Off |
| Hamming | Off |

### Geometry - Common

| | |
|---|---|
| Slice group | 1 |
| Slices | 85 |
| Dist. factor | 0 % |
| Position | L0.0 P13.6 H7.5 mm |
| Orientation | T > C-20.0 |
| Phase enc. dir. | A >> P |
| FoV read | 208 mm |
| FoV phase | 100.0 % |
| Slice thickness | 1.60 mm |
| TR | 1000 ms |
| Multi-slice mode | Interleaved |
| Series | Interleaved |
| Multi-band accel. factor | 5 |

### Geometry - AutoAlign

| | |
|---|---|
| Slice group | 1 |
| Position | L0.0 P13.6 H7.5 mm |
| Orientation | T > C-20.0 |
| Phase enc. dir. | A >> P |
| AutoAlign | Head > Brain |
| Initial Position | L0.0 P13.6 H7.5 |
| L | 0.0 mm |
| P | 13.6 mm |
| H | 7.5 mm |
| Initial Rotation | -0.52 deg |
| Initial Orientation | T > C |
| T > C | -20.0 |
| > S | 0.0 |

### Geometry - Saturation

| | |
|---|---|
| Fat suppr. | Fat sat. |
| Special sat. | None |

### Geometry - Tim Planning Suite

| | |
|---|---|
| Set-n-Go Protocol | Off |
| Table position | H |
| Table position | 0 mm |
| Inline Composing | Off |

### System - Miscellaneous

| | |
|---|---|
| Positioning mode | FIX |
| Table position | H |
| Table position | 0 mm |
| MSMA | S - C - T |
| Sagittal | R >> L |
| Coronal | A >> P |
| Transversal | F >> H |
| Coil Combine Mode | Sum of Squares |
| Matrix Optimization | Off |
| AutoAlign | Head > Brain |
| Coil Select Mode | Off - AutoCoilSelect |

### System - Adjustments

| | |
|---|---|
| B0 Shim mode | Advanced |
| B1 Shim mode | TrueForm |
| Confirm freq. adjustment | Off |
| Assume Dominant Fat | Off |
| Assume Silicone | Off |
| Adjustment Tolerance | Auto |

### System - Adjust Volume

| | |
|---|---|
| ! Position | L0.7 P13.2 H9.5 mm |
| ! Orientation | T > C-15.6 > S0.3 |
| ! Rotation | 0.00 deg |
| ! A >> P | 170 mm |
| ! R >> L | 130 mm |
| ! F >> H | 120 mm |
| Reset | Off |

### System - Tx/Rx

| | |
|---|---|
| Frequency 1H | 297.207726 MHz |
| Correction factor | 1 |
| Gain | High |
| Img. Scale Cor. | 1.000 |
| Reset | Off |
| ? Ref. amplitude 1H | 0.000 V |

### Physio - Signal1

| | |
|---|---|
| 1st Signal/Mode | None |
| TR | 1000 ms |
| Multi-band accel. factor | 5 |

### BOLD

| | |
|---|---|
| GLM Statistics | Off |
| Dynamic t-maps | Off |
| Ignore meas. at start | 0 |
| Ignore after transition | 0 |
| Model transition states | On |
| Temp. highpass filter | On |
| Threshold | 4.00 |
| Paradigm size | 20 |
| Meas[1] | Baseline |
| Meas[2] | Baseline |
| Meas[3] | Baseline |
| Meas[4] | Baseline |
| Meas[5] | Baseline |
| Meas[6] | Baseline |
| Meas[7] | Baseline |
| Meas[8] | Baseline |
| Meas[9] | Baseline |
| Meas[10] | Baseline |
| Meas[11] | Active |
| Meas[12] | Active |
| Meas[13] | Active |
| Meas[14] | Active |
| Meas[15] | Active |
| Meas[16] | Active |
| Meas[17] | Active |
| Meas[18] | Active |
| Meas[19] | Active |
| Meas[20] | Active |
| Motion correction | Off |
| Spatial filter | Off |
| Measurements | 468 |
| Delay in TR | 0 ms |
| Multiple series | Off |

### Sequence - Part 1

| | |
|---|---|
| Introduction | Off |
| Contrasts | 1 |
| Flow comp. | No |
| Multi-slice mode | Interleaved |
| Free echo spacing | Off |
| Echo spacing | 0.64 ms |
| Bandwidth | 1924 Hz/Px |

### Sequence - Part 2

| | |
|---|---|
| EPI factor | 130 |
| Gradient mode | Normal |
| RF spoiling | Off |

### Sequence - Special

| | |
|---|---|
| Excite pulse duration | 5760 us |
| Single-band images | On |
| MB LeakBlock kernel | Off |
| MB dual kernel | Off |
| MB RF phase scramble | On |
| SENSE1 coil combine | Off |
| Invert RO/PE polarity | Off |
| PF omits higher k-space | Off |
| Disable freq. update | Off |
| Force equal slice timing | Off |
| Online multi-band recon. | Online |
| FFT scale factor | 0.60 |
| Fat saturation FA | 110.0 deg |
| GRE iPAT ref. FA | 12.0 deg |
| Physio recording | Off |
| Triggering scheme | Standard |

**\\USER\PHCP\functional\final\BOLD_COP_task2_AP**

TA: 6:31 PM: FIX Voxel size: 1.6×1.6×1.6 mmPAT: 2 Rel. SNR: 1.00 : epfid

### Properties

| | |
|---|---|
| Prio recon | Off |
| Load images to viewer | Off |
| Inline movie | Off |
| Auto store images | On |
| Load images to stamp segments | Off |
| Load images to graphic segments | Off |
| Auto open inline display | Off |
| Auto close inline display | Off |
| Start measurement without further preparation | Off |
| Wait for user to start | On |
| Start measurements | Single measurement |

### Routine

| | |
|---|---|
| Slice group | 1 |
| Slices | 85 |
| Dist. factor | 0 % |
| Position | L0.0 P13.6 H7.5 mm |
| Orientation | T > C-20.0 |
| Phase enc. dir. | A >> P |
| AutoAlign | Head > Brain |
| Phase oversampling | 0 % |
| FoV read | 208 mm |
| FoV phase | 100.0 % |
| Slice thickness | 1.60 mm |
| TR | 1000 ms |
| TE | 22.20 ms |
| Multi-band accel. factor | 5 |
| Filter | None |
| Coil elements | A32 |

### Contrast - Common

| | |
|---|---|
| TR | 1000 ms |
| TE | 22.20 ms |
| MTC | Off |
| Magn. preparation | None |
| Flip angle | 45 deg |
| Fat suppr. | Fat sat. |

### Contrast - Dynamic

| | |
|---|---|
| Averaging mode | Long term |
| Reconstruction | Magnitude |
| Measurements | 312 |
| Delay in TR | 0 ms |
| Multiple series | Off |

### Resolution - Common

| | |
|---|---|
| FoV read | 208 mm |
| FoV phase | 100.0 % |
| Slice thickness | 1.60 mm |
| Base resolution | 130 |
| Phase resolution | 100 % |
| Phase partial Fourier | 7/8 |
| Interpolation | Off |

### Resolution - iPAT

| | |
|---|---|
| PAT mode | GRAPPA |
| Accel. factor PE | 2 |
| Ref. lines PE | 96 |
| Reference scan mode | GRE |

### Resolution - Filter Image

| | |
|---|---|
| Distortion Corr. | Off |
| Prescan Normalize | Off |

### Resolution - Filter Rawdata

| | |
|---|---|
| Raw filter | Off |
| Elliptical filter | Off |
| Hamming | Off |

### Geometry - Common

| | |
|---|---|
| Slice group | 1 |
| Slices | 85 |
| Dist. factor | 0 % |
| Position | L0.0 P13.6 H7.5 mm |
| Orientation | T > C-20.0 |
| Phase enc. dir. | A >> P |
| FoV read | 208 mm |
| FoV phase | 100.0 % |
| Slice thickness | 1.60 mm |
| TR | 1000 ms |
| Multi-slice mode | Interleaved |
| Series | Interleaved |
| Multi-band accel. factor | 5 |

### Geometry - AutoAlign

| | |
|---|---|
| Slice group | 1 |
| Position | L0.0 P13.6 H7.5 mm |
| Orientation | T > C-20.0 |
| Phase enc. dir. | A >> P |
| AutoAlign | Head > Brain |
| Initial Position | L0.0 P13.6 H7.5 |
| L | 0.0 mm |
| P | 13.6 mm |
| H | 7.5 mm |
| Initial Rotation | -0.52 deg |
| Initial Orientation | T > C |
| T > C | -20.0 |
| > S | 0.0 |

### Geometry - Saturation

| | |
|---|---|
| Fat suppr. | Fat sat. |
| Special sat. | None |

### Geometry - Tim Planning Suite

| | |
|---|---|
| Set-n-Go Protocol | Off |
| Table position | H |
| Table position | 0 mm |
| Inline Composing | Off |

### System - Miscellaneous

| | |
|---|---|
| Positioning mode | FIX |
| Table position | H |
| Table position | 0 mm |
| MSMA | S - C - T |
| Sagittal | R >> L |
| Coronal | A >> P |
| Transversal | F >> H |
| Coil Combine Mode | Sum of Squares |
| Matrix Optimization | Off |
| AutoAlign | Head > Brain |
| Coil Select Mode | Off - AutoCoilSelect |

### System - Adjustments

| | |
|---|---|
| B0 Shim mode | Advanced |
| B1 Shim mode | TrueForm |
| Confirm freq. adjustment | Off |
| Assume Dominant Fat | Off |
| Assume Silicone | Off |
| Adjustment Tolerance | Auto |

### System - Adjust Volume

| | |
|---|---|
| ! Position | L0.7 P13.2 H9.5 mm |
| ! Orientation | T > C-15.6 > S0.3 |
| ! Rotation | 0.00 deg |
| ! A >> P | 170 mm |
| ! R >> L | 130 mm |
| ! F >> H | 120 mm |
| Reset | Off |

### System - Tx/Rx

| | |
|---|---|
| Frequency 1H | 297.207726 MHz |
| Correction factor | 1 |
| Gain | High |
| Img. Scale Cor. | 1.000 |
| Reset | Off |
| ? Ref. amplitude 1H | 0.000 V |

### Physio - Signal1

| | |
|---|---|
| 1st Signal/Mode | None |
| TR | 1000 ms |
| Multi-band accel. factor | 5 |

### BOLD

| | |
|---|---|
| GLM Statistics | Off |
| Dynamic t-maps | Off |
| Ignore meas. at start | 0 |
| Ignore after transition | 0 |
| Model transition states | On |
| Temp. highpass filter | On |
| Threshold | 4.00 |
| Paradigm size | 20 |
| Meas[1] | Baseline |
| Meas[2] | Baseline |
| Meas[3] | Baseline |
| Meas[4] | Baseline |
| Meas[5] | Baseline |
| Meas[6] | Baseline |
| Meas[7] | Baseline |
| Meas[8] | Baseline |
| Meas[9] | Baseline |
| Meas[10] | Baseline |
| Meas[11] | Active |
| Meas[12] | Active |
| Meas[13] | Active |
| Meas[14] | Active |
| Meas[15] | Active |
| Meas[16] | Active |
| Meas[17] | Active |
| Meas[18] | Active |
| Meas[19] | Active |
| Meas[20] | Active |
| Motion correction | Off |
| Spatial filter | Off |
| Measurements | 312 |
| Delay in TR | 0 ms |
| Multiple series | Off |

### Sequence - Part 1

| | |
|---|---|
| Introduction | Off |
| Contrasts | 1 |
| Flow comp. | No |
| Multi-slice mode | Interleaved |
| Free echo spacing | Off |
| Echo spacing | 0.64 ms |
| Bandwidth | 1924 Hz/Px |

### Sequence - Part 2

| | |
|---|---|
| EPI factor | 130 |
| Gradient mode | Normal |
| RF spoiling | Off |

### Sequence - Special

| | |
|---|---|
| Excite pulse duration | 5760 us |
| Single-band images | On |
| MB LeakBlock kernel | Off |
| MB dual kernel | Off |
| MB RF phase scramble | On |
| SENSE1 coil combine | Off |
| Invert RO/PE polarity | Off |
| PF omits higher k-space | Off |
| Disable freq. update | Off |
| Force equal slice timing | Off |
| Online multi-band recon. | Online |
| FFT scale factor | 0.60 |
| Fat saturation FA | 110.0 deg |
| GRE iPAT ref. FA | 12.0 deg |
| Physio recording | Off |
| Triggering scheme | Standard |

| \\USER\PHCP\functional\final\BOLD_PRF2_AP |
|---|
| TA: 6:43 PM: FIX Voxel size: 1.6×1.6×1.6 mmPAT: 2 Rel. SNR: 1.00 : epfid |

### Properties

| | |
|---|---|
| Prio recon | Off |
| Load images to viewer | Off |
| Inline movie | Off |
| Auto store images | On |
| Load images to stamp segments | Off |
| Load images to graphic segments | Off |
| Auto open inline display | Off |
| Auto close inline display | Off |
| Start measurement without further preparation | Off |
| Wait for user to start | On |
| Start measurements | Single measurement |

### Routine

| | |
|---|---|
| Slice group | 1 |
| Slices | 85 |
| Dist. factor | 0 % |
| Position | L0.0 P13.6 H7.5 mm |
| Orientation | T > C-20.0 |
| Phase enc. dir. | A >> P |
| AutoAlign | Head > Brain |
| Phase oversampling | 0 % |
| FoV read | 208 mm |
| FoV phase | 100.0 % |
| Slice thickness | 1.60 mm |
| TR | 1000 ms |
| TE | 22.20 ms |
| Multi-band accel. factor | 5 |
| Filter | None |
| Coil elements | A32 |

### Contrast - Common

| | |
|---|---|
| TR | 1000 ms |
| TE | 22.20 ms |
| MTC | Off |
| Magn. preparation | None |
| Flip angle | 45 deg |
| Fat suppr. | Fat sat. |

### Contrast - Dynamic

| | |
|---|---|
| Averaging mode | Long term |
| Reconstruction | Magnitude |
| Measurements | 324 |
| Delay in TR | 0 ms |
| Multiple series | Off |

### Resolution - Common

| | |
|---|---|
| FoV read | 208 mm |
| FoV phase | 100.0 % |
| Slice thickness | 1.60 mm |
| Base resolution | 130 |
| Phase resolution | 100 % |
| Phase partial Fourier | 7/8 |
| Interpolation | Off |

### Resolution - iPAT

| | |
|---|---|
| PAT mode | GRAPPA |
| Accel. factor PE | 2 |
| Ref. lines PE | 96 |
| Reference scan mode | GRE |

### Resolution - Filter Image

| | |
|---|---|
| Distortion Corr. | Off |
| Prescan Normalize | Off |

### Resolution - Filter Rawdata

| | |
|---|---|
| Raw filter | Off |
| Elliptical filter | Off |
| Hamming | Off |

### Geometry - Common

| | |
|---|---|
| Slice group | 1 |
| Slices | 85 |
| Dist. factor | 0 % |
| Position | L0.0 P13.6 H7.5 mm |
| Orientation | T > C-20.0 |
| Phase enc. dir. | A >> P |
| FoV read | 208 mm |
| FoV phase | 100.0 % |
| Slice thickness | 1.60 mm |
| TR | 1000 ms |
| Multi-slice mode | Interleaved |
| Series | Interleaved |
| Multi-band accel. factor | 5 |

### Geometry - AutoAlign

| | |
|---|---|
| Slice group | 1 |
| Position | L0.0 P13.6 H7.5 mm |
| Orientation | T > C-20.0 |
| Phase enc. dir. | A >> P |
| AutoAlign | Head > Brain |
| Initial Position | L0.0 P13.6 H7.5 |
| L | 0.0 mm |
| P | 13.6 mm |
| H | 7.5 mm |
| Initial Rotation | -0.52 deg |
| Initial Orientation | T > C |
| T > C | -20.0 |
| > S | 0.0 |

### Geometry - Saturation

| | |
|---|---|
| Fat suppr. | Fat sat. |
| Special sat. | None |

### Geometry - Tim Planning Suite

| | |
|---|---|
| Set-n-Go Protocol | Off |
| Table position | H |
| Table position | 0 mm |
| Inline Composing | Off |

### System - Miscellaneous

| | |
|---|---|
| Positioning mode | FIX |
| Table position | H |
| Table position | 0 mm |
| MSMA | S - C - T |
| Sagittal | R >> L |
| Coronal | A >> P |
| Transversal | F >> H |
| Coil Combine Mode | Sum of Squares |
| Matrix Optimization | Off |
| AutoAlign | Head > Brain |
| Coil Select Mode | Off - AutoCoilSelect |

### System - Adjustments

| | |
|---|---|
| B0 Shim mode | Advanced |
| B1 Shim mode | TrueForm |
| Confirm freq. adjustment | Off |
| Assume Dominant Fat | Off |
| Assume Silicone | Off |
| Adjustment Tolerance | Auto |

### System - Adjust Volume

| | |
|---|---|
| ! Position | L0.7 P13.2 H9.5 mm |
| ! Orientation | T > C-15.6 > S0.3 |
| ! Rotation | 0.00 deg |
| ! A >> P | 170 mm |
| ! R >> L | 130 mm |
| ! F >> H | 120 mm |
| Reset | Off |

### System - Tx/Rx

| | |
|---|---|
| Frequency 1H | 297.207726 MHz |
| Correction factor | 1 |
| Gain | High |
| Img. Scale Cor. | 1.000 |
| Reset | Off |
| ? Ref. amplitude 1H | 0.000 V |

### Physio - Signal1

| | |
|---|---|
| 1st Signal/Mode | None |
| TR | 1000 ms |
| Multi-band accel. factor | 5 |

### BOLD

| | |
|---|---|
| GLM Statistics | Off |
| Dynamic t-maps | Off |
| Ignore meas. at start | 0 |
| Ignore after transition | 0 |
| Model transition states | On |
| Temp. highpass filter | On |
| Threshold | 4.00 |
| Paradigm size | 20 |
| Meas[1] | Baseline |
| Meas[2] | Baseline |
| Meas[3] | Baseline |
| Meas[4] | Baseline |
| Meas[5] | Baseline |
| Meas[6] | Baseline |
| Meas[7] | Baseline |
| Meas[8] | Baseline |
| Meas[9] | Baseline |
| Meas[10] | Baseline |
| Meas[11] | Active |
| Meas[12] | Active |
| Meas[13] | Active |
| Meas[14] | Active |
| Meas[15] | Active |
| Meas[16] | Active |
| Meas[17] | Active |
| Meas[18] | Active |
| Meas[19] | Active |
| Meas[20] | Active |
| Motion correction | Off |
| Spatial filter | Off |
| Measurements | 324 |
| Delay in TR | 0 ms |
| Multiple series | Off |

### Sequence - Part 1

| | |
|---|---|
| Introduction | Off |
| Contrasts | 1 |
| Flow comp. | No |
| Multi-slice mode | Interleaved |
| Free echo spacing | Off |
| Echo spacing | 0.64 ms |
| Bandwidth | 1924 Hz/Px |

### Sequence - Part 2

| | |
|---|---|
| EPI factor | 130 |
| Gradient mode | Normal |
| RF spoiling | Off |

### Sequence - Special

| | |
|---|---|
| Excite pulse duration | 5760 us |
| Single-band images | On |
| MB LeakBlock kernel | Off |
| MB dual kernel | Off |
| MB RF phase scramble | On |
| SENSE1 coil combine | Off |
| Invert RO/PE polarity | Off |
| PF omits higher k-space | Off |
| Disable freq. update | Off |
| Force equal slice timing | Off |
| Online multi-band recon. | Online |
| FFT scale factor | 0.60 |
| Fat saturation FA | 110.0 deg |
| GRE iPAT ref. FA | 12.0 deg |
| Physio recording | Off |
| Triggering scheme | Standard |
