## Supplementary material for "The Psychosis Human Connectome Project: Design and rationale for studies of visual neurophysiology": MRS Protocol

**Table of contents**

\\USER
PHCP
MRS
final
localizer
AAScout_32ch
localizer aligned
t1inplane 64sl occ 1p25iso 2p5sTR 160FOV
OCC fastestmap lin
OCC fastestmap all
OCC fastestmap all
OCC fastestmap all
OCC fastestmap lin6
OCC linewidth check
OCC steam eja cal fa 50 10 7
OCC steam eja cal fa 80 3 10
OCC steam eja ws check
OCC steam eja metab
OCC steam eja w1
OCC steam eja w4
OCC steam eja w1 noOVS
OCC steam eja ws FA optimization
OCC steam eja ws delay 7
PFC localizer
PFC AAScout_32ch
localizer aligned
PFC t1inplane 64sl occ 1p25iso 2p5sTR 160FOV
PFC fastestmap lin
PFC fastestmap all
PFC fastestmap all
PFC fastestmap all
PFC fastestmap lin6
PFC linewidth check
PFC steam eja cal fa 50 10 7
PFC steam eja cal fa 80 3 10
PFC steam eja ws check
PFC steam eja metab
PFC steam eja w1
PFC steam eja w4
PFC steam eja w1 noOVS
PFC steam eja ws FA optimization
PFC steam eja ws delay 7

| \\USER\PHCP\MRS\final\localizer |
|---|
| TA: 0:13 PM: REF Voxel size: 0.5×0.5×7.0 mmPAT: Off Rel. SNR: 1.00 : qfl |

### Properties

| | |
|---|---|
| Prio recon | Off |
| Load images to viewer | On |
| Inline movie | Off |
| Auto store images | On |
| Load images to stamp segments | On |
| Load images to graphic segments | On |
| Auto open inline display | On |
| Auto close inline display | Off |
| Start measurement without further preparation | Off |
| Wait for user to start | Off |
| Start measurements | Single measurement |

### Routine

| | |
|---|---|
| Slice group | 1 |
| Slices | 1 |
| Dist. factor | 20 % |
| Position | Isocenter |
| Orientation | Sagittal |
| Phase enc. dir. | A >> P |
| Slice group | 2 |
| Slices | 1 |
| Dist. factor | 20 % |
| Position | Isocenter |
| Orientation | Transversal |
| Phase enc. dir. | A >> P |
| Slice group | 3 |
| Slices | 1 |
| Dist. factor | 20 % |
| Position | Isocenter |
| Orientation | Coronal |
| Phase enc. dir. | R >> L |
| AutoAlign | --- |
| Phase oversampling | 0 % |
| FoV read | 250 mm |
| FoV phase | 100.0 % |
| Slice thickness | 7.0 mm |
| TR | 8.6 ms |
| TE | 4.00 ms |
| Averages | 2 |
| Concatenations | 3 |
| Filter | Elliptical filter |
| Coil elements | OCC;PFC |

### Contrast - Common

| | |
|---|---|
| TR | 8.6 ms |
| TE | 4.00 ms |
| TD | 0 ms |
| MTC | Off |
| Magn. preparation | None |
| Flip angle | 20 deg |
| Fat suppr. | None |
| Water suppr. | None |
| SWI | Off |

### Contrast - Dynamic

| | |
|---|---|
| Averages | 2 |
| Averaging mode | Short term |
| Reconstruction | Magnitude |
| Measurements | 1 |

| | |
|---|---|
| Tim CT mode | Off |
| Slices | 1 |
| Slice thickness | 7.0 mm |
| Dist. factor | 20 % |
| FoV read | 250 mm |
| FoV phase | 100.0 % |
| Segments | 1 |

#### System - Miscellaneous

| | |
|---|---|
| Positioning mode | REF |
| Table position | F |
| Table position | 0 mm |
| MSMA | S - C - T |
| Sagittal | R >> L |
| Coronal | A >> P |
| Transversal | F >> H |
| Coil Combine Mode | Sum of Squares |
| Save uncombined | Off |
| Matrix Optimization | Off |
| AutoAlign | --- |
| Coil Select Mode | Off - AutoCoilSelect |

#### System - Adjustments

| | |
|---|---|
| B0 Shim mode | Tune up |
| B1 Shim mode | TrueForm |
| Confirm freq. adjustment | Off |
| Assume Dominant Fat | Off |
| Assume Silicone | Off |
| Adjustment Tolerance | Auto |

#### System - Adjust Volume

| | |
|---|---|
| Position | Isocenter |
| Orientation | Transversal |
| Rotation | 0.00 deg |
| A >> P | 263 mm |
| R >> L | 350 mm |
| F >> H | 350 mm |
| Reset | Off |

#### System - Tx/Rx

| | |
|---|---|
| Frequency 1H | 297.207726 MHz |
| Correction factor | 1 |
| Gain | High |
| Img. Scale Cor. | 1.000 |
| Reset | Off |
| ? Ref. amplitude 1H | 0.000 V |

#### Physio - Signal1

| | |
|---|---|
| 1st Signal/Mode | None |
| TR | 8.6 ms |
| Concatenations | 3 |
| Segments | 1 |

#### Physio - Cardiac

| | |
|---|---|
| Tagging | None |
| Magn. preparation | None |
| Fat suppr. | None |
| Dark blood | Off |
| FoV read | 250 mm |
| FoV phase | 100.0 % |
| Phase resolution | 90 % |

#### Physio - PACE

| | |
|---|---|
| Resp. control | Off |
| Concatenations | 3 |

#### Inline - Common

| | |
|---|---|
| Subtract | Off |
| Measurements | 1 |
| StdDev | Off |
| Liver registration | Off |
| Save original images | On |

#### Inline - MIP

| | |
|---|---|
| MIP-Sag | Off |
| MIP-Cor | Off |
| MIP-Tra | Off |
| MIP-Time | Off |
| Save original images | On |

#### Inline - Soft Tissue

| | |
|---|---|
| Wash - In | Off |
| Wash - Out | Off |
| TTP | Off |
| PEI | Off |
| MIP - time | Off |
| Measurements | 1 |

#### Inline - Composing

| | |
|---|---|
| Inline Composing | Off |
| Distortion Corr. | Off |

#### Inline - MapIt

| | |
|---|---|
| Save original images | On |
| MapIt | None |
| Flip angle | 20 deg |
| Measurements | 1 |
| Contrasts | 1 |
| TR | 8.6 ms |
| TE | 4.00 ms |

#### Sequence - Part 1

| | |
|---|---|
| Introduction | On |

### Sequence - Part 1

| | |
|---|---|
| Dimension | 2D |
| Phase stabilisation | Off |
| Asymmetric echo | Allowed |
| Contrasts | 1 |
| Flow comp. | No |
| Multi-slice mode | Sequential |
| Bandwidth | 320 Hz/Px |

### Geometry - AutoAlign

| | |
|---|---|
| Slab group | 1 |
| Position | L0.0 A22.2 F11.5 mm |
| Orientation | Sagittal |
| Phase enc. dir. | A >> P |
| Initial Position | Isocenter |
| L | 0.0 mm |
| P | 0.0 mm |
| H | 0.0 mm |
| Initial Rotation | 0.00 deg |
| Initial Orientation | Transversal |

### Sequence - Assistant

| | |
|---|---|
| Mode | Off |

| \\USER\PHCP\MRS\final\localizer aligned |
|---|
| TA: 0:28 PM: FIX Voxel size: 0.5×0.5×5.0 mmPAT: Off Rel. SNR: 1.00 : qfl |

### Properties

| | |
|---|---|
| Prio recon | Off |
| Load images to viewer | On |
| Inline movie | Off |
| Auto store images | On |
| Load images to stamp segments | On |
| Load images to graphic segments | On |
| Auto open inline display | Off |
| Auto close inline display | Off |
| Start measurement without further preparation | On |
| Wait for user to start | Off |
| Start measurements | Single measurement |

#### Sequence - Assistant

| | |
|---|---|
| Mode | Off |

| \\USER\PHCP\MRS\final\t1inplane 64sl occc 1p25iso 2p5sTR 160FOV |
|---|
| TA: 4:02 PM: FIX Voxel size: 1.3×1.3×1.3 mmPAT: Off Rel. SNR: 1.00 : tfl |

### Properties

| | |
|---|---|
| Prio recon | Off |
| Load images to viewer | On |
| Inline movie | Off |
| Auto store images | On |
| Load images to stamp segments | On |
| Load images to graphic segments | On |
| Auto open inline display | Off |
| Auto close inline display | Off |
| Start measurement without further preparation | Off |
| Wait for user to start | Off |
| Start measurements | Single measurement |

### Routine

| | |
|---|---|
| Slab group | 1 |
| Slabs | 1 |
| Dist. factor | 50 % |
| Position | L1.8 A1.2 H7.3 mm |
| Orientation | Sagittal |
| Phase enc. dir. | A >> P |
| AutoAlign | Head > Brain |
| Phase oversampling | 0 % |
| Slice oversampling | 0.0 % |
| Slices per slab | 64 |
| FoV read | 160 mm |
| FoV phase | 100.0 % |
| Slice thickness | 1.25 mm |
| TR | 2500.0 ms |
| TE | 3.2 ms |
| Averages | 1 |
| Concatenations | 1 |
| Filter | None |
| Coil elements | OCC |

### Contrast - Common

| | |
|---|---|
| TR | 2500.0 ms |
| TE | 3.2 ms |
| Magn. preparation | Non-sel. IR |
| TI | 1500 ms |
| Flip angle | 5.0 deg |
| Fat suppr. | None |
| Water suppr. | None |

### Contrast - Dynamic

| | |
|---|---|
| Averages | 1 |
| Averaging mode | Long term |
| Reconstruction | Magnitude |
| Measurements | 1 |
| Multiple series | Each measurement |

### Resolution - Common

| | |
|---|---|
| FoV read | 160 mm |
| FoV phase | 100.0 % |
| Slice thickness | 1.25 mm |
| Base resolution | 128 |
| Phase resolution | 100 % |
| Slice resolution | 100 % |
| Phase partial Fourier | 6/8 |
| Slice partial Fourier | Off |
| Interpolation | Off |

### Resolution - iPAT

| | |
|---|---|
| PAT mode | None |

### Resolution - Filter Image

| | |
|---|---|
| Image Filter | Off |
| Distortion Corr. | Off |
| Prescan Normalize | Off |
| Normalize | Off |
| B1 filter | Off |

### Resolution - Filter Rawdata

| | |
|---|---|
| Raw filter | Off |
| Elliptical filter | Off |

### Geometry - Common

| | |
|---|---|
| Slab group | 1 |
| Slabs | 1 |
| Dist. factor | 50 % |
| Position | L1.8 A1.2 H7.3 mm |
| Orientation | Sagittal |
| Phase enc. dir. | A >> P |
| Slice oversampling | 0.0 % |
| Slices per slab | 64 |
| FoV read | 160 mm |
| FoV phase | 100.0 % |
| Slice thickness | 1.25 mm |
| TR | 2500.0 ms |
| Multi-slice mode | Single shot |
| Series | Ascending |
| Concatenations | 1 |

### Geometry - AutoAlign

| | |
|---|---|
| Slab group | 1 |
| Position | L1.8 A1.2 H7.3 mm |
| Orientation | Sagittal |
| Phase enc. dir. | A >> P |
| AutoAlign | Head > Brain |
| Initial Position | L1.8 A1.2 H7.3 |
| L | 1.8 mm |
| A | 1.2 mm |
| H | 7.3 mm |
| Initial Rotation | 0.00 deg |
| Initial Orientation | Sagittal |

### Geometry - Navigator

### Geometry - Tim Planning Suite

| | |
|---|---|
| Set-n-Go Protocol | Off |
| Table position | H |
| Table position | 0 mm |
| Inline Composing | Off |

### System - Miscellaneous

| | |
|---|---|
| Positioning mode | FIX |
| Table position | H |
| Table position | 0 mm |
| MSMA | S - C - T |
| Sagittal | R >> L |
| Coronal | A >> P |
| Transversal | F >> H |
| Coil Combine Mode | Adaptive Combine |

### System - Miscellaneous

| | |
|---|---|
| Save uncombined | Off |
| Matrix Optimization | Off |
| AutoAlign | Head > Brain |
| Coil Select Mode | Off - All |

### System - Adjustments

| | |
|---|---|
| B0 Shim mode | Tune up |
| B1 Shim mode | TrueForm |
| Confirm freq. adjustment | Off |
| Assume Dominant Fat | Off |
| Assume Silicone | Off |
| Adjustment Tolerance | Auto |

### System - Adjust Volume

| | |
|---|---|
| ! Position | L1.8 P18.8 H15.1 mm |
| ! Orientation | T > C-18.8 |
| ! Rotation | 0.00 deg |
| ! A >> P | 64 mm |
| ! R >> L | 148 mm |
| ! F >> H | 82 mm |
| Reset | Off |

### System - Tx/Rx

| | |
|---|---|
| Frequency 1H | 297.207726 MHz |
| Correction factor | 1 |
| Gain | High |
| Img. Scale Cor. | 1.000 |
| Reset | Off |
| ? Ref. amplitude 1H | 0.000 V |

### Physio - Signal1

| | |
|---|---|
| 1st Signal/Mode | None |
| TR | 2500.0 ms |
| Concatenations | 1 |

### Physio - Cardiac

| | |
|---|---|
| Magn. preparation | Non-sel. IR |
| TI | 1500 ms |
| Fat suppr. | None |
| Dark blood | Off |
| FoV read | 160 mm |
| FoV phase | 100.0 % |
| Phase resolution | 100 % |

### Physio - PACE

| | |
|---|---|
| Resp. control | Off |
| Concatenations | 1 |

### Inline - Common

| | |
|---|---|
| Subtract | Off |
| Measurements | 1 |
| StdDev | Off |
| Save original images | On |

### Inline - MIP

| | |
|---|---|
| MIP-Sag | Off |
| MIP-Cor | Off |
| MIP-Tra | Off |
| MIP-Time | Off |
| Save original images | On |

### Inline - Composing

| | |
|---|---|
| Inline Composing | Off |

### Inline - Composing

| | |
|---|---|
| Distortion Corr. | Off |

### Inline - MapIt

| | |
|---|---|
| Save original images | On |
| MapIt | None |
| Flip angle | 5.0 deg |
| Measurements | 1 |
| TR | 2500.0 ms |
| TE | 3.2 ms |

### Sequence - Part 1

| | |
|---|---|
| Introduction | On |
| Dimension | 3D |
| Elliptical scanning | Off |
| Reordering | Linear |
| Asymmetric echo | Allowed |
| Flow comp. | No |
| Multi-slice mode | Single shot |
| Echo spacing | 7.9 ms |
| Bandwidth | 180 Hz/Px |

### Sequence - Part 2

| | |
|---|---|
| RF pulse type | Normal |
| Gradient mode | Normal |
| Excitation | Slab-sel. |
| RF spoiling | On |
| Incr. Gradient spoiling | Off |
| Turbo factor | 64 |

### Sequence - Nuclei

| | |
|---|---|
| TX/RX Nucleus | 1H |
| TX/RX delta frequency | 0 Hz |
| TX Nucleus | None |
| TX delta frequency | 0 Hz |
| Coil elements | OCC |

### Sequence - Assistant

| | |
|---|---|
| Mode | Off |

| \\USER\PHCP\MRS\final\OCC fastestmap lin |
|---|
| TA: 6.1 s PM: FIX VoI: 18 ×30 ×18 mmRel. SNR: 1.00 : fastmp |

### Properties

| | |
|---|---|
| Prio recon | Off |
| Load images to viewer | On |
| Inline movie | Off |
| Auto store images | On |
| Load images to stamp segments | Off |
| Load images to graphic segments | Off |
| Auto open inline display | On |
| Auto close inline display | Off |
| Start measurement without further preparation | Off |
| Wait for user to start | Off |
| Start measurements | Single measurement |

### Routine

| | |
|---|---|
| Position | L1.2 P49.6 H8.9 mm |
| Orientation | Transversal |
| Rotation | 0 deg |
| VoI R >> L | 30 mm |
| VoI R >> L | 30 mm |
| VoI F >> H | 18 mm |
| TR | 2000 ms |
| TE | 46.00 ms |
| Averages | 1 |
| Filter | None |
| Coil elements | OCC |

### Contrast

| | |
|---|---|
| TR | 2000 ms |
| TE | 46.00 ms |
| Tau | 5.00 ms |
| Averages | 1 |
| Excite flip angle | 90 deg |
| Refocus flip angle | 180 deg |
| Measurements | 1 |

### Resolution - Common

| | |
|---|---|
| Vector size | 256 |

### Geometry - Common

| | |
|---|---|
| Position | L1.2 P49.6 H8.9 mm |
| Orientation | Transversal |
| Rotation | 0 deg |
| VoI R >> L | 30 mm |
| VoI A >> P | 18 mm |
| VoI F >> H | 18 mm |

### Geometry - AutoAlign

| | |
|---|---|
| AutoAlign | Head > Brain |
| Initial Position | L1.2 P49.6 H8.9 |
| L | 1.2 mm |
| P | 49.6 mm |
| H | 8.9 mm |
| Initial Rotation | 0.00 deg |
| Initial Orientation | Transversal |

### System - Miscellaneous

| | |
|---|---|
| Positioning mode | FIX |
| Table position | H |
| Table position | 0 mm |
| MSMA | S - C - T |

### System - Miscellaneous

| | |
|---|---|
| Sagittal | R >> L |
| Coronal | A >> P |
| Transversal | F >> H |
| Save uncombined | Off |
| AutoAlign | Head > Brain |
| Coil Select Mode | Off - AutoCoilSelect |

### System - Adjustments

| | |
|---|---|
| B0 Shim mode | Tune up |
| B1 Shim mode | TrueForm |
| Adj. water suppr. | Off |
| Confirm freq. adjustment | Off |
| Assume Dominant Fat | Off |
| Assume Silicone | Off |
| Adjustment Tolerance | Auto |

### System - Adjust Volume

| | |
|---|---|
| Position | L1.2 P49.6 H8.9 mm |
| Orientation | Transversal |
| Rotation | 0.00 deg |
| A >> P | 18 mm |
| R >> L | 30 mm |
| F >> H | 18 mm |
| Reset | Off |

### System - Tx/Rx

| | |
|---|---|
| Frequency 1H | 297.207726 MHz |
| Correction factor | 1 |
| Gain | High |
| Img. Scale Cor. | 1.000 |
| Reset | Off |
| ? Ref. amplitude 1H | 0.000 V |

### Physio - Signal1

| | |
|---|---|
| 1st Signal/Mode | None |
| TR | 2000 ms |

### Sequence - Common

| | |
|---|---|
| Delta frequency | 0.0 ppm |
| Phase cycling | None |
| Bandwidth | 100000 Hz |
| Acquisition duration | 2 ms |

### Sequence - Special

| | |
|---|---|
| Type of fit | Linear 3-proj |
| VoI fit factor | 100 % |
| Force spherical fit VoI | Off Force spherical fit VoI |
| Save plots to database | Off |
| Refocus pulses | Normal |
| Excite pulse duration | 6400 us |
| Refocus pulse duration | 5120 us |
| Bar FoV | 384 mm |
| Bar thickness | 10.0 mm |
| Inversion pulse | Off |
| Multi-echo acquisition | On |
| Number of echoes | 4 |

| \\USER\PHCP\MRS\final\OCC fastestmap all |
|---|
| TA: 0:12 PM: FIX Vol: 18 ×30 ×18 mmRel. SNR: 1.00 : fastmp |

### Properties

| | |
|---|---|
| Prio recon | Off |
| Load images to viewer | On |
| Inline movie | Off |
| Auto store images | On |
| Load images to stamp segments | Off |
| Load images to graphic segments | Off |
| Auto open inline display | On |
| Auto close inline display | Off |
| Start measurement without further preparation | Off |
| Wait for user to start | Off |
| Start measurements | Single measurement |

### Routine

| | |
|---|---|
| Position | L1.2 P49.6 H8.9 mm |
| Orientation | Transversal |
| Rotation | 0 deg |
| VoI R >> L | 30 mm |
| VoI R >> L | 30 mm |
| VoI F >> H | 18 mm |
| TR | 2000 ms |
| TE | 46.00 ms |
| Averages | 1 |
| Filter | None |
| Coil elements | OCC |

### Contrast

| | |
|---|---|
| TR | 2000 ms |
| TE | 46.00 ms |
| Tau | 5.00 ms |
| Averages | 1 |
| Excite flip angle | 90 deg |
| Refocus flip angle | 180 deg |
| Measurements | 1 |

### Resolution - Common

| | |
|---|---|
| Vector size | 256 |

### Geometry - Common

| | |
|---|---|
| Position | L1.2 P49.6 H8.9 mm |
| Orientation | Transversal |
| Rotation | 0 deg |
| VoI R >> L | 30 mm |
| VoI A >> P | 18 mm |
| VoI F >> H | 18 mm |

### Geometry - AutoAlign

| | |
|---|---|
| AutoAlign | Head > Brain |
| Initial Position | L1.2 P49.6 H8.9 |
| L | 1.2 mm |
| P | 49.6 mm |
| H | 8.9 mm |
| Initial Rotation | 0.00 deg |
| Initial Orientation | Transversal |

### System - Miscellaneous

| | |
|---|---|
| Positioning mode | FIX |
| Table position | H |
| Table position | 0 mm |
| MSMA | S - C - T |

### System - Miscellaneous

| | |
|---|---|
| Sagittal | R >> L |
| Coronal | A >> P |
| Transversal | F >> H |
| Save uncombined | Off |
| AutoAlign | Head > Brain |
| Coil Select Mode | Off - AutoCoilSelect |

### System - Adjustments

| | |
|---|---|
| B0 Shim mode | Tune up |
| B1 Shim mode | TrueForm |
| Adj. water suppr. | Off |
| Confirm freq. adjustment | Off |
| Assume Dominant Fat | Off |
| Assume Silicone | Off |
| Adjustment Tolerance | Auto |

### System - Adjust Volume

| | |
|---|---|
| Position | L1.2 P49.6 H8.9 mm |
| Orientation | Transversal |
| Rotation | 0.00 deg |
| A >> P | 18 mm |
| R >> L | 30 mm |
| F >> H | 18 mm |
| Reset | Off |

### System - Tx/Rx

| | |
|---|---|
| Frequency 1H | 297.207726 MHz |
| Correction factor | 1 |
| Gain | High |
| Img. Scale Cor. | 1.000 |
| Reset | Off |
| ? Ref. amplitude 1H | 0.000 V |

### Physio - Signal1

| | |
|---|---|
| 1st Signal/Mode | None |
| TR | 2000 ms |

### Sequence - Common

| | |
|---|---|
| Delta frequency | 0.0 ppm |
| Phase cycling | None |
| Bandwidth | 100000 Hz |
| Acquisition duration | 2 ms |

### Sequence - Special

| | |
|---|---|
| Type of fit | Full 6-proj |
| VoI fit factor | 100 % |
| Force spherical fit VoI | Off Force spherical fit VoI |
| Save plots to database | Off |
| Refocus pulses | Normal |
| Excite pulse duration | 6400 us |
| Refocus pulse duration | 5120 us |
| Bar FoV | 384 mm |
| Bar thickness | 10.0 mm |
| Inversion pulse | Off |
| Multi-echo acquisition | On |
| Number of echoes | 4 |

| \\USER\PHCP\MRS\final\OCC fastestmap all |
|---|
| TA: 0:12 PM: FIX Vol: 18 ×30 ×18 mmRel. SNR: 1.00 : fastmp |

### Properties

| | |
|---|---|
| Prio recon | Off |
| Load images to viewer | On |
| Inline movie | Off |
| Auto store images | On |
| Load images to stamp segments | Off |
| Load images to graphic segments | Off |
| Auto open inline display | Off |
| Auto close inline display | Off |
| Start measurement without further preparation | Off |
| Wait for user to start | Off |
| Start measurements | Single measurement |

### Routine

| | |
|---|---|
| Position | L1.2 P49.6 H8.9 mm |
| Orientation | Transversal |
| Rotation | 0 deg |
| Vol R >> L | 30 mm |
| Vol R >> L | 30 mm |
| Vol F >> H | 18 mm |
| TR | 2000 ms |
| TE | 46.00 ms |
| Averages | 1 |
| Filter | None |
| Coil elements | OCC |

### Contrast

| | |
|---|---|
| TR | 2000 ms |
| TE | 46.00 ms |
| Tau | 5.00 ms |
| Averages | 1 |
| Excite flip angle | 90 deg |
| Refocus flip angle | 180 deg |
| Measurements | 1 |

### Resolution - Common

| | |
|---|---|
| Vector size | 256 |

### Geometry - Common

| | |
|---|---|
| Position | L1.2 P49.6 H8.9 mm |
| Orientation | Transversal |
| Rotation | 0 deg |
| Vol R >> L | 30 mm |
| Vol A >> P | 18 mm |
| Vol F >> H | 18 mm |

### Geometry - AutoAlign

| | |
|---|---|
| AutoAlign | Head > Brain |
| Initial Position | L1.2 P49.6 H8.9 |
| L | 1.2 mm |
| P | 49.6 mm |
| H | 8.9 mm |
| Initial Rotation | 0.00 deg |
| Initial Orientation | Transversal |

### System - Miscellaneous

| | |
|---|---|
| Positioning mode | FIX |
| Table position | H |
| Table position | 0 mm |
| MSMA | S - C - T |

### System - Miscellaneous

| | |
|---|---|
| Sagittal | R >> L |
| Coronal | A >> P |
| Transversal | F >> H |
| Save uncombined | Off |
| AutoAlign | Head > Brain |
| Coil Select Mode | Off - AutoCoilSelect |

### System - Adjustments

| | |
|---|---|
| B0 Shim mode | Tune up |
| B1 Shim mode | TrueForm |
| Adj. water suppr. | Off |
| Confirm freq. adjustment | Off |
| Assume Dominant Fat | Off |
| Assume Silicone | Off |
| Adjustment Tolerance | Auto |

### System - Adjust Volume

| | |
|---|---|
| Position | L1.2 P49.6 H8.9 mm |
| Orientation | Transversal |
| Rotation | 0.00 deg |
| A >> P | 18 mm |
| R >> L | 30 mm |
| F >> H | 18 mm |
| Reset | Off |

### System - Tx/Rx

| | |
|---|---|
| Frequency 1H | 297.207726 MHz |
| Correction factor | 1 |
| Gain | High |
| Img. Scale Cor. | 1.000 |
| Reset | Off |
| ? Ref. amplitude 1H | 0.000 V |

### Physio - Signal1

| | |
|---|---|
| 1st Signal/Mode | None |
| TR | 2000 ms |

### Sequence - Common

| | |
|---|---|
| Delta frequency | 0.0 ppm |
| Phase cycling | None |
| Bandwidth | 100000 Hz |
| Acquisition duration | 2 ms |

### Sequence - Special

| | |
|---|---|
| Type of fit | Full 6-proj |
| Vol fit factor | 100 % |
| Force spherical fit Vol | Off Force spherical fit Vol |
| Save plots to database | Off |
| Refocus pulses | Normal |
| Excite pulse duration | 6400 us |
| Refocus pulse duration | 5120 us |
| Bar FoV | 384 mm |
| Bar thickness | 10.0 mm |
| Inversion pulse | Off |
| Multi-echo acquisition | On |
| Number of echoes | 4 |

**\\USER\PHCP\MRS\final\OCC fastestmap all**

TA: 0:12 PM: FIX Vol: 18 ×30 ×18 mmRel. SNR: 1.00 : fastmp

### Properties

| | |
|---|---|
| Prio recon | Off |
| Load images to viewer | On |
| Inline movie | Off |
| Auto store images | On |
| Load images to stamp segments | Off |
| Load images to graphic segments | Off |
| Auto open inline display | Off |
| Auto close inline display | Off |
| Start measurement without further preparation | Off |
| Wait for user to start | Off |
| Start measurements | Single measurement |

### Routine

| | |
|---|---|
| Position | L1.2 P49.6 H8.9 mm |
| Orientation | Transversal |
| Rotation | 0 deg |
| Vol R >> L | 30 mm |
| Vol R >> L | 30 mm |
| Vol F >> H | 18 mm |
| TR | 2000 ms |
| TE | 46.00 ms |
| Averages | 1 |
| Filter | None |
| Coil elements | OCC |

### Contrast

| | |
|---|---|
| TR | 2000 ms |
| TE | 46.00 ms |
| Tau | 5.00 ms |
| Averages | 1 |
| Excite flip angle | 90 deg |
| Refocus flip angle | 180 deg |
| Measurements | 1 |

### Resolution - Common

| | |
|---|---|
| Vector size | 256 |

### Geometry - Common

| | |
|---|---|
| Position | L1.2 P49.6 H8.9 mm |
| Orientation | Transversal |
| Rotation | 0 deg |
| Vol R >> L | 30 mm |
| Vol A >> P | 18 mm |
| Vol F >> H | 18 mm |

### Geometry - AutoAlign

| | |
|---|---|
| AutoAlign | Head > Brain |
| Initial Position | L1.2 P49.6 H8.9 |
| L | 1.2 mm |
| P | 49.6 mm |
| H | 8.9 mm |
| Initial Rotation | 0.00 deg |
| Initial Orientation | Transversal |

### System - Miscellaneous

| | |
|---|---|
| Positioning mode | FIX |
| Table position | H |
| Table position | 0 mm |
| MSMA | S - C - T |
| Sagittal | R >> L |
| Coronal | A >> P |
| Transversal | F >> H |
| Save uncombined | Off |
| AutoAlign | Head > Brain |
| Coil Select Mode | Off - AutoCoilSelect |

### System - Adjustments

| | |
|---|---|
| B0 Shim mode | Tune up |
| B1 Shim mode | TrueForm |
| Adj. water suppr. | Off |
| Confirm freq. adjustment | Off |
| Assume Dominant Fat | Off |
| Assume Silicone | Off |
| Adjustment Tolerance | Auto |

### System - Adjust Volume

| | |
|---|---|
| Position | L1.2 P49.6 H8.9 mm |
| Orientation | Transversal |
| Rotation | 0.00 deg |
| A >> P | 18 mm |
| R >> L | 30 mm |
| F >> H | 18 mm |
| Reset | Off |

### System - Tx/Rx

| | |
|---|---|
| Frequency 1H | 297.207726 MHz |
| Correction factor | 1 |
| Gain | High |
| Img. Scale Cor. | 1.000 |
| Reset | Off |
| ? Ref. amplitude 1H | 0.000 V |

### Physio - Signal1

| | |
|---|---|
| 1st Signal/Mode | None |
| TR | 2000 ms |

### Sequence - Common

| | |
|---|---|
| Delta frequency | 0.0 ppm |
| Phase cycling | None |
| Bandwidth | 100000 Hz |
| Acquisition duration | 2 ms |

### Sequence - Special

| | |
|---|---|
| Type of fit | Full 6-proj |
| Vol fit factor | 100 % |
| Force spherical fit Vol | Off Force spherical fit Vol |
| Save plots to database | Off |
| Refocus pulses | Normal |
| Excite pulse duration | 6400 us |
| Refocus pulse duration | 5120 us |
| Bar FoV | 384 mm |
| Bar thickness | 10.0 mm |
| Inversion pulse | Off |
| Multi-echo acquisition | On |
| Number of echoes | 4 |

| \\USER\PHCP\MRS\final\OCC fastestmap lin6 |
|---|
| TA: 0:24 PM: FIX Vol: 18 ×30 ×18 mmRel. SNR: 1.00 : fastmp |

### Properties

| | |
|---|---|
| Prio recon | Off |
| Load images to viewer | On |
| Inline movie | Off |
| Auto store images | On |
| Load images to stamp segments | Off |
| Load images to graphic segments | Off |
| Auto open inline display | On |
| Auto close inline display | Off |
| Start measurement without further preparation | Off |
| Wait for user to start | Off |
| Start measurements | Single measurement |

### Routine

| | |
|---|---|
| Position | L1.2 P49.6 H8.9 mm |
| Orientation | Transversal |
| Rotation | 0 deg |
| VoI R >> L | 30 mm |
| VoI R >> L | 30 mm |
| VoI F >> H | 18 mm |
| TR | 2000 ms |
| TE | 46.00 ms |
| Averages | 1 |
| Filter | None |
| Coil elements | OCC |

### Contrast

| | |
|---|---|
| TR | 2000 ms |
| TE | 46.00 ms |
| Tau | 20.00 ms |
| Averages | 1 |
| Excite flip angle | 90 deg |
| Refocus flip angle | 180 deg |
| Measurements | 1 |

### Resolution - Common

| | |
|---|---|
| Vector size | 256 |

### Geometry - Common

| | |
|---|---|
| Position | L1.2 P49.6 H8.9 mm |
| Orientation | Transversal |
| Rotation | 0 deg |
| VoI R >> L | 30 mm |
| VoI A >> P | 18 mm |
| VoI F >> H | 18 mm |

### Geometry - AutoAlign

| | |
|---|---|
| AutoAlign | Head > Brain |
| Initial Position | L1.2 P49.6 H8.9 |
| L | 1.2 mm |
| P | 49.6 mm |
| H | 8.9 mm |
| Initial Rotation | 0.00 deg |
| Initial Orientation | Transversal |

### System - Miscellaneous

| | |
|---|---|
| Positioning mode | FIX |
| Table position | H |
| Table position | 0 mm |
| MSMA | S - C - T |

### System - Miscellaneous

| | |
|---|---|
| Sagittal | R >> L |
| Coronal | A >> P |
| Transversal | F >> H |
| Save uncombined | Off |
| AutoAlign | Head > Brain |
| Coil Select Mode | Off - AutoCoilSelect |

### System - Adjustments

| | |
|---|---|
| B0 Shim mode | Tune up |
| B1 Shim mode | TrueForm |
| Adj. water suppr. | Off |
| Confirm freq. adjustment | Off |
| Assume Dominant Fat | Off |
| Assume Silicone | Off |
| Adjustment Tolerance | Auto |

### System - Adjust Volume

| | |
|---|---|
| Position | L1.2 P49.6 H8.9 mm |
| Orientation | Transversal |
| Rotation | 0.00 deg |
| A >> P | 18 mm |
| R >> L | 30 mm |
| F >> H | 18 mm |
| Reset | Off |

### System - Tx/Rx

| | |
|---|---|
| Frequency 1H | 297.207726 MHz |
| Correction factor | 1 |
| Gain | High |
| Img. Scale Cor. | 1.000 |
| Reset | Off |
| ? Ref. amplitude 1H | 0.000 V |

### Physio - Signal1

| | |
|---|---|
| 1st Signal/Mode | None |
| TR | 2000 ms |

### Sequence - Common

| | |
|---|---|
| Delta frequency | 0.0 ppm |
| Phase cycling | None |
| Bandwidth | 100000 Hz |
| Acquisition duration | 2 ms |

### Sequence - Special

| | |
|---|---|
| Type of fit | Linear 6-proj |
| VoI fit factor | 100 % |
| Force spherical fit VoI | Off Force spherical fit VoI |
| Save plots to database | Off |
| Refocus pulses | Normal |
| Excite pulse duration | 6400 us |
| Refocus pulse duration | 5120 us |
| Bar FoV | 384 mm |
| Bar thickness | 10.0 mm |
| Inversion pulse | Off |
| Multi-echo acquisition | Off |

| \\USER\PHCP\MRS\final\OCC linewidth check |
|---|
| TA: 0:23 PM: FIX Vol: 18 ×30 ×18 mmRel. SNR: 1.00 : steam |

### Properties

| | |
|---|---|
| Prio recon | Off |
| Load images to viewer | On |
| Inline movie | Off |
| Auto store images | On |
| Load images to stamp segments | Off |
| Load images to graphic segments | Off |
| Auto open inline display | Off |
| Auto close inline display | Off |
| Start measurement without further preparation | Off |
| Wait for user to start | Off |
| Start measurements | Single measurement |

### Routine

| | |
|---|---|
| Position | L1.2 P49.6 H8.9 mm |
| Orientation | Transversal |
| Rotation | 0 deg |
| Vol R >> L | 30 mm |
| Vol R >> L | 30 mm |
| Vol F >> H | 18 mm |
| TR | 5000 ms |
| TE | 8.00 ms |
| Averages | 1 |
| Filter | None |
| Coil elements | OCC |

### Contrast

| | |
|---|---|
| TR | 5000 ms |
| TE | 8.00 ms |
| TM | 32.00 ms |
| Averages | 1 |
| Flip angle | 90 deg |
| VAPOR | Only RF off |
| VAPOR suppr. | Water suppr. |
| Water s. BW | 135 Hz |
| Water s. delta pos. | 0.00 ppm |
| Measurements | 1 |

### Resolution - Common

| | |
|---|---|
| Vector size | 2048 |

### Geometry - Common

| | |
|---|---|
| Position | L1.2 P49.6 H8.9 mm |
| Orientation | Transversal |
| Rotation | 0 deg |
| Vol R >> L | 30 mm |
| Vol A >> P | 18 mm |
| Vol F >> H | 18 mm |

### Geometry - AutoAlign

| | |
|---|---|
| AutoAlign | Head > Brain |
| Initial Position | L1.2 P49.6 H8.9 |
| L | 1.2 mm |
| P | 49.6 mm |
| H | 8.9 mm |
| Initial Rotation | 0.00 deg |
| Initial Orientation | Transversal |

### System - Miscellaneous

| | |
|---|---|
| Positioning mode | FIX |

### System - Miscellaneous

| | |
|---|---|
| Table position | H |
| Table position | 0 mm |
| MSMA | S - C - T |
| Sagittal | R >> L |
| Coronal | A >> P |
| Transversal | F >> H |
| Save uncombined | Off |
| AutoAlign | Head > Brain |
| Coil Select Mode | Off - AutoCoilSelect |

### System - Adjustments

| | |
|---|---|
| B0 Shim mode | Brain |
| B1 Shim mode | TrueForm |
| Adj. water suppr. | Off |
| Confirm freq. adjustment | Off |
| Assume Dominant Fat | Off |
| Assume Silicone | Off |
| Adjustment Tolerance | Auto |

### System - Adjust Volume

| | |
|---|---|
| Position | L1.2 P49.6 H8.9 mm |
| Orientation | Transversal |
| Rotation | 0.00 deg |
| A >> P | 18 mm |
| R >> L | 30 mm |
| F >> H | 18 mm |
| Reset | Off |

### System - Tx/Rx

| | |
|---|---|
| Frequency 1H | 297.207726 MHz |
| Correction factor | 1 |
| Gain | High |
| Img. Scale Cor. | 1.000 |
| Reset | Off |
| ? Ref. amplitude 1H | 0.000 V |

### Physio - Signal1

| | |
|---|---|
| 1st Signal/Mode | None |
| TR | 5000 ms |

### Sequence - Common

| | |
|---|---|
| Introduction | On |
| Preparation scans | 2 |
| Delta frequency | 0.0 ppm |
| Phase cycling | Auto |
| Bandwidth | 6000 Hz |
| Acquisition duration | 341 ms |
| Remove oversampling | On |

### Sequence - Nuclei

| | |
|---|---|
| TX/RX Nucleus | 1H |
| TX/RX delta frequency | 0 Hz |
| TX Nucleus | None |
| TX delta frequency | 0 Hz |
| Coil elements | OCC |

### Sequence - Special

| | |
|---|---|
| RF pulse duration | 1280 us |
| Spoiler max. amplitude | 30.0 mT/m |
| Refocus grad. factor | 1.00 |

### Sequence - Special

| | |
|---|---|
| OVS slab thickness | 100.0 mm |
| OVS slab pos. offset | 5.0 mm |
| Spoiler duration | 1500 us |
| Acq. window shift | 200 us |
| Min. settling delay | 100 us |
| Gradient ramp time | 200 us |
| VAPOR flip angle | 60 deg |
| VAPOR delay 8 | 16 ms |
| VAPOR delay 7 | 66 ms |
| OVS pulse duration | 2560 us |
| OVS flip angle RO | 90 deg |
| OVS flip angle PH | 90 deg |
| OVS flip angle SL | 90 deg |
| VAPOR delay 6 | 71 ms |
| VAPOR delay 5 | 112 ms |
| VAPOR delay 4 | 115 ms |
| VAPOR delay 3 | 132 ms |
| VAPOR delay 2 | 110 ms |
| VAPOR delay 1 | 160 ms |
| Enable OVS | On |
| Send ref. scans | Off |
| Inversion pulse | Off |
| Symmetric RF pulses | On |
| Invert SS grad. pol. | Off |
| Shift RO frequency | Off |
| Debug loop type | None |

| \\USER\PHCP\MRS\final\OCC steam eja cal fa 50 10 7 |
|---|
| TA: 0:53 PM: FIX Vol: 18 ×30 ×18 mmRel. SNR: 1.00 : steam |

### Properties

| | |
|---|---|
| Prio recon | Off |
| Load images to viewer | On |
| Inline movie | Off |
| Auto store images | On |
| Load images to stamp segments | Off |
| Load images to graphic segments | Off |
| Auto open inline display | On |
| Auto close inline display | Off |
| Start measurement without further preparation | Off |
| Wait for user to start | Off |
| Start measurements | Single measurement |

### Routine

| | |
|---|---|
| Position | L1.2 P49.6 H8.9 mm |
| Orientation | Transversal |
| Rotation | 0 deg |
| Vol R >> L | 30 mm |
| Vol R >> L | 30 mm |
| Vol F >> H | 18 mm |
| TR | 5000 ms |
| TE | 8.00 ms |
| Averages | 1 |
| Filter | None |
| Coil elements | OCC |

### Contrast

| | |
|---|---|
| TR | 5000 ms |
| TE | 8.00 ms |
| TM | 32.00 ms |
| Averages | 1 |
| Flip angle | 50 deg |
| VAPOR | Only RF off |
| VAPOR suppr. | Water suppr. |
| Water s. BW | 135 Hz |
| Water s. delta pos. | 0.00 ppm |
| Measurements | 7 |

### Resolution - Common

| | |
|---|---|
| Vector size | 2048 |

### Geometry - Common

| | |
|---|---|
| Position | L1.2 P49.6 H8.9 mm |
| Orientation | Transversal |
| Rotation | 0 deg |
| Vol R >> L | 30 mm |
| Vol A >> P | 18 mm |
| Vol F >> H | 18 mm |

### Geometry - AutoAlign

| | |
|---|---|
| AutoAlign | Head > Brain |
| Initial Position | L1.2 P49.6 H8.9 |
| L | 1.2 mm |
| P | 49.6 mm |
| H | 8.9 mm |
| Initial Rotation | 0.00 deg |
| Initial Orientation | Transversal |

### System - Miscellaneous

| | |
|---|---|
| Positioning mode | FIX |

### System - Miscellaneous

| | |
|---|---|
| Table position | H |
| Table position | 0 mm |
| MSMA | S - C - T |
| Sagittal | R >> L |
| Coronal | A >> P |
| Transversal | F >> H |
| Save uncombined | Off |
| AutoAlign | Head > Brain |
| Coil Select Mode | Off - AutoCoilSelect |

### System - Adjustments

| | |
|---|---|
| B0 Shim mode | Brain |
| B1 Shim mode | TrueForm |
| Adj. water suppr. | Off |
| Confirm freq. adjustment | Off |
| Assume Dominant Fat | Off |
| Assume Silicone | Off |
| Adjustment Tolerance | Auto |

### System - Adjust Volume

| | |
|---|---|
| Position | L1.2 P49.6 H8.9 mm |
| Orientation | Transversal |
| Rotation | 0.00 deg |
| A >> P | 18 mm |
| R >> L | 30 mm |
| F >> H | 18 mm |
| Reset | Off |

### System - Tx/Rx

| | |
|---|---|
| Frequency 1H | 297.207726 MHz |
| Correction factor | 1 |
| Gain | High |
| Img. Scale Cor. | 1.000 |
| Reset | Off |
| ? Ref. amplitude 1H | 0.000 V |

### Physio - Signal1

| | |
|---|---|
| 1st Signal/Mode | None |
| TR | 5000 ms |

### Sequence - Common

| | |
|---|---|
| Introduction | On |
| Preparation scans | 2 |
| Delta frequency | 0.0 ppm |
| Phase cycling | Auto |
| Bandwidth | 6000 Hz |
| Acquisition duration | 341 ms |
| Remove oversampling | On |

### Sequence - Nuclei

| | |
|---|---|
| TX/RX Nucleus | 1H |
| TX/RX delta frequency | 0 Hz |
| TX Nucleus | None |
| TX delta frequency | 0 Hz |
| Coil elements | OCC |

### Sequence - Special

| | |
|---|---|
| RF pulse duration | 1520 us |
| Spoiler max. amplitude | 30.0 mT/m |
| Refocus grad. factor | 1.00 |

### Sequence - Special

| | |
|---|---|
| OVS slab thickness | 100.0 mm |
| OVS slab pos. offset | 5.0 mm |
| Spoiler duration | 1500 us |
| Acq. window shift | 200 us |
| Min. settling delay | 100 us |
| Gradient ramp time | 200 us |
| VAPOR flip angle | 60 deg |
| VAPOR delay 8 | 16 ms |
| VAPOR delay 7 | 66 ms |
| OVS pulse duration | 2560 us |
| OVS flip angle RO | 90 deg |
| OVS flip angle PH | 90 deg |
| OVS flip angle SL | 90 deg |
| VAPOR delay 6 | 71 ms |
| VAPOR delay 5 | 112 ms |
| VAPOR delay 4 | 115 ms |
| VAPOR delay 3 | 132 ms |
| VAPOR delay 2 | 110 ms |
| VAPOR delay 1 | 160 ms |
| Enable OVS | On |
| Weighted averages | Off |
| Send ref. scans | Off |
| Inversion pulse | Off |
| Symmetric RF pulses | On |
| Invert SS grad. pol. | Off |
| Shift RO frequency | Off |
| Debug loop type | Flip angle |
| Flip angle inc. | 10 deg |
| Measurements | 7 |

| \\USER\PHCP\MRS\final\OCC steam eja cal fa 80 3 10 |
|---|
| TA: 1:08 PM: FIX Vol: 18 ×30 ×18 mmRel. SNR: 1.00 : steam |

### Properties

| | |
|---|---|
| Prio recon | Off |
| Load images to viewer | On |
| Inline movie | Off |
| Auto store images | On |
| Load images to stamp segments | Off |
| Load images to graphic segments | Off |
| Auto open inline display | Off |
| Auto close inline display | Off |
| Start measurement without further preparation | Off |
| Wait for user to start | Off |
| Start measurements | Single measurement |

### Routine

| | |
|---|---|
| Position | L1.2 P49.6 H8.9 mm |
| Orientation | Transversal |
| Rotation | 0 deg |
| Vol R >> L | 30 mm |
| Vol R >> L | 30 mm |
| Vol F >> H | 18 mm |
| TR | 5000 ms |
| TE | 8.00 ms |
| Averages | 1 |
| Filter | None |
| Coil elements | OCC |

### Contrast

| | |
|---|---|
| TR | 5000 ms |
| TE | 8.00 ms |
| TM | 32.00 ms |
| Averages | 1 |
| Flip angle | 80 deg |
| VAPOR | Only RF off |
| VAPOR suppr. | Water suppr. |
| Water s. BW | 135 Hz |
| Water s. delta pos. | 0.00 ppm |
| Measurements | 10 |

### Resolution - Common

| | |
|---|---|
| Vector size | 2048 |

### Geometry - Common

| | |
|---|---|
| Position | L1.2 P49.6 H8.9 mm |
| Orientation | Transversal |
| Rotation | 0 deg |
| Vol R >> L | 30 mm |
| Vol A >> P | 18 mm |
| Vol F >> H | 18 mm |

### Geometry - AutoAlign

| | |
|---|---|
| AutoAlign | Head > Brain |
| Initial Position | L1.2 P49.6 H8.9 |
| L | 1.2 mm |
| P | 49.6 mm |
| H | 8.9 mm |
| Initial Rotation | 0.00 deg |
| Initial Orientation | Transversal |

### System - Miscellaneous

| | |
|---|---|
| Positioning mode | FIX |

### System - Miscellaneous

| | |
|---|---|
| Table position | H |
| Table position | 0 mm |
| MSMA | S - C - T |
| Sagittal | R >> L |
| Coronal | A >> P |
| Transversal | F >> H |
| Save uncombined | Off |
| AutoAlign | Head > Brain |
| Coil Select Mode | Off - AutoCoilSelect |

### System - Adjustments

| | |
|---|---|
| B0 Shim mode | Brain |
| B1 Shim mode | TrueForm |
| Adj. water suppr. | Off |
| Confirm freq. adjustment | Off |
| Assume Dominant Fat | Off |
| Assume Silicone | Off |
| Adjustment Tolerance | Auto |

### System - Adjust Volume

| | |
|---|---|
| Position | L1.2 P49.6 H8.9 mm |
| Orientation | Transversal |
| Rotation | 0.00 deg |
| A >> P | 18 mm |
| R >> L | 30 mm |
| F >> H | 18 mm |
| Reset | Off |

### System - Tx/Rx

| | |
|---|---|
| Frequency 1H | 297.207726 MHz |
| Correction factor | 1 |
| Gain | High |
| Img. Scale Cor. | 1.000 |
| Reset | Off |
| ? Ref. amplitude 1H | 0.000 V |

### Physio - Signal1

| | |
|---|---|
| 1st Signal/Mode | None |
| TR | 5000 ms |

### Sequence - Common

| | |
|---|---|
| Introduction | On |
| Preparation scans | 2 |
| Delta frequency | 0.0 ppm |
| Phase cycling | Auto |
| Bandwidth | 6000 Hz |
| Acquisition duration | 341 ms |
| Remove oversampling | On |

### Sequence - Nuclei

| | |
|---|---|
| TX/RX Nucleus | 1H |
| TX/RX delta frequency | 0 Hz |
| TX Nucleus | None |
| TX delta frequency | 0 Hz |
| Coil elements | OCC |

### Sequence - Special

| | |
|---|---|
| RF pulse duration | 1520 us |
| Spoiler max. amplitude | 30.0 mT/m |
| Refocus grad. factor | 1.00 |

### Sequence - Special

| | |
|---|---|
| OVS slab thickness | 100.0 mm |
| OVS slab pos. offset | 5.0 mm |
| Spoiler duration | 1500 us |
| Acq. window shift | 200 us |
| Min. settling delay | 100 us |
| Gradient ramp time | 200 us |
| VAPOR flip angle | 60 deg |
| VAPOR delay 8 | 16 ms |
| VAPOR delay 7 | 66 ms |
| OVS pulse duration | 2560 us |
| OVS flip angle RO | 90 deg |
| OVS flip angle PH | 90 deg |
| OVS flip angle SL | 90 deg |
| VAPOR delay 6 | 71 ms |
| VAPOR delay 5 | 112 ms |
| VAPOR delay 4 | 115 ms |
| VAPOR delay 3 | 132 ms |
| VAPOR delay 2 | 110 ms |
| VAPOR delay 1 | 160 ms |
| Enable OVS | On |
| Weighted averages | Off |
| Send ref. scans | Off |
| Inversion pulse | Off |
| Symmetric RF pulses | On |
| Invert SS grad. pol. | Off |
| Shift RO frequency | Off |
| Debug loop type | Flip angle |
| Flip angle inc. | 3 deg |
| Measurements | 10 |

| \\USER\PHCP\MRS\final\OCC steam eja ws check |
|---|
| TA: 0:38 PM: FIX Vol: 18 ×30 ×18 mmRel. SNR: 1.00 : steam |

### Properties

| | |
|---|---|
| Prio recon | Off |
| Load images to viewer | On |
| Inline movie | Off |
| Auto store images | On |
| Load images to stamp segments | Off |
| Load images to graphic segments | Off |
| Auto open inline display | Off |
| Auto close inline display | Off |
| Start measurement without further preparation | Off |
| Wait for user to start | Off |
| Start measurements | Single measurement |

### Routine

| | |
|---|---|
| Position | L1.2 P49.6 H8.9 mm |
| Orientation | Transversal |
| Rotation | 0 deg |
| Vol R >> L | 30 mm |
| Vol R >> L | 30 mm |
| Vol F >> H | 18 mm |
| TR | 5000 ms |
| TE | 8.00 ms |
| Averages | 4 |
| Filter | None |
| Coil elements | OCC |

### Contrast

| | |
|---|---|
| TR | 5000 ms |
| TE | 8.00 ms |
| TM | 32.00 ms |
| Averages | 4 |
| Flip angle | 90 deg |
| VAPOR | Enabled |
| VAPOR suppr. | Water suppr. |
| Water s. BW | 135 Hz |
| Water s. delta pos. | 0.00 ppm |
| Measurements | 1 |

### Resolution - Common

| | |
|---|---|
| Vector size | 2048 |

### Geometry - Common

| | |
|---|---|
| Position | L1.2 P49.6 H8.9 mm |
| Orientation | Transversal |
| Rotation | 0 deg |
| Vol R >> L | 30 mm |
| Vol A >> P | 18 mm |
| Vol F >> H | 18 mm |

### Geometry - AutoAlign

| | |
|---|---|
| AutoAlign | Head > Brain |
| Initial Position | L1.2 P49.6 H8.9 |
| L | 1.2 mm |
| P | 49.6 mm |
| H | 8.9 mm |
| Initial Rotation | 0.00 deg |
| Initial Orientation | Transversal |

### System - Miscellaneous

| | |
|---|---|
| Positioning mode | FIX |

### System - Miscellaneous

| | |
|---|---|
| Table position | H |
| Table position | 0 mm |
| MSMA | S - C - T |
| Sagittal | R >> L |
| Coronal | A >> P |
| Transversal | F >> H |
| Save uncombined | Off |
| AutoAlign | Head > Brain |
| Coil Select Mode | Off - AutoCoilSelect |

### System - Adjustments

| | |
|---|---|
| B0 Shim mode | Brain |
| B1 Shim mode | TrueForm |
| Adj. water suppr. | Off |
| Confirm freq. adjustment | Off |
| Assume Dominant Fat | Off |
| Assume Silicone | Off |
| Adjustment Tolerance | Auto |

### System - Adjust Volume

| | |
|---|---|
| Position | L1.2 P49.6 H8.9 mm |
| Orientation | Transversal |
| Rotation | 0.00 deg |
| A >> P | 18 mm |
| R >> L | 30 mm |
| F >> H | 18 mm |
| Reset | Off |

### System - Tx/Rx

| | |
|---|---|
| Frequency 1H | 297.207726 MHz |
| Correction factor | 1 |
| Gain | High |
| Img. Scale Cor. | 1.000 |
| Reset | Off |
| ? Ref. amplitude 1H | 0.000 V |

### Physio - Signal1

| | |
|---|---|
| 1st Signal/Mode | None |
| TR | 5000 ms |

### Sequence - Common

| | |
|---|---|
| Introduction | On |
| Preparation scans | 2 |
| Delta frequency | -1.7 ppm |
| Phase cycling | Auto |
| Bandwidth | 6000 Hz |
| Acquisition duration | 341 ms |
| Remove oversampling | On |

### Sequence - Nuclei

| | |
|---|---|
| TX/RX Nucleus | 1H |
| TX/RX delta frequency | 0 Hz |
| TX Nucleus | None |
| TX delta frequency | 0 Hz |
| Coil elements | OCC |

### Sequence - Special

| | |
|---|---|
| RF pulse duration | 1280 us |
| Spoiler max. amplitude | 30.0 mT/m |
| Refocus grad. factor | 1.00 |

### Sequence - Special

| | |
|---|---|
| OVS slab thickness | 100.0 mm |
| OVS slab pos. offset | 5.0 mm |
| Spoiler duration | 1500 us |
| Acq. window shift | 200 us |
| Min. settling delay | 100 us |
| Gradient ramp time | 200 us |
| VAPOR flip angle | 72 deg |
| VAPOR delay 8 | 16 ms |
| VAPOR delay 7 | 66 ms |
| OVS pulse duration | 2560 us |
| OVS flip angle RO | 90 deg |
| OVS flip angle PH | 90 deg |
| OVS flip angle SL | 90 deg |
| VAPOR delay 6 | 71 ms |
| VAPOR delay 5 | 112 ms |
| VAPOR delay 4 | 115 ms |
| VAPOR delay 3 | 132 ms |
| VAPOR delay 2 | 110 ms |
| VAPOR delay 1 | 160 ms |
| Enable OVS | On |
| Resolve averages | On |
| Send ref. scans | Off |
| Inversion pulse | Off |
| Symmetric RF pulses | On |
| Invert SS grad. pol. | Off |
| Shift RO frequency | Off |
| Debug loop type | None |

| \\USER\PHCP\MRS\final\OCC steam eja metab |
|---|
| TA: 8:33 PM: FIX Vol: 18 ×30 ×18 mmRel. SNR: 1.00 : steam |

### Properties

| | |
|---|---|
| Prio recon | Off |
| Load images to viewer | On |
| Inline movie | Off |
| Auto store images | On |
| Load images to stamp segments | Off |
| Load images to graphic segments | Off |
| Auto open inline display | Off |
| Auto close inline display | Off |
| Start measurement without further preparation | Off |
| Wait for user to start | Off |
| Start measurements | Single measurement |

### Routine

| | |
|---|---|
| Position | L1.2 P49.6 H8.9 mm |
| Orientation | Transversal |
| Rotation | 0 deg |
| Vol R >> L | 30 mm |
| Vol R >> L | 30 mm |
| Vol F >> H | 18 mm |
| TR | 5000 ms |
| TE | 8.00 ms |
| Averages | 96 |
| Filter | None |
| Coil elements | OCC |

### Contrast

| | |
|---|---|
| TR | 5000 ms |
| TE | 8.00 ms |
| TM | 32.00 ms |
| Averages | 96 |
| Flip angle | 90 deg |
| VAPOR | Enabled |
| VAPOR suppr. | Water suppr. |
| Water s. BW | 135 Hz |
| Water s. delta pos. | 0.00 ppm |
| Measurements | 1 |

### Resolution - Common

| | |
|---|---|
| Vector size | 2048 |

### Geometry - Common

| | |
|---|---|
| Position | L1.2 P49.6 H8.9 mm |
| Orientation | Transversal |
| Rotation | 0 deg |
| Vol R >> L | 30 mm |
| Vol A >> P | 18 mm |
| Vol F >> H | 18 mm |

### Geometry - AutoAlign

| | |
|---|---|
| AutoAlign | Head > Brain |
| Initial Position | L1.2 P49.6 H8.9 |
| L | 1.2 mm |
| P | 49.6 mm |
| H | 8.9 mm |
| Initial Rotation | 0.00 deg |
| Initial Orientation | Transversal |

### System - Miscellaneous

| | |
|---|---|
| Positioning mode | FIX |

### System - Miscellaneous

| | |
|---|---|
| Table position | H |
| Table position | 0 mm |
| MSMA | S - C - T |
| Sagittal | R >> L |
| Coronal | A >> P |
| Transversal | F >> H |
| Save uncombined | Off |
| AutoAlign | Head > Brain |
| Coil Select Mode | Off - AutoCoilSelect |

### System - Adjustments

| | |
|---|---|
| B0 Shim mode | Brain |
| B1 Shim mode | TrueForm |
| Adj. water suppr. | Off |
| Confirm freq. adjustment | Off |
| Assume Dominant Fat | Off |
| Assume Silicone | Off |
| Adjustment Tolerance | Auto |

### System - Adjust Volume

| | |
|---|---|
| Position | L1.2 P49.6 H8.9 mm |
| Orientation | Transversal |
| Rotation | 0.00 deg |
| A >> P | 18 mm |
| R >> L | 30 mm |
| F >> H | 18 mm |
| Reset | Off |

### System - Tx/Rx

| | |
|---|---|
| Frequency 1H | 297.207726 MHz |
| Correction factor | 1 |
| Gain | High |
| Img. Scale Cor. | 1.000 |
| Reset | Off |
| ? Ref. amplitude 1H | 0.000 V |

### Physio - Signal1

| | |
|---|---|
| 1st Signal/Mode | None |
| TR | 5000 ms |

### Sequence - Common

| | |
|---|---|
| Introduction | On |
| Preparation scans | 2 |
| Delta frequency | -1.7 ppm |
| Phase cycling | Auto |
| Bandwidth | 6000 Hz |
| Acquisition duration | 341 ms |
| Remove oversampling | On |

### Sequence - Nuclei

| | |
|---|---|
| TX/RX Nucleus | 1H |
| TX/RX delta frequency | 0 Hz |
| TX Nucleus | None |
| TX delta frequency | 0 Hz |
| Coil elements | OCC |

### Sequence - Special

| | |
|---|---|
| RF pulse duration | 1280 us |
| Spoiler max. amplitude | 30.0 mT/m |
| Refocus grad. factor | 1.00 |

### Sequence - Special

| | |
|---|---|
| OVS slab thickness | 100.0 mm |
| OVS slab pos. offset | 5.0 mm |
| Spoiler duration | 1500 us |
| Acq. window shift | 200 us |
| Min. settling delay | 100 us |
| Gradient ramp time | 200 us |
| VAPOR flip angle | 72 deg |
| VAPOR delay 8 | 16 ms |
| VAPOR delay 7 | 66 ms |
| OVS pulse duration | 2560 us |
| OVS flip angle RO | 90 deg |
| OVS flip angle PH | 90 deg |
| OVS flip angle SL | 90 deg |
| VAPOR delay 6 | 71 ms |
| VAPOR delay 5 | 112 ms |
| VAPOR delay 4 | 115 ms |
| VAPOR delay 3 | 132 ms |
| VAPOR delay 2 | 110 ms |
| VAPOR delay 1 | 160 ms |
| Enable OVS | On |
| Resolve averages | On |
| Send ref. scans | Off |
| Inversion pulse | Off |
| Symmetric RF pulses | On |
| Invert SS grad. pol. | Off |
| Shift RO frequency | Off |
| Debug loop type | None |

| \\USER\PHCP\MRS\final\OCC steam eja w1 |
|---|
| TA: 0:23 PM: FIX Vol: 18 ×30 ×18 mmRel. SNR: 1.00 : steam |

### Properties

| | |
|---|---|
| Prio recon | Off |
| Load images to viewer | On |
| Inline movie | Off |
| Auto store images | On |
| Load images to stamp segments | Off |
| Load images to graphic segments | Off |
| Auto open inline display | Off |
| Auto close inline display | Off |
| Start measurement without further preparation | Off |
| Wait for user to start | Off |
| Start measurements | Single measurement |

### Routine

| | |
|---|---|
| Position | L1.2 P49.6 H8.9 mm |
| Orientation | Transversal |
| Rotation | 0 deg |
| Vol R >> L | 30 mm |
| Vol R >> L | 30 mm |
| Vol F >> H | 18 mm |
| TR | 5000 ms |
| TE | 8.00 ms |
| Averages | 1 |
| Filter | None |
| Coil elements | OCC |

### Contrast

| | |
|---|---|
| TR | 5000 ms |
| TE | 8.00 ms |
| TM | 32.00 ms |
| Averages | 1 |
| Flip angle | 90 deg |
| VAPOR | Only RF off |
| VAPOR suppr. | Water suppr. |
| Water s. BW | 135 Hz |
| Water s. delta pos. | 0.00 ppm |
| Measurements | 1 |

### Resolution - Common

| | |
|---|---|
| Vector size | 2048 |

### Geometry - Common

| | |
|---|---|
| Position | L1.2 P49.6 H8.9 mm |
| Orientation | Transversal |
| Rotation | 0 deg |
| Vol R >> L | 30 mm |
| Vol A >> P | 18 mm |
| Vol F >> H | 18 mm |

### Geometry - AutoAlign

| | |
|---|---|
| AutoAlign | Head > Brain |
| Initial Position | L1.2 P49.6 H8.9 |
| L | 1.2 mm |
| P | 49.6 mm |
| H | 8.9 mm |
| Initial Rotation | 0.00 deg |
| Initial Orientation | Transversal |

### System - Miscellaneous

| | |
|---|---|
| Positioning mode | FIX |

### System - Miscellaneous

| | |
|---|---|
| Table position | H |
| Table position | 0 mm |
| MSMA | S - C - T |
| Sagittal | R >> L |
| Coronal | A >> P |
| Transversal | F >> H |
| Save uncombined | Off |
| AutoAlign | Head > Brain |
| Coil Select Mode | Off - AutoCoilSelect |

### System - Adjustments

| | |
|---|---|
| B0 Shim mode | Brain |
| B1 Shim mode | TrueForm |
| Adj. water suppr. | Off |
| Confirm freq. adjustment | Off |
| Assume Dominant Fat | Off |
| Assume Silicone | Off |
| Adjustment Tolerance | Auto |

### System - Adjust Volume

| | |
|---|---|
| Position | L1.2 P49.6 H8.9 mm |
| Orientation | Transversal |
| Rotation | 0.00 deg |
| A >> P | 18 mm |
| R >> L | 30 mm |
| F >> H | 18 mm |
| Reset | Off |

### System - Tx/Rx

| | |
|---|---|
| Frequency 1H | 297.207726 MHz |
| Correction factor | 1 |
| Gain | High |
| Img. Scale Cor. | 1.000 |
| Reset | Off |
| ? Ref. amplitude 1H | 0.000 V |

### Physio - Signal1

| | |
|---|---|
| 1st Signal/Mode | None |
| TR | 5000 ms |

### Sequence - Common

| | |
|---|---|
| Introduction | On |
| Preparation scans | 2 |
| Delta frequency | 0.0 ppm |
| Phase cycling | Auto |
| Bandwidth | 6000 Hz |
| Acquisition duration | 341 ms |
| Remove oversampling | On |

### Sequence - Nuclei

| | |
|---|---|
| TX/RX Nucleus | 1H |
| TX/RX delta frequency | 0 Hz |
| TX Nucleus | None |
| TX delta frequency | 0 Hz |
| Coil elements | OCC |

### Sequence - Special

| | |
|---|---|
| RF pulse duration | 1280 us |
| Spoiler max. amplitude | 30.0 mT/m |
| Refocus grad. factor | 1.00 |

**Sequence - Special**

| | |
|---|---|
| OVS slab thickness | 100.0 mm |
| OVS slab pos. offset | 5.0 mm |
| Spoiler duration | 1500 us |
| Acq. window shift | 200 us |
| Min. settling delay | 100 us |
| Gradient ramp time | 200 us |
| VAPOR flip angle | 60 deg |
| VAPOR delay 8 | 16 ms |
| VAPOR delay 7 | 66 ms |
| OVS pulse duration | 2560 us |
| OVS flip angle RO | 90 deg |
| OVS flip angle PH | 90 deg |
| OVS flip angle SL | 90 deg |
| VAPOR delay 6 | 71 ms |
| VAPOR delay 5 | 112 ms |
| VAPOR delay 4 | 115 ms |
| VAPOR delay 3 | 132 ms |
| VAPOR delay 2 | 110 ms |
| VAPOR delay 1 | 160 ms |
| Enable OVS | On |
| Send ref. scans | Off |
| Inversion pulse | Off |
| Symmetric RF pulses | On |
| Invert SS grad. pol. | Off |
| Shift RO frequency | Off |
| Debug loop type | None |

| \\USER\PHCP\MRS\final\OCC steam eja w4 |
|---|
| TA: 0:38 PM: FIX Vol: 18 ×30 ×18 mmRel. SNR: 1.00 : steam |

### Properties

| | |
|---|---|
| Prio recon | Off |
| Load images to viewer | On |
| Inline movie | Off |
| Auto store images | On |
| Load images to stamp segments | Off |
| Load images to graphic segments | Off |
| Auto open inline display | Off |
| Auto close inline display | Off |
| Start measurement without further preparation | Off |
| Wait for user to start | Off |
| Start measurements | Single measurement |

### Routine

| | |
|---|---|
| Position | L1.2 P49.6 H8.9 mm |
| Orientation | Transversal |
| Rotation | 0 deg |
| Vol R >> L | 30 mm |
| Vol R >> L | 30 mm |
| Vol F >> H | 18 mm |
| TR | 5000 ms |
| TE | 8.00 ms |
| Averages | 4 |
| Filter | None |
| Coil elements | OCC |

### Contrast

| | |
|---|---|
| TR | 5000 ms |
| TE | 8.00 ms |
| TM | 32.00 ms |
| Averages | 4 |
| Flip angle | 90 deg |
| VAPOR | Only RF off |
| VAPOR suppr. | Water suppr. |
| Water s. BW | 135 Hz |
| Water s. delta pos. | 0.00 ppm |
| Measurements | 1 |

### Resolution - Common

| | |
|---|---|
| Vector size | 2048 |

### Geometry - Common

| | |
|---|---|
| Position | L1.2 P49.6 H8.9 mm |
| Orientation | Transversal |
| Rotation | 0 deg |
| Vol R >> L | 30 mm |
| Vol A >> P | 18 mm |
| Vol F >> H | 18 mm |

### Geometry - AutoAlign

| | |
|---|---|
| AutoAlign | Head > Brain |
| Initial Position | L1.2 P49.6 H8.9 |
| L | 1.2 mm |
| P | 49.6 mm |
| H | 8.9 mm |
| Initial Rotation | 0.00 deg |
| Initial Orientation | Transversal |

### System - Miscellaneous

| | |
|---|---|
| Positioning mode | FIX |

### System - Miscellaneous

| | |
|---|---|
| Table position | H |
| Table position | 0 mm |
| MSMA | S - C - T |
| Sagittal | R >> L |
| Coronal | A >> P |
| Transversal | F >> H |
| Save uncombined | Off |
| AutoAlign | Head > Brain |
| Coil Select Mode | Off - AutoCoilSelect |

### System - Adjustments

| | |
|---|---|
| B0 Shim mode | Brain |
| B1 Shim mode | TrueForm |
| Adj. water suppr. | Off |
| Confirm freq. adjustment | Off |
| Assume Dominant Fat | Off |
| Assume Silicone | Off |
| Adjustment Tolerance | Auto |

### System - Adjust Volume

| | |
|---|---|
| Position | L1.2 P49.6 H8.9 mm |
| Orientation | Transversal |
| Rotation | 0.00 deg |
| A >> P | 18 mm |
| R >> L | 30 mm |
| F >> H | 18 mm |
| Reset | Off |

### System - Tx/Rx

| | |
|---|---|
| Frequency 1H | 297.207726 MHz |
| Correction factor | 1 |
| Gain | High |
| Img. Scale Cor. | 1.000 |
| Reset | Off |
| ? Ref. amplitude 1H | 0.000 V |

### Physio - Signal1

| | |
|---|---|
| 1st Signal/Mode | None |
| TR | 5000 ms |

### Sequence - Common

| | |
|---|---|
| Introduction | On |
| Preparation scans | 2 |
| Delta frequency | 0.0 ppm |
| Phase cycling | Auto |
| Bandwidth | 6000 Hz |
| Acquisition duration | 341 ms |
| Remove oversampling | On |

### Sequence - Nuclei

| | |
|---|---|
| TX/RX Nucleus | 1H |
| TX/RX delta frequency | 0 Hz |
| TX Nucleus | None |
| TX delta frequency | 0 Hz |
| Coil elements | OCC |

### Sequence - Special

| | |
|---|---|
| RF pulse duration | 1280 us |
| Spoiler max. amplitude | 30.0 mT/m |
| Refocus grad. factor | 1.00 |

#### Sequence - Special

| | |
|---|---|
| OVS slab thickness | 100.0 mm |
| OVS slab pos. offset | 5.0 mm |
| Spoiler duration | 1500 us |
| Acq. window shift | 200 us |
| Min. settling delay | 100 us |
| Gradient ramp time | 200 us |
| VAPOR flip angle | 60 deg |
| VAPOR delay 8 | 16 ms |
| VAPOR delay 7 | 66 ms |
| OVS pulse duration | 2560 us |
| OVS flip angle RO | 90 deg |
| OVS flip angle PH | 90 deg |
| OVS flip angle SL | 90 deg |
| VAPOR delay 6 | 71 ms |
| VAPOR delay 5 | 112 ms |
| VAPOR delay 4 | 115 ms |
| VAPOR delay 3 | 132 ms |
| VAPOR delay 2 | 110 ms |
| VAPOR delay 1 | 160 ms |
| Enable OVS | On |
| Resolve averages | On |
| Send ref. scans | Off |
| Inversion pulse | Off |
| Symmetric RF pulses | On |
| Invert SS grad. pol. | Off |
| Shift RO frequency | Off |
| Debug loop type | None |

| \\USER\PHCP\MRS\final\OCC steam eja w1 noOVS |
|---|
| TA: 0:23 PM: FIX Vol: 18 ×30 ×18 mmRel. SNR: 1.00 : steam |

### Properties

| | |
|---|---|
| Prio recon | Off |
| Load images to viewer | On |
| Inline movie | Off |
| Auto store images | On |
| Load images to stamp segments | Off |
| Load images to graphic segments | Off |
| Auto open inline display | Off |
| Auto close inline display | Off |
| Start measurement without further preparation | Off |
| Wait for user to start | Off |
| Start measurements | Single measurement |

### Routine

| | |
|---|---|
| Position | L1.2 P49.6 H8.9 mm |
| Orientation | Transversal |
| Rotation | 0 deg |
| Vol R >> L | 30 mm |
| Vol R >> L | 30 mm |
| Vol F >> H | 18 mm |
| TR | 5000 ms |
| TE | 8.00 ms |
| Averages | 1 |
| Filter | None |
| Coil elements | OCC |

### Contrast

| | |
|---|---|
| TR | 5000 ms |
| TE | 8.00 ms |
| TM | 32.00 ms |
| Averages | 1 |
| Flip angle | 90 deg |
| VAPOR | Only RF off |
| VAPOR suppr. | Water suppr. |
| Water s. BW | 135 Hz |
| Water s. delta pos. | 0.00 ppm |
| Measurements | 1 |

### Resolution - Common

| | |
|---|---|
| Vector size | 2048 |

### Geometry - Common

| | |
|---|---|
| Position | L1.2 P49.6 H8.9 mm |
| Orientation | Transversal |
| Rotation | 0 deg |
| Vol R >> L | 30 mm |
| Vol A >> P | 18 mm |
| Vol F >> H | 18 mm |

### Geometry - AutoAlign

| | |
|---|---|
| AutoAlign | Head > Brain |
| Initial Position | L1.2 P49.6 H8.9 |
| L | 1.2 mm |
| P | 49.6 mm |
| H | 8.9 mm |
| Initial Rotation | 0.00 deg |
| Initial Orientation | Transversal |

### System - Miscellaneous

| | |
|---|---|
| Positioning mode | FIX |

### System - Miscellaneous

| | |
|---|---|
| Table position | H |
| Table position | 0 mm |
| MSMA | S - C - T |
| Sagittal | R >> L |
| Coronal | A >> P |
| Transversal | F >> H |
| Save uncombined | Off |
| AutoAlign | Head > Brain |
| Coil Select Mode | Off - AutoCoilSelect |

### System - Adjustments

| | |
|---|---|
| B0 Shim mode | Brain |
| B1 Shim mode | TrueForm |
| Adj. water suppr. | Off |
| Confirm freq. adjustment | Off |
| Assume Dominant Fat | Off |
| Assume Silicone | Off |
| Adjustment Tolerance | Auto |

### System - Adjust Volume

| | |
|---|---|
| Position | L1.2 P49.6 H8.9 mm |
| Orientation | Transversal |
| Rotation | 0.00 deg |
| A >> P | 18 mm |
| R >> L | 30 mm |
| F >> H | 18 mm |
| Reset | Off |

### System - Tx/Rx

| | |
|---|---|
| Frequency 1H | 297.207726 MHz |
| Correction factor | 1 |
| Gain | High |
| Img. Scale Cor. | 1.000 |
| Reset | Off |
| ? Ref. amplitude 1H | 0.000 V |

### Physio - Signal1

| | |
|---|---|
| 1st Signal/Mode | None |
| TR | 5000 ms |

### Sequence - Common

| | |
|---|---|
| Introduction | On |
| Preparation scans | 2 |
| Delta frequency | 0.0 ppm |
| Phase cycling | Auto |
| Bandwidth | 6000 Hz |
| Acquisition duration | 341 ms |
| Remove oversampling | On |

### Sequence - Nuclei

| | |
|---|---|
| TX/RX Nucleus | 1H |
| TX/RX delta frequency | 0 Hz |
| TX Nucleus | None |
| TX delta frequency | 0 Hz |
| Coil elements | OCC |

### Sequence - Special

| | |
|---|---|
| RF pulse duration | 1280 us |
| Spoiler max. amplitude | 30.0 mT/m |
| Refocus grad. factor | 1.00 |

#### Sequence - Special

| | |
|---|---|
| Spoiler duration | 1500 us |
| Acq. window shift | 200 us |
| Min. settling delay | 100 us |
| Gradient ramp time | 200 us |
| VAPOR flip angle | 60 deg |
| VAPOR delay 8 | 16 ms |
| VAPOR delay 7 | 66 ms |
| VAPOR delay 6 | 71 ms |
| VAPOR delay 5 | 112 ms |
| VAPOR delay 4 | 115 ms |
| VAPOR delay 3 | 132 ms |
| VAPOR delay 2 | 110 ms |
| VAPOR delay 1 | 160 ms |
| Enable OVS | Off |
| Send ref. scans | Off |
| Inversion pulse | Off |
| Symmetric RF pulses | On |
| Invert SS grad. pol. | Off |
| Shift RO frequency | Off |
| Debug loop type | None |

| \\USER\PHCP\MRS\final\OCC steam eja ws FA optimization |
|---|
| TA: 1:08 PM: FIX Vol: 18 ×30 ×18 mmRel. SNR: 1.00 : steam |

### Properties

| | |
|---|---|
| Prio recon | Off |
| Load images to viewer | On |
| Inline movie | Off |
| Auto store images | On |
| Load images to stamp segments | Off |
| Load images to graphic segments | Off |
| Auto open inline display | Off |
| Auto close inline display | Off |
| Start measurement without further preparation | Off |
| Wait for user to start | Off |
| Start measurements | Single measurement |

### Routine

| | |
|---|---|
| Position | L1.2 P49.6 H8.9 mm |
| Orientation | Transversal |
| Rotation | 0 deg |
| Vol R >> L | 30 mm |
| Vol R >> L | 30 mm |
| Vol F >> H | 18 mm |
| TR | 5000 ms |
| TE | 8.00 ms |
| Averages | 1 |
| Filter | None |
| Coil elements | OCC |

### Contrast

| | |
|---|---|
| TR | 5000 ms |
| TE | 8.00 ms |
| TM | 32.00 ms |
| Averages | 1 |
| Flip angle | 90 deg |
| VAPOR | Enabled |
| VAPOR suppr. | Water suppr. |
| Water s. BW | 135 Hz |
| Water s. delta pos. | 0.00 ppm |
| Measurements | 10 |

### Resolution - Common

| | |
|---|---|
| Vector size | 2048 |

### Geometry - Common

| | |
|---|---|
| Position | L1.2 P49.6 H8.9 mm |
| Orientation | Transversal |
| Rotation | 0 deg |
| Vol R >> L | 30 mm |
| Vol A >> P | 18 mm |
| Vol F >> H | 18 mm |

### Geometry - AutoAlign

| | |
|---|---|
| AutoAlign | Head > Brain |
| Initial Position | L1.2 P49.6 H8.9 |
| L | 1.2 mm |
| P | 49.6 mm |
| H | 8.9 mm |
| Initial Rotation | 0.00 deg |
| Initial Orientation | Transversal |

### System - Miscellaneous

| | |
|---|---|
| Positioning mode | FIX |

### System - Miscellaneous

| | |
|---|---|
| Table position | H |
| Table position | 0 mm |
| MSMA | S - C - T |
| Sagittal | R >> L |
| Coronal | A >> P |
| Transversal | F >> H |
| Save uncombined | Off |
| AutoAlign | Head > Brain |
| Coil Select Mode | Off - AutoCoilSelect |

### System - Adjustments

| | |
|---|---|
| B0 Shim mode | Brain |
| B1 Shim mode | TrueForm |
| Adj. water suppr. | Off |
| Confirm freq. adjustment | Off |
| Assume Dominant Fat | Off |
| Assume Silicone | Off |
| Adjustment Tolerance | Auto |

### System - Adjust Volume

| | |
|---|---|
| Position | L1.2 P49.6 H8.9 mm |
| Orientation | Transversal |
| Rotation | 0.00 deg |
| A >> P | 18 mm |
| R >> L | 30 mm |
| F >> H | 18 mm |
| Reset | Off |

### System - Tx/Rx

| | |
|---|---|
| Frequency 1H | 297.207726 MHz |
| Correction factor | 1 |
| Gain | High |
| Img. Scale Cor. | 1.000 |
| Reset | Off |
| ? Ref. amplitude 1H | 0.000 V |

### Physio - Signal1

| | |
|---|---|
| 1st Signal/Mode | None |
| TR | 5000 ms |

### Sequence - Common

| | |
|---|---|
| Introduction | On |
| Preparation scans | 2 |
| Delta frequency | -1.7 ppm |
| Phase cycling | Auto |
| Bandwidth | 6000 Hz |
| Acquisition duration | 341 ms |
| Remove oversampling | On |

### Sequence - Nuclei

| | |
|---|---|
| TX/RX Nucleus | 1H |
| TX/RX delta frequency | 0 Hz |
| TX Nucleus | None |
| TX delta frequency | 0 Hz |
| Coil elements | OCC |

### Sequence - Special

| | |
|---|---|
| RF pulse duration | 1280 us |
| Spoiler max. amplitude | 30.0 mT/m |
| Refocus grad. factor | 1.00 |

### Sequence - Special

| | |
|---|---|
| OVS slab thickness | 100.0 mm |
| OVS slab pos. offset | 5.0 mm |
| Spoiler duration | 1500 us |
| Acq. window shift | 200 us |
| Min. settling delay | 100 us |
| Gradient ramp time | 200 us |
| VAPOR flip angle | 60 deg |
| VAPOR delay 8 | 16 ms |
| VAPOR delay 7 | 66 ms |
| OVS pulse duration | 2560 us |
| OVS flip angle RO | 90 deg |
| OVS flip angle PH | 90 deg |
| OVS flip angle SL | 90 deg |
| VAPOR delay 6 | 71 ms |
| VAPOR delay 5 | 112 ms |
| VAPOR delay 4 | 115 ms |
| VAPOR delay 3 | 132 ms |
| VAPOR delay 2 | 110 ms |
| VAPOR delay 1 | 160 ms |
| Enable OVS | On |
| Weighted averages | Off |
| Send ref. scans | Off |
| Inversion pulse | Off |
| Symmetric RF pulses | On |
| Invert SS grad. pol. | Off |
| Shift RO frequency | Off |
| Debug loop type | WS FA |
| WS FA inc. | 2 deg |
| Measurements | 10 |

| \\USER\PHCP\MRS\final\OCC steam eja ws delay 7 |
|---|
| TA: 1:08 PM: FIX Vol: 18 ×30 ×18 mmRel. SNR: 1.00 : steam |

### Properties

| | |
|---|---|
| Prio recon | Off |
| Load images to viewer | On |
| Inline movie | Off |
| Auto store images | On |
| Load images to stamp segments | Off |
| Load images to graphic segments | Off |
| Auto open inline display | Off |
| Auto close inline display | Off |
| Start measurement without further preparation | Off |
| Wait for user to start | Off |
| Start measurements | Single measurement |

### Routine

| | |
|---|---|
| Position | L1.2 P49.6 H8.9 mm |
| Orientation | Transversal |
| Rotation | 0 deg |
| Vol R >> L | 30 mm |
| Vol R >> L | 30 mm |
| Vol F >> H | 18 mm |
| TR | 5000 ms |
| TE | 8.00 ms |
| Averages | 1 |
| Filter | None |
| Coil elements | OCC |

### Contrast

| | |
|---|---|
| TR | 5000 ms |
| TE | 8.00 ms |
| TM | 32.00 ms |
| Averages | 1 |
| Flip angle | 90 deg |
| VAPOR | Enabled |
| VAPOR suppr. | Water suppr. |
| Water s. BW | 135 Hz |
| Water s. delta pos. | 0.00 ppm |
| Measurements | 10 |

### Resolution - Common

| | |
|---|---|
| Vector size | 2048 |

### Geometry - Common

| | |
|---|---|
| Position | L1.2 P49.6 H8.9 mm |
| Orientation | Transversal |
| Rotation | 0 deg |
| Vol R >> L | 30 mm |
| Vol A >> P | 18 mm |
| Vol F >> H | 18 mm |

### Geometry - AutoAlign

| | |
|---|---|
| AutoAlign | Head > Brain |
| Initial Position | L1.2 P49.6 H8.9 |
| L | 1.2 mm |
| P | 49.6 mm |
| H | 8.9 mm |
| Initial Rotation | 0.00 deg |
| Initial Orientation | Transversal |

### System - Miscellaneous

| | |
|---|---|
| Positioning mode | FIX |

### System - Miscellaneous

| | |
|---|---|
| Table position | H |
| Table position | 0 mm |
| MSMA | S - C - T |
| Sagittal | R >> L |
| Coronal | A >> P |
| Transversal | F >> H |
| Save uncombined | Off |
| AutoAlign | Head > Brain |
| Coil Select Mode | Off - AutoCoilSelect |

### System - Adjustments

| | |
|---|---|
| B0 Shim mode | Brain |
| B1 Shim mode | TrueForm |
| Adj. water suppr. | Off |
| Confirm freq. adjustment | Off |
| Assume Dominant Fat | Off |
| Assume Silicone | Off |
| Adjustment Tolerance | Auto |

### System - Adjust Volume

| | |
|---|---|
| Position | L1.2 P49.6 H8.9 mm |
| Orientation | Transversal |
| Rotation | 0.00 deg |
| A >> P | 18 mm |
| R >> L | 30 mm |
| F >> H | 18 mm |
| Reset | Off |

### System - Tx/Rx

| | |
|---|---|
| Frequency 1H | 297.207726 MHz |
| Correction factor | 1 |
| Gain | High |
| Img. Scale Cor. | 1.000 |
| Reset | Off |
| ? Ref. amplitude 1H | 0.000 V |

### Physio - Signal1

| | |
|---|---|
| 1st Signal/Mode | None |
| TR | 5000 ms |

### Sequence - Common

| | |
|---|---|
| Introduction | On |
| Preparation scans | 2 |
| Delta frequency | -1.7 ppm |
| Phase cycling | Auto |
| Bandwidth | 6000 Hz |
| Acquisition duration | 341 ms |
| Remove oversampling | On |

### Sequence - Nuclei

| | |
|---|---|
| TX/RX Nucleus | 1H |
| TX/RX delta frequency | 0 Hz |
| TX Nucleus | None |
| TX delta frequency | 0 Hz |
| Coil elements | OCC |

### Sequence - Special

| | |
|---|---|
| RF pulse duration | 1280 us |
| Spoiler max. amplitude | 30.0 mT/m |
| Refocus grad. factor | 1.00 |

### Sequence - Special

| | |
|---|---|
| OVS slab thickness | 100.0 mm |
| OVS slab pos. offset | 5.0 mm |
| Spoiler duration | 1500 us |
| Acq. window shift | 200 us |
| Min. settling delay | 100 us |
| Gradient ramp time | 200 us |
| VAPOR flip angle | 60 deg |
| VAPOR delay 8 | 16 ms |
| VAPOR delay 7 | 66 ms |
| OVS pulse duration | 2560 us |
| OVS flip angle RO | 90 deg |
| OVS flip angle PH | 90 deg |
| OVS flip angle SL | 90 deg |
| VAPOR delay 6 | 71 ms |
| VAPOR delay 5 | 112 ms |
| VAPOR delay 4 | 115 ms |
| VAPOR delay 3 | 132 ms |
| VAPOR delay 2 | 110 ms |
| VAPOR delay 1 | 160 ms |
| Enable OVS | On |
| Weighted averages | Off |
| Send ref. scans | Off |
| Inversion pulse | Off |
| Symmetric RF pulses | On |
| Invert SS grad. pol. | Off |
| Shift RO frequency | Off |
| Debug loop type | WS delay 7 |
| WS delay 7 inc. | 3 ms |
| Measurements | 10 |

**\\USER\PHCP\MRS\final\PFC localizer**

TA: 0:13 PM: REF Voxel size: 0.5×0.5×7.0 mmPAT: Off Rel. SNR: 1.00 : qfl

### Properties

| | |
|---|---|
| Prio recon | Off |
| Load images to viewer | On |
| Inline movie | Off |
| Auto store images | On |
| Load images to stamp segments | On |
| Load images to graphic segments | On |
| Auto open inline display | On |
| Auto close inline display | Off |
| Start measurement without further preparation | Off |
| Wait for user to start | Off |
| Start measurements | Single measurement |

### Routine

| | |
|---|---|
| Slice group | 1 |
| Slices | 1 |
| Dist. factor | 20 % |
| Position | Isocenter |
| Orientation | Sagittal |
| Phase enc. dir. | A >> P |
| Slice group | 2 |
| Slices | 1 |
| Dist. factor | 20 % |
| Position | Isocenter |
| Orientation | Transversal |
| Phase enc. dir. | A >> P |
| Slice group | 3 |
| Slices | 1 |
| Dist. factor | 20 % |
| Position | Isocenter |
| Orientation | Coronal |
| Phase enc. dir. | R >> L |
| AutoAlign | --- |
| Phase oversampling | 0 % |
| FoV read | 250 mm |
| FoV phase | 100.0 % |
| Slice thickness | 7.0 mm |
| TR | 8.6 ms |
| TE | 4.00 ms |
| Averages | 2 |
| Concatenations | 3 |
| Filter | Elliptical filter |
| Coil elements | OCC;PFC |

### Contrast - Common

| | |
|---|---|
| TR | 8.6 ms |
| TE | 4.00 ms |
| TD | 0 ms |
| MTC | Off |
| Magn. preparation | None |
| Flip angle | 20 deg |
| Fat suppr. | None |
| Water suppr. | None |
| SWI | Off |

### Contrast - Dynamic

| | |
|---|---|
| Averages | 2 |
| Averaging mode | Short term |
| Reconstruction | Magnitude |
| Measurements | 1 |

| | |
|---|---|
| Tim CT mode | Off |
| Slices | 1 |
| Slice thickness | 7.0 mm |
| Dist. factor | 20 % |
| FoV read | 250 mm |
| FoV phase | 100.0 % |
| Segments | 1 |

### System - Miscellaneous

| | |
|---|---|
| Positioning mode | REF |
| Table position | F |
| Table position | 0 mm |
| MSMA | S - C - T |
| Sagittal | R >> L |
| Coronal | A >> P |
| Transversal | F >> H |
| Coil Combine Mode | Sum of Squares |
| Save uncombined | Off |
| Matrix Optimization | Off |
| AutoAlign | --- |
| Coil Select Mode | Off - AutoCoilSelect |

### System - Adjustments

| | |
|---|---|
| B0 Shim mode | Tune up |
| B1 Shim mode | TrueForm |
| Confirm freq. adjustment | Off |
| Assume Dominant Fat | Off |
| Assume Silicone | Off |
| Adjustment Tolerance | Auto |

### System - Adjust Volume

| | |
|---|---|
| Position | Isocenter |
| Orientation | Transversal |
| Rotation | 0.00 deg |
| A >> P | 263 mm |
| R >> L | 350 mm |
| F >> H | 350 mm |
| Reset | Off |

### System - Tx/Rx

| | |
|---|---|
| Frequency 1H | 297.207726 MHz |
| Correction factor | 1 |
| Gain | High |
| Img. Scale Cor. | 1.000 |
| Reset | Off |
| ? Ref. amplitude 1H | 0.000 V |

### Physio - Signal1

| | |
|---|---|
| 1st Signal/Mode | None |
| TR | 8.6 ms |
| Concatenations | 3 |
| Segments | 1 |

### Physio - Cardiac

| | |
|---|---|
| Tagging | None |
| Magn. preparation | None |
| Fat suppr. | None |
| Dark blood | Off |
| FoV read | 250 mm |
| FoV phase | 100.0 % |
| Phase resolution | 90 % |

### Physio - PACE

| | |
|---|---|
| Resp. control | Off |
| Concatenations | 3 |

### Inline - Common

| | |
|---|---|
| Subtract | Off |
| Measurements | 1 |
| StdDev | Off |
| Liver registration | Off |
| Save original images | On |

### Inline - MIP

| | |
|---|---|
| MIP-Sag | Off |
| MIP-Cor | Off |
| MIP-Tra | Off |
| MIP-Time | Off |
| Save original images | On |

### Inline - Soft Tissue

| | |
|---|---|
| Wash - In | Off |
| Wash - Out | Off |
| TTP | Off |
| PEI | Off |
| MIP - time | Off |
| Measurements | 1 |

### Inline - Composing

| | |
|---|---|
| Inline Composing | Off |
| Distortion Corr. | Off |

### Inline - MapIt

| | |
|---|---|
| Save original images | On |
| MapIt | None |
| Flip angle | 20 deg |
| Measurements | 1 |
| Contrasts | 1 |
| TR | 8.6 ms |
| TE | 4.00 ms |

### Sequence - Part 1

| | |
|---|---|
| Introduction | On |

#### Sequence - Part 1

| | |
|---|---|
| Dimension | 2D |
| Phase stabilisation | Off |
| Asymmetric echo | Allowed |
| Contrasts | 1 |
| Flow comp. | No |
| Multi-slice mode | Sequential |
| Bandwidth | 320 Hz/Px |

### Geometry - AutoAlign

| | |
|---|---|
| Slab group | 1 |
| Position | L0.0 A22.2 F11.5 mm |
| Orientation | Sagittal |
| Phase enc. dir. | A >> P |
| Initial Position | Isocenter |
| L | 0.0 mm |
| P | 0.0 mm |
| H | 0.0 mm |
| Initial Rotation | 0.00 deg |
| Initial Orientation | Transversal |

### Sequence - Assistant

| | |
|---|---|
| Mode | Off |

| \\USER\PHCP\MRS\final\localizer aligned |
|---|
| TA: 0:28 PM: FIX Voxel size: 0.5×0.5×5.0 mmPAT: Off Rel. SNR: 1.00 : qfl |

### Properties

| | |
|---|---|
| Prio recon | Off |
| Load images to viewer | On |
| Inline movie | Off |
| Auto store images | On |
| Load images to stamp segments | On |
| Load images to graphic segments | On |
| Auto open inline display | Off |
| Auto close inline display | Off |
| Start measurement without further preparation | On |
| Wait for user to start | Off |
| Start measurements | Single measurement |

**Sequence - Assistant**

| | |
|---|---|
| Mode | Off |

| \\USER\PHCP\MRS\final\PFC t1inplane 64sl occ 1p25iso 2p5sTR 160FOV |
|---|
| TA: 4:02 PM: FIX Voxel size: 1.3×1.3×1.3 mmPAT: Off Rel. SNR: 1.00 : tfl |

### Properties

| | |
|---|---|
| Prio recon | Off |
| Load images to viewer | On |
| Inline movie | Off |
| Auto store images | On |
| Load images to stamp segments | On |
| Load images to graphic segments | On |
| Auto open inline display | Off |
| Auto close inline display | Off |
| Start measurement without further preparation | Off |
| Wait for user to start | Off |
| Start measurements | Single measurement |

### Routine

| | |
|---|---|
| Slab group | 1 |
| Slabs | 1 |
| Dist. factor | 50 % |
| Position | L1.8 A1.2 H7.3 mm |
| Orientation | Sagittal |
| Phase enc. dir. | A >> P |
| AutoAlign | Head > Brain |
| Phase oversampling | 0 % |
| Slice oversampling | 0.0 % |
| Slices per slab | 64 |
| FoV read | 160 mm |
| FoV phase | 100.0 % |
| Slice thickness | 1.25 mm |
| TR | 2500.0 ms |
| TE | 3.2 ms |
| Averages | 1 |
| Concatenations | 1 |
| Filter | None |
| Coil elements | PFC |

### Contrast - Common

| | |
|---|---|
| TR | 2500.0 ms |
| TE | 3.2 ms |
| Magn. preparation | Non-sel. IR |
| TI | 1500 ms |
| Flip angle | 5.0 deg |
| Fat suppr. | None |
| Water suppr. | None |

### Contrast - Dynamic

| | |
|---|---|
| Averages | 1 |
| Averaging mode | Long term |
| Reconstruction | Magnitude |
| Measurements | 1 |
| Multiple series | Each measurement |

### Resolution - Common

| | |
|---|---|
| FoV read | 160 mm |
| FoV phase | 100.0 % |
| Slice thickness | 1.25 mm |
| Base resolution | 128 |
| Phase resolution | 100 % |
| Slice resolution | 100 % |
| Phase partial Fourier | 6/8 |
| Slice partial Fourier | Off |
| Interpolation | Off |

### Resolution - iPAT

| | |
|---|---|
| PAT mode | None |

### Resolution - Filter Image

| | |
|---|---|
| Image Filter | Off |
| Distortion Corr. | Off |
| Prescan Normalize | Off |
| Normalize | Off |
| B1 filter | Off |

### Resolution - Filter Rawdata

| | |
|---|---|
| Raw filter | Off |
| Elliptical filter | Off |

### Geometry - Common

| | |
|---|---|
| Slab group | 1 |
| Slabs | 1 |
| Dist. factor | 50 % |
| Position | L1.8 A1.2 H7.3 mm |
| Orientation | Sagittal |
| Phase enc. dir. | A >> P |
| Slice oversampling | 0.0 % |
| Slices per slab | 64 |
| FoV read | 160 mm |
| FoV phase | 100.0 % |
| Slice thickness | 1.25 mm |
| TR | 2500.0 ms |
| Multi-slice mode | Single shot |
| Series | Ascending |
| Concatenations | 1 |

### Geometry - AutoAlign

| | |
|---|---|
| Slab group | 1 |
| Position | L1.8 A1.2 H7.3 mm |
| Orientation | Sagittal |
| Phase enc. dir. | A >> P |
| AutoAlign | Head > Brain |
| Initial Position | L1.8 A1.2 H7.3 |
| L | 1.8 mm |
| A | 1.2 mm |
| H | 7.3 mm |
| Initial Rotation | 0.00 deg |
| Initial Orientation | Sagittal |

### Geometry - Navigator

### Geometry - Tim Planning Suite

| | |
|---|---|
| Set-n-Go Protocol | Off |
| Table position | H |
| Table position | 0 mm |
| Inline Composing | Off |

### System - Miscellaneous

| | |
|---|---|
| Positioning mode | FIX |
| Table position | H |
| Table position | 0 mm |
| MSMA | S - C - T |
| Sagittal | R >> L |
| Coronal | A >> P |
| Transversal | F >> H |
| Coil Combine Mode | Adaptive Combine |

### System - Miscellaneous

| | |
|---|---|
| Save uncombined | Off |
| Matrix Optimization | Off |
| AutoAlign | Head > Brain |
| Coil Select Mode | Off - All |

### System - Adjustments

| | |
|---|---|
| B0 Shim mode | Tune up |
| B1 Shim mode | TrueForm |
| Confirm freq. adjustment | Off |
| Assume Dominant Fat | Off |
| Assume Silicone | Off |
| Adjustment Tolerance | Auto |

### System - Adjust Volume

| | |
|---|---|
| ! Position | L1.8 P18.8 H15.1 mm |
| ! Orientation | T > C-18.8 |
| ! Rotation | 0.00 deg |
| ! A >> P | 64 mm |
| ! R >> L | 148 mm |
| ! F >> H | 82 mm |
| Reset | Off |

### System - Tx/Rx

| | |
|---|---|
| Frequency 1H | 297.207726 MHz |
| Correction factor | 1 |
| Gain | High |
| Img. Scale Cor. | 1.000 |
| Reset | Off |
| ? Ref. amplitude 1H | 0.000 V |

### Physio - Signal1

| | |
|---|---|
| 1st Signal/Mode | None |
| TR | 2500.0 ms |
| Concatenations | 1 |

### Physio - Cardiac

| | |
|---|---|
| Magn. preparation | Non-sel. IR |
| TI | 1500 ms |
| Fat suppr. | None |
| Dark blood | Off |
| FoV read | 160 mm |
| FoV phase | 100.0 % |
| Phase resolution | 100 % |

### Physio - PACE

| | |
|---|---|
| Resp. control | Off |
| Concatenations | 1 |

### Inline - Common

| | |
|---|---|
| Subtract | Off |
| Measurements | 1 |
| StdDev | Off |
| Save original images | On |

### Inline - MIP

| | |
|---|---|
| MIP-Sag | Off |
| MIP-Cor | Off |
| MIP-Tra | Off |
| MIP-Time | Off |
| Save original images | On |

### Inline - Composing

| | |
|---|---|
| Inline Composing | Off |

### Inline - Composing

| | |
|---|---|
| Distortion Corr. | Off |

### Inline - MapIt

| | |
|---|---|
| Save original images | On |
| MapIt | None |
| Flip angle | 5.0 deg |
| Measurements | 1 |
| TR | 2500.0 ms |
| TE | 3.2 ms |

### Sequence - Part 1

| | |
|---|---|
| Introduction | On |
| Dimension | 3D |
| Elliptical scanning | Off |
| Reordering | Linear |
| Asymmetric echo | Allowed |
| Flow comp. | No |
| Multi-slice mode | Single shot |
| Echo spacing | 7.9 ms |
| Bandwidth | 180 Hz/Px |

### Sequence - Part 2

| | |
|---|---|
| RF pulse type | Normal |
| Gradient mode | Normal |
| Excitation | Slab-sel. |
| RF spoiling | On |
| Incr. Gradient spoiling | Off |
| Turbo factor | 64 |

### Sequence - Nuclei

| | |
|---|---|
| TX/RX Nucleus | 1H |
| TX/RX delta frequency | 0 Hz |
| TX Nucleus | None |
| TX delta frequency | 0 Hz |
| Coil elements | PFC |

### Sequence - Assistant

| | |
|---|---|
| Mode | Off |

| \\USER\PHCP\MRS\final\PFC fastestmap lin |
|---|
| TA: 6.1 s PM: FIX Vol: 30 ×30 ×15 mmRel. SNR: 1.00 : fastmp |

### Properties

| | |
|---|---|
| Prio recon | Off |
| Load images to viewer | On |
| Inline movie | Off |
| Auto store images | On |
| Load images to stamp segments | Off |
| Load images to graphic segments | Off |
| Auto open inline display | On |
| Auto close inline display | Off |
| Start measurement without further preparation | Off |
| Wait for user to start | Off |
| Start measurements | Single measurement |

### Routine

| | |
|---|---|
| Position | L1.2 P49.6 H8.9 mm |
| Orientation | C > T30.0 |
| Rotation | 0 deg |
| VoI F >> H | 30 mm |
| VoI F >> H | 30 mm |
| VoI A >> P | 15 mm |
| TR | 2000 ms |
| TE | 46.00 ms |
| Averages | 1 |
| Filter | None |
| Coil elements | PFC |

### Contrast

| | |
|---|---|
| TR | 2000 ms |
| TE | 46.00 ms |
| Tau | 5.00 ms |
| Averages | 1 |
| Excite flip angle | 90 deg |
| Refocus flip angle | 180 deg |
| Measurements | 1 |

### Resolution - Common

| | |
|---|---|
| Vector size | 256 |

### Geometry - Common

| | |
|---|---|
| Position | L1.2 P49.6 H8.9 mm |
| Orientation | C > T30.0 |
| Rotation | 0 deg |
| VoI F >> H | 30 mm |
| VoI R >> L | 30 mm |
| VoI A >> P | 15 mm |

### Geometry - AutoAlign

| | |
|---|---|
| AutoAlign | Head > Brain |
| Initial Position | L1.2 P49.6 H8.9 |
| L | 1.2 mm |
| P | 49.6 mm |
| H | 8.9 mm |
| Initial Rotation | 0.00 deg |
| Initial Orientation | C > T |
| C > T | 30.0 |
| > S | 0.0 |

### System - Miscellaneous

| | |
|---|---|
| Positioning mode | FIX |
| Table position | H |
| Table position | 0 mm |
| MSMA | S - C - T |
| Sagittal | R >> L |
| Coronal | A >> P |
| Transversal | F >> H |
| Save uncombined | Off |
| AutoAlign | Head > Brain |
| Coil Select Mode | Off - AutoCoilSelect |

### System - Adjustments

| | |
|---|---|
| B0 Shim mode | Tune up |
| B1 Shim mode | TrueForm |
| Adj. water suppr. | Off |
| Confirm freq. adjustment | Off |
| Assume Dominant Fat | Off |
| Assume Silicone | Off |
| Adjustment Tolerance | Auto |

### System - Adjust Volume

| | |
|---|---|
| Position | L1.2 P49.6 H8.9 mm |
| Orientation | C > T30.0 |
| Rotation | 0.00 deg |
| R >> L | 30 mm |
| F >> H | 30 mm |
| A >> P | 15 mm |
| Reset | Off |

### System - Tx/Rx

| | |
|---|---|
| Frequency 1H | 297.207726 MHz |
| Correction factor | 1 |
| Gain | High |
| Img. Scale Cor. | 1.000 |
| Reset | Off |
| ? Ref. amplitude 1H | 0.000 V |

### Physio - Signal1

| | |
|---|---|
| 1st Signal/Mode | None |
| TR | 2000 ms |

### Sequence - Common

| | |
|---|---|
| Delta frequency | 0.0 ppm |
| Phase cycling | None |
| Bandwidth | 100000 Hz |
| Acquisition duration | 2 ms |

### Sequence - Special

| | |
|---|---|
| Type of fit | Linear 3-proj |
| VoI fit factor | 100 % |
| Force spherical fit VoI | Off Force spherical fit VoI |
| Save plots to database | Off |
| Refocus pulses | Normal |
| Excite pulse duration | 6400 us |
| Refocus pulse duration | 5120 us |
| Bar FoV | 384 mm |
| Bar thickness | 10.0 mm |
| Inversion pulse | Off |
| Multi-echo acquisition | On |
| Number of echoes | 4 |

| **\\USER\PHCP\MRS\final\PFC fastestmap all** |
|---|
| TA: 0:12 PM: FIX Vol: 30 ×30 ×15 mmRel. SNR: 1.00 : fastmp |

### Properties

| | |
|---|---|
| Prio recon | Off |
| Load images to viewer | On |
| Inline movie | Off |
| Auto store images | On |
| Load images to stamp segments | Off |
| Load images to graphic segments | Off |
| Auto open inline display | On |
| Auto close inline display | Off |
| Start measurement without further preparation | Off |
| Wait for user to start | Off |
| Start measurements | Single measurement |

### Routine

| | |
|---|---|
| Position | L1.2 P49.6 H8.9 mm |
| Orientation | C > T30.0 |
| Rotation | 0 deg |
| VoI F >> H | 30 mm |
| VoI F >> H | 30 mm |
| VoI A >> P | 15 mm |
| TR | 2000 ms |
| TE | 46.00 ms |
| Averages | 1 |
| Filter | None |
| Coil elements | PFC |

### Contrast

| | |
|---|---|
| TR | 2000 ms |
| TE | 46.00 ms |
| Tau | 5.00 ms |
| Averages | 1 |
| Excite flip angle | 90 deg |
| Refocus flip angle | 180 deg |
| Measurements | 1 |

### Resolution - Common

| | |
|---|---|
| Vector size | 256 |

### Geometry - Common

| | |
|---|---|
| Position | L1.2 P49.6 H8.9 mm |
| Orientation | C > T30.0 |
| Rotation | 0 deg |
| VoI F >> H | 30 mm |
| VoI R >> L | 30 mm |
| VoI A >> P | 15 mm |

### Geometry - AutoAlign

| | |
|---|---|
| AutoAlign | Head > Brain |
| Initial Position | L1.2 P49.6 H8.9 |
| L | 1.2 mm |
| P | 49.6 mm |
| H | 8.9 mm |
| Initial Rotation | 0.00 deg |
| Initial Orientation | C > T |
| C > T | 30.0 |
| > S | 0.0 |

### System - Miscellaneous

| | |
|---|---|
| Positioning mode | FIX |
| Table position | H |
| Table position | 0 mm |
| MSMA | S - C - T |
| Sagittal | R >> L |
| Coronal | A >> P |
| Transversal | F >> H |
| Save uncombined | Off |
| AutoAlign | Head > Brain |
| Coil Select Mode | Off - AutoCoilSelect |

### System - Adjustments

| | |
|---|---|
| B0 Shim mode | Tune up |
| B1 Shim mode | TrueForm |
| Adj. water suppr. | Off |
| Confirm freq. adjustment | Off |
| Assume Dominant Fat | Off |
| Assume Silicone | Off |
| Adjustment Tolerance | Auto |

### System - Adjust Volume

| | |
|---|---|
| Position | L1.2 P49.6 H8.9 mm |
| Orientation | C > T30.0 |
| Rotation | 0.00 deg |
| R >> L | 30 mm |
| F >> H | 30 mm |
| A >> P | 15 mm |
| Reset | Off |

### System - Tx/Rx

| | |
|---|---|
| Frequency 1H | 297.207726 MHz |
| Correction factor | 1 |
| Gain | High |
| Img. Scale Cor. | 1.000 |
| Reset | Off |
| ? Ref. amplitude 1H | 0.000 V |

### Physio - Signal1

| | |
|---|---|
| 1st Signal/Mode | None |
| TR | 2000 ms |

### Sequence - Common

| | |
|---|---|
| Delta frequency | 0.0 ppm |
| Phase cycling | None |
| Bandwidth | 100000 Hz |
| Acquisition duration | 2 ms |

### Sequence - Special

| | |
|---|---|
| Type of fit | Full 6-proj |
| VoI fit factor | 100 % |
| Force spherical fit VoI | Off Force spherical fit VoI |
| Save plots to database | Off |
| Refocus pulses | Normal |
| Excite pulse duration | 6400 us |
| Refocus pulse duration | 5120 us |
| Bar FoV | 384 mm |
| Bar thickness | 10.0 mm |
| Inversion pulse | Off |
| Multi-echo acquisition | On |
| Number of echoes | 4 |

| \\USER\PHCP\MRS\final\PFC fastestmap all |
|---|
| TA: 0:12 PM: FIX Vol: 30 ×30 ×15 mmRel. SNR: 1.00 : fastmp |

### Properties

| | |
|---|---|
| Prio recon | Off |
| Load images to viewer | On |
| Inline movie | Off |
| Auto store images | On |
| Load images to stamp segments | Off |
| Load images to graphic segments | Off |
| Auto open inline display | Off |
| Auto close inline display | Off |
| Start measurement without further preparation | Off |
| Wait for user to start | Off |
| Start measurements | Single measurement |

### Routine

| | |
|---|---|
| Position | L1.2 P49.6 H8.9 mm |
| Orientation | C > T30.0 |
| Rotation | 0 deg |
| VoI F >> H | 30 mm |
| VoI F >> H | 30 mm |
| VoI A >> P | 15 mm |
| TR | 2000 ms |
| TE | 46.00 ms |
| Averages | 1 |
| Filter | None |
| Coil elements | PFC |

### Contrast

| | |
|---|---|
| TR | 2000 ms |
| TE | 46.00 ms |
| Tau | 5.00 ms |
| Averages | 1 |
| Excite flip angle | 90 deg |
| Refocus flip angle | 180 deg |
| Measurements | 1 |

### Resolution - Common

| | |
|---|---|
| Vector size | 256 |

### Geometry - Common

| | |
|---|---|
| Position | L1.2 P49.6 H8.9 mm |
| Orientation | C > T30.0 |
| Rotation | 0 deg |
| VoI F >> H | 30 mm |
| VoI R >> L | 30 mm |
| VoI A >> P | 15 mm |

### Geometry - AutoAlign

| | |
|---|---|
| AutoAlign | Head > Brain |
| Initial Position | L1.2 P49.6 H8.9 |
| L | 1.2 mm |
| P | 49.6 mm |
| H | 8.9 mm |
| Initial Rotation | 0.00 deg |
| Initial Orientation | C > T |
| C > T | 30.0 |
| > S | 0.0 |

### System - Miscellaneous

| | |
|---|---|
| Positioning mode | FIX |
| Table position | H |
| Table position | 0 mm |
| MSMA | S - C - T |
| Sagittal | R >> L |
| Coronal | A >> P |
| Transversal | F >> H |
| Save uncombined | Off |
| AutoAlign | Head > Brain |
| Coil Select Mode | Off - AutoCoilSelect |

### System - Adjustments

| | |
|---|---|
| B0 Shim mode | Tune up |
| B1 Shim mode | TrueForm |
| Adj. water suppr. | Off |
| Confirm freq. adjustment | Off |
| Assume Dominant Fat | Off |
| Assume Silicone | Off |
| Adjustment Tolerance | Auto |

### System - Adjust Volume

| | |
|---|---|
| Position | L1.2 P49.6 H8.9 mm |
| Orientation | C > T30.0 |
| Rotation | 0.00 deg |
| R >> L | 30 mm |
| F >> H | 30 mm |
| A >> P | 15 mm |
| Reset | Off |

### System - Tx/Rx

| | |
|---|---|
| Frequency 1H | 297.207726 MHz |
| Correction factor | 1 |
| Gain | High |
| Img. Scale Cor. | 1.000 |
| Reset | Off |
| ? Ref. amplitude 1H | 0.000 V |

### Physio - Signal1

| | |
|---|---|
| 1st Signal/Mode | None |
| TR | 2000 ms |

### Sequence - Common

| | |
|---|---|
| Delta frequency | 0.0 ppm |
| Phase cycling | None |
| Bandwidth | 100000 Hz |
| Acquisition duration | 2 ms |

### Sequence - Special

| | |
|---|---|
| Type of fit | Full 6-proj |
| VoI fit factor | 100 % |
| Force spherical fit VoI | Off Force spherical fit VoI |
| Save plots to database | Off |
| Refocus pulses | Normal |
| Excite pulse duration | 6400 us |
| Refocus pulse duration | 5120 us |
| Bar FoV | 384 mm |
| Bar thickness | 10.0 mm |
| Inversion pulse | Off |
| Multi-echo acquisition | On |
| Number of echoes | 4 |

| \\USER\PHCP\MRS\final\PFC fastestmap all |
|---|
| TA: 0:12 PM: FIX Vol: 30 ×30 ×15 mmRel. SNR: 1.00 : fastmp |

### Properties

| | |
|---|---|
| Prio recon | Off |
| Load images to viewer | On |
| Inline movie | Off |
| Auto store images | On |
| Load images to stamp segments | Off |
| Load images to graphic segments | Off |
| Auto open inline display | Off |
| Auto close inline display | Off |
| Start measurement without further preparation | Off |
| Wait for user to start | Off |
| Start measurements | Single measurement |

### Routine

| | |
|---|---|
| Position | L1.2 P49.6 H8.9 mm |
| Orientation | C > T30.0 |
| Rotation | 0 deg |
| VoI F >> H | 30 mm |
| VoI F >> H | 30 mm |
| VoI A >> P | 15 mm |
| TR | 2000 ms |
| TE | 46.00 ms |
| Averages | 1 |
| Filter | None |
| Coil elements | PFC |

### Contrast

| | |
|---|---|
| TR | 2000 ms |
| TE | 46.00 ms |
| Tau | 5.00 ms |
| Averages | 1 |
| Excite flip angle | 90 deg |
| Refocus flip angle | 180 deg |
| Measurements | 1 |

### Resolution - Common

| | |
|---|---|
| Vector size | 256 |

### Geometry - Common

| | |
|---|---|
| Position | L1.2 P49.6 H8.9 mm |
| Orientation | C > T30.0 |
| Rotation | 0 deg |
| VoI F >> H | 30 mm |
| VoI R >> L | 30 mm |
| VoI A >> P | 15 mm |

### Geometry - AutoAlign

| | |
|---|---|
| AutoAlign | Head > Brain |
| Initial Position | L1.2 P49.6 H8.9 |
| L | 1.2 mm |
| P | 49.6 mm |
| H | 8.9 mm |
| Initial Rotation | 0.00 deg |
| Initial Orientation | C > T |
| C > T | 30.0 |
| > S | 0.0 |

### System - Miscellaneous

| | |
|---|---|
| Positioning mode | FIX |
| Table position | H |

### System - Miscellaneous

| | |
|---|---|
| Table position | 0 mm |
| MSMA | S - C - T |
| Sagittal | R >> L |
| Coronal | A >> P |
| Transversal | F >> H |
| Save uncombined | Off |
| AutoAlign | Head > Brain |
| Coil Select Mode | Off - AutoCoilSelect |

### System - Adjustments

| | |
|---|---|
| B0 Shim mode | Tune up |
| B1 Shim mode | TrueForm |
| Adj. water suppr. | Off |
| Confirm freq. adjustment | Off |
| Assume Dominant Fat | Off |
| Assume Silicone | Off |
| Adjustment Tolerance | Auto |

### System - Adjust Volume

| | |
|---|---|
| Position | L1.2 P49.6 H8.9 mm |
| Orientation | C > T30.0 |
| Rotation | 0.00 deg |
| R >> L | 30 mm |
| F >> H | 30 mm |
| A >> P | 15 mm |
| Reset | Off |

### System - Tx/Rx

| | |
|---|---|
| Frequency 1H | 297.207726 MHz |
| Correction factor | 1 |
| Gain | High |
| Img. Scale Cor. | 1.000 |
| Reset | Off |
| ? Ref. amplitude 1H | 0.000 V |

### Physio - Signal1

| | |
|---|---|
| 1st Signal/Mode | None |
| TR | 2000 ms |

### Sequence - Common

| | |
|---|---|
| Delta frequency | 0.0 ppm |
| Phase cycling | None |
| Bandwidth | 100000 Hz |
| Acquisition duration | 2 ms |

### Sequence - Special

| | |
|---|---|
| Type of fit | Full 6-proj |
| VoI fit factor | 100 % |
| Force spherical fit VoI | Off Force spherical fit VoI |
| Save plots to database | Off |
| Refocus pulses | Normal |
| Excite pulse duration | 6400 us |
| Refocus pulse duration | 5120 us |
| Bar FoV | 384 mm |
| Bar thickness | 10.0 mm |
| Inversion pulse | Off |
| Multi-echo acquisition | On |
| Number of echoes | 4 |

| \\USER\PHCP\MRS\final\PFC fastestmap lin6 |
|---|
| TA: 0:24 PM: FIX VoI: 30 ×30 ×15 mmRel. SNR: 1.00 : fastmp |

### Properties

| | |
|---|---|
| Prio recon | Off |
| Load images to viewer | On |
| Inline movie | Off |
| Auto store images | On |
| Load images to stamp segments | Off |
| Load images to graphic segments | Off |
| Auto open inline display | On |
| Auto close inline display | Off |
| Start measurement without further preparation | Off |
| Wait for user to start | Off |
| Start measurements | Single measurement |

### Routine

| | |
|---|---|
| Position | L1.2 P49.6 H8.9 mm |
| Orientation | C > T30.0 |
| Rotation | 0 deg |
| VoI F >> H | 30 mm |
| VoI F >> H | 30 mm |
| VoI A >> P | 15 mm |
| TR | 2000 ms |
| TE | 46.00 ms |
| Averages | 1 |
| Filter | None |
| Coil elements | PFC |

### Contrast

| | |
|---|---|
| TR | 2000 ms |
| TE | 46.00 ms |
| Tau | 20.00 ms |
| Averages | 1 |
| Excite flip angle | 90 deg |
| Refocus flip angle | 180 deg |
| Measurements | 1 |

### Resolution - Common

| | |
|---|---|
| Vector size | 256 |

### Geometry - Common

| | |
|---|---|
| Position | L1.2 P49.6 H8.9 mm |
| Orientation | C > T30.0 |
| Rotation | 0 deg |
| VoI F >> H | 30 mm |
| VoI R >> L | 30 mm |
| VoI A >> P | 15 mm |

### Geometry - AutoAlign

| | |
|---|---|
| AutoAlign | Head > Brain |
| Initial Position | L1.2 P49.6 H8.9 |
| L | 1.2 mm |
| P | 49.6 mm |
| H | 8.9 mm |
| Initial Rotation | 0.00 deg |
| Initial Orientation | C > T |
| C > T | 30.0 |
| > S | 0.0 |

### System - Miscellaneous

| | |
|---|---|
| Positioning mode | FIX |
| Table position | H |
| Table position | 0 mm |
| MSMA | S - C - T |
| Sagittal | R >> L |
| Coronal | A >> P |
| Transversal | F >> H |
| Save uncombined | Off |
| AutoAlign | Head > Brain |
| Coil Select Mode | Off - AutoCoilSelect |

### System - Adjustments

| | |
|---|---|
| B0 Shim mode | Tune up |
| B1 Shim mode | TrueForm |
| Adj. water suppr. | Off |
| Confirm freq. adjustment | Off |
| Assume Dominant Fat | Off |
| Assume Silicone | Off |
| Adjustment Tolerance | Auto |

### System - Adjust Volume

| | |
|---|---|
| Position | L1.2 P49.6 H8.9 mm |
| Orientation | C > T30.0 |
| Rotation | 0.00 deg |
| R >> L | 30 mm |
| F >> H | 30 mm |
| A >> P | 15 mm |
| Reset | Off |

### System - Tx/Rx

| | |
|---|---|
| Frequency 1H | 297.207726 MHz |
| Correction factor | 1 |
| Gain | High |
| Img. Scale Cor. | 1.000 |
| Reset | Off |
| ? Ref. amplitude 1H | 0.000 V |

### Physio - Signal1

| | |
|---|---|
| 1st Signal/Mode | None |
| TR | 2000 ms |

### Sequence - Common

| | |
|---|---|
| Delta frequency | 0.0 ppm |
| Phase cycling | None |
| Bandwidth | 100000 Hz |
| Acquisition duration | 2 ms |

### Sequence - Special

| | |
|---|---|
| Type of fit | Linear 6-proj |
| VoI fit factor | 100 % |
| Force spherical fit VoI | Off Force spherical fit VoI |
| Save plots to database | Off |
| Refocus pulses | Normal |
| Excite pulse duration | 6400 us |
| Refocus pulse duration | 5120 us |
| Bar FoV | 384 mm |
| Bar thickness | 10.0 mm |
| Inversion pulse | Off |
| Multi-echo acquisition | Off |

| \\USER\PHCP\MRS\final\PFC linewidth check |
|---|
| TA: 0:23 PM: FIX Vol: 30 ×30 ×15 mmRel. SNR: 1.00 : steam |

### Properties

| | |
|---|---|
| Prio recon | Off |
| Load images to viewer | On |
| Inline movie | Off |
| Auto store images | On |
| Load images to stamp segments | Off |
| Load images to graphic segments | Off |
| Auto open inline display | Off |
| Auto close inline display | Off |
| Start measurement without further preparation | Off |
| Wait for user to start | Off |
| Start measurements | Single measurement |

### Routine

| | |
|---|---|
| Position | L1.2 P49.6 H8.9 mm |
| Orientation | C > T30.0 |
| Rotation | 0 deg |
| Vol F >> H | 30 mm |
| Vol F >> H | 30 mm |
| Vol A >> P | 15 mm |
| TR | 5000 ms |
| TE | 8.00 ms |
| Averages | 1 |
| Filter | None |
| Coil elements | PFC |

### Contrast

| | |
|---|---|
| TR | 5000 ms |
| TE | 8.00 ms |
| TM | 32.00 ms |
| Averages | 1 |
| Flip angle | 90 deg |
| VAPOR | Only RF off |
| VAPOR suppr. | Water suppr. |
| Water s. BW | 135 Hz |
| Water s. delta pos. | 0.00 ppm |
| Measurements | 1 |

### Resolution - Common

| | |
|---|---|
| Vector size | 2048 |

### Geometry - Common

| | |
|---|---|
| Position | L1.2 P49.6 H8.9 mm |
| Orientation | C > T30.0 |
| Rotation | 0 deg |
| Vol F >> H | 30 mm |
| Vol R >> L | 30 mm |
| Vol A >> P | 15 mm |

### Geometry - AutoAlign

| | |
|---|---|
| AutoAlign | Head > Brain |
| Initial Position | L1.2 P49.6 H8.9 |
| L | 1.2 mm |
| P | 49.6 mm |
| H | 8.9 mm |
| Initial Rotation | 0.00 deg |
| Initial Orientation | C > T |
| C > T | 30.0 |
| > S | 0.0 |

### System - Miscellaneous

| | |
|---|---|
| Positioning mode | FIX |
| Table position | H |
| Table position | 0 mm |
| MSMA | S - C - T |
| Sagittal | R >> L |
| Coronal | A >> P |
| Transversal | F >> H |
| Save uncombined | Off |
| AutoAlign | Head > Brain |
| Coil Select Mode | Off - AutoCoilSelect |

### System - Adjustments

| | |
|---|---|
| B0 Shim mode | Brain |
| B1 Shim mode | TrueForm |
| Adj. water suppr. | Off |
| Confirm freq. adjustment | Off |
| Assume Dominant Fat | Off |
| Assume Silicone | Off |
| Adjustment Tolerance | Auto |

### System - Adjust Volume

| | |
|---|---|
| Position | L1.2 P49.6 H8.9 mm |
| Orientation | C > T30.0 |
| Rotation | 0.00 deg |
| R >> L | 30 mm |
| F >> H | 30 mm |
| A >> P | 15 mm |
| Reset | Off |

### System - Tx/Rx

| | |
|---|---|
| Frequency 1H | 297.207726 MHz |
| Correction factor | 1 |
| Gain | High |
| Img. Scale Cor. | 1.000 |
| Reset | Off |
| ? Ref. amplitude 1H | 0.000 V |

### Physio - Signal1

| | |
|---|---|
| 1st Signal/Mode | None |
| TR | 5000 ms |

### Sequence - Common

| | |
|---|---|
| Introduction | On |
| Preparation scans | 2 |
| Delta frequency | 0.0 ppm |
| Phase cycling | Auto |
| Bandwidth | 6000 Hz |
| Acquisition duration | 341 ms |
| Remove oversampling | On |

### Sequence - Nuclei

| | |
|---|---|
| TX/RX Nucleus | 1H |
| TX/RX delta frequency | 0 Hz |
| TX Nucleus | None |
| TX delta frequency | 0 Hz |
| Coil elements | PFC |

### Sequence - Special

| | |
|---|---|
| RF pulse duration | 1280 us |
| Spoiler max. amplitude | 30.0 mT/m |

### Sequence - Special

| | |
|---|---|
| Refocus grad. factor | 1.00 |
| OVS slab thickness | 100.0 mm |
| OVS slab pos. offset | 5.0 mm |
| Spoiler duration | 1500 us |
| Acq. window shift | 200 us |
| Min. settling delay | 100 us |
| Gradient ramp time | 200 us |
| VAPOR flip angle | 60 deg |
| VAPOR delay 8 | 16 ms |
| VAPOR delay 7 | 66 ms |
| OVS pulse duration | 2560 us |
| OVS flip angle RO | 90 deg |
| OVS flip angle PH | 90 deg |
| OVS flip angle SL | 90 deg |
| VAPOR delay 6 | 71 ms |
| VAPOR delay 5 | 112 ms |
| VAPOR delay 4 | 115 ms |
| VAPOR delay 3 | 132 ms |
| VAPOR delay 2 | 110 ms |
| VAPOR delay 1 | 160 ms |
| Enable OVS | On |
| Send ref. scans | Off |
| Inversion pulse | Off |
| Symmetric RF pulses | On |
| Invert SS grad. pol. | Off |
| Shift RO frequency | Off |
| Debug loop type | None |

| \\USER\PHCP\MRS\final\PFC steam eja cal fa 50 10 7 |
|---|
| TA: 0:53 PM: FIX Vol: 30 ×30 ×15 mmRel. SNR: 1.00 : steam |

### Properties

| | |
|---|---|
| Prio recon | Off |
| Load images to viewer | On |
| Inline movie | Off |
| Auto store images | On |
| Load images to stamp segments | Off |
| Load images to graphic segments | Off |
| Auto open inline display | On |
| Auto close inline display | Off |
| Start measurement without further preparation | Off |
| Wait for user to start | Off |
| Start measurements | Single measurement |

### Routine

| | |
|---|---|
| Position | L1.2 P49.6 H8.9 mm |
| Orientation | C > T30.0 |
| Rotation | 0 deg |
| Vol F >> H | 30 mm |
| Vol F >> H | 30 mm |
| Vol A >> P | 15 mm |
| TR | 5000 ms |
| TE | 8.00 ms |
| Averages | 1 |
| Filter | None |
| Coil elements | PFC |

### Contrast

| | |
|---|---|
| TR | 5000 ms |
| TE | 8.00 ms |
| TM | 32.00 ms |
| Averages | 1 |
| Flip angle | 50 deg |
| VAPOR | Only RF off |
| VAPOR suppr. | Water suppr. |
| Water s. BW | 135 Hz |
| Water s. delta pos. | 0.00 ppm |
| Measurements | 7 |

### Resolution - Common

| | |
|---|---|
| Vector size | 2048 |

### Geometry - Common

| | |
|---|---|
| Position | L1.2 P49.6 H8.9 mm |
| Orientation | C > T30.0 |
| Rotation | 0 deg |
| Vol F >> H | 30 mm |
| Vol R >> L | 30 mm |
| Vol A >> P | 15 mm |

### Geometry - AutoAlign

| | |
|---|---|
| AutoAlign | Head > Brain |
| Initial Position | L1.2 P49.6 H8.9 |
| L | 1.2 mm |
| P | 49.6 mm |
| H | 8.9 mm |
| Initial Rotation | 0.00 deg |
| Initial Orientation | C > T |
| C > T | 30.0 |
| > S | 0.0 |

### System - Miscellaneous

| | |
|---|---|
| Positioning mode | FIX |
| Table position | H |
| Table position | 0 mm |
| MSMA | S - C - T |
| Sagittal | R >> L |
| Coronal | A >> P |
| Transversal | F >> H |
| Save uncombined | Off |
| AutoAlign | Head > Brain |
| Coil Select Mode | Off - AutoCoilSelect |

### System - Adjustments

| | |
|---|---|
| B0 Shim mode | Brain |
| B1 Shim mode | TrueForm |
| Adj. water suppr. | Off |
| Confirm freq. adjustment | Off |
| Assume Dominant Fat | Off |
| Assume Silicone | Off |
| Adjustment Tolerance | Auto |

### System - Adjust Volume

| | |
|---|---|
| Position | L1.2 P49.6 H8.9 mm |
| Orientation | C > T30.0 |
| Rotation | 0.00 deg |
| R >> L | 30 mm |
| F >> H | 30 mm |
| A >> P | 15 mm |
| Reset | Off |

### System - Tx/Rx

| | |
|---|---|
| Frequency 1H | 297.207726 MHz |
| Correction factor | 1 |
| Gain | High |
| Img. Scale Cor. | 1.000 |
| Reset | Off |
| ? Ref. amplitude 1H | 0.000 V |

### Physio - Signal1

| | |
|---|---|
| 1st Signal/Mode | None |
| TR | 5000 ms |

### Sequence - Common

| | |
|---|---|
| Introduction | On |
| Preparation scans | 2 |
| Delta frequency | 0.0 ppm |
| Phase cycling | Auto |
| Bandwidth | 6000 Hz |
| Acquisition duration | 341 ms |
| Remove oversampling | On |

### Sequence - Nuclei

| | |
|---|---|
| TX/RX Nucleus | 1H |
| TX/RX delta frequency | 0 Hz |
| TX Nucleus | None |
| TX delta frequency | 0 Hz |
| Coil elements | PFC |

### Sequence - Special

| | |
|---|---|
| RF pulse duration | 1520 us |
| Spoiler max. amplitude | 30.0 mT/m |

#### Sequence - Special

| | |
|---|---|
| Refocus grad. factor | 1.00 |
| OVS slab thickness | 100.0 mm |
| OVS slab pos. offset | 5.0 mm |
| Spoiler duration | 1500 us |
| Acq. window shift | 200 us |
| Min. settling delay | 100 us |
| Gradient ramp time | 200 us |
| VAPOR flip angle | 60 deg |
| VAPOR delay 8 | 16 ms |
| VAPOR delay 7 | 66 ms |
| OVS pulse duration | 2560 us |
| OVS flip angle RO | 90 deg |
| OVS flip angle PH | 90 deg |
| OVS flip angle SL | 90 deg |
| VAPOR delay 6 | 71 ms |
| VAPOR delay 5 | 112 ms |
| VAPOR delay 4 | 115 ms |
| VAPOR delay 3 | 132 ms |
| VAPOR delay 2 | 110 ms |
| VAPOR delay 1 | 160 ms |
| Enable OVS | On |
| Weighted averages | Off |
| Send ref. scans | Off |
| Inversion pulse | Off |
| Symmetric RF pulses | On |
| Invert SS grad. pol. | Off |
| Shift RO frequency | Off |
| Debug loop type | Flip angle |
| Flip angle inc. | 10 deg |
| Measurements | 7 |

**\\USER\PHCP\MRS\final\PFC steam eja cal fa 80 3 10**

TA: 1:08 PM: FIX Vol: 30 ×30 ×15 mmRel. SNR: 1.00 : steam

### Properties

| | |
|---|---|
| Prio recon | Off |
| Load images to viewer | On |
| Inline movie | Off |
| Auto store images | On |
| Load images to stamp segments | Off |
| Load images to graphic segments | Off |
| Auto open inline display | Off |
| Auto close inline display | Off |
| Start measurement without further preparation | Off |
| Wait for user to start | Off |
| Start measurements | Single measurement |

### Routine

| | |
|---|---|
| Position | L1.2 P49.6 H8.9 mm |
| Orientation | C > T30.0 |
| Rotation | 0 deg |
| Vol F >> H | 30 mm |
| Vol F >> H | 30 mm |
| Vol A >> P | 15 mm |
| TR | 5000 ms |
| TE | 8.00 ms |
| Averages | 1 |
| Filter | None |
| Coil elements | PFC |

### Contrast

| | |
|---|---|
| TR | 5000 ms |
| TE | 8.00 ms |
| TM | 32.00 ms |
| Averages | 1 |
| Flip angle | 80 deg |
| VAPOR | Only RF off |
| VAPOR suppr. | Water suppr. |
| Water s. BW | 135 Hz |
| Water s. delta pos. | 0.00 ppm |
| Measurements | 10 |

### Resolution - Common

| | |
|---|---|
| Vector size | 2048 |

### Geometry - Common

| | |
|---|---|
| Position | L1.2 P49.6 H8.9 mm |
| Orientation | C > T30.0 |
| Rotation | 0 deg |
| Vol F >> H | 30 mm |
| Vol R >> L | 30 mm |
| Vol A >> P | 15 mm |

### Geometry - AutoAlign

| | |
|---|---|
| AutoAlign | Head > Brain |
| Initial Position | L1.2 P49.6 H8.9 |
| L | 1.2 mm |
| P | 49.6 mm |
| H | 8.9 mm |
| Initial Rotation | 0.00 deg |
| Initial Orientation | C > T |
| C > T | 30.0 |
| > S | 0.0 |

### System - Miscellaneous

| | |
|---|---|
| Positioning mode | FIX |
| Table position | H |
| Table position | 0 mm |
| MSMA | S - C - T |
| Sagittal | R >> L |
| Coronal | A >> P |
| Transversal | F >> H |
| Save uncombined | Off |
| AutoAlign | Head > Brain |
| Coil Select Mode | Off - AutoCoilSelect |

### System - Adjustments

| | |
|---|---|
| B0 Shim mode | Brain |
| B1 Shim mode | TrueForm |
| Adj. water suppr. | Off |
| Confirm freq. adjustment | Off |
| Assume Dominant Fat | Off |
| Assume Silicone | Off |
| Adjustment Tolerance | Auto |

### System - Adjust Volume

| | |
|---|---|
| Position | L1.2 P49.6 H8.9 mm |
| Orientation | C > T30.0 |
| Rotation | 0.00 deg |
| R >> L | 30 mm |
| F >> H | 30 mm |
| A >> P | 15 mm |
| Reset | Off |

### System - Tx/Rx

| | |
|---|---|
| Frequency 1H | 297.207726 MHz |
| Correction factor | 1 |
| Gain | High |
| Img. Scale Cor. | 1.000 |
| Reset | Off |
| ? Ref. amplitude 1H | 0.000 V |

### Physio - Signal1

| | |
|---|---|
| 1st Signal/Mode | None |
| TR | 5000 ms |

### Sequence - Common

| | |
|---|---|
| Introduction | On |
| Preparation scans | 2 |
| Delta frequency | 0.0 ppm |
| Phase cycling | Auto |
| Bandwidth | 6000 Hz |
| Acquisition duration | 341 ms |
| Remove oversampling | On |

### Sequence - Nuclei

| | |
|---|---|
| TX/RX Nucleus | 1H |
| TX/RX delta frequency | 0 Hz |
| TX Nucleus | None |
| TX delta frequency | 0 Hz |
| Coil elements | PFC |

### Sequence - Special

| | |
|---|---|
| RF pulse duration | 1520 us |
| Spoiler max. amplitude | 30.0 mT/m |

### Sequence - Special

| | |
|---|---|
| Refocus grad. factor | 1.00 |
| OVS slab thickness | 100.0 mm |
| OVS slab pos. offset | 5.0 mm |
| Spoiler duration | 1500 us |
| Acq. window shift | 200 us |
| Min. settling delay | 100 us |
| Gradient ramp time | 200 us |
| VAPOR flip angle | 60 deg |
| VAPOR delay 8 | 16 ms |
| VAPOR delay 7 | 66 ms |
| OVS pulse duration | 2560 us |
| OVS flip angle RO | 90 deg |
| OVS flip angle PH | 90 deg |
| OVS flip angle SL | 90 deg |
| VAPOR delay 6 | 71 ms |
| VAPOR delay 5 | 112 ms |
| VAPOR delay 4 | 115 ms |
| VAPOR delay 3 | 132 ms |
| VAPOR delay 2 | 110 ms |
| VAPOR delay 1 | 160 ms |
| Enable OVS | On |
| Weighted averages | Off |
| Send ref. scans | Off |
| Inversion pulse | Off |
| Symmetric RF pulses | On |
| Invert SS grad. pol. | Off |
| Shift RO frequency | Off |
| Debug loop type | Flip angle |
| Flip angle inc. | 3 deg |
| Measurements | 10 |

| \\USER\PHCP\MRS\final\PFC steam eja ws check |
|---|
| TA: 0:38 PM: FIX Vol: 30 ×30 ×15 mmRel. SNR: 1.00 : steam |

### Properties

| | |
|---|---|
| Prio recon | Off |
| Load images to viewer | On |
| Inline movie | Off |
| Auto store images | On |
| Load images to stamp segments | Off |
| Load images to graphic segments | Off |
| Auto open inline display | Off |
| Auto close inline display | Off |
| Start measurement without further preparation | Off |
| Wait for user to start | Off |
| Start measurements | Single measurement |

### Routine

| | |
|---|---|
| Position | L1.2 P49.6 H8.9 mm |
| Orientation | C > T30.0 |
| Rotation | 0 deg |
| Vol F >> H | 30 mm |
| Vol F >> H | 30 mm |
| Vol A >> P | 15 mm |
| TR | 5000 ms |
| TE | 8.00 ms |
| Averages | 4 |
| Filter | None |
| Coil elements | PFC |

### Contrast

| | |
|---|---|
| TR | 5000 ms |
| TE | 8.00 ms |
| TM | 32.00 ms |
| Averages | 4 |
| Flip angle | 90 deg |
| VAPOR | Enabled |
| VAPOR suppr. | Water suppr. |
| Water s. BW | 135 Hz |
| Water s. delta pos. | 0.00 ppm |
| Measurements | 1 |

### Resolution - Common

| | |
|---|---|
| Vector size | 2048 |

### Geometry - Common

| | |
|---|---|
| Position | L1.2 P49.6 H8.9 mm |
| Orientation | C > T30.0 |
| Rotation | 0 deg |
| Vol F >> H | 30 mm |
| Vol R >> L | 30 mm |
| Vol A >> P | 15 mm |

### Geometry - AutoAlign

| | |
|---|---|
| AutoAlign | Head > Brain |
| Initial Position | L1.2 P49.6 H8.9 |
| L | 1.2 mm |
| P | 49.6 mm |
| H | 8.9 mm |
| Initial Rotation | 0.00 deg |
| Initial Orientation | C > T |
| C > T | 30.0 |
| > S | 0.0 |

### System - Miscellaneous

| | |
|---|---|
| Positioning mode | FIX |
| Table position | H |
| Table position | 0 mm |
| MSMA | S - C - T |
| Sagittal | R >> L |
| Coronal | A >> P |
| Transversal | F >> H |
| Save uncombined | Off |
| AutoAlign | Head > Brain |
| Coil Select Mode | Off - AutoCoilSelect |

### System - Adjustments

| | |
|---|---|
| B0 Shim mode | Brain |
| B1 Shim mode | TrueForm |
| Adj. water suppr. | Off |
| Confirm freq. adjustment | Off |
| Assume Dominant Fat | Off |
| Assume Silicone | Off |
| Adjustment Tolerance | Auto |

### System - Adjust Volume

| | |
|---|---|
| Position | L1.2 P49.6 H8.9 mm |
| Orientation | C > T30.0 |
| Rotation | 0.00 deg |
| R >> L | 30 mm |
| F >> H | 30 mm |
| A >> P | 15 mm |
| Reset | Off |

### System - Tx/Rx

| | |
|---|---|
| Frequency 1H | 297.207726 MHz |
| Correction factor | 1 |
| Gain | High |
| Img. Scale Cor. | 1.000 |
| Reset | Off |
| ? Ref. amplitude 1H | 0.000 V |

### Physio - Signal1

| | |
|---|---|
| 1st Signal/Mode | None |
| TR | 5000 ms |

### Sequence - Common

| | |
|---|---|
| Introduction | On |
| Preparation scans | 2 |
| Delta frequency | -1.7 ppm |
| Phase cycling | Auto |
| Bandwidth | 6000 Hz |
| Acquisition duration | 341 ms |
| Remove oversampling | On |

### Sequence - Nuclei

| | |
|---|---|
| TX/RX Nucleus | 1H |
| TX/RX delta frequency | 0 Hz |
| TX Nucleus | None |
| TX delta frequency | 0 Hz |
| Coil elements | PFC |

### Sequence - Special

| | |
|---|---|
| RF pulse duration | 1280 us |
| Spoiler max. amplitude | 30.0 mT/m |

### Sequence - Special

| | |
|---|---|
| Refocus grad. factor | 1.00 |
| OVS slab thickness | 100.0 mm |
| OVS slab pos. offset | 5.0 mm |
| Spoiler duration | 1500 us |
| Acq. window shift | 200 us |
| Min. settling delay | 100 us |
| Gradient ramp time | 200 us |
| VAPOR flip angle | 72 deg |
| VAPOR delay 8 | 16 ms |
| VAPOR delay 7 | 66 ms |
| OVS pulse duration | 2560 us |
| OVS flip angle RO | 90 deg |
| OVS flip angle PH | 90 deg |
| OVS flip angle SL | 90 deg |
| VAPOR delay 6 | 71 ms |
| VAPOR delay 5 | 112 ms |
| VAPOR delay 4 | 115 ms |
| VAPOR delay 3 | 132 ms |
| VAPOR delay 2 | 110 ms |
| VAPOR delay 1 | 160 ms |
| Enable OVS | On |
| Resolve averages | On |
| Send ref. scans | Off |
| Inversion pulse | Off |
| Symmetric RF pulses | On |
| Invert SS grad. pol. | Off |
| Shift RO frequency | Off |
| Debug loop type | None |

| \\USER\PHCP\MRS\final\PFC steam eja metab |
|---|
| TA: 8:33 PM: FIX Vol: 30 ×30 ×15 mmRel. SNR: 1.00 : steam |

### Properties

| | |
|---|---|
| Prio recon | Off |
| Load images to viewer | On |
| Inline movie | Off |
| Auto store images | On |
| Load images to stamp segments | Off |
| Load images to graphic segments | Off |
| Auto open inline display | Off |
| Auto close inline display | Off |
| Start measurement without further preparation | Off |
| Wait for user to start | Off |
| Start measurements | Single measurement |

### Routine

| | |
|---|---|
| Position | L1.2 P49.6 H8.9 mm |
| Orientation | C > T30.0 |
| Rotation | 0 deg |
| Vol F >> H | 30 mm |
| Vol F >> H | 30 mm |
| Vol A >> P | 15 mm |
| TR | 5000 ms |
| TE | 8.00 ms |
| Averages | 96 |
| Filter | None |
| Coil elements | PFC |

### Contrast

| | |
|---|---|
| TR | 5000 ms |
| TE | 8.00 ms |
| TM | 32.00 ms |
| Averages | 96 |
| Flip angle | 90 deg |
| VAPOR | Enabled |
| VAPOR suppr. | Water suppr. |
| Water s. BW | 135 Hz |
| Water s. delta pos. | 0.00 ppm |
| Measurements | 1 |

### Resolution - Common

| | |
|---|---|
| Vector size | 2048 |

### Geometry - Common

| | |
|---|---|
| Position | L1.2 P49.6 H8.9 mm |
| Orientation | C > T30.0 |
| Rotation | 0 deg |
| Vol F >> H | 30 mm |
| Vol R >> L | 30 mm |
| Vol A >> P | 15 mm |

### Geometry - AutoAlign

| | |
|---|---|
| AutoAlign | Head > Brain |
| Initial Position | L1.2 P49.6 H8.9 |
| L | 1.2 mm |
| P | 49.6 mm |
| H | 8.9 mm |
| Initial Rotation | 0.00 deg |
| Initial Orientation | C > T |
| C > T | 30.0 |
| > S | 0.0 |

### System - Miscellaneous

| | |
|---|---|
| Positioning mode | FIX |
| Table position | H |
| Table position | 0 mm |
| MSMA | S - C - T |
| Sagittal | R >> L |
| Coronal | A >> P |
| Transversal | F >> H |
| Save uncombined | Off |
| AutoAlign | Head > Brain |
| Coil Select Mode | Off - AutoCoilSelect |

### System - Adjustments

| | |
|---|---|
| B0 Shim mode | Brain |
| B1 Shim mode | TrueForm |
| Adj. water suppr. | Off |
| Confirm freq. adjustment | Off |
| Assume Dominant Fat | Off |
| Assume Silicone | Off |
| Adjustment Tolerance | Auto |

### System - Adjust Volume

| | |
|---|---|
| Position | L1.2 P49.6 H8.9 mm |
| Orientation | C > T30.0 |
| Rotation | 0.00 deg |
| R >> L | 30 mm |
| F >> H | 30 mm |
| A >> P | 15 mm |
| Reset | Off |

### System - Tx/Rx

| | |
|---|---|
| Frequency 1H | 297.207726 MHz |
| Correction factor | 1 |
| Gain | High |
| Img. Scale Cor. | 1.000 |
| Reset | Off |
| ? Ref. amplitude 1H | 0.000 V |

### Physio - Signal1

| | |
|---|---|
| 1st Signal/Mode | None |
| TR | 5000 ms |

### Sequence - Common

| | |
|---|---|
| Introduction | On |
| Preparation scans | 2 |
| Delta frequency | -1.7 ppm |
| Phase cycling | Auto |
| Bandwidth | 6000 Hz |
| Acquisition duration | 341 ms |
| Remove oversampling | On |

### Sequence - Nuclei

| | |
|---|---|
| TX/RX Nucleus | 1H |
| TX/RX delta frequency | 0 Hz |
| TX Nucleus | None |
| TX delta frequency | 0 Hz |
| Coil elements | PFC |

### Sequence - Special

| | |
|---|---|
| RF pulse duration | 1280 us |
| Spoiler max. amplitude | 30.0 mT/m |

**Sequence - Special**

| | |
|---|---|
| Refocus grad. factor | 1.00 |
| OVS slab thickness | 100.0 mm |
| OVS slab pos. offset | 5.0 mm |
| Spoiler duration | 1500 us |
| Acq. window shift | 200 us |
| Min. settling delay | 100 us |
| Gradient ramp time | 200 us |
| VAPOR flip angle | 72 deg |
| VAPOR delay 8 | 16 ms |
| VAPOR delay 7 | 66 ms |
| OVS pulse duration | 2560 us |
| OVS flip angle RO | 90 deg |
| OVS flip angle PH | 90 deg |
| OVS flip angle SL | 90 deg |
| VAPOR delay 6 | 71 ms |
| VAPOR delay 5 | 112 ms |
| VAPOR delay 4 | 115 ms |
| VAPOR delay 3 | 132 ms |
| VAPOR delay 2 | 110 ms |
| VAPOR delay 1 | 160 ms |
| Enable OVS | On |
| Resolve averages | On |
| Send ref. scans | Off |
| Inversion pulse | Off |
| Symmetric RF pulses | On |
| Invert SS grad. pol. | Off |
| Shift RO frequency | Off |
| Debug loop type | None |

**\\USER\PHCP\MRS\final\PFC steam eja w1**

TA: 0:23 PM: FIX Vol: 30 ×30 ×15 mmRel. SNR: 1.00 : steam

### Properties

| | |
|---|---|
| Prio recon | Off |
| Load images to viewer | On |
| Inline movie | Off |
| Auto store images | On |
| Load images to stamp segments | Off |
| Load images to graphic segments | Off |
| Auto open inline display | Off |
| Auto close inline display | Off |
| Start measurement without further preparation | Off |
| Wait for user to start | Off |
| Start measurements | Single measurement |

### Routine

| | |
|---|---|
| Position | L1.2 P49.6 H8.9 mm |
| Orientation | C > T30.0 |
| Rotation | 0 deg |
| Vol F >> H | 30 mm |
| Vol F >> H | 30 mm |
| Vol A >> P | 15 mm |
| TR | 5000 ms |
| TE | 8.00 ms |
| Averages | 1 |
| Filter | None |
| Coil elements | PFC |

### Contrast

| | |
|---|---|
| TR | 5000 ms |
| TE | 8.00 ms |
| TM | 32.00 ms |
| Averages | 1 |
| Flip angle | 90 deg |
| VAPOR | Only RF off |
| VAPOR suppr. | Water suppr. |
| Water s. BW | 135 Hz |
| Water s. delta pos. | 0.00 ppm |
| Measurements | 1 |

### Resolution - Common

| | |
|---|---|
| Vector size | 2048 |

### Geometry - Common

| | |
|---|---|
| Position | L1.2 P49.6 H8.9 mm |
| Orientation | C > T30.0 |
| Rotation | 0 deg |
| Vol F >> H | 30 mm |
| Vol R >> L | 30 mm |
| Vol A >> P | 15 mm |

### Geometry - AutoAlign

| | |
|---|---|
| AutoAlign | Head > Brain |
| Initial Position | L1.2 P49.6 H8.9 |
| L | 1.2 mm |
| P | 49.6 mm |
| H | 8.9 mm |
| Initial Rotation | 0.00 deg |
| Initial Orientation | C > T |
| C > T | 30.0 |
| > S | 0.0 |

### System - Miscellaneous

| | |
|---|---|
| Positioning mode | FIX |
| Table position | H |
| Table position | 0 mm |
| MSMA | S - C - T |
| Sagittal | R >> L |
| Coronal | A >> P |
| Transversal | F >> H |
| Save uncombined | Off |
| AutoAlign | Head > Brain |
| Coil Select Mode | Off - AutoCoilSelect |

### System - Adjustments

| | |
|---|---|
| B0 Shim mode | Brain |
| B1 Shim mode | TrueForm |
| Adj. water suppr. | Off |
| Confirm freq. adjustment | Off |
| Assume Dominant Fat | Off |
| Assume Silicone | Off |
| Adjustment Tolerance | Auto |

### System - Adjust Volume

| | |
|---|---|
| Position | L1.2 P49.6 H8.9 mm |
| Orientation | C > T30.0 |
| Rotation | 0.00 deg |
| R >> L | 30 mm |
| F >> H | 30 mm |
| A >> P | 15 mm |
| Reset | Off |

### System - Tx/Rx

| | |
|---|---|
| Frequency 1H | 297.207726 MHz |
| Correction factor | 1 |
| Gain | High |
| Img. Scale Cor. | 1.000 |
| Reset | Off |
| ? Ref. amplitude 1H | 0.000 V |

### Physio - Signal1

| | |
|---|---|
| 1st Signal/Mode | None |
| TR | 5000 ms |

### Sequence - Common

| | |
|---|---|
| Introduction | On |
| Preparation scans | 2 |
| Delta frequency | 0.0 ppm |
| Phase cycling | Auto |
| Bandwidth | 6000 Hz |
| Acquisition duration | 341 ms |
| Remove oversampling | On |

### Sequence - Nuclei

| | |
|---|---|
| TX/RX Nucleus | 1H |
| TX/RX delta frequency | 0 Hz |
| TX Nucleus | None |
| TX delta frequency | 0 Hz |
| Coil elements | PFC |

### Sequence - Special

| | |
|---|---|
| RF pulse duration | 1280 us |
| Spoiler max. amplitude | 30.0 mT/m |

### Sequence - Special

| | |
|---|---|
| Refocus grad. factor | 1.00 |
| OVS slab thickness | 100.0 mm |
| OVS slab pos. offset | 5.0 mm |
| Spoiler duration | 1500 us |
| Acq. window shift | 200 us |
| Min. settling delay | 100 us |
| Gradient ramp time | 200 us |
| VAPOR flip angle | 60 deg |
| VAPOR delay 8 | 16 ms |
| VAPOR delay 7 | 66 ms |
| OVS pulse duration | 2560 us |
| OVS flip angle RO | 90 deg |
| OVS flip angle PH | 90 deg |
| OVS flip angle SL | 90 deg |
| VAPOR delay 6 | 71 ms |
| VAPOR delay 5 | 112 ms |
| VAPOR delay 4 | 115 ms |
| VAPOR delay 3 | 132 ms |
| VAPOR delay 2 | 110 ms |
| VAPOR delay 1 | 160 ms |
| Enable OVS | On |
| Send ref. scans | Off |
| Inversion pulse | Off |
| Symmetric RF pulses | On |
| Invert SS grad. pol. | Off |
| Shift RO frequency | Off |
| Debug loop type | None |

| \\USER\PHCP\MRS\final\PFC steam eja w4 |
|---|
| TA: 0:38 PM: FIX Vol: 30 ×30 ×15 mmRel. SNR: 1.00 : steam |

### Properties

| | |
|---|---|
| Prio recon | Off |
| Load images to viewer | On |
| Inline movie | Off |
| Auto store images | On |
| Load images to stamp segments | Off |
| Load images to graphic segments | Off |
| Auto open inline display | Off |
| Auto close inline display | Off |
| Start measurement without further preparation | Off |
| Wait for user to start | Off |
| Start measurements | Single measurement |

### Routine

| | |
|---|---|
| Position | L1.2 P49.6 H8.9 mm |
| Orientation | C > T30.0 |
| Rotation | 0 deg |
| Vol F >> H | 30 mm |
| Vol F >> H | 30 mm |
| Vol A >> P | 15 mm |
| TR | 5000 ms |
| TE | 8.00 ms |
| Averages | 4 |
| Filter | None |
| Coil elements | PFC |

### Contrast

| | |
|---|---|
| TR | 5000 ms |
| TE | 8.00 ms |
| TM | 32.00 ms |
| Averages | 4 |
| Flip angle | 90 deg |
| VAPOR | Only RF off |
| VAPOR suppr. | Water suppr. |
| Water s. BW | 135 Hz |
| Water s. delta pos. | 0.00 ppm |
| Measurements | 1 |

### Resolution - Common

| | |
|---|---|
| Vector size | 2048 |

### Geometry - Common

| | |
|---|---|
| Position | L1.2 P49.6 H8.9 mm |
| Orientation | C > T30.0 |
| Rotation | 0 deg |
| Vol F >> H | 30 mm |
| Vol R >> L | 30 mm |
| Vol A >> P | 15 mm |

### Geometry - AutoAlign

| | |
|---|---|
| AutoAlign | Head > Brain |
| Initial Position | L1.2 P49.6 H8.9 |
| L | 1.2 mm |
| P | 49.6 mm |
| H | 8.9 mm |
| Initial Rotation | 0.00 deg |
| Initial Orientation | C > T |
| C > T | 30.0 |
| > S | 0.0 |

### System - Miscellaneous

| | |
|---|---|
| Positioning mode | FIX |
| Table position | H |
| Table position | 0 mm |
| MSMA | S - C - T |
| Sagittal | R >> L |
| Coronal | A >> P |
| Transversal | F >> H |
| Save uncombined | Off |
| AutoAlign | Head > Brain |
| Coil Select Mode | Off - AutoCoilSelect |

### System - Adjustments

| | |
|---|---|
| B0 Shim mode | Brain |
| B1 Shim mode | TrueForm |
| Adj. water suppr. | Off |
| Confirm freq. adjustment | Off |
| Assume Dominant Fat | Off |
| Assume Silicone | Off |
| Adjustment Tolerance | Auto |

### System - Adjust Volume

| | |
|---|---|
| Position | L1.2 P49.6 H8.9 mm |
| Orientation | C > T30.0 |
| Rotation | 0.00 deg |
| R >> L | 30 mm |
| F >> H | 30 mm |
| A >> P | 15 mm |
| Reset | Off |

### System - Tx/Rx

| | |
|---|---|
| Frequency 1H | 297.207726 MHz |
| Correction factor | 1 |
| Gain | High |
| Img. Scale Cor. | 1.000 |
| Reset | Off |
| ? Ref. amplitude 1H | 0.000 V |

### Physio - Signal1

| | |
|---|---|
| 1st Signal/Mode | None |
| TR | 5000 ms |

### Sequence - Common

| | |
|---|---|
| Introduction | On |
| Preparation scans | 2 |
| Delta frequency | 0.0 ppm |
| Phase cycling | Auto |
| Bandwidth | 6000 Hz |
| Acquisition duration | 341 ms |
| Remove oversampling | On |

### Sequence - Nuclei

| | |
|---|---|
| TX/RX Nucleus | 1H |
| TX/RX delta frequency | 0 Hz |
| TX Nucleus | None |
| TX delta frequency | 0 Hz |
| Coil elements | PFC |

### Sequence - Special

| | |
|---|---|
| RF pulse duration | 1280 us |
| Spoiler max. amplitude | 30.0 mT/m |

### Sequence - Special

| | |
|---|---|
| Refocus grad. factor | 1.00 |
| OVS slab thickness | 100.0 mm |
| OVS slab pos. offset | 5.0 mm |
| Spoiler duration | 1500 us |
| Acq. window shift | 200 us |
| Min. settling delay | 100 us |
| Gradient ramp time | 200 us |
| VAPOR flip angle | 60 deg |
| VAPOR delay 8 | 16 ms |
| VAPOR delay 7 | 66 ms |
| OVS pulse duration | 2560 us |
| OVS flip angle RO | 90 deg |
| OVS flip angle PH | 90 deg |
| OVS flip angle SL | 90 deg |
| VAPOR delay 6 | 71 ms |
| VAPOR delay 5 | 112 ms |
| VAPOR delay 4 | 115 ms |
| VAPOR delay 3 | 132 ms |
| VAPOR delay 2 | 110 ms |
| VAPOR delay 1 | 160 ms |
| Enable OVS | On |
| Resolve averages | On |
| Send ref. scans | Off |
| Inversion pulse | Off |
| Symmetric RF pulses | On |
| Invert SS grad. pol. | Off |
| Shift RO frequency | Off |
| Debug loop type | None |

| \\USER\PHCP\MRS\final\PFC steam eja w1 noOVS |
|---|
| TA: 0:23 PM: FIX Vol: 30 ×30 ×15 mmRel. SNR: 1.00 : steam |

### Properties

| | |
|---|---|
| Prio recon | Off |
| Load images to viewer | On |
| Inline movie | Off |
| Auto store images | On |
| Load images to stamp segments | Off |
| Load images to graphic segments | Off |
| Auto open inline display | Off |
| Auto close inline display | Off |
| Start measurement without further preparation | Off |
| Wait for user to start | Off |
| Start measurements | Single measurement |

### Routine

| | |
|---|---|
| Position | L1.2 P49.6 H8.9 mm |
| Orientation | C > T30.0 |
| Rotation | 0 deg |
| Vol F >> H | 30 mm |
| Vol F >> H | 30 mm |
| Vol A >> P | 15 mm |
| TR | 5000 ms |
| TE | 8.00 ms |
| Averages | 1 |
| Filter | None |
| Coil elements | PFC |

### Contrast

| | |
|---|---|
| TR | 5000 ms |
| TE | 8.00 ms |
| TM | 32.00 ms |
| Averages | 1 |
| Flip angle | 90 deg |
| VAPOR | Only RF off |
| VAPOR suppr. | Water suppr. |
| Water s. BW | 135 Hz |
| Water s. delta pos. | 0.00 ppm |
| Measurements | 1 |

### Resolution - Common

| | |
|---|---|
| Vector size | 2048 |

### Geometry - Common

| | |
|---|---|
| Position | L1.2 P49.6 H8.9 mm |
| Orientation | C > T30.0 |
| Rotation | 0 deg |
| Vol F >> H | 30 mm |
| Vol R >> L | 30 mm |
| Vol A >> P | 15 mm |

### Geometry - AutoAlign

| | |
|---|---|
| AutoAlign | Head > Brain |
| Initial Position | L1.2 P49.6 H8.9 |
| L | 1.2 mm |
| P | 49.6 mm |
| H | 8.9 mm |
| Initial Rotation | 0.00 deg |
| Initial Orientation | C > T |
| C > T | 30.0 |
| > S | 0.0 |

### System - Miscellaneous

| | |
|---|---|
| Positioning mode | FIX |
| Table position | H |
| Table position | 0 mm |
| MSMA | S - C - T |
| Sagittal | R >> L |
| Coronal | A >> P |
| Transversal | F >> H |
| Save uncombined | Off |
| AutoAlign | Head > Brain |
| Coil Select Mode | Off - AutoCoilSelect |

### System - Adjustments

| | |
|---|---|
| B0 Shim mode | Brain |
| B1 Shim mode | TrueForm |
| Adj. water suppr. | Off |
| Confirm freq. adjustment | Off |
| Assume Dominant Fat | Off |
| Assume Silicone | Off |
| Adjustment Tolerance | Auto |

### System - Adjust Volume

| | |
|---|---|
| Position | L1.2 P49.6 H8.9 mm |
| Orientation | C > T30.0 |
| Rotation | 0.00 deg |
| R >> L | 30 mm |
| F >> H | 30 mm |
| A >> P | 15 mm |
| Reset | Off |

### System - Tx/Rx

| | |
|---|---|
| Frequency 1H | 297.207726 MHz |
| Correction factor | 1 |
| Gain | High |
| Img. Scale Cor. | 1.000 |
| Reset | Off |
| ? Ref. amplitude 1H | 0.000 V |

### Physio - Signal1

| | |
|---|---|
| 1st Signal/Mode | None |
| TR | 5000 ms |

### Sequence - Common

| | |
|---|---|
| Introduction | On |
| Preparation scans | 2 |
| Delta frequency | 0.0 ppm |
| Phase cycling | Auto |
| Bandwidth | 6000 Hz |
| Acquisition duration | 341 ms |
| Remove oversampling | On |

### Sequence - Nuclei

| | |
|---|---|
| TX/RX Nucleus | 1H |
| TX/RX delta frequency | 0 Hz |
| TX Nucleus | None |
| TX delta frequency | 0 Hz |
| Coil elements | PFC |

### Sequence - Special

| | |
|---|---|
| RF pulse duration | 1280 us |
| Spoiler max. amplitude | 30.0 mT/m |

### Sequence - Special

| | |
|---|---|
| Refocus grad. factor | 1.00 |
| Spoiler duration | 1500 us |
| Acq. window shift | 200 us |
| Min. settling delay | 100 us |
| Gradient ramp time | 200 us |
| VAPOR flip angle | 60 deg |
| VAPOR delay 8 | 16 ms |
| VAPOR delay 7 | 66 ms |
| VAPOR delay 6 | 71 ms |
| VAPOR delay 5 | 112 ms |
| VAPOR delay 4 | 115 ms |
| VAPOR delay 3 | 132 ms |
| VAPOR delay 2 | 110 ms |
| VAPOR delay 1 | 160 ms |
| Enable OVS | Off |
| Send ref. scans | Off |
| Inversion pulse | Off |
| Symmetric RF pulses | On |
| Invert SS grad. pol. | Off |
| Shift RO frequency | Off |
| Debug loop type | None |

**\\USER\PHCP\MRS\final\PFC steam eja ws FA optimization**

TA: 1:08 PM: FIX Vol: 30 ×30 ×15 mmRel. SNR: 1.00 : steam

### Properties

| | |
|---|---|
| Prio recon | Off |
| Load images to viewer | On |
| Inline movie | Off |
| Auto store images | On |
| Load images to stamp segments | Off |
| Load images to graphic segments | Off |
| Auto open inline display | Off |
| Auto close inline display | Off |
| Start measurement without further preparation | Off |
| Wait for user to start | Off |
| Start measurements | Single measurement |

### Routine

| | |
|---|---|
| Position | L1.2 P49.6 H8.9 mm |
| Orientation | C > T30.0 |
| Rotation | 0 deg |
| Vol F >> H | 30 mm |
| Vol F >> H | 30 mm |
| Vol A >> P | 15 mm |
| TR | 5000 ms |
| TE | 8.00 ms |
| Averages | 1 |
| Filter | None |
| Coil elements | PFC |

### Contrast

| | |
|---|---|
| TR | 5000 ms |
| TE | 8.00 ms |
| TM | 32.00 ms |
| Averages | 1 |
| Flip angle | 90 deg |
| VAPOR | Enabled |
| VAPOR suppr. | Water suppr. |
| Water s. BW | 135 Hz |
| Water s. delta pos. | 0.00 ppm |
| Measurements | 10 |

### Resolution - Common

| | |
|---|---|
| Vector size | 2048 |

### Geometry - Common

| | |
|---|---|
| Position | L1.2 P49.6 H8.9 mm |
| Orientation | C > T30.0 |
| Rotation | 0 deg |
| Vol F >> H | 30 mm |
| Vol R >> L | 30 mm |
| Vol A >> P | 15 mm |

### Geometry - AutoAlign

| | |
|---|---|
| AutoAlign | Head > Brain |
| Initial Position | L1.2 P49.6 H8.9 |
| L | 1.2 mm |
| P | 49.6 mm |
| H | 8.9 mm |
| Initial Rotation | 0.00 deg |
| Initial Orientation | C > T |
| C > T | 30.0 |
| > S | 0.0 |

### System - Miscellaneous

| | |
|---|---|
| Positioning mode | FIX |
| Table position | H |
| Table position | 0 mm |
| MSMA | S - C - T |
| Sagittal | R >> L |
| Coronal | A >> P |
| Transversal | F >> H |
| Save uncombined | Off |
| AutoAlign | Head > Brain |
| Coil Select Mode | Off - AutoCoilSelect |

### System - Adjustments

| | |
|---|---|
| B0 Shim mode | Brain |
| B1 Shim mode | TrueForm |
| Adj. water suppr. | Off |
| Confirm freq. adjustment | Off |
| Assume Dominant Fat | Off |
| Assume Silicone | Off |
| Adjustment Tolerance | Auto |

### System - Adjust Volume

| | |
|---|---|
| Position | L1.2 P49.6 H8.9 mm |
| Orientation | C > T30.0 |
| Rotation | 0.00 deg |
| R >> L | 30 mm |
| F >> H | 30 mm |
| A >> P | 15 mm |
| Reset | Off |

### System - Tx/Rx

| | |
|---|---|
| Frequency 1H | 297.207726 MHz |
| Correction factor | 1 |
| Gain | High |
| Img. Scale Cor. | 1.000 |
| Reset | Off |
| ? Ref. amplitude 1H | 0.000 V |

### Physio - Signal1

| | |
|---|---|
| 1st Signal/Mode | None |
| TR | 5000 ms |

### Sequence - Common

| | |
|---|---|
| Introduction | On |
| Preparation scans | 2 |
| Delta frequency | -1.7 ppm |
| Phase cycling | Auto |
| Bandwidth | 6000 Hz |
| Acquisition duration | 341 ms |
| Remove oversampling | On |

### Sequence - Nuclei

| | |
|---|---|
| TX/RX Nucleus | 1H |
| TX/RX delta frequency | 0 Hz |
| TX Nucleus | None |
| TX delta frequency | 0 Hz |
| Coil elements | PFC |

### Sequence - Special

| | |
|---|---|
| RF pulse duration | 1280 us |
| Spoiler max. amplitude | 30.0 mT/m |

### Sequence - Special

| | |
|---|---|
| Refocus grad. factor | 1.00 |
| OVS slab thickness | 100.0 mm |
| OVS slab pos. offset | 5.0 mm |
| Spoiler duration | 1500 us |
| Acq. window shift | 200 us |
| Min. settling delay | 100 us |
| Gradient ramp time | 200 us |
| VAPOR flip angle | 60 deg |
| VAPOR delay 8 | 16 ms |
| VAPOR delay 7 | 66 ms |
| OVS pulse duration | 2560 us |
| OVS flip angle RO | 90 deg |
| OVS flip angle PH | 90 deg |
| OVS flip angle SL | 90 deg |
| VAPOR delay 6 | 71 ms |
| VAPOR delay 5 | 112 ms |
| VAPOR delay 4 | 115 ms |
| VAPOR delay 3 | 132 ms |
| VAPOR delay 2 | 110 ms |
| VAPOR delay 1 | 160 ms |
| Enable OVS | On |
| Weighted averages | Off |
| Send ref. scans | Off |
| Inversion pulse | Off |
| Symmetric RF pulses | On |
| Invert SS grad. pol. | Off |
| Shift RO frequency | Off |
| Debug loop type | WS FA |
| WS FA inc. | 2 deg |
| Measurements | 10 |

| \\USER\PHCP\MRS\final\PFC steam eja ws delay 7 |
|---|
| TA: 1:08 PM: FIX Vol: 30 ×30 ×15 mmRel. SNR: 1.00 : steam |

### Properties

| | |
|---|---|
| Prio recon | Off |
| Load images to viewer | On |
| Inline movie | Off |
| Auto store images | On |
| Load images to stamp segments | Off |
| Load images to graphic segments | Off |
| Auto open inline display | Off |
| Auto close inline display | Off |
| Start measurement without further preparation | Off |
| Wait for user to start | Off |
| Start measurements | Single measurement |

### Routine

| | |
|---|---|
| Position | L1.2 P49.6 H8.9 mm |
| Orientation | C > T30.0 |
| Rotation | 0 deg |
| Vol F >> H | 30 mm |
| Vol F >> H | 30 mm |
| Vol A >> P | 15 mm |
| TR | 5000 ms |
| TE | 8.00 ms |
| Averages | 1 |
| Filter | None |
| Coil elements | PFC |

### Contrast

| | |
|---|---|
| TR | 5000 ms |
| TE | 8.00 ms |
| TM | 32.00 ms |
| Averages | 1 |
| Flip angle | 90 deg |
| VAPOR | Enabled |
| VAPOR suppr. | Water suppr. |
| Water s. BW | 135 Hz |
| Water s. delta pos. | 0.00 ppm |
| Measurements | 10 |

### Resolution - Common

| | |
|---|---|
| Vector size | 2048 |

### Geometry - Common

| | |
|---|---|
| Position | L1.2 P49.6 H8.9 mm |
| Orientation | C > T30.0 |
| Rotation | 0 deg |
| Vol F >> H | 30 mm |
| Vol R >> L | 30 mm |
| Vol A >> P | 15 mm |

### Geometry - AutoAlign

| | |
|---|---|
| AutoAlign | Head > Brain |
| Initial Position | L1.2 P49.6 H8.9 |
| L | 1.2 mm |
| P | 49.6 mm |
| H | 8.9 mm |
| Initial Rotation | 0.00 deg |
| Initial Orientation | C > T |
| C > T | 30.0 |
| > S | 0.0 |

### System - Miscellaneous

| | |
|---|---|
| Positioning mode | FIX |
| Table position | H |
| Table position | 0 mm |
| MSMA | S - C - T |
| Sagittal | R >> L |
| Coronal | A >> P |
| Transversal | F >> H |
| Save uncombined | Off |
| AutoAlign | Head > Brain |
| Coil Select Mode | Off - AutoCoilSelect |

### System - Adjustments

| | |
|---|---|
| B0 Shim mode | Brain |
| B1 Shim mode | TrueForm |
| Adj. water suppr. | Off |
| Confirm freq. adjustment | Off |
| Assume Dominant Fat | Off |
| Assume Silicone | Off |
| Adjustment Tolerance | Auto |

### System - Adjust Volume

| | |
|---|---|
| Position | L1.2 P49.6 H8.9 mm |
| Orientation | C > T30.0 |
| Rotation | 0.00 deg |
| R >> L | 30 mm |
| F >> H | 30 mm |
| A >> P | 15 mm |
| Reset | Off |

### System - Tx/Rx

| | |
|---|---|
| Frequency 1H | 297.207726 MHz |
| Correction factor | 1 |
| Gain | High |
| Img. Scale Cor. | 1.000 |
| Reset | Off |
| ? Ref. amplitude 1H | 0.000 V |

### Physio - Signal1

| | |
|---|---|
| 1st Signal/Mode | None |
| TR | 5000 ms |

### Sequence - Common

| | |
|---|---|
| Introduction | On |
| Preparation scans | 2 |
| Delta frequency | -1.7 ppm |
| Phase cycling | Auto |
| Bandwidth | 6000 Hz |
| Acquisition duration | 341 ms |
| Remove oversampling | On |

### Sequence - Nuclei

| | |
|---|---|
| TX/RX Nucleus | 1H |
| TX/RX delta frequency | 0 Hz |
| TX Nucleus | None |
| TX delta frequency | 0 Hz |
| Coil elements | PFC |

### Sequence - Special

| | |
|---|---|
| RF pulse duration | 1280 us |
| Spoiler max. amplitude | 30.0 mT/m |

### Sequence - Special

| | |
|---|---|
| Refocus grad. factor | 1.00 |
| OVS slab thickness | 100.0 mm |
| OVS slab pos. offset | 5.0 mm |
| Spoiler duration | 1500 us |
| Acq. window shift | 200 us |
| Min. settling delay | 100 us |
| Gradient ramp time | 200 us |
| VAPOR flip angle | 60 deg |
| VAPOR delay 8 | 16 ms |
| VAPOR delay 7 | 66 ms |
| OVS pulse duration | 2560 us |
| OVS flip angle RO | 90 deg |
| OVS flip angle PH | 90 deg |
| OVS flip angle SL | 90 deg |
| VAPOR delay 6 | 71 ms |
| VAPOR delay 5 | 112 ms |
| VAPOR delay 4 | 115 ms |
| VAPOR delay 3 | 132 ms |
| VAPOR delay 2 | 110 ms |
| VAPOR delay 1 | 160 ms |
| Enable OVS | On |
| Weighted averages | Off |
| Send ref. scans | Off |
| Inversion pulse | Off |
| Symmetric RF pulses | On |
| Invert SS grad. pol. | Off |
| Shift RO frequency | Off |
| Debug loop type | WS delay 7 |
| WS delay 7 inc. | 3 ms |
| Measurements | 10 |
